# Monitoring of plasma and urine tumor-derived DNA to inform bladder-sparing approaches for patients with muscle-invasive bladder cancer

**DOI:** 10.64898/2026.01.14.26344033

**Authors:** Matthew D. Galsky, Sudeh Izadmehr, Menggang Yu, Samuel D. Curtis, Christopher Douville, Maria Popoli, Janine Ptak, Lisa Dobbyn, Natalie Silliman, Kevin G. Chan, Tanya B. Dorff, Jeremy P. Cetnar, Brock O’Neil, Anishka D’Souza, Ronac Mamtani, Christos E. Kyriakopoulos, Rachel Brody, Evita Sadimin, Reza Mehrazin, Diego Chowell, John Sfakianos, Siamak Daneshmand, Sumanta K. Pal, Chetan Bettegowda, Kenneth W. Kinzler, Nickolas Papadopoulos, Bert Vogelstein, Yuxuan Wang

**Affiliations:** Division of Hematology and Medical Oncology, Icahn School of Medicine at Mount Sinai, New York, NY 10029; Tisch Cancer Institute, Icahn School of Medicine at Mount Sinai, New York, NY 10029; Department of Biostatistics, University of Michigan, Ann Arbor, Michigan 48109; Ludwig Center for Cancer Genetics and Therapeutics, Johns Hopkins University School of Medicine, Baltimore, MD 21287; Department of Urology, City of Hope Comprehensive Cancer Center, Duarte, CA 91010; Department of Medical Oncology & Therapeutics, City of Hope Comprehensive Cancer Center, Duarte, CA 91010; Division of Hematology and Medical Oncology, Oregon Health and Science University, Portland, OR; Department of Urology, University of Utah, Salt Lake City, UT 84132; Division of Hematology and Medical Oncology, Keck School of Medicine of USC, Norris Comprehensive Cancer Center, Los Angeles, CA 90089; Division of Hematology and Medical Oncology, University of Pennsylvania Abramson Cancer Center, Philadelphia, PA 19104; Division of Hematology and Medical Oncology, University of Wisconsin Carbone Cancer Center, Madison, WI 53792; Department of Pathology, Molecular and Cell-based Medicine, Icahn School of Medicine at Mount Sinai, New York, NY 10029; Department of Pathology, City of Hope Comprehensive Cancer Center, Duarte, CA, USA; Department of Urology, Icahn School of Medicine at Mount Sinai, Tisch Cancer Institute, New York, NY 10029; Precision Immunology Institute, Icahn School of Medicine at Mount Sinai, New York, NY 10029; Department of Urology, Keck School of Medicine of USC, Norris Comprehensive Cancer Center, Los Angeles, CA 90033

**Author notes:** Contributed equally (Bert Vogelstein, Yuxuan Wang).

**Keywords:** Biomarker, ctDNA, utDNA, bladder cancer

## Abstract

We previously reported initial results from a clinical trial testing a strategy in which patients with muscle-invasive bladder cancer (MIBC) achieving a clinical complete response after cystoscopic resection of the bladder tumor plus systemic therapy could forgo removal of their entire bladder (cystectomy). While the results were highly promising, a subset of patients omitting initial cystectomy developed recurrence highlighting the need for biomarkers to refine selection of patients for this approach. We here report long-term follow-up of these patients and investigate whether tumor DNA in the plasma (ctDNA) or urine (utDNA) could inform prognosis and the need for cystectomy. Three-year bladder-intact survival among patients with a complete clinical response following four rounds of systemic therapy was 69%. Metastatic risk was significantly higher for patients with detectable versus undetectable ctDNA pre-systemic therapy (HR 4.68; 95% CI 1.10-43.35; log-rank p=0.036). Only 4.5% of patients with undetectable baseline ctDNA developed metastatic disease. Undetectable ctDNA before or after systemic therapy was associated with extremely low metastatic risk. Urine utDNA was more sensitive than plasma ctDNA at detecting residual disease within the bladder, and detectable urine utDNA in patients with a complete clinical response was associated with shorter bladder-intact survival (HR 6.47, 95% CI 1.34-31.31; log-rank p=0.008). These findings establish the conceptual and experimental foundation for incorporating ctDNA and utDNA assays into the management of patients with MIBC, particularly with respect to the need for cystectomy.

**Significance Statement:** Radical cystectomy—surgical removal of the bladder and creation of permanent urinary diversion—has historically been used to treat muscle-invasive bladder cancer, but is life-altering with permanent changes to urinary function, sexuality, and daily routine. We show here that systemic therapy, without cystectomy, can lead to durable remission in a subset of patients with muscle-invasive bladder cancer. Furthermore, we show that the measurement of circulating tumor DNA (ctDNA) and urine tumor DNA (utDNA) can identify patients who are most likely to remain cancer-free following this systemic therapy and bladder-preserving approach.

## Introduction

Bladder cancer is initially diagnosed through cystoscopic resection of the bladder tumor, known as a TURBT (Transurethral Resection of Bladder Tumor). For muscle-invasive bladder cancer (MIBC), this procedure serves primarily a diagnostic rather than therapeutic purpose. Radical cystectomy, surgical removal of the bladder, therefore remains the standard curative-intent treatment for MIBC. However, this procedure requires permanent urinary diversion, has detrimental effects on patients’ psychosocial health, and carries a peri-operative mortality risk of 2-10% (1–3). Notwithstanding these challenges, ∼50% of patients with MIBC treated with cystectomy alone develop metastatic recurrence (4, 5).

Given the risk of metastatic recurrence with cystectomy alone, systemic therapy administered prior to cystectomy with the goal of eradicating microscopic metastatic disease (also referred to as neoadjuvant therapy) has evolved as a standard treatment approach for MIBC (4, 5). Pathologic examination of the bladder after cystectomy reveals that 30-40% of patients with MIBC achieve a pathologic complete response (pCR) to systemic therapy, with no detectable cancer cells in the removed bladder (6, 7). This observation has raised the hypothesis that a subset of patients with MIBC may be cured without the need to remove their bladders (8–10). We previously reported the results of a phase II trial exploring a new treatment paradigm for MIBC, in which cystectomy was omitted in patients achieving a *clinical* complete response (cCR), i.e., these patients had no evidence of cancer detected by a series of examinations after systemic therapy, including a repeat TURBT (11). Herein, we report extended follow-up of the patients enrolled in this trial and explore the potential of personalized circulating tumor DNA (ctDNA) and urine tumor DNA (utDNA) assays to further inform the prognosis of these patients.

## Results

### Clinical Outcomes with Extended Follow-up

HCRN GU16-257 (NCT03558087) enrolled patients with cT2-4aN0M0 urothelial cancers of the bladder. Following TURBT, patients received four cycles of the pyrimidine analog gemcitabine, DNA cross-linking agent cisplatin, and the immune checkpoint inhibitor nivolumab. They then underwent clinical restaging, encompassing a second cystoscopy with biopsies, urine cytology, and imaging (Fig. 1A) (11). Those patients achieving a cCR could elect to forgo cystectomy and receive 8 additional cycles of nivolumab (cycles 5-12) followed by surveillance. Patients not achieving a cCR were advised to undergo cystectomy. Among 76 enrolled patients, 72 completed clinical restaging, and 33/76 (43%) achieved a cCR; only one patient with a cCR opted for immediate cystectomy. The median follow-up of patients achieving a cCR at the final data lock was 47.3 months (range, 18.4-59.9 months). On landmark analysis from the time of restaging, patients who did, versus did not, achieve a cCR demonstrated a 3-year metastasis-free survival (MFS) rate of 90% versus 64% (log-rank p=0.009) and 3-year overall survival rate of 97% versus 72% (log-rank p=0.002), respectively (Fig. 1B and C). Among patients achieving a cCR, the 3-year bladder intact survival rate was 69% (95% CI 55%, 87%; Fig. 1D).

**Fig. 1.**
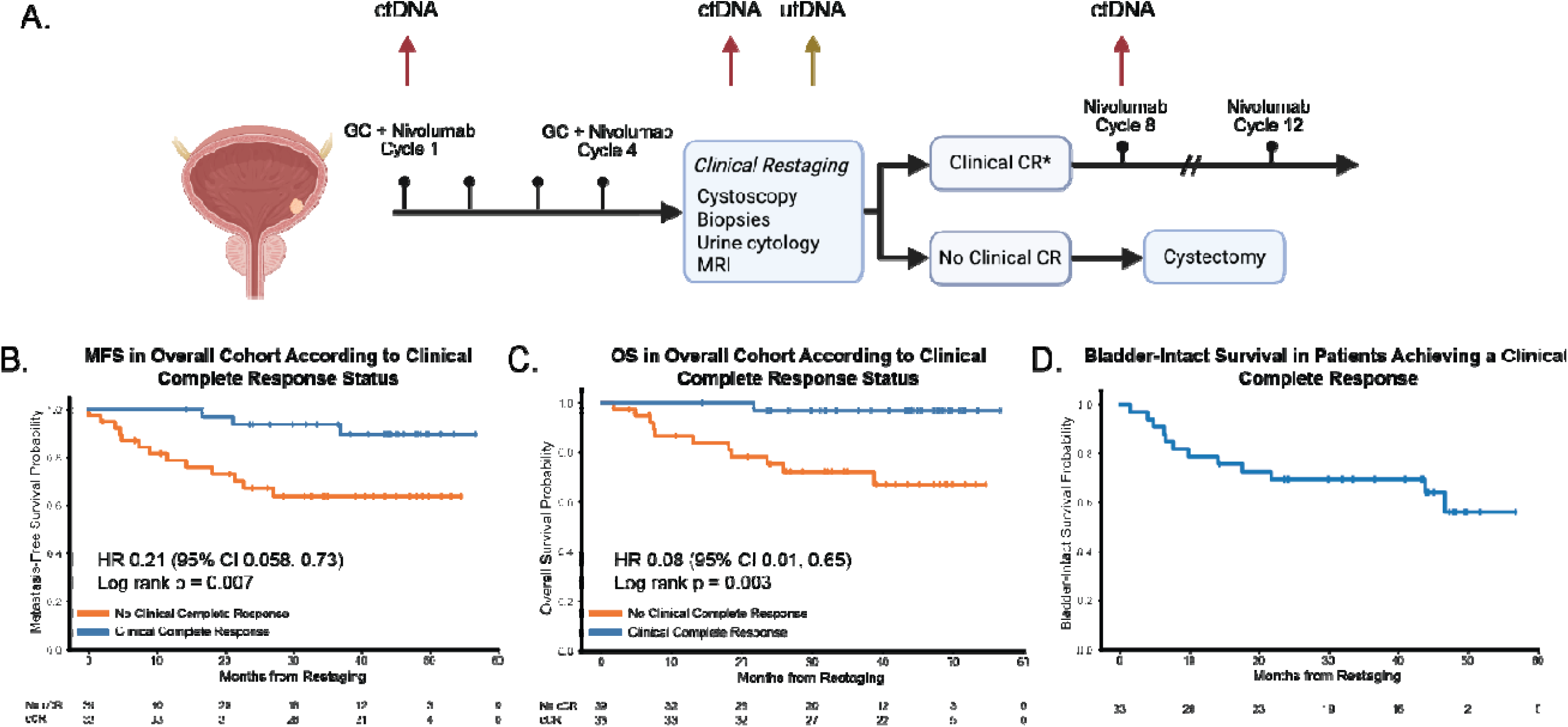
HCRN GU16-257 phase 2 trial extended follow-up. (A) HCRN GU16-257 clinical trial schema and timing of blood and urine collection for circulating tumor DNA (ctDNA) and urine tumor DNA (utDNA) testing. Cisplatin-eligible patients with muscle-invasive bladder cancer were treated with four cycles of gemcitabine, cisplatin, plus nivolumab (GC + Nivolumab). Clinical restaging was performed after cycle 4. Patients achieving a clinical complete response (cCR) could forgo cystectomy and receive eight additional doses of nivolumab followed by surveillance. Patients not achieving a cCR were advised to proceed with radical cystectomy. Plasma was collected for ctDNA testing on cycle 1 day 1, at the time of clinical restaging, and cycle 8 day 1. Urine was collected for utDNA testing at the time of clinical restaging. (B) Metastasis-free survival (MFS) according to cCR versus no cCR using post-cycle 4 restaging as the “landmark” time point (n□=□72; four patients were excluded who did not undergo clinical response assessment). (C) Overall survival (OS) according to cCR versus no cCR using the landmark timepoint (n□=□72; four patients were excluded who did not undergo clinical response assessment). (D) Bladder-intact survival among patients achieving a cCR (n=33 patients who achieved a cCR, including one patient who opted for immediate cystectomy). *Choice of cystectomy versus proceeding with additional systemic therapy was based on patient decision. Panel (A) was created with BioRender.

### Overview of ctDNA and utDNA Analysis

We hypothesized that tumor-informed ctDNA and utDNA, as measures of subclinical molecular residual disease (MRD), could help determine which of the patients described above were most likely to be cured without cystectomy (Figs. 2A, 3A, and *SI Appendix* S1) (12). We retrospectively applied a personalized, tumor-informed platform to characterize ctDNA and utDNA on all available samples from the HCRN GU16-257 study (*SI Appendix* Fig. S2). For each patient, up to 96 mutations (candidate mutations) were selected from whole-genome sequencing (WGS) data of DNA from the primary bladder tumor as well as from matched white blood cells (WBC). These candidate mutations were carefully selected based on the quality of the sequencing data and other criteria designed to minimize errors in subsequent sequencing analysis (see Materials and Methods). Note that virtually all chosen mutations were passengers rather than drivers. These passenger mutations were selected for diagnostic purposes because they were clonal with high mutant allele frequencies in the primary tumors, and were unlikely to be artifacts of library preparation or sequencing (such as a subset of transitions or other mutations with high technical backgrounds). The candidate mutations were then assessed in DNA from the primary tumor and WBC using a highly specific and sensitive assay termed “v96” (see Materials and Methods) (13, 14). The high sensitivity and specificity of v96 results from the careful selection of the mutations to be assessed, the relatively large number of tumor-specific mutations assessed, the high conversion efficiency of original DNA to library DNA, and the independent sequencing of both the Watson and Crick strands of the DNA molecules (13, 14). Candidate mutations were considered *bona fide* somatic mutations if the v96 assay confirmed their presence in primary tumor DNA at the frequency expected from the WGS data and absent from WBC DNA. These *bona fide* mutations were then tested in plasma and urine samples to detect ctDNA and utDNA, respectively. We employed a locked-in assay with a prespecified definition for “detectable” and with the assay team blinded to clinical outcomes (see Materials and Methods). Using our prespecified definition, any sample containing more than one mutant molecule detected by the v96 assay was scored as ‘detectable’ (aka, ‘positive’).”

**Fig. 2.**
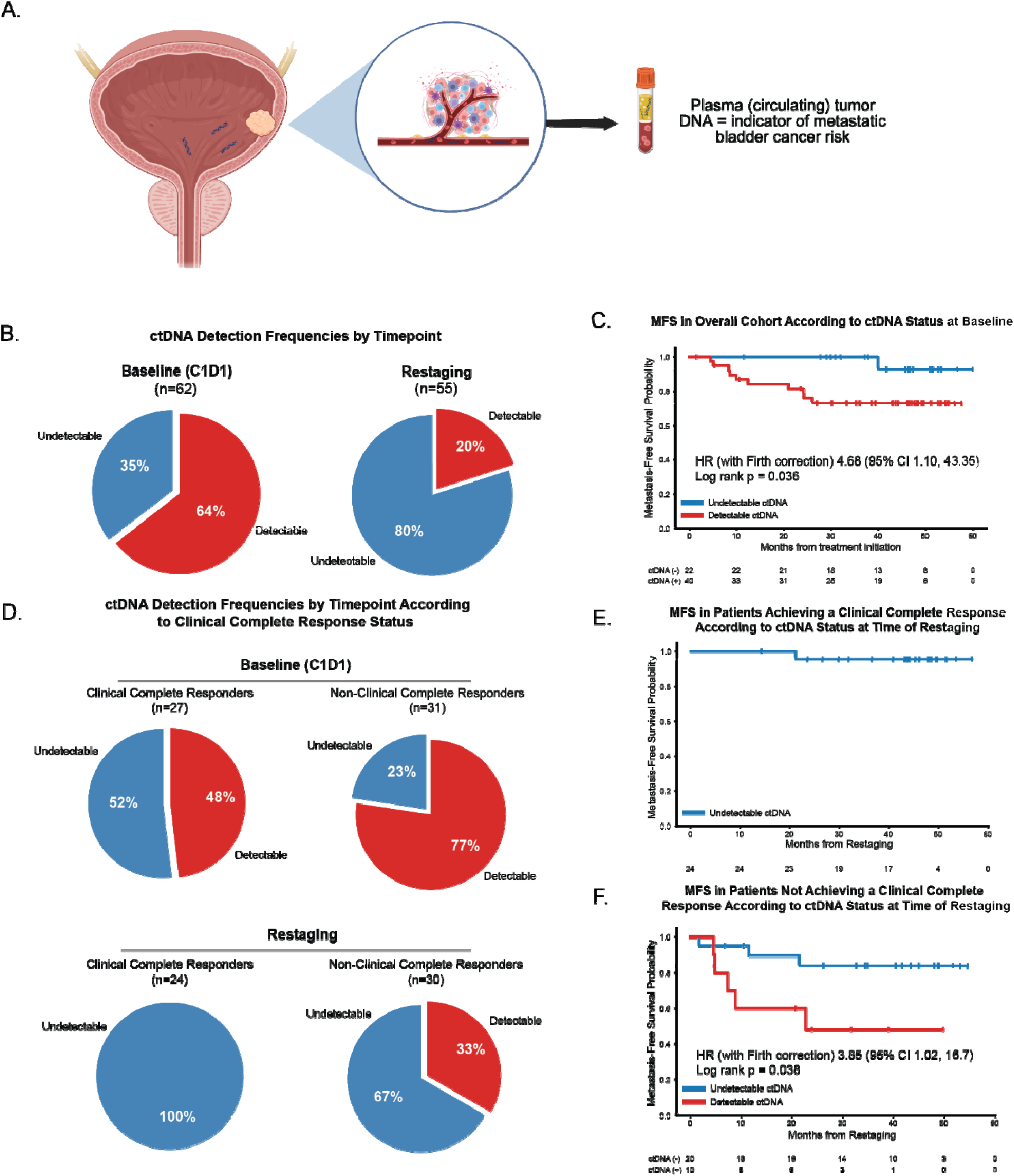
Association between circulating tumor DNA (ctDNA) results and outcomes among patients enrolled on HCRN GU16-257. (A) Illustration highlighting hypothesis that ctDNA may serve as a measure of subclinical metastatic cancer bladder cancer. (B) ctDNA detection frequencies according to testing timepoint (sample sizes at each timepoint limited to patients with available biospecimens at those timepoints). (C) Metastasis-free survival according to ctDNA (detectable versus undetectable) on cycle 1 day 1 (n=62; includes all patients with evaluable cycle 1 day 1 ctDNA data). (D) ctDNA detection frequencies at cycle 1 day 1 and restaging according to cCR status (sample sizes at each timepoint limited to patients with available biospecimens; 4 patients excluded from the cycle 1 day 1 analysis and 1 patient excluded from the restaging analysis who did not undergo a clinical restaging assessment). (E) Metastasis-free survival (landmark analysis based on time of restaging) among patients achieving a cCR, all of whom had undetectable ctDNA at the time of restaging (sample size limited to patients with available biospecimens). (F) Metastasis-free survival (landmark analysis based on time of restaging) among patients not achieving a cCR according to ctDNA status at the time of restaging (sample size limited to patients with available biospecimens). One event in a patient with undetectable ctDNA was a death related to post-cystectomy sepsis. Panel (A) was created with BioRender.

**Fig. 3.**
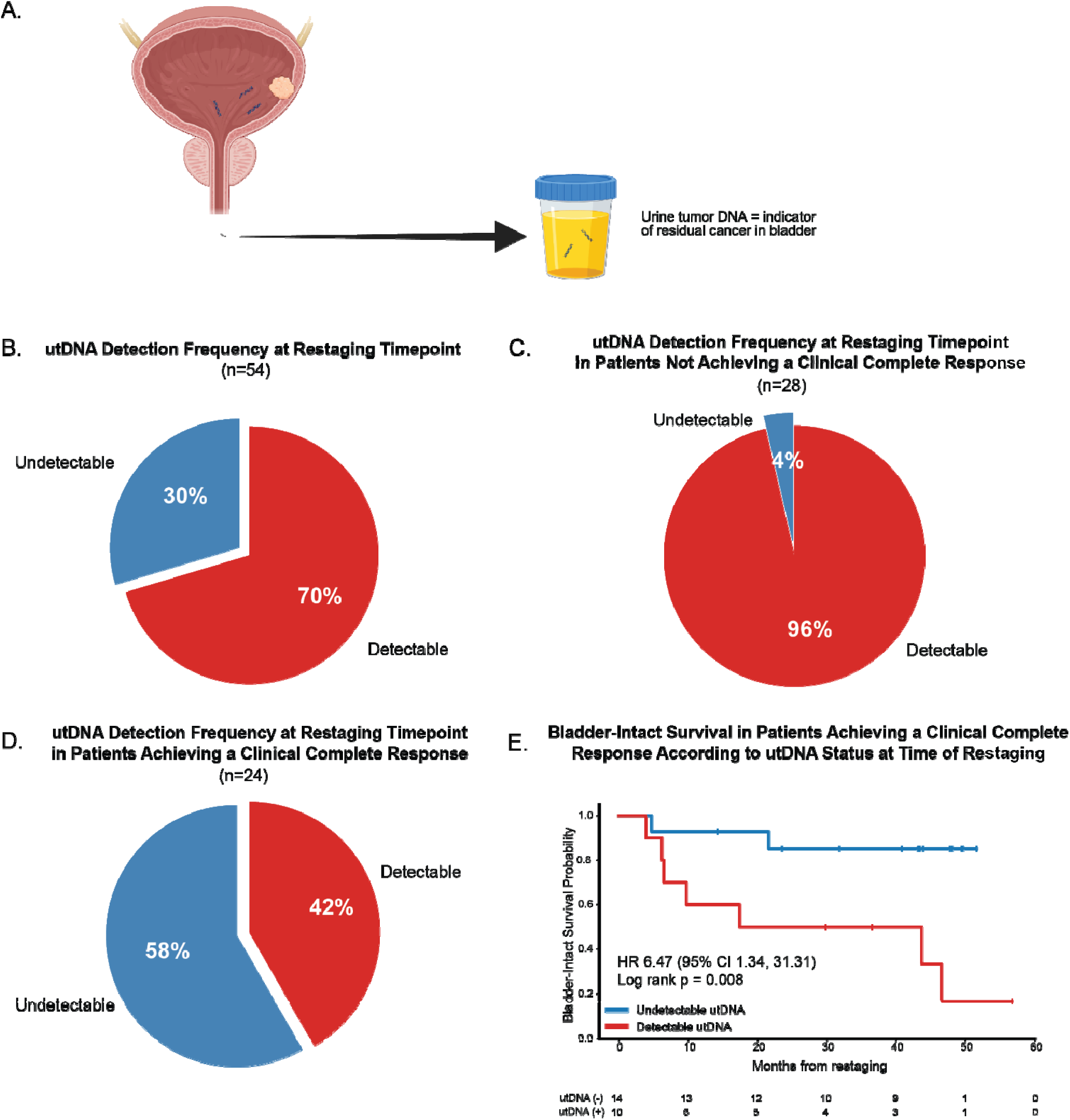
Association between urine tumor DNA (utDNA) results and outcomes among patients enrolled on HCRN GU16-257. (A) Illustration highlighting hypothesis that utDNA may serve as a measure of subclinical local bladder cancer, even in patients with a clinical complete response to systemic therapy (i.e., no evidence of cancer on restaging biopsies and urine cytology). (B) utDNA detection frequency at the restaging timepoint in the overall study population(n=54, sample size limited to patients with available biospecimens). (C) utDNA detection frequency at the restaging timepoint in patients not achieving a clinical complete response (n=28, sample size limited to patients with available biospecimens and clinical restaging data). (D) utDNA detection frequency at the restaging timepoint in patients achieving a clinical complete response (n=24, sample size limited to patients with available biospecimens and clinical restaging data). (E) Bladder-intact survival (landmark analysis at time of restaging) among patients achieving a clinical complete response according to utDNA at the restaging timepoint (n=24, sample size limited to patients with available biospecimens). Panel (A) was created with BioRender. Two patients were omitted from the analyses of utDNA versus clinical complete response because they did not undergo clinical restaging assessment.

Among 76 patients enrolled in the trial, tumor-informed panels tracking a median of 93 *bona fide* somatic mutations per patient (range, 24-96) could be generated for 65 patients (*SI Appendix* Fig. S2 and Tables S1 and S2). For the other eleven patients, sufficient tumor for DNA analysis was unavailable. We assessed plasma ctDNA at three timepoints (Fig. 1A): Baseline (day 1 of the first cycle of systemic therapy, also called C1D1); Restaging (after four cycles of systemic therapy) and for patients achieving a cCR, after four additional cycles of maintenance therapy (day 1 of Cycle 8). Urine tumor DNA was assessed only at the restaging timepoint because that was the timepoint at which a decision for cystectomy had first to be made and because that was the only time point at which the clinical protocol enabled urine collection (Fig. 1A).

### Association of ctDNA with Clinical Outcomes

The ctDNA detection frequencies at baseline and restaging are shown in Fig. 2B: ctDNA was detectable in 64% of patients at baseline but in only 20% of patients after four cycles of systemic therapy (p<0.0001, 2-tailed Fisher’s Exact test). No patient demonstrated conversion of ctDNA from undetectable to detectable from baseline to restaging (*SI Appendix*, Fig. S2B and Tables S1 and S2). All patients who could be examined on cycle 8 day 1 had undetectable ctDNA (*SI Appendix,* Table S2).

We next explored the prognostic value of baseline ctDNA in the cohort (n=62 patients available for analysis; see Materials and Methods). Patients with undetectable versus detectable ctDNA at baseline were at exceptionally low risk of developing metastatic recurrence. Only one patient in the cohort with baseline undetectable ctDNA experienced metastatic recurrence (Metastasis-free survival (MFS) for detectable versus undetectable ctDNA (HR 4.68; 95% CI 1.10-43.35; log-rank p=0.036; Fig. 2C). Notably, patients with undetectable versus detectable baseline ctDNA also had a higher likelihood of achieving a cCR (Fig. 2D; p=0.029, 2-tailed Fisher’s Exact test).

To assess whether ctDNA testing had the potential to guide treatment decisions, we analyzed ctDNA status at the restaging timepoint in patients with (n=24) and without (n=30) a cCR. No patients achieving a cCR had detectable ctDNA at the time of restaging (Fig. 2D) and only one patient in this subgroup experienced metastatic recurrence (Fig. 2E). Among patients not achieving a cCR, 67% had undetectable ctDNA at restaging (Fig. 2D). Despite not achieving a cCR, ctDNA status at restaging (prior to cystectomy) was strongly associated with MFS. Patients with detectable ctDNA had significantly shorter MFS compared to those with undetectable ctDNA (HR, 3.85; 95% CI 1.02, 16.7; log-rank p=0.038; Fig. 2F).

### Association of utDNA with Clinical Outcomes

Urine tumor DNA was assessable at restaging in 54 patients (*SI Appendix,* Fig. S2 and Table S2). In contrast to ctDNA, utDNA was detectable in most patients (70%) at the time of restaging, illustrating its higher sensitivity for residual disease within the bladder (p=0.0242 by 2-tailed Fisher’s Exact test, Fig. 3B). However, there was a difference in utDNA detectability between patients who did versus did not achieve a cCR (40% and 96%, respectively; p<0.0001 by 2-tailed Fisher’s Exact test, Fig. 3C and D). Patients achieving a cCR with detectable versus undetectable utDNA at restaging demonstrated inferior bladder-intact survival (HR 6.47, 95% CI 1.34-31.31; log-rank p=0.008; Fig. 3E). Among patients with a cCR, 10 had detectable utDNA at the time of restaging and 7/10 have subsequently undergone cystectomy. Importantly, by definition, at the time of restaging (the same timepoint at which urine was tested for utDNA) all patients with a cCR were cancer-negative on urine cytology and had no evidence of malignancy on histopathologic examination of their bladder biopsies.

The outcomes of the 33 patients achieving a cCR, annotated with ctDNA and utDNA status, are detailed in Figure 4. Among patients with a cCR, high-grade recurrences in the bladder occurred in 70% (7/10) with detectable utDNA at restaging and 29% (4/14) with undetectable utDNA at restaging. The median time from detectable utDNA at the restaging timepoint to high-grade recurrence was 12.9 months (range, 5.6-55.1 months). Thirteen patients with a cCR experienced local recurrence and were treated as follows: 9 underwent radical cystectomy, 1 underwent partial cystectomy, 1 received intravesical therapy for low-grade recurrence, and 2 patients with non-invasive papillary recurrences opted for continued surveillance after repeat transurethral resection.

**Fig. 4.**
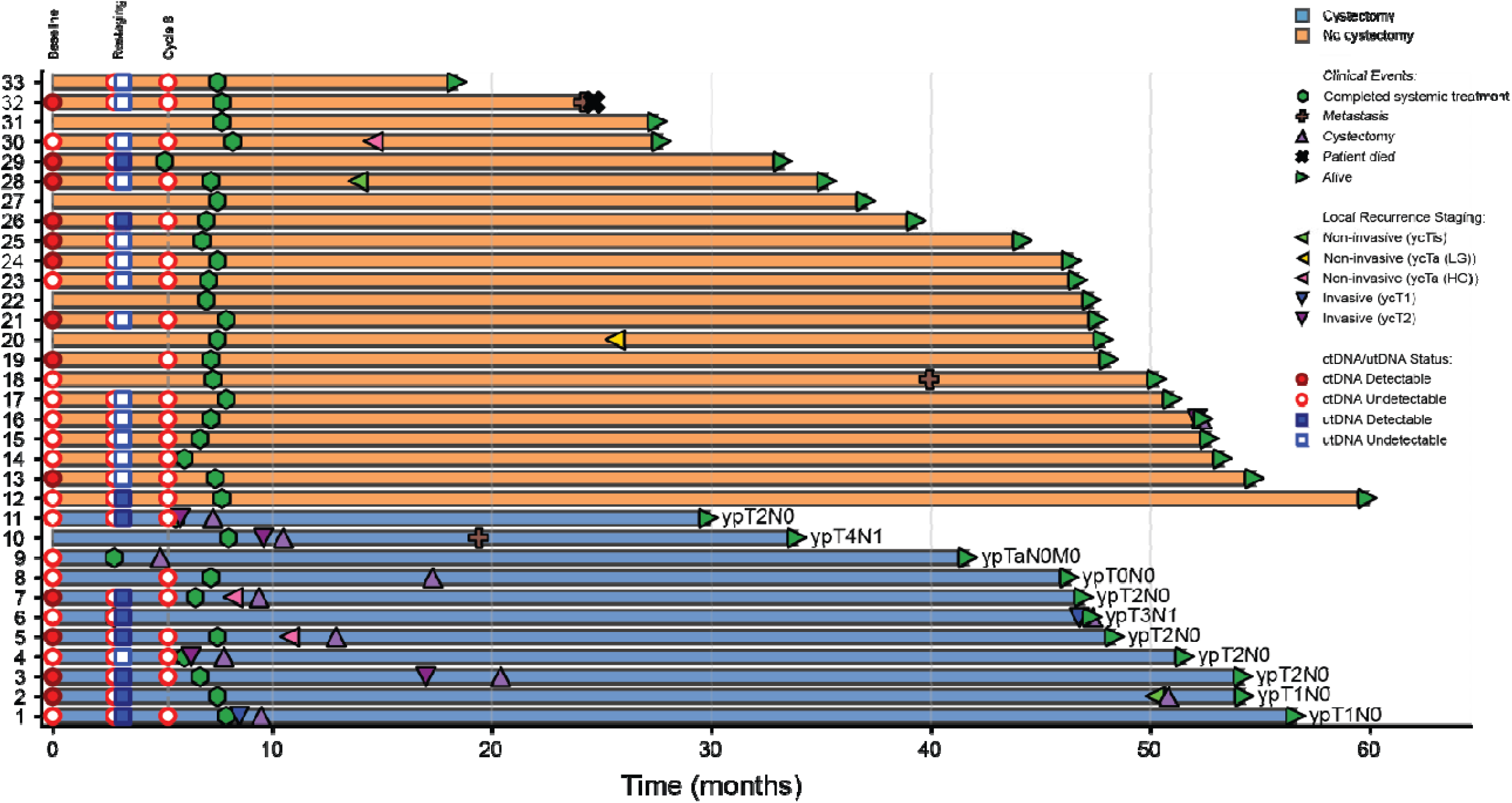
Swimmers lane plot detailing outcomes of patients achieving a clinical complete response annotated with circulating tumor DNA (ctDNA) and urine tumor DNA (utDNA) and clinical event data. (n=33 patients achieving a clinical complete response). Final pathologic stage indicated at the ends of the lanes in patients who underwent cystectomy. Patient 8 underwent cystectomy for suspicious MRI findings but had a pathological complete response. Patient 9 opted for immediate cystectomy. Patient 16 developed an invasive recurrence but underwent partial cystectomy.

## Discussion

The results described above demonstrate that a subset of patients with MIBC achieving a stringently defined cCR can achieve durable bladder-intact survival with systemic chemotherapy plus immune checkpoint blockade following a diagnostic resection of bladder tumor through cystoscopy. Since publication of our initial results, additional evidence has bolstered these findings (4, 15, 16). Advancing this treatment paradigm requires further investigation. In particular, whether forgoing immediate cystectomy in patients with a cCR is non-inferior to immediate cystectomy for all patients requires a randomized study. The feasibility of such a randomization is unknown and is being evaluated in the NEO-BLAST trial (NCT06537154).

Tools to further refine the identification of patients who could most safely forgo immediate cystectomy are, therefore, highly desirable. We here evaluated a personalized approach to MRD detection in plasma and urine, employing highly sensitive and specific sequencing assays to identify subclinical disease. To our knowledge, this study is the first to use a highly sensitive sequencing-based technology to retrospectively explore the relationship between such MRD assays and clinical outcomes in the setting of a response-guided bladder-sparing paradigm in MIBC. We identified five key findings that, if further validated, could inform future response-guided bladder-sparing treatment strategies (*SI Appendix* Fig. S1):

ctDNA Finding 1: In the context of this trial with gemcitabine, cisplatin, and nivolumab, patients with undetectable ctDNA using the v96 assay demonstrated an extremely low risk of developing metastatic disease. In particular, only 1/22 patients with undetectable baseline ctDNA developed metastatic disease) despite a large subset of patients not undergoing conventional definitive local therapy (cystectomy or radiation). These findings underline the safety of surveillance in patients achieving a cCR using the protocol described herein. Outcomes could of course differ if other systemic regimens are used for treatment or less sensitive assays are used to detect ctDNA.

ctDNA Finding 2: Patients with detectable ctDNA after systemic therapy were highly unlikely to achieve a cCR. These findings align with the extremely low likelihood of a *pCR* at cystectomy in patients with detectable ctDNA after systemic therapy (16) and such patients could potentially avoid invasive cystoscopic biopsies as a component of clinical restaging in future response-guided bladder-sparing studies.

ctDNA Finding 3: Despite pathologic evidence of residual cancer in the bladder after TURBT plus systemic therapy in patients not achieving a cCR, undetectable versus detectable ctDNA at the time of restaging was still associated with improved MFS. This finding reinforces the high curative potential of local therapy in this setting. Conversely, patients without a cCR who had detectable ctDNA after initial systemic therapy might benefit from alternative systemic therapy rather than proceeding directly to cystectomy.

utDNA Finding 1: utDNA was detected in 96% of patients not achieving a cCR. Furthermore, patients with detectable utDNA in the setting of a cCR—despite having no evidence of cancer on cystoscopic biopsies or urine cytology—demonstrated significantly inferior bladder-intact survival (HR 6.47, p=0.008). These findings strongly support a role of utDNA for identifying subclinical residual disease, as such patients may benefit from early integration of local therapy. Whether utDNA could complement or potentially replace invasive cystoscopic surveillance to define the need for local intervention, and whether different local therapy approaches (cystectomy versus radiation versus novel intravesical therapies) are indicated based on the combination of cCR status and utDNA results, warrant prospective investigation.

utDNA Finding 2: The role of utDNA in guiding the optimal duration of maintenance systemic therapy in patients achieving a cCR remains to be determined. Three patients achieving a cCR with detectable utDNA at restaging had no evidence of local or distant recurrence at 34+ months follow-up. All patients achieving a cCR in HCRN GU16-257 received maintenance nivolumab after the restaging utDNA timepoint, raising the possibility that either (a) these detectable findings at the time of restaging were false positive values or (b) the additional cycles of immune checkpoint blockade deepened the response to treatment and “cleared” utDNA. Serial utDNA analysis before, during, and after maintenance systemic therapy could be incorporated into future prospective studies to distinguish between these possibilities.

There are potential limitations to our analysis. We reported bladder-intact overall survival (BIOS) rather than bladder-intact event-free survival (BIEFS) as BIOS was prespecified as an endpoint when our study was designed. Bladder-intact overall survival differs from BIEFS in that muscle-invasive and metastatic recurrences are not considered events for BIOS. However, these endpoints were nearly synonymous for the purposes of our analyses as all patients with muscle-invasive recurrence underwent cystectomy and only one patient with a cCR and metastatic recurrence was alive with an intact bladder by the end of the trial. Our ctDNA and utDNA analyses are exploratory and were performed retrospectively using samples collected in the HCRN GU16-257 trial. Despite being among the first and largest prospective trials testing response-guided bladder-sparing treatment in MIBC, our sample size for ctDNA and utDNA analyses was relatively small. Our analyses employed a highly sensitive and specific tumor-informed ctDNA and utDNA assay performed in an academic laboratory. These assays are not yet available for routine use. However, several companies are now developing tumor-informed ctDNA assays. The reduced cost of sequencing and further technical improvements, combined with clinical trials and regulatory approvals, will hopefully enable such testing to be routinely performed (17–19). Urine was only banked at the restaging timepoint in HCRN GU16-257. Whether utDNA in patients achieving a cCR can similarly detect other types of cancer recurrences in the bladder (across different grades, stages, and timing of recurrences), and whether uniform treatment is required for all types of local recurrences, remains unknown. Serial utDNA testing may further improve performance in detecting subclinical cancer in the bladder. Several aspects of utDNA analysis, including the optimal sample type, remain poorly standardized. Based on our data and others, purification of DNA from both urine supernatant and cell pellets may improve performance (20, 21). The therapeutic landscape has evolved since HCRN GU16-257, most notably with the combination of enfortumab vedotin plus pembrolizumab (22). This regimen is now being tested within a response-guided bladder-sparing approach in HCRN GU22-598 (NCT06809140).

This extended follow-up confirms that a uniformly assessed and stringently defined cCR identifies a subset of patients with MIBC achieving durable bladder-intact survival after TURBT plus systemic therapy. The integration of tumor-informed ctDNA and utDNA assays yields complementary molecular insights based on the risk for subclinical metastatic and local residual disease, respectively. Despite the need for further investigation, our results establish a framework for incorporating MRD detection into response-guided bladder-sparing approaches in MIBC (*SI Appendix*, Fig. S1).

## Materials and Methods

HCRN GU 16-257 was a phase 2 investigator-initiated multicenter clinical trial. Cisplatin-eligible patients with muscle-invasive urothelial cancer of the bladder (cT2-T4aN0M0) enrolled at seven medical centers received treatment with gemcitabine, cisplatin, plus nivolumab following TURBT (Fig. 1A). Clinical restaging was performed after cycle 4. Clinical restaging included magnetic resonance imaging of the abdomen and pelvis [or computed tomography (CT) if MRI was contraindicated], CT of the chest, rigid cystoscopy with biopsies, and urine cytology. Transurethral resection of any visible tumor and/or the prior tumor site was performed. In addition, biopsies were obtained from the following sites in the bladder: trigone, left, right, anterior, posterior, dome. In men, prostatic urethral biopsies were performed. A clinical complete response (cCR) was defined as meeting all of the following: (a) no evidence of malignancy on biopsy with the exception of low grade papillary (Ta) tumors, (b) no malignant cells on urine cytology, (c) no evidence of local or metastatic disease on cross-sectional imaging. Residual bladder wall changes on cross-sectional imaging were interpreted by the treating investigator in consultation with the local radiologist and in the context of the bladder biopsy results. Patients achieving a cCR were offered the option to proceed with radical cystectomy versus retain their bladder and receive 8 additional doses of nivolumab followed by surveillance. Patients not achieving a cCR were advised to proceed with radical cystectomy. The surveillance schedule is outlined in the protocol (9). The study was conducted in accordance with the Declaration of Helsinki. The protocol was approved by local ethics committees at the Icahn School of Medicine at Mount Sinai, City of Hope Comprehensive Cancer Centers, Huntsman Cancer Institute University of Utah, Oregon Health and Science University, Penn Medicine Abramson Cancer Center, Rutgers Cancer Institute of New Jersey, University of Southern California, and University of Wisconsin and written informed consent was provided by all patients prior to enrollment. The trial was registered at clinicaltrials.gov (NCT03558087). The trial design, inclusion and exclusion criteria, and initial clinical results have been previously described (11).

### Biospecimen Collection, Processing, and Banking

Archival tumor tissue was collected from formalin-fixed paraffin-embedded (FFPE) diagnostic MIBC specimens (unstained 4-micrometer sections on positively charged slides with at least 20% tumor content). Slides were freshly cut, unbaked, and shipped to the Hoosier Cancer Research Network Biorepository. Baseline samples for tumors were obtained prior to chemotherapy in all but 11 patients; in these 11 patients, baseline archival tumor tissue had been exhausted and on-treatment specimens were used to identify tumor-specific mutations (see below). A hematoxylin and eosin-stained slide was evaluated for tumor content.

Blood samples for circulating tumor DNA (ctDNA) analysis were collected at predefined timepoints including pre-dose cycle 1 day 1, time of restaging (post-cycle 4), and cycle 8 day 1. Blood was collected into 10 mL lavender-top EDTA tubes and processed within 30 minutes of draw by centrifugation at ≤3000 RCF for 10 minutes. Plasma was separated from leukocytes (WBCs) and transferred to −80°C storage. The plasma was equally divided into nine labeled 2 mL DNase-free cryovials, then immediately transferred to −80°C storage. Though plasma samples were always obtained at the baseline timepoint (C1D1, before systemic chemotherapy), WBCs for germline sequencing were obtained from baseline in all but 8 patients; in these 8 patients, baseline WBCs were unavailable and on-treatment samples were used to identify tumor-specific mutations (i.e., those present in the tumor DNA but no the WBC DNA, as explained below).

Urine samples for tumor DNA (utDNA) analysis were collected at the time of restaging (post-cycle 4) into provided specimen cups and transferred to 50 mL conical tubes. Urine was centrifuged at 450 rpm for 15 minutes at room temperature, after which the supernatant was aliquoted into four 15 mL tubes (≤10 mL per tube) and the remaining cell pellet was resuspended in 0.5-1 mL of residual urine, transferred to a 2 mL cryovial with 10% DMSO final concentration, and frozen at −80°C. All biospecimens were shipped overnight on dry ice to the Hoosier Cancer Research Network biorepository for banking and frozen at −80°C for subsequent analysis. In *SI Appendix* Table S2, samples marked “urine” indicate that the “whole” urine sample (supernatant plus pellet) was used for DNA purification.

### Circulating tumor DNA and urine tumor DNA analysis

For ctDNA, 6 mL of plasma were purified using BioChain cfDNA Extraction Kit (BioChain, cat #K5011610) using the manufacturer’s recommended protocol as previously described (23). Leukocyte DNA was purified from whole blood and sheared to an average size of ∼200bp using a Hielscher UIP400MTP sonicator prior to library preparation as previously described (23). DNA was purified from macrodissected tumors as previously described (24). For urine DNA, cell pellets, urine supernatants, or 1.5-15 mL (median 8 mL) whole urine samples were purified as previously described (25). cfDNA and utDNA were quantified using an Agilent TapeStation instrument. SaferSeqS libraries were made from plasma cfDNA, or sheared DNA from urine, tumors, and leukocytes as previously described (13) with the following modifications. KAPA HiFi HotStart ReadyMix (Roche) was used to amplify the DNA following ligations using the following conditions: 98 °C for 45 s, followed by 8 cycles of 98 °C for 15 s, 60 °C for 30 s and 72 °C for 30 seconds. The amplified DNA was cleaned up using 1.8 X SPRI beads and eluted in 100 uL EB buffer (Qiagen).

Whole genome sequencing (WGS) was performed on an Illumina NovaSeq 6000 instrument or a Complete Genomics T7 instrument to a depth of ∼30X for primary tumors and matched normal leukocytes as previously described (14, 23). Briefly, FASTQ files were generated using Illumina’s bcl2fastq or by Complete Genomic’s Ztron Lite Server. Adapter sequences were removed with Cutadapt (https://cutadapt.readthedocs.io/en/stable/). The trimmed sequences were then aligned to hg38 reference genome with BWA-MEM with default settings (https://github.com/lh3/bwa). Duplicate sequencing clusters were removed with Picard (http://broadinstitute.github.io/picard). Variants in the plasma sample were called using Mutect2 (https://gatk.broadinstitute.org/hc/en-us/articles/360037593851-Mutect2) using the same patient’s leukocytes as the matched normal (26). Any mutation present in the matched leukocytes were excluded from further analysis. Up to 96 candidate, tumor-specific mutations per patient were selected after excluding mutations in repetitive regions, regions with difficult alignments to the reference genome (hg38), regions that were difficult to amplify efficiently, regions containing single nucleotide polymorphisms, or transitions at CpG sites. These exclusions were informed through prior analysis of samples from individuals without cancer, using whole genome or targeted sequencing. The candidate mutations described above were then used to design a personalized assay (“personalized mutation panel”; *SI Appendix* Table S1). For each of the candidate mutations chosen, primers were designed as described previously (13). Primers for all candidate mutations were combined into a single tube for each patient. A hemi-nested, two-stage PCR protocol was used to amplify the regions containing the candidate mutations as described previously, except that KAPA HiFi HotStart polymerase was used for amplification ReadyMix (Roche, Indianapolis, IN; cat # KR0370) (13). Following sequencing on a Complete Genomics T7 instrument, the data were evaluated using SaferSeqS as described (with source code available at https://zenodo.org/records/4588264) (13). ctDNA and utDNA analysis were performed by investigators who were blinded to the clinical outcome. Any sample in which more than one mutant molecule was detected by this assay was scored as “detectable” (aka, “positive”). When there was more than one urine sample type (e.g., pellet and supernatant) available for a patient at restaging, we scored it as “detectable” if any one of the samples contained more than one mutant molecule. The results for each sample are recorded in *SI Appendix* Table S2. The sequencing data generated during the study is deposited in the database of EU Genome–Phenome Archive under accession code EGAS00001008304.

### Statistical Analysis

The clinical endpoints included in this analysis are specified in the protocol though this is a post-hoc analysis utilizing extended follow-up data. The ctDNA and utDNA analyses are post-hoc and exploratory though a plan was outlined prior to the analysis (https://www.protocols.io/view/protocol-for-retrospective-analysis-of-circulating-j8nlkyeqdg5r/v1). Rates were calculated using percentages and compared among different groups using Fisher’s exact test. The Kaplan–Meier method was used to estimate metastasis-free, bladder-intact, and overall survival. Comparisons of time-to-event distributions between groups were made with the log-rank tests. Univariable Cox proportional hazard regression models were used to estimate the hazard ratios and corresponding 95% CIs for metastasis-free, bladder-intact, and overall survival. When comparing time-to-event outcomes based on variables from the restaging timepoint (i.e., cCR status, ctDNA at restaging, utDNA at restaging), landmark analyses were conducted using the restaging timepoints as the landmark time (i.e., time 0). Given the low number of metastasis-free survival events in the undetectable ctDNA subgroups, the Firth correction (27, 28) was applied to the univariate Cox model to ensure a more robust estimation of the hazard ratios. Due to the exploratory nature of our analysis, P values less than 0.05 were deemed statistically significant without further multiplicity adjustments. All statistical analyses were performed using SAS software version 9.4 (SAS Institute) and RStudio version 4.5.0 (R Core Team).

## Data Availability

All data produced in the present study are available upon reasonable request to the authors.

## Author contributions

Conception and design: MDG, BV, YW; Financial Support: MDG; Administrative support: MDG; Provision of study materials: MDG, SD, KGC, TBD, JPC, BO, AD, RM, CK, RM, SG, JS, SKP; Sample processing and sequencing: MP, JP, LD, NS. Collection and assembly of data: MDG, MY, SI, BV, YW; Data analysis and interpretation: MDG, MY, SD, CD, SI, BV, YW; Manuscript writing: MDG, MY, BV, YW; Final approval of manuscript: all authors; Accountable for all aspects of work: MDG.

## Acknowledgements

The authors would also like to thank Cherie Blair, Kathy Judge, Deborah Cummings-Thomas, and Matthew Smith for technical assistance, as well as the patients in the study for their courage and generosity. The trial was funded by Bristol Myers-Squibb. This study was supported by National Institutes of Health grant R21NS113016 (CB), National Institutes of Health grant RA37CA230400 (CB), National Institutes of Health grant U01CA230691 (NP, CB), Oncology Core CA 06973 (BV, KWK, NP), The Virginia and D.K. Ludwig Fund for Cancer Research (BV, KWK, NP, CB), The Commonwealth Fund (NP), Thomas M Hohman Memorial Cancer Research Fund (CB), The Sol Goldman Sequencing Facility at Johns Hopkins (BV), The Benjamin Baker Endowment (YW, CD), Swim Across America (CD), Burroughs Wellcome Career Award for Medical Scientists (CB), The V Foundation for Cancer Research (YW), P30 CA196521 (MDG), and Department of Defense Award No. HT9425-25-1-0489 (MDG). This work utilized the services of the Tisch Cancer Institute Biorepository and Pathology CoRE at the Icahn School of Medicine at Mount Sinai.

