## Supplemental Tables 1 and 2, Supplemental Figures 1 and 2 for "Monitoring of plasma and urine tumor-derived DNA to inform bladder-sparing approaches for patients with muscle-invasive bladder cancer"

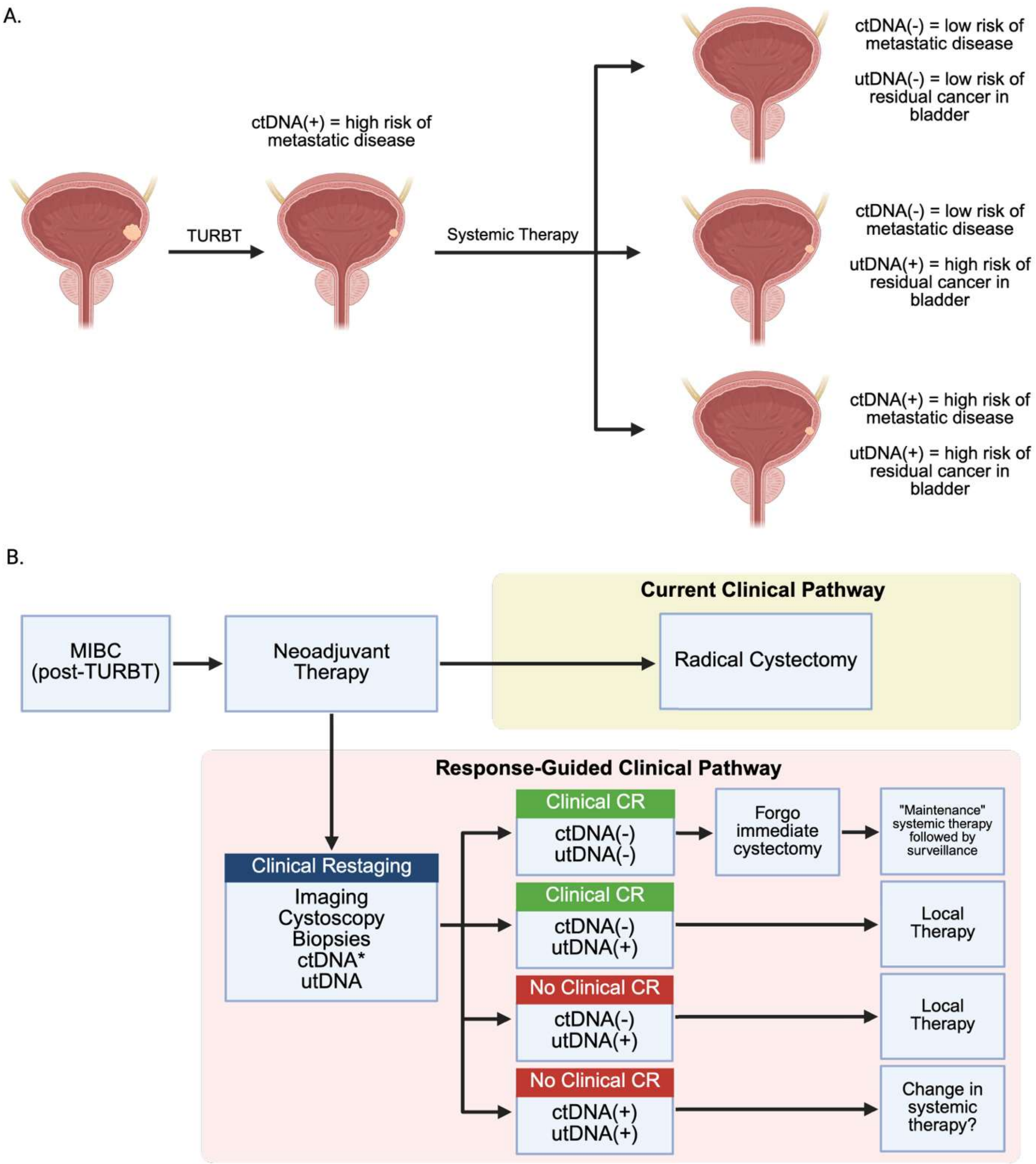


**Fig. S1. Hypothesis regarding the role of circulating tumor DNA and urine tumor DNA in a response-guided bladder-sparing approach to muscle-invasive bladder cancer.** (B) Hypothesis related to complimentary information derived from ctDNA and utDNA testing in the setting of systemic therapy for MIBC. (B) Potential future role of ctDNA and utDNA testing in the setting of a response-guided bladder-sparing approach versus current standard clinical practice. ctDNA, circulating tumor DNA; ctDNA(+), detectable ctDNA; ctDNA(-), undetectable ctDNA; utDNA, urine tumor DNA; utDNA(+), detectable utDNA; utDNA(-), undetectable utDNA; MIBC, muscle-invasive bladder cancer; Clinical CR, clinical complete response. *Patients with ctDNA(+) testing at the clinical restaging timepoint could potentially forgo restaging cystoscopy and biopsies. Figure was created with BioRender.

A.

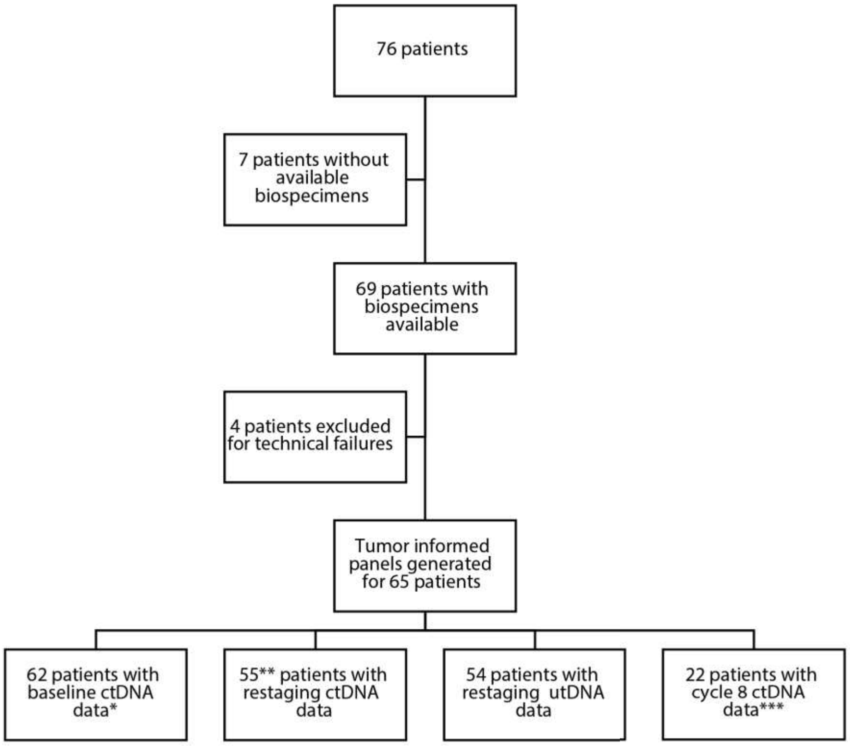


B.

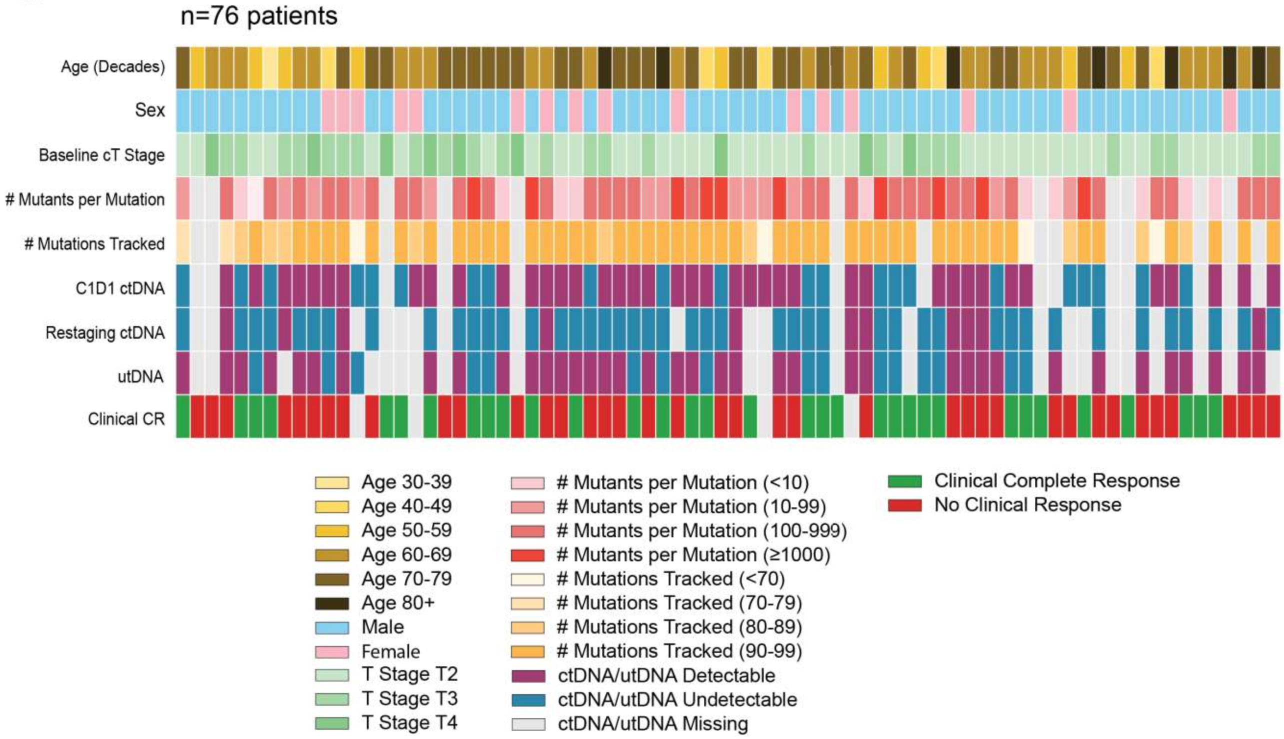


**Fig. S2. Data included in the long-term follow-up of HCRN GU16-257 and ctDNA and utDNA analyses.** (A) CONSORT diagram detailing inclusion and exclusion of patients enrolled on HCRN GU16-257 for circulating tumor DNA and urine tumor DNA analyses. (B) Heatmap detailing patient, tumor, ctDNA, and utDNA characteristics in patients enrolled on HCRN GU16-257 (n=76) and outlining missing data.

*Excludes 3 patients without cycle 1 day 1 plasma available but with plasma available at later timepoints

**51 patients had matched cycle 1 day 1 and restaging plasma ctDNA values

***Among 32 patients who achieved a clinical CR and proceeded without immediate cystectomy and continuing “maintenance” nivolumab.

**Table S1: Mutations included in personalized panels for each patient (genomic coordinates refer to hg38).**

| Patient | Hugo Symbol | Chromosome | Position | Variant Classification | Variant Type | From | To |
|---|---|---|---|---|---|---|---|
| INDIA 8544 | ENST00000413056 | chr3 | 187933255 | Intron | SBS | G | C |
| INDIA 8544 | Unknown | chr3 | 55200331 | IGR | SBS | G | C |
| INDIA 8544 | ENST00000652126 | chr14 | 39597627 | Intron | SBS | C | G |
| INDIA 8544 | CD99L2 | chrX | 150769092 | Missense_Mutation | SBS | G | C |
| INDIA 8544 | MAP2 | chr2 | 209472470 | Intron | SBS | G | C |
| INDIA 8544 | PCNX2 | chr1 | 233143250 | Intron | SBS | C | G |
| INDIA 8544 | LINC00111 | chr21 | 41678120 | 5'Flank | SBS | G | C |
| INDIA 8544 | Unknown | chr17 | 77789487 | IGR | SBS | G | C |
| INDIA 8544 | SNCAIP | chr5 | 122401946 | Intron | SBS | C | G |
| INDIA 8544 | ABCF2 | chr7 | 151221041 | Intron | SBS | C | G |
| INDIA 8544 | Unknown | chr8 | 61954138 | IGR | SBS | C | G |
| INDIA 8544 | CAMK1D | chr10 | 12813243 | Intron | DEL | T | - |
| INDIA 8544 | BMP7 | chr20 | 57200018 | Intron | SBS | C | G |
| INDIA 8544 | Unknown | chrX | 35247229 | IGR | SBS | G | C |
| INDIA 8544 | Unknown | chr5 | 105429468 | IGR | SBS | G | C |
| INDIA 8544 | FTO | chr16 | 53895762 | Intron | SBS | G | C |
| INDIA 8544 | Unknown | chr7 | 111691666 | IGR | SBS | G | C |
| INDIA 8544 | LINC02133 | chr16 | 47830087 | Intron | SBS | G | T |
| INDIA 8544 | SKAP1 | chr17 | 48256669 | Intron | SBS | C | G |
| INDIA 8544 | DSCAM | chr21 | 40258247 | Intron | SBS | C | G |
| INDIA 8544 | Unknown | chr13 | 112173078 | IGR | SBS | G | C |
| INDIA 8544 | SYN3 | chr22 | 32651649 | Intron | SBS | G | C |
| INDIA 8544 | ENST00000669498 | chr6 | 46100563 | Intron | SBS | G | C |
| INDIA 8544 | PPP1R14C | chr6 | 150148535 | Intron | SBS | C | G |
| INDIA 8544 | CIT | chr12 | 119691542 | Intron | SBS | G | C |
| INDIA 8544 | Unknown | chr14 | 75320548 | IGR | SBS | C | G |
| INDIA 8544 | Unknown | chr5 | 71780064 | IGR | INS | - | A |
| INDIA 8544 | GABBR2 | chr9 | 98596020 | Intron | DEL | T | - |
| INDIA 8544 | ITGA9 | chr3 | 37817050 | Intron | SBS | C | G |
| INDIA 8544 | PDPN | chr1 | 13582875 | 5'Flank | SBS | G | C |
| INDIA 8544 | NARF | chr17 | 82483953 | Intron | SBS | G | C |
| INDIA 8544 | LINC03000 | chr5 | 164406349 | Intron | SBS | A | T |
| INDIA 8544 | ENST00000660050 | chr10 | 132503515 | 3'Flank | SBS | C | G |
| INDIA 8544 | LIG3 | chr17 | 35008393 | 3'UTR | DEL | CATGTGCA | - |
| INDIA 8544 | COL4A2 | chr13 | 110425358 | Intron | SBS | G | C |
| INDIA 8544 | TAPT1-AS1 | chr4 | 16272253 | Intron | SBS | C | G |
| INDIA 8544 | ENST00000563114 | chr16 | 78052353 | Intron | SBS | C | G |
| INDIA 8544 | Unknown | chr14 | 26224795 | IGR | SBS | C | G |
| INDIA 8544 | EML6 | chr2 | 54722603 | 5'Flank | SBS | C | G |
| INDIA 8544 | TRPM8 | chr2 | 233975426 | Intron | SBS | C | G |
| INDIA 8544 | NARF | chr17 | 82484761 | Intron | SBS | G | C |
| INDIA 8544 | RBBP7 | chrX | 16854362 | Intron | SBS | G | C |
| INDIA 8544 | EXOSC7 | chr3 | 45016607 | 3'Flank | SBS | G | C |
| INDIA 8544 | DTX3L | chr3 | 122572369 | 3'UTR | SBS | C | G |
| INDIA 8544 | ASAP1 | chr8 | 130300830 | Intron | SBS | C | G |
| INDIA 8544 | ENST00000605292 | chr20 | 10801670 | Intron | SBS | C | G |
| INDIA 8544 | Unknown | chr4 | 35724511 | IGR | SBS | C | G |
| INDIA 8544 | Unknown | chr1 | 175768733 | IGR | SBS | G | C |
| INDIA 8544 | PARD3 | chr10 | 34363985 | Intron | SBS | A | T |
| INDIA 8544 | Unknown | chr12 | 109644214 | IGR | SBS | G | C |
| INDIA 8544 | RBFOX1 | chr16 | 7256189 | Intron | SBS | G | C |
| INDIA 8544 | ENST00000654007 | chr2 | 14737607 | Intron | SBS | C | G |
| INDIA 8544 | DRG2 | chr17 | 18109078 | 3'Flank | SBS | G | C |
| INDIA 8544 | ENST00000661295 | chr8 | 37327809 | RNA | SBS | G | C |
| INDIA 8544 | SKI | chr1 | 2278430 | Intron | SBS | G | C |
| INDIA 8544 | Unknown | chr2 | 476102 | IGR | SBS | C | G |
| INDIA 8544 | AP3B1 | chr5 | 77996165 | 3'Flank | SBS | A | C |
| INDIA 8544 | EPHB4 | chr7 | 100825531 | Intron | SBS | C | G |
| INDIA 8544 | ZC3H12A-DT | chr1 | 37406794 | Intron | SBS | T | G |
| INDIA 8544 | Unknown | chr4 | 174402483 | IGR | SBS | G | C |
| INDIA 8544 | ENST00000404626 | chr7 | 3144775 | 3'Flank | SBS | C | G |
| INDIA 8544 | TMEM132C | chr12 | 128502533 | Intron | SBS | G | C |
| INDIA 8544 | FOXJ3 | chr1 | 42184237 | Intron | SBS | C | G |

| | | | | | | | |
|---|---|---|---|---|---|---|---|
| INDIA 8544 | RICTOR | chr5 | 39074683 | 5'Flank | SBS | G | T |
| INDIA 8544 | SPAM1 | chr7 | 123931050 | Intron | SBS | C | G |
| INDIA 8544 | ABCF2 | chr7 | 151220690 | Intron | SBS | C | G |
| INDIA 8544 | Unknown | chr6 | 160334991 | IGR | SBS | C | G |
| INDIA 8544 | COX10-DT | chr17 | 13956022 | Intron | SBS | A | T |
| INDIA 8544 | PACRGL | chr4 | 20728898 | 3'UTR | SBS | A | T |
| INDIA 8544 | Unknown | chr17 | 73971078 | IGR | SBS | G | C |
| INDIA 8544 | RERE | chr1 | 8541425 | Intron | SBS | G | C |
| INDIA 8544 | DRG2 | chr17 | 18106994 | Intron | SBS | G | C |
| INDIA 8544 | VIRMA | chr8 | 94510620 | Silent | SBS | G | C |
| INDIA 8544 | GRM7 | chr3 | 7346538 | Intron | SBS | G | C |
| INDIA 8544 | PEMT | chr17 | 17555436 | Intron | SBS | C | G |
| INDIA 8547 | LAMC1 | chr1 | 183076864 | Intron | SBS | G | C |
| INDIA 8547 | Unknown | chrX | 120418616 | IGR | SBS | G | C |
| INDIA 8547 | KCNIP4 | chr4 | 21753935 | Intron | SBS | C | A |
| INDIA 8547 | RALGAPA2 | chr20 | 20444373 | Intron | SBS | C | G |
| INDIA 8547 | Unknown | chr10 | 85513035 | IGR | SBS | T | G |
| INDIA 8547 | PAPSS2 | chr10 | 87668759 | Intron | SBS | G | C |
| INDIA 8547 | NPS | chr10 | 127556230 | 3'Flank | SBS | G | C |
| INDIA 8547 | Unknown | chr7 | 21260804 | IGR | SBS | C | G |
| INDIA 8547 | ENST00000684900 | chr11 | 88429928 | 3'Flank | INS | - | GCTT |
| INDIA 8547 | DPEP2 | chr16 | 67992615 | Silent | SBS | G | C |
| INDIA 8547 | Unknown | chr1 | 112795857 | IGR | SBS | C | G |
| INDIA 8547 | UBE2Q2 | chr15 | 75868107 | Intron | SBS | C | G |
| INDIA 8547 | LINC01320 | chr2 | 34348299 | Intron | SBS | G | C |
| INDIA 8547 | IRGM | chr5 | 150847589 | Intron | SBS | C | G |
| INDIA 8547 | PLXNA4 | chr7 | 132218690 | Intron | SBS | C | A |
| INDIA 8547 | LYVE1 | chr11 | 10564688 | Intron | SBS | T | A |
| INDIA 8547 | GABRG3 | chr15 | 27538782 | 3'UTR | DEL | C | - |
| INDIA 8547 | ETFRF1 | chr12 | 25203351 | Intron | SBS | C | G |
| INDIA 8547 | LINC01036 | chr1 | 187368360 | Intron | SBS | T | A |
| INDIA 8547 | PARD3B | chr2 | 204665465 | Intron | SBS | C | G |
| INDIA 8547 | CPED1 | chr7 | 121114397 | Intron | DEL | T | - |
| INDIA 8547 | ABRAXAS2 | chr10 | 124802312 | Intron | SBS | C | G |
| INDIA 8547 | Unknown | chr4 | 115690777 | IGR | SBS | C | G |
| INDIA 8547 | Unknown | chr6 | 112045037 | IGR | SBS | C | G |
| INDIA 8547 | DYSF | chr2 | 71620442 | Intron | SBS | G | C |
| INDIA 8547 | Unknown | chr3 | 134464699 | IGR | SBS | C | G |
| INDIA 8547 | COL4A2 | chr13 | 110439913 | Intron | SBS | C | G |
| INDIA 8547 | NEFH | chr22 | 29478418 | 5'Flank | SBS | C | G |
| INDIA 8547 | PCMT1 | chr6 | 149811233 | 3'UTR | SBS | T | G |
| INDIA 8547 | RAI14 | chr5 | 34684474 | Intron | SBS | T | A |
| INDIA 8547 | SGMS1 | chr10 | 50376334 | Intron | DEL | T | - |
| INDIA 8547 | KBTBD11 | chr8 | 1991613 | Intron | SBS | C | G |
| INDIA 8547 | LRRK2 | chr12 | 40327971 | Intron | SBS | T | A |
| INDIA 8547 | DYRK2 | chr12 | 67648298 | 5'Flank | SBS | A | T |
| INDIA 8547 | AGBL1 | chr15 | 86228499 | Intron | SBS | G | C |
| INDIA 8547 | ABHD2 | chr15 | 89157448 | Intron | SBS | G | C |
| INDIA 8547 | LINC02882 | chr12 | 73835784 | Intron | SBS | C | G |
| INDIA 8547 | GAS7 | chr17 | 10088536 | Intron | SBS | G | T |
| INDIA 8547 | Unknown | chr1 | 165131744 | IGR | SBS | C | G |
| INDIA 8547 | TMEM108 | chr3 | 133079754 | Intron | SBS | C | G |
| INDIA 8547 | ATP1A2 | chr1 | 160128114 | Intron | SBS | C | G |
| INDIA 8547 | ATP7B | chr13 | 51981455 | Intron | SBS | C | G |
| INDIA 8547 | Unknown | chr18 | 11273054 | IGR | SBS | C | G |
| INDIA 8547 | Unknown | chr17 | 44588843 | IGR | SBS | G | C |
| INDIA 8547 | LAMC2 | chr1 | 183220972 | Intron | SBS | C | G |
| INDIA 8547 | SULF1 | chr8 | 69468689 | Intron | SBS | A | T |
| INDIA 8547 | Unknown | chr11 | 116709105 | IGR | SBS | G | C |
| INDIA 8547 | TPD52 | chr8 | 80064249 | Intron | SBS | C | G |
| INDIA 8547 | HS6ST3 | chr13 | 96745580 | Intron | SBS | C | A |
| INDIA 8547 | CTCF | chr16 | 67642247 | 3'Flank | INS | - | T |
| INDIA 8547 | PPP1R9A | chr7 | 95290368 | 3'UTR | SBS | C | G |
| INDIA 8547 | RBFOX1 | chr16 | 7644556 | Intron | SBS | G | C |
| INDIA 8547 | DSCAM | chr21 | 40547707 | Intron | SBS | C | G |
| INDIA 8547 | CTBP2 | chr10 | 125100371 | Intron | DEL | G | - |

| | | | | | | | |
|---|---|---|---|---|---|---|---|
| INDIA 8547 | RAB3B | chr1 | 51927005 | Intron | SBS | G | C |
| INDIA 8547 | DPT | chr1 | 168709275 | Intron | SBS | G | C |
| INDIA 8547 | ENST00000654974 | chr1 | 209244181 | Intron | SBS | G | C |
| INDIA 8547 | KCNH7 | chr2 | 162486196 | Intron | SBS | C | G |
| INDIA 8547 | TTN | chr2 | 178717540 | Missense_Mutation | SBS | G | T |
| INDIA 8547 | RBMS3-AS3 | chr3 | 29253379 | Intron | SBS | C | G |
| INDIA 8547 | Unknown | chr7 | 1204602 | IGR | SBS | G | C |
| INDIA 8547 | CPVL | chr7 | 29115707 | Intron | SBS | G | C |
| INDIA 8547 | PTK2B | chr8 | 27420658 | Missense_Mutation | SBS | T | A |
| INDIA 8547 | BTBD11 | chr12 | 107509296 | Intron | SBS | C | G |
| INDIA 8547 | ENST00000657016 | chr13 | 53252916 | Intron | SBS | C | G |
| INDIA 8547 | DPF3 | chr14 | 72728483 | Intron | SBS | T | A |
| INDIA 8547 | ENST00000637635 | chr16 | 50411018 | Intron | DEL | C | - |
| INDIA 8547 | CDH13 | chr16 | 82732589 | Intron | SBS | C | G |
| INDIA 8547 | PLCB1 | chr20 | 8311954 | Intron | SBS | T | A |
| INDIA 8547 | ENST00000554814 | chr14 | 81838858 | Intron | DEL | T | - |
| INDIA 8547 | LINC01234 | chr12 | 113679850 | Intron | SBS | T | A |
| INDIA 8548 | RAP1B | chr12 | 68658856 | Intron | SBS | G | C |
| INDIA 8548 | Unknown | chr13 | 87058649 | IGR | SBS | A | T |
| INDIA 8548 | AOPEP | chr9 | 95046609 | Intron | SBS | C | G |
| INDIA 8548 | PDHA2 | chr4 | 95839847 | 5'Flank | SBS | G | C |
| INDIA 8548 | Unknown | chr4 | 18398076 | IGR | SBS | G | C |
| INDIA 8548 | COG3 | chr13 | 45462126 | 5'Flank | SBS | A | T |
| INDIA 8548 | ATXN10 | chr22 | 45785240 | Intron | SBS | C | G |
| INDIA 8548 | USH2A | chr1 | 215874118 | Intron | SBS | C | A |
| INDIA 8548 | ELP3 | chr8 | 28102511 | Intron | SBS | C | G |
| INDIA 8548 | AP5B1 | chr11 | 65785556 | 5'Flank | SBS | C | G |
| INDIA 8548 | Unknown | chr12 | 120678338 | IGR | SBS | G | C |
| INDIA 8548 | UBN1 | chr16 | 4876324 | Intron | SBS | C | G |
| INDIA 8548 | ENST00000580422 | chr17 | 15413388 | Intron | SBS | C | A |
| INDIA 8548 | MGAT3 | chr22 | 39461288 | Intron | SBS | G | T |
| INDIA 8548 | MYDGF | chr19 | 4661792 | Intron | SBS | G | T |
| INDIA 8548 | RUNX1 | chr21 | 34986262 | Intron | SBS | C | G |
| INDIA 8548 | Unknown | chr5 | 175123431 | IGR | SBS | C | G |
| INDIA 8548 | HEATR1 | chr1 | 236553575 | Intron | SBS | C | G |
| INDIA 8548 | MBNL1-AS1 | chr3 | 152243358 | 3'Flank | SBS | T | G |
| INDIA 8548 | WDFY3 | chr4 | 84777480 | Intron | SBS | G | C |
| INDIA 8548 | ANKRD10 | chr13 | 110880029 | Missense_Mutation | SBS | G | C |
| INDIA 8548 | ATP13A4 | chr3 | 193545412 | Intron | SBS | G | C |
| INDIA 8548 | FAT3 | chr11 | 92554716 | Intron | SBS | T | A |
| INDIA 8548 | SLC49A3 | chr4 | 693323 | 5'Flank | SBS | C | G |
| INDIA 8548 | FLNC | chr7 | 128843174 | Intron | SBS | G | T |
| INDIA 8548 | Unknown | chr11 | 116704751 | IGR | SBS | C | G |
| INDIA 8548 | ENST00000560360 | chr15 | 98192734 | Intron | SBS | T | A |
| INDIA 8548 | RRAD | chr16 | 66926646 | 5'Flank | SBS | G | C |
| INDIA 8548 | LDLRAD4 | chr18 | 13625036 | Intron | SBS | C | A |
| INDIA 8548 | Unknown | chr20 | 50055021 | IGR | SBS | G | T |
| INDIA 8548 | PM20D1 | chr1 | 205852179 | 5'Flank | SBS | C | G |
| INDIA 8548 | SEC16A | chr9 | 136477295 | Nonsense_Mutation | SBS | A | T |
| INDIA 8548 | ABCB9 | chr12 | 122950076 | Intron | SBS | C | G |
| INDIA 8548 | Unknown | chr13 | 112121184 | IGR | SBS | C | G |
| INDIA 8548 | RPS6KA2 | chr6 | 166603469 | Intron | SBS | G | C |
| INDIA 8548 | FLII | chr17 | 18247312 | Frame_Shift_Del | DEL | CCACC | - |
| INDIA 8548 | Unknown | chr18 | 52008617 | IGR | SBS | C | G |
| INDIA 8548 | Unknown | chr6 | 52299815 | IGR | SBS | G | C |
| INDIA 8548 | IRX3 | chr16 | 54283488 | 3'UTR | SBS | G | C |
| INDIA 8548 | Unknown | chr10 | 9923681 | IGR | SBS | C | G |
| INDIA 8548 | GPC6 | chr13 | 93687782 | Intron | SBS | A | T |
| INDIA 8548 | LINC02534 | chr6 | 115630548 | 3'Flank | SBS | C | G |
| INDIA 8548 | SDK1 | chr7 | 4237979 | Intron | SBS | C | G |
| INDIA 8548 | RBFOX3 | chr17 | 79315411 | Intron | SBS | C | G |
| INDIA 8548 | CACNG6 | chr19 | 53992342 | 5'UTR | SBS | C | G |
| INDIA 8548 | EYA2 | chr20 | 47004842 | Intron | SBS | G | C |
| INDIA 8548 | ZBTB8B | chr1 | 32492676 | 3'UTR | SBS | G | C |
| INDIA 8548 | JARID2 | chr6 | 15432841 | Intron | SBS | A | C |
| INDIA 8548 | Unknown | chr5 | 2251934 | IGR | SBS | C | G |

| | | | | | | | |
|---|---|---|---|---|---|---|---|
| INDIA 8548 | CCDC88B | chr11 | 64352475 | Intron | SBS | A | T |
| INDIA 8548 | LINC01907 | chr2 | 230694230 | Intron | SBS | A | C |
| INDIA 8548 | THAP4 | chr2 | 241628392 | Intron | SBS | C | G |
| INDIA 8548 | ZBTB20 | chr3 | 114321805 | 3'UTR | SBS | A | T |
| INDIA 8548 | CHN2 | chr7 | 29393962 | Intron | SBS | G | C |
| INDIA 8548 | Unknown | chr8 | 141313518 | IGR | SBS | C | G |
| INDIA 8548 | MAP2K6 | chr17 | 69436169 | Intron | SBS | C | G |
| INDIA 8548 | Unknown | chrX | 137453403 | IGR | SBS | C | G |
| INDIA 8548 | PLXDC2 | chr10 | 19970919 | Intron | SBS | C | G |
| INDIA 8548 | TCF7L2 | chr10 | 113083607 | Intron | SBS | T | A |
| INDIA 8548 | Unknown | chr1 | 168735752 | IGR | SBS | G | C |
| INDIA 8548 | UNK | chr17 | 75824776 | 3'UTR | SBS | G | C |
| INDIA 8548 | Unknown | chr1 | 40897655 | IGR | SBS | C | G |
| INDIA 8548 | GOSR1 | chr17 | 30477043 | 5'Flank | SBS | T | A |
| INDIA 8548 | IRAG1 | chr11 | 10619424 | Intron | SBS | C | G |
| INDIA 8548 | ZNF710 | chr15 | 90035313 | Intron | SBS | C | G |
| INDIA 8548 | Unknown | chr16 | 86604487 | IGR | SBS | G | C |
| INDIA 8548 | RUNX2-AS1 | chr6 | 45569578 | 3'Flank | SBS | G | C |
| INDIA 8548 | MIR1471 | chr2 | 231891739 | 3'Flank | SBS | G | C |
| INDIA 8548 | CSNK1A1 | chr5 | 149549794 | Intron | INS | - | G |
| INDIA 8548 | ZMPSTE24 | chr1 | 40259900 | Intron | SBS | C | A |
| INDIA 8548 | HK2 | chr2 | 74886254 | Intron | SBS | G | C |
| INDIA 8548 | LINC02568 | chr15 | 63412274 | Intron | SBS | G | T |
| INDIA 8548 | DDB1 | chr11 | 61302164 | Intron | SBS | C | G |
| INDIA 8548 | OR5A1 | chr11 | 59447648 | 3'UTR | SBS | G | C |
| INDIA 8548 | Unknown | chr1 | 109783437 | IGR | DEL | AACCTG | - |
| INDIA 8548 | FANCA | chr16 | 89792332 | Intron | SBS | G | C |
| INDIA 8548 | LINC02543 | chr6 | 15998317 | Intron | SBS | C | G |
| INDIA 8548 | LINC01065 | chr13 | 53140324 | Intron | SBS | T | A |
| INDIA 8548 | ZMIZ1 | chr10 | 79224523 | Intron | SBS | A | T |
| INDIA 8548 | EPHA2 | chr1 | 16125893 | Intron | SBS | C | G |
| INDIA 8548 | NUS1 | chr6 | 117695607 | Intron | SBS | T | G |
| INDIA 8548 | FOXN2 | chr2 | 48324952 | Intron | SBS | G | C |
| INDIA 8548 | ENST00000564193 | chr16 | 47017787 | Intron | SBS | G | C |
| INDIA 8548 | KATNIP | chr16 | 27693211 | Intron | SBS | C | G |
| INDIA 8548 | Unknown | chr21 | 34346575 | IGR | SBS | A | T |
| INDIA 8549 | RIPOR2 | chr6 | 24897110 | Intron | SBS | C | G |
| INDIA 8549 | EOLA2-DT | chrX | 150161607 | Intron | SBS | C | G |
| INDIA 8549 | RPL26 | chr17 | 8372686 | 3'Flank | SBS | C | G |
| INDIA 8549 | Unknown | chr6 | 153747082 | IGR | SBS | G | C |
| INDIA 8549 | Unknown | chr18 | 29378734 | IGR | SBS | A | C |
| INDIA 8549 | Unknown | chr8 | 8508313 | IGR | SBS | C | G |
| INDIA 8549 | ENST00000411785 | chr2 | 66235946 | Intron | SBS | C | G |
| INDIA 8549 | TNNI3K | chr1 | 74466693 | Intron | SBS | G | C |
| INDIA 8549 | RNF4 | chr4 | 2498065 | Intron | SBS | G | C |
| INDIA 8549 | Unknown | chr9 | 108298068 | IGR | SBS | C | G |
| INDIA 8549 | FSD2 | chr15 | 82769791 | Missense_Mutation | SBS | G | T |
| INDIA 8549 | Unknown | chr16 | 50000826 | IGR | SBS | G | C |
| INDIA 8549 | ENST00000562656 | chr16 | 65329001 | Intron | SBS | G | C |
| INDIA 8549 | DOCK5 | chr8 | 25317041 | Missense_Mutation | SBS | C | G |
| INDIA 8549 | EBPL | chr13 | 49695247 | 5'Flank | SBS | C | G |
| INDIA 8549 | NALF1 | chr13 | 107573473 | Intron | SBS | C | G |
| INDIA 8549 | Unknown | chr7 | 149040632 | IGR | SBS | G | C |
| INDIA 8549 | SPOP | chr17 | 49622667 | Intron | SBS | C | G |
| INDIA 8549 | TTC7B | chr14 | 90677167 | Intron | SBS | C | G |
| INDIA 8549 | SLC8A1-AS1 | chr2 | 39894453 | Intron | SBS | C | G |
| INDIA 8549 | ZNF32-AS3 | chr10 | 43665405 | Intron | SBS | C | G |
| INDIA 8549 | Unknown | chr13 | 93106202 | IGR | SBS | G | C |
| INDIA 8549 | Unknown | chr5 | 82898517 | IGR | SBS | G | C |
| INDIA 8549 | TOM1L1 | chr17 | 54949720 | Intron | SBS | G | C |
| INDIA 8549 | KAZN | chr1 | 14655439 | Intron | SBS | G | C |
| INDIA 8549 | ILDR1 | chr3 | 122016664 | Intron | SBS | C | G |
| INDIA 8549 | CRAT37 | chr15 | 91556510 | Intron | SBS | C | G |
| INDIA 8549 | UTRN | chr6 | 144743082 | Intron | SBS | G | C |
| INDIA 8549 | CHN2 | chr7 | 29421890 | Intron | SBS | G | C |
| INDIA 8549 | RN7SL720P | chr9 | 17054703 | 3'Flank | SBS | G | C |

| | | | | | | | |
|---|---|---|---|---|---|---|---|
| INDIA 8549 | TOX3 | chr16 | 52471650 | Intron | SBS | C | G |
| INDIA 8549 | IQCA1 | chr2 | 236417272 | Intron | SBS | C | G |
| INDIA 8549 | ENST00000511431 | chr4 | 19683541 | Intron | SBS | A | C |
| INDIA 8549 | MCTP2 | chr15 | 94346666 | Intron | SBS | G | C |
| INDIA 8549 | Unknown | chr8 | 21380836 | IGR | SBS | C | G |
| INDIA 8549 | LRRC4C | chr11 | 41003308 | Intron | SBS | G | C |
| INDIA 8549 | B4GALT6 | chr18 | 31645143 | Intron | SBS | C | G |
| INDIA 8549 | TPGS1 | chr19 | 504579 | 5'Flank | SBS | C | G |
| INDIA 8549 | ENST00000446355 | chr7 | 116465313 | Intron | SBS | C | G |
| INDIA 8549 | MTAP | chr9 | 21815726 | Intron | SBS | G | C |
| INDIA 8549 | ENST00000444750 | chr1 | 212191519 | 3'Flank | SBS | G | C |
| INDIA 8549 | TTC30B | chr2 | 177548584 | 3'Flank | SBS | C | G |
| INDIA 8549 | RTN4RL1 | chr17 | 2018388 | Intron | SBS | C | G |
| INDIA 8549 | SULF2 | chr20 | 47664366 | Intron | SBS | G | C |
| INDIA 8549 | TOM1L2 | chr17 | 17855238 | Intron | SBS | G | C |
| INDIA 8549 | CYP1B1 | chr2 | 38071109 | Silent | SBS | G | C |
| INDIA 8549 | ENST00000659342 | chr11 | 22138672 | Intron | SBS | G | C |
| INDIA 8549 | TMEM178A | chr2 | 39677244 | Intron | SBS | C | G |
| INDIA 8549 | Unknown | chr9 | 21268649 | IGR | SBS | C | G |
| INDIA 8549 | LINC01332 | chr5 | 74336676 | 5'Flank | SBS | G | C |
| INDIA 8549 | RBP7 | chr1 | 10006108 | Intron | SBS | C | G |
| INDIA 8549 | KDM3A | chr2 | 86477688 | Intron | SBS | C | G |
| INDIA 8549 | Unknown | chr3 | 35835620 | IGR | SBS | G | C |
| INDIA 8549 | DCLK3 | chr3 | 36707621 | 3'Flank | SBS | G | C |
| INDIA 8549 | ENST00000510095 | chr4 | 11679653 | Intron | SBS | C | G |
| INDIA 8549 | VGLL2 | chr6 | 117268923 | Intron | SBS | G | C |
| INDIA 8549 | OR8K5 | chr11 | 56162004 | 5'Flank | SBS | G | C |
| INDIA 8549 | WASF3 | chr13 | 26601875 | Intron | SBS | G | C |
| INDIA 8549 | PSTPIP2 | chr18 | 46010963 | Intron | SBS | C | G |
| INDIA 8549 | Unknown | chr8 | 80600995 | IGR | SBS | C | G |
| INDIA 8549 | MTMR2 | chr11 | 95854004 | Intron | SBS | G | C |
| INDIA 8549 | KASH5 | chr19 | 49401406 | Intron | SBS | C | G |
| INDIA 8549 | SH2D3A | chr19 | 6767209 | Intron | SBS | G | C |
| INDIA 8549 | LINC02789 | chr1 | 199284736 | Intron | SBS | A | T |
| INDIA 8549 | DPYSL3 | chr5 | 147405307 | Intron | SBS | C | G |
| INDIA 8549 | LINC01833 | chr2 | 44920752 | 3'Flank | SBS | C | G |
| INDIA 8549 | DGKD | chr2 | 233376198 | Intron | SBS | C | G |
| INDIA 8549 | SLC38A8 | chr16 | 84011087 | Intron | SBS | G | C |
| INDIA 8549 | KLHL14 | chr18 | 32670267 | 3'Flank | DEL | C | - |
| INDIA 8549 | Unknown | chr2 | 174057319 | IGR | SBS | C | G |
| INDIA 8549 | Unknown | chr10 | 30748793 | IGR | SBS | C | G |
| INDIA 8549 | FOXJ1 | chr17 | 76140910 | Intron | SBS | C | G |
| INDIA 8549 | LRMDA | chr10 | 75496039 | Intron | SBS | C | G |
| INDIA 8549 | Unknown | chr2 | 30836364 | IGR | SBS | G | C |
| INDIA 8549 | AP1S3 | chr2 | 223728867 | Intron | SBS | C | G |
| INDIA 8549 | LINC01629 | chr14 | 76958640 | 5'Flank | SBS | G | C |
| INDIA 8549 | SHISA6 | chr17 | 11352429 | Intron | SBS | G | C |
| INDIA 8549 | MIR1-1HG | chr20 | 62574795 | 3'Flank | SBS | C | G |
| INDIA 8549 | RALB | chr2 | 120292171 | Intron | SBS | C | G |
| INDIA 8549 | PTPRG | chr3 | 61874289 | Intron | SBS | G | C |
| INDIA 8549 | PDZRN4 | chr12 | 41301756 | Intron | SBS | C | G |
| INDIA 8549 | FBN1 | chr15 | 48612409 | Intron | SBS | G | C |
| INDIA 8549 | Unknown | chr2 | 101631020 | IGR | SBS | C | G |
| INDIA 8549 | Unknown | chr2 | 133931459 | IGR | SBS | A | C |
| INDIA 8549 | PCDHGA1 | chr5 | 141326694 | 5'Flank | SBS | G | C |
| INDIA 8549 | ENST00000524772 | chr11 | 112797264 | 3'Flank | SBS | G | C |
| INDIA 8549 | ENST00000414039 | chr1 | 94323473 | Intron | SBS | G | C |
| INDIA 8549 | MROH2B | chr5 | 41064614 | Intron | SBS | C | G |
| INDIA 8549 | FAM178B | chr2 | 96882820 | Intron | SBS | C | G |
| INDIA 8549 | Unknown | chr1 | 23352553 | IGR | SBS | G | C |
| INDIA 8549 | EVA1B | chr1 | 36317068 | 3'Flank | SBS | G | C |
| INDIA 8549 | TMEM52B | chr12 | 10179088 | 5'UTR | SBS | G | C |
| INDIA 8549 | PTPN14 | chr1 | 214441416 | Intron | SBS | C | G |
| INDIA 8549 | CPNE4 | chr3 | 131575460 | Intron | SBS | C | G |
| INDIA 8549 | Unknown | chr18 | 7549018 | IGR | SBS | C | G |
| INDIA 8549 | Unknown | chr18 | 73480626 | IGR | SBS | G | C |

| | | | | | | | |
|---|---|---|---|---|---|---|---|
| INDIA 8550 | LRIG3-DT | chr12 | 58926800 | Intron | SBS | A | T |
| INDIA 8550 | ENST00000702412 | chr10 | 23473495 | Intron | SBS | G | C |
| INDIA 8550 | ABCB7 | chrX | 75155902 | Intron | INS | - | T |
| INDIA 8550 | Unknown | chr5 | 98046959 | IGR | DEL | G | - |
| INDIA 8550 | Unknown | chr20 | 16850027 | IGR | SBS | C | A |
| INDIA 8550 | PFKFB1 | chrX | 54934926 | Intron | SBS | G | C |
| INDIA 8550 | Unknown | chr3 | 190194676 | IGR | SBS | C | G |
| INDIA 8550 | ME3 | chr11 | 86473722 | Intron | SBS | G | C |
| INDIA 8550 | PSMD7-DT | chr16 | 74239055 | Intron | SBS | G | C |
| INDIA 8550 | ENST00000651680 | chr2 | 145660229 | Intron | SBS | T | A |
| INDIA 8550 | RABGAP1L | chr1 | 174825271 | Intron | SBS | G | C |
| INDIA 8550 | Unknown | chr4 | 58435931 | IGR | SBS | C | G |
| INDIA 8550 | ENST00000661809 | chr4 | 126538420 | Intron | SBS | G | C |
| INDIA 8550 | SEMA3C | chr7 | 80785949 | Intron | SBS | C | G |
| INDIA 8550 | Unknown | chr14 | 82092949 | IGR | SBS | C | G |
| INDIA 8550 | TFDP2 | chr3 | 141959806 | Missense_Mutation | SBS | C | G |
| INDIA 8550 | ENST00000654219 | chr2 | 76278269 | Intron | SBS | C | G |
| INDIA 8550 | ZNF423 | chr16 | 49584157 | Intron | SBS | G | C |
| INDIA 8550 | SIL1 | chr5 | 139214882 | Intron | SBS | C | G |
| INDIA 8550 | GRIN2B | chr12 | 13749957 | Intron | SBS | G | C |
| INDIA 8550 | SPTB | chr14 | 64799531 | Intron | SBS | G | C |
| INDIA 8550 | IL34 | chr16 | 70656563 | Intron | SBS | C | G |
| INDIA 8550 | SLC7A13 | chr8 | 86274520 | Intron | SBS | C | G |
| INDIA 8550 | NKAIN2 | chr6 | 124769726 | Intron | SBS | G | C |
| INDIA 8550 | SEMA3C | chr7 | 80818267 | Intron | SBS | C | G |
| INDIA 8550 | SNTB1 | chr8 | 120771086 | Intron | SBS | A | C |
| INDIA 8550 | OLFM3 | chr1 | 101914475 | Intron | SBS | G | T |
| INDIA 8550 | ENST00000431862 | chr1 | 159342929 | 3'Flank | SBS | C | A |
| INDIA 8550 | LINC01619 | chr12 | 92054529 | Intron | SBS | A | T |
| INDIA 8550 | DPP6 | chr7 | 154372976 | Intron | DEL | C | - |
| INDIA 8550 | SPTAN1 | chr9 | 128608135 | Frame_Shift_Del | DEL | C | - |
| INDIA 8550 | Unknown | chr17 | 34443752 | IGR | SBS | G | C |
| INDIA 8550 | LCORL | chr4 | 18012293 | Intron | SBS | C | G |
| INDIA 8550 | Unknown | chr2 | 149237590 | IGR | SBS | G | C |
| INDIA 8550 | ENST00000556361 | chr14 | 66281518 | Intron | SBS | G | C |
| INDIA 8550 | LINC02254 | chr15 | 97495170 | Intron | SBS | G | C |
| INDIA 8550 | Unknown | chr2 | 35730150 | IGR | SBS | C | G |
| INDIA 8550 | Unknown | chr3 | 135548115 | IGR | SBS | C | G |
| INDIA 8550 | ENST00000651847 | chr5 | 126096980 | Intron | SBS | T | A |
| INDIA 8550 | ALK | chr2 | 29320581 | Intron | SBS | C | G |
| INDIA 8550 | ENST00000447311 | chr6 | 142566910 | Intron | SBS | G | C |
| INDIA 8550 | Unknown | chr10 | 120104755 | IGR | SBS | C | G |
| INDIA 8550 | PKN2-AS1 | chr1 | 88228392 | Intron | SBS | G | C |
| INDIA 8550 | CSNK1G1 | chr15 | 64234435 | Intron | SBS | C | G |
| INDIA 8550 | Unknown | chr3 | 168991930 | IGR | SBS | C | G |
| INDIA 8550 | DSCAML1 | chr11 | 117648034 | Intron | SBS | C | A |
| INDIA 8550 | PLEKHG1 | chr6 | 150620310 | Intron | INS | - | T |
| INDIA 8550 | ENST00000649475 | chr3 | 14788816 | 5'Flank | SBS | C | G |
| INDIA 8550 | LMTK2 | chr7 | 98159770 | Intron | SBS | G | C |
| INDIA 8550 | Unknown | chr8 | 48375433 | IGR | SBS | C | G |
| INDIA 8550 | ENST00000657880 | chr2 | 121972670 | Intron | SBS | C | G |
| INDIA 8550 | Unknown | chr8 | 142945569 | IGR | SBS | C | G |
| INDIA 8550 | Unknown | chr1 | 232667526 | IGR | SBS | C | G |
| INDIA 8550 | ENST00000649194 | chr18 | 37660326 | Intron | SBS | C | A |
| INDIA 8550 | LINC-PINT | chr7 | 131029647 | Intron | SBS | C | G |
| INDIA 8550 | GRHL3 | chr1 | 24349645 | Intron | SBS | A | T |
| INDIA 8550 | MPPED2 | chr11 | 30517673 | Intron | SBS | A | T |
| INDIA 8550 | Unknown | chr20 | 41862503 | IGR | SBS | C | G |
| INDIA 8550 | PRDM1 | chr6 | 106078080 | Intron | SBS | G | C |
| INDIA 8550 | DNAH9 | chr17 | 11808558 | Intron | SBS | G | C |
| INDIA 8550 | SLIT3 | chr5 | 168984348 | Intron | SBS | G | C |
| INDIA 8550 | Unknown | chr2 | 1781273 | IGR | SBS | G | T |
| INDIA 8550 | ENST00000660587 | chr7 | 34237968 | Intron | SBS | C | G |
| INDIA 8550 | ENST00000397264 | chr10 | 5946037 | 5'Flank | SBS | C | G |
| INDIA 8550 | Unknown | chr10 | 36899846 | IGR | SBS | A | T |
| INDIA 8550 | SPAG16 | chr2 | 213989021 | Intron | SBS | G | C |

| | | | | | | | |
|---|---|---|---|---|---|---|---|
| INDIA 8550 | CLEC16A | chr16 | 11017113 | Intron | SBS | A | C |
| INDIA 8550 | Unknown | chr6 | 12474402 | IGR | SBS | G | C |
| INDIA 8550 | KLHL25 | chr15 | 85793142 | Intron | SBS | C | G |
| INDIA 8550 | CACNB4 | chr2 | 151987652 | Intron | SBS | C | G |
| INDIA 8550 | Unknown | chr7 | 86505240 | IGR | SBS | G | C |
| INDIA 8550 | PRDM8 | chr4 | 80197061 | 5'Flank | SBS | G | C |
| INDIA 8550 | PDE4B | chr1 | 66307742 | Intron | SBS | G | C |
| INDIA 8550 | Unknown | chr4 | 147279704 | IGR | SBS | C | G |
| INDIA 8550 | Unknown | chr7 | 22083294 | IGR | SBS | G | C |
| INDIA 8550 | GRM3 | chr7 | 86654483 | Intron | SBS | G | C |
| INDIA 8550 | FYN | chr6 | 111761200 | Intron | SBS | G | C |
| INDIA 8550 | RNF32-DT | chr7 | 156506665 | Intron | SBS | C | G |
| INDIA 8550 | FLRT2 | chr14 | 85569436 | Intron | DEL | G | - |
| INDIA 8550 | SPECC1L | chr22 | 24408703 | Intron | SBS | T | A |
| INDIA 8550 | LCE6A | chr1 | 152841785 | 5'Flank | SBS | G | T |
| INDIA 8550 | TLX3 | chr5 | 171317045 | 3'Flank | SBS | G | T |
| INDIA 8550 | ENPP6 | chr4 | 184176571 | Intron | SBS | G | C |
| INDIA 8550 | Unknown | chr22 | 48396461 | IGR | SBS | T | A |
| INDIA 8551 | Unknown | chr12 | 73486868 | IGR | SBS | T | A |
| INDIA 8551 | EPM2AIP1 | chr3 | 36982604 | 3'Flank | SBS | C | G |
| INDIA 8551 | MICAL2 | chr11 | 12186980 | Intron | SBS | G | C |
| INDIA 8551 | Unknown | chr2 | 156273959 | IGR | SBS | G | C |
| INDIA 8551 | MYO7B | chr2 | 127569955 | Intron | SBS | G | C |
| INDIA 8551 | FGFR1 | chr8 | 38451750 | Intron | DEL | G | - |
| INDIA 8551 | Unknown | chrX | 20680239 | IGR | SBS | C | G |
| INDIA 8551 | ENST00000658975 | chr9 | 121392807 | Intron | SBS | C | G |
| INDIA 8551 | PLCB1 | chr20 | 8230234 | Intron | SBS | C | G |
| INDIA 8551 | LINC01829 | chr2 | 67351981 | Intron | DEL | T | - |
| INDIA 8551 | WASHC5 | chr8 | 125075662 | Intron | DEL | AA | - |
| INDIA 8551 | ABL1 | chr9 | 130780485 | Intron | SBS | C | G |
| INDIA 8551 | Unknown | chr7 | 98088324 | IGR | SBS | C | G |
| INDIA 8551 | Unknown | chr7 | 149741629 | IGR | SBS | G | C |
| INDIA 8551 | LINC02427 | chr4 | 184512928 | Intron | SBS | G | C |
| INDIA 8551 | ALCAM | chr3 | 105540702 | Intron | SBS | G | C |
| INDIA 8551 | U8 | chr2 | 86347457 | 5'Flank | SBS | G | C |
| INDIA 8551 | ZNF469 | chr16 | 88433164 | Missense_Mutation | SBS | G | C |
| INDIA 8551 | LINC01807 | chr2 | 228668835 | Intron | DEL | CTA | - |
| INDIA 8551 | CNNM3 | chr2 | 96829113 | Missense_Mutation | SBS | G | C |
| INDIA 8551 | Unknown | chr19 | 7582613 | IGR | SBS | C | G |
| INDIA 8551 | ARID1B | chr6 | 156876678 | Intron | DEL | TC | - |
| INDIA 8551 | NEIL2 | chr8 | 11784177 | Intron | SBS | G | C |
| INDIA 8551 | Unknown | chr8 | 40475997 | IGR | SBS | A | T |
| INDIA 8551 | Unknown | chr8 | 141685632 | IGR | SBS | C | G |
| INDIA 8551 | SPRY4-AS1 | chr5 | 142530220 | Intron | SBS | C | G |
| INDIA 8551 | ZFHX2 | chr14 | 23530764 | Intron | SBS | C | G |
| INDIA 8551 | Unknown | chr3 | 104124181 | IGR | SBS | G | C |
| INDIA 8551 | FOXP2 | chr7 | 114129409 | Intron | SBS | T | A |
| INDIA 8551 | SETBP1 | chr18 | 44977595 | Intron | SBS | T | G |
| INDIA 8551 | Unknown | chr18 | 38420690 | IGR | SBS | C | G |
| INDIA 8551 | Unknown | chr1 | 2889321 | IGR | SBS | C | A |
| INDIA 8551 | Unknown | chr10 | 2285409 | IGR | SBS | C | A |
| INDIA 8551 | CTIF | chr18 | 48632241 | Intron | SBS | G | C |
| INDIA 8551 | LINC01513 | chr5 | 10482899 | 3'Flank | SBS | G | C |
| INDIA 8551 | LMCD1-AS1 | chr3 | 8249275 | Intron | SBS | G | C |
| INDIA 8551 | FAM171A1 | chr10 | 15367847 | Intron | SBS | C | G |
| INDIA 8551 | POLD4 | chr11 | 67358171 | 5'Flank | SBS | G | C |
| INDIA 8551 | ATXN7L1 | chr7 | 105700058 | Intron | SBS | G | C |
| INDIA 8551 | LYN | chr8 | 55979533 | Intron | SBS | G | C |
| INDIA 8551 | Unknown | chr11 | 110935134 | IGR | SBS | G | C |
| INDIA 8551 | CHST9 | chr18 | 27136484 | Intron | SBS | C | G |
| INDIA 8551 | TECR | chr19 | 14538486 | Intron | SBS | G | C |
| INDIA 8551 | LSAMP | chr3 | 116339180 | Intron | DEL | AG | - |
| INDIA 8551 | FNDC3A | chr13 | 48974895 | 5'Flank | INS | - | A |
| INDIA 8551 | Unknown | chr10 | 8758192 | IGR | SBS | C | G |
| INDIA 8551 | DNMT3A | chr2 | 25294647 | Intron | SBS | C | G |
| INDIA 8551 | Unknown | chr1 | 163697440 | IGR | SBS | G | C |

| | | | | | | | |
|---|---|---|---|---|---|---|---|
| INDIA 8551 | MYO18B | chr22 | 25991602 | Intron | SBS | G | C |
| INDIA 8551 | Unknown | chr17 | 56699762 | IGR | SBS | G | C |
| INDIA 8551 | UIMC1 | chr5 | 176937808 | Intron | SBS | G | C |
| INDIA 8551 | TMTC2 | chr12 | 82688076 | Intron | SBS | G | C |
| INDIA 8551 | Unknown | chr6 | 169200937 | IGR | SBS | C | G |
| INDIA 8551 | Unknown | chr1 | 59936618 | IGR | SBS | C | G |
| INDIA 8551 | TMEM150B | chr19 | 55321155 | Intron | SBS | C | G |
| INDIA 8551 | NLRP4 | chr19 | 55858243 | Missense_Mutation | SBS | C | A |
| INDIA 8551 | DPP6 | chr7 | 154518048 | Intron | SBS | T | A |
| INDIA 8551 | Unknown | chr6 | 49651728 | IGR | SBS | A | T |
| INDIA 8551 | Unknown | chr6 | 117239692 | IGR | SBS | C | G |
| INDIA 8551 | LINC02888 | chr7 | 17412676 | Intron | INS | - | AGA |
| INDIA 8551 | MYOT | chr5 | 137880500 | Intron | SBS | C | G |
| INDIA 8551 | GABRG3 | chr15 | 27176738 | Intron | SBS | G | C |
| INDIA 8551 | LITAF | chr16 | 11585413 | Intron | DEL | TTTC | - |
| INDIA 8551 | ENST00000641859 | chr22 | 39274055 | Intron | SBS | T | A |
| INDIA 8551 | GNAO1-DT | chr16 | 56023745 | Intron | SBS | G | C |
| INDIA 8551 | ENGASE | chr17 | 79074677 | 5'Flank | SBS | C | G |
| INDIA 8551 | TMEM169 | chr2 | 216089109 | Intron | SBS | G | C |
| INDIA 8551 | RNU6-604P | chr7 | 151901090 | 3'Flank | SBS | G | C |
| INDIA 8551 | LIMK2 | chr22 | 31227372 | Intron | SBS | G | C |
| INDIA 8551 | OBSCN | chr1 | 228349776 | Intron | SBS | C | G |
| INDIA 8551 | WWC2 | chr4 | 183118996 | Intron | SBS | G | C |
| INDIA 8551 | SIGLECL1 | chr19 | 51251821 | Intron | SBS | C | G |
| INDIA 8551 | LARGE1 | chr22 | 33583111 | Intron | SBS | C | G |
| INDIA 8551 | LINC02542 | chr6 | 81930262 | Intron | SBS | C | G |
| INDIA 8551 | USE1 | chr19 | 17213688 | 5'Flank | SBS | C | G |
| INDIA 8551 | PAPLN | chr14 | 73272647 | Missense_Mutation | SBS | C | G |
| INDIA 8551 | Unknown | chr15 | 56491929 | IGR | DEL | A | - |
| INDIA 8551 | Unknown | chr22 | 33952580 | IGR | SBS | C | G |
| INDIA 8551 | CCNF | chr16 | 2436454 | Intron | SBS | C | G |
| INDIA 8551 | ERC2 | chr3 | 56017882 | Intron | SBS | C | G |
| INDIA 8551 | Unknown | chr2 | 236864033 | IGR | SBS | C | G |
| INDIA 8551 | FAM135B | chr8 | 138483456 | Intron | SBS | G | T |
| INDIA 8551 | MIR4300HG | chr11 | 82528622 | Intron | SBS | G | C |
| INDIA 8552 | Unknown | chr2 | 3172277 | IGR | SBS | C | G |
| INDIA 8552 | ADAMTSL1 | chr9 | 18652953 | Intron | SBS | G | C |
| INDIA 8552 | Unknown | chr9 | 131838356 | IGR | SBS | C | G |
| INDIA 8552 | SFMBT2 | chr10 | 7312940 | Intron | SBS | C | G |
| INDIA 8552 | Unknown | chr10 | 5466648 | IGR | SBS | G | C |
| INDIA 8552 | Unknown | chr15 | 55018678 | IGR | SBS | G | C |
| INDIA 8552 | WDR24 | chr16 | 683442 | 3'Flank | SBS | C | A |
| INDIA 8552 | RIMS2 | chr8 | 103920543 | Intron | SBS | C | G |
| INDIA 8552 | OR2C3 | chr1 | 247527456 | 3'UTR | SBS | T | G |
| INDIA 8552 | LINC01735 | chr1 | 208606807 | RNA | SBS | C | G |
| INDIA 8552 | Unknown | chr13 | 104241358 | IGR | SBS | A | C |
| INDIA 8552 | MRAP2 | chr6 | 84060247 | Intron | SBS | C | G |
| INDIA 8552 | SNX29 | chr16 | 12412858 | Intron | SBS | C | G |
| INDIA 8552 | PUDP | chrX | 6931168 | Intron | SBS | C | G |
| INDIA 8552 | Unknown | chr2 | 147737764 | IGR | SBS | G | C |
| INDIA 8552 | Unknown | chr21 | 34000890 | IGR | SBS | G | C |
| INDIA 8552 | ENST00000583662 | chr17 | 17612149 | 3'Flank | SBS | C | G |
| INDIA 8552 | PUS10 | chr2 | 61016804 | Intron | SBS | C | G |
| INDIA 8552 | Unknown | chr2 | 128497878 | IGR | SBS | C | G |
| INDIA 8552 | CADPS | chr3 | 62447561 | Intron | SBS | C | G |
| INDIA 8552 | CYYR1 | chr21 | 26492357 | Intron | SBS | G | C |
| INDIA 8552 | IPO9 | chr1 | 201833159 | Intron | SBS | G | C |
| INDIA 8552 | Unknown | chr2 | 124976944 | IGR | SBS | C | G |
| INDIA 8552 | CUL4B | chrX | 120549861 | Intron | SBS | T | G |
| INDIA 8552 | FILIP1 | chr6 | 75444318 | Intron | SBS | G | C |
| INDIA 8552 | ATP8A2 | chr13 | 25919347 | Intron | SBS | G | C |
| INDIA 8552 | EFNB3 | chr17 | 7711384 | 3'Flank | SBS | G | C |
| INDIA 8552 | REL-DT | chr2 | 60849701 | Intron | SBS | C | G |
| INDIA 8552 | Unknown | chr1 | 125066075 | IGR | SBS | C | A |
| INDIA 8552 | Unknown | chr1 | 125066076 | IGR | SBS | C | A |
| INDIA 8552 | Unknown | chr2 | 239745582 | IGR | SBS | C | A |

| | | | | | | | |
|---|---|---|---|---|---|---|---|
| INDIA 8552 | HIVEP2 | chr6 | 142843083 | Intron | SBS | C | G |
| INDIA 8552 | TRHR | chr8 | 109116911 | Intron | SBS | A | T |
| INDIA 8552 | Unknown | chr17 | 70661338 | IGR | SBS | G | C |
| INDIA 8552 | DGAT2 | chr11 | 75789912 | Intron | SBS | C | G |
| INDIA 8552 | ENST00000700867 | chr14 | 97176325 | 5'Flank | SBS | C | G |
| INDIA 8552 | GALNTL6 | chr4 | 171860889 | Intron | SBS | G | C |
| INDIA 8552 | MLPH | chr2 | 237498637 | Intron | SBS | G | C |
| INDIA 8552 | DEF8 | chr16 | 89951552 | Intron | SBS | C | G |
| INDIA 8552 | Unknown | chr9 | 135296925 | IGR | SBS | G | C |
| INDIA 8552 | RCN1 | chr11 | 32090714 | 5'Flank | SBS | G | C |
| INDIA 8552 | KCNK9 | chr8 | 139683157 | Intron | SBS | G | C |
| INDIA 8552 | Unknown | chr14 | 99057752 | IGR | SBS | G | C |
| INDIA 8552 | EN2-DT | chr7 | 155449146 | Intron | SBS | G | C |
| INDIA 8552 | CSMD1 | chr8 | 4023463 | Intron | SBS | G | C |
| INDIA 8552 | TPD52 | chr8 | 80192031 | Intron | SBS | A | C |
| INDIA 8552 | RORB | chr9 | 74565522 | Intron | SBS | G | C |
| INDIA 8552 | ENST00000554926 | chr14 | 76537777 | Intron | SBS | C | G |
| INDIA 8552 | CRAT37 | chr15 | 91623400 | Intron | SBS | C | G |
| INDIA 8552 | LINC02987 | chr19 | 27937391 | Intron | SBS | C | G |
| INDIA 8552 | PARD3 | chr10 | 34218506 | Intron | SBS | C | G |
| INDIA 8552 | CTIF | chr18 | 48675643 | Intron | SBS | G | C |
| INDIA 8552 | TAFA1 | chr3 | 68212917 | Intron | SBS | C | G |
| INDIA 8552 | Unknown | chr4 | 20169237 | IGR | SBS | T | A |
| INDIA 8552 | ENST00000681682 | chr4 | 111317561 | Intron | SBS | G | C |
| INDIA 8552 | Unknown | chr7 | 153532563 | IGR | SBS | C | G |
| INDIA 8552 | QSOX2 | chr9 | 136236265 | Intron | SBS | C | G |
| INDIA 8552 | CYYR1 | chr21 | 26524461 | Intron | SBS | T | G |
| INDIA 8552 | Unknown | chrX | 132601350 | IGR | SBS | C | G |
| INDIA 8552 | ENST00000507435 | chr5 | 4604523 | Intron | SBS | C | G |
| INDIA 8552 | CCND3 | chr6 | 42044548 | Intron | SBS | G | C |
| INDIA 8552 | SMARCAL1 | chr2 | 216447399 | Intron | SBS | G | C |
| INDIA 8552 | ANK2 | chr4 | 113071364 | Intron | SBS | G | C |
| INDIA 8552 | ENST00000420572 | chr6 | 22949209 | Intron | SBS | C | G |
| INDIA 8552 | SLC12A1 | chr15 | 48265555 | Intron | SBS | G | C |
| INDIA 8552 | Unknown | chr9 | 71477917 | IGR | SBS | G | C |
| INDIA 8552 | Unknown | chr16 | 77003534 | IGR | SBS | G | C |
| INDIA 8552 | ZKSCAN5 | chr7 | 99512746 | Intron | SBS | G | C |
| INDIA 8552 | ENST00000664509 | chr7 | 431947 | Intron | SBS | G | T |
| INDIA 8552 | ATP1A1 | chr1 | 116374567 | Intron | SBS | T | A |
| INDIA 8552 | Unknown | chr9 | 105910201 | IGR | SBS | G | C |
| INDIA 8552 | RAPGEF4 | chr2 | 172894323 | Intron | SBS | G | C |
| INDIA 8552 | MYO15A | chr17 | 18166928 | Intron | SBS | C | G |
| INDIA 8552 | ATP8A1 | chr4 | 42483549 | Intron | SBS | C | G |
| INDIA 8552 | HDLBP | chr2 | 241269951 | Intron | SBS | C | G |
| INDIA 8552 | CFAP221 | chr2 | 119600074 | Intron | SBS | C | G |
| INDIA 8552 | MAN2A2 | chr15 | 90923971 | 3'Flank | SBS | G | C |
| INDIA 8552 | ARSG | chr17 | 68347136 | Missense_Mutation | SBS | C | G |
| INDIA 8552 | Unknown | chr2 | 149528086 | IGR | SBS | C | G |
| INDIA 8552 | ENST00000383808 | chr3 | 10007865 | Intron | SBS | G | C |
| INDIA 8552 | MCF2L2 | chr3 | 183332233 | Intron | SBS | C | G |
| INDIA 8552 | MCF2L2 | chr3 | 183332464 | Intron | SBS | C | G |
| INDIA 8552 | Unknown | chr3 | 190269351 | IGR | SBS | T | A |
| INDIA 8552 | ENST00000511431 | chr4 | 19773789 | Intron | SBS | C | G |
| INDIA 8552 | ENST00000641328 | chr4 | 143737276 | Intron | SBS | T | A |
| INDIA 8552 | Unknown | chr4 | 186934772 | IGR | SBS | C | G |
| INDIA 8552 | Unknown | chr5 | 12502034 | IGR | SBS | G | C |
| INDIA 8552 | ADAMTS6 | chr5 | 65190001 | Intron | SBS | C | G |
| INDIA 8552 | SH3BGRL2 | chr6 | 79683804 | Intron | SBS | C | G |
| INDIA 8552 | IGF2BP3 | chr7 | 23367625 | Intron | SBS | C | G |
| INDIA 8552 | Unknown | chr8 | 52100991 | IGR | SBS | G | C |
| INDIA 8552 | FLJ46284 | chr8 | 92716084 | RNA | SBS | C | G |
| INDIA 8552 | KIAA1217 | chr10 | 24241688 | Intron | SBS | C | G |
| INDIA 8552 | Unknown | chr10 | 108222403 | IGR | SBS | G | C |
| INDIA 8553 | Unknown | chr12 | 86860510 | IGR | DEL | AG | - |
| INDIA 8553 | ENST00000652510 | chr20 | 39655299 | 3'Flank | SBS | A | T |
| INDIA 8553 | TAB3 | chrX | 30840776 | Intron | DEL | G | - |

| | | | | | | | |
|---|---|---|---|---|---|---|---|
| INDIA 8553 | CDKAL1 | chr6 | 21033162 | Intron | SBS | G | C |
| INDIA 8553 | PRKN | chr6 | 162227216 | Intron | SBS | G | C |
| INDIA 8553 | SRGAP1 | chr12 | 64015673 | Intron | SBS | G | C |
| INDIA 8553 | ITSN1 | chr21 | 33684359 | Intron | SBS | G | C |
| INDIA 8553 | TMEM74 | chr8 | 108635407 | Intron | SBS | C | G |
| INDIA 8553 | ENST00000484222 | chr3 | 67311251 | 3'Flank | SBS | T | A |
| INDIA 8553 | ENST00000509416 | chr4 | 28580079 | Intron | SBS | A | T |
| INDIA 8553 | KC6 | chr18 | 41586839 | Intron | SBS | G | C |
| INDIA 8553 | RBM5 | chr3 | 50086614 | 5'Flank | SBS | C | G |
| INDIA 8553 | ATR | chr3 | 142560342 | Missense_Mutation | SBS | C | G |
| INDIA 8553 | CYSTM1 | chr5 | 140227407 | Intron | SBS | G | C |
| INDIA 8553 | Unknown | chr12 | 72887800 | IGR | SBS | C | G |
| INDIA 8553 | ENST00000433210 | chr21 | 19303866 | 3'Flank | SBS | T | A |
| INDIA 8553 | LUZP4 | chrX | 115290714 | Intron | SBS | C | G |
| INDIA 8553 | Unknown | chrX | 140367687 | IGR | DEL | T | - |
| INDIA 8553 | LINC01505 | chr9 | 106598171 | Intron | SBS | C | G |
| INDIA 8553 | CNTN4 | chr3 | 2768597 | Intron | DEL | T | - |
| INDIA 8553 | ATP8A2 | chr13 | 25862279 | Intron | SBS | G | C |
| INDIA 8553 | Unknown | chr1 | 184213355 | IGR | SBS | T | A |
| INDIA 8553 | KCNIP1 | chr5 | 170362991 | Intron | SBS | C | G |
| INDIA 8553 | GRIK2 | chr6 | 100964600 | Intron | SBS | C | G |
| INDIA 8553 | COBL | chr7 | 51238789 | Intron | SBS | A | T |
| INDIA 8553 | Unknown | chr16 | 7741755 | IGR | SBS | C | G |
| INDIA 8553 | KIF1C | chr17 | 5028036 | 3'UTR | SBS | C | G |
| INDIA 8553 | PTPRT | chr20 | 43189094 | Intron | SBS | G | C |
| INDIA 8553 | ENST00000429588 | chr21 | 36432383 | Intron | DEL | C | - |
| INDIA 8553 | PHC2-AS1 | chr1 | 33354875 | Intron | SBS | G | C |
| INDIA 8553 | NCMAP | chr1 | 24569370 | Intron | SBS | C | G |
| INDIA 8553 | Unknown | chr9 | 25863402 | IGR | SBS | C | G |
| INDIA 8553 | AOAH | chr7 | 36564798 | Intron | SBS | C | G |
| INDIA 8553 | Unknown | chr10 | 92787067 | IGR | DEL | GT | - |
| INDIA 8553 | ENST00000554245 | chr14 | 100049500 | 3'Flank | SBS | G | C |
| INDIA 8553 | CDH11 | chr16 | 65037631 | Intron | SBS | G | C |
| INDIA 8553 | KCTD16 | chr5 | 144298511 | Intron | SBS | C | G |
| INDIA 8553 | ENST00000646617 | chr5 | 162753909 | Intron | SBS | T | A |
| INDIA 8553 | ENST00000665012 | chr8 | 90304445 | Intron | SBS | C | G |
| INDIA 8553 | ABCA12 | chr2 | 215140805 | 5'Flank | SBS | G | C |
| INDIA 8553 | SSUH2 | chr3 | 8707063 | Intron | SBS | A | T |
| INDIA 8553 | FGF12 | chr3 | 192438697 | Intron | SBS | G | C |
| INDIA 8553 | ENST00000629853 | chr6 | 14954040 | Intron | SBS | C | G |
| INDIA 8553 | Unknown | chr11 | 17678735 | IGR | DEL | T | - |
| INDIA 8553 | ENST00000549930 | chr12 | 93092773 | Intron | SBS | C | G |
| INDIA 8553 | Unknown | chr15 | 69896341 | IGR | SBS | C | G |
| INDIA 8553 | FHOD1 | chr16 | 67232567 | Intron | SBS | A | T |
| INDIA 8553 | LINC00687 | chr20 | 11831912 | Intron | SBS | C | G |
| INDIA 8553 | XAGE1B | chrX | 52509226 | 3'Flank | SBS | C | G |
| INDIA 8553 | Unknown | chrX | 96456182 | IGR | SBS | C | A |
| INDIA 8553 | PRDM6 | chr5 | 123182434 | Intron | SBS | G | C |
| INDIA 8553 | Unknown | chr3 | 84303482 | IGR | SBS | T | A |
| INDIA 8553 | ENST00000444770 | chr10 | 63836246 | Intron | SBS | C | G |
| INDIA 8553 | NXPH4 | chr12 | 57220547 | Intron | SBS | G | C |
| INDIA 8553 | Unknown | chr15 | 78428367 | IGR | SBS | C | G |
| INDIA 8553 | Unknown | chr2 | 56703148 | IGR | DEL | A | - |
| INDIA 8553 | LSAMP | chr3 | 116855993 | Intron | SBS | C | G |
| INDIA 8553 | EGFR | chr7 | 55187231 | Intron | SBS | G | C |
| INDIA 8553 | Unknown | chr14 | 24766849 | IGR | SBS | C | G |
| INDIA 8553 | Unknown | chr16 | 48795111 | IGR | SBS | C | G |
| INDIA 8553 | ENST00000593140 | chr19 | 30661250 | 3'Flank | SBS | G | C |
| INDIA 8553 | UBE2E2 | chr3 | 23234562 | Intron | SBS | A | T |
| INDIA 8553 | LINC02006 | chr3 | 153975356 | Intron | SBS | G | C |
| INDIA 8553 | Unknown | chr4 | 37169574 | IGR | SBS | G | C |
| INDIA 8553 | CMYA5 | chr5 | 79731413 | Nonsense_Mutation | SBS | C | G |
| INDIA 8553 | RN7SL250P | chr8 | 54417697 | 5'Flank | SBS | C | G |
| INDIA 8553 | TRHDE | chr12 | 72512119 | Intron | SBS | G | C |
| INDIA 8553 | Unknown | chr13 | 37474272 | IGR | SBS | A | T |
| INDIA 8553 | LRRC49 | chr15 | 70914840 | Intron | SBS | C | G |

| | | | | | | | |
|---|---|---|---|---|---|---|---|
| INDIA 8553 | LINC02131 | chr16 | 77605665 | Intron | SBS | C | G |
| INDIA 8553 | SETBP1 | chr18 | 44703975 | Intron | SBS | G | C |
| INDIA 8553 | OLFM2 | chr19 | 9920558 | Intron | SBS | C | G |
| INDIA 8553 | PDYN | chr20 | 1980722 | Silent | SBS | C | G |
| INDIA 8553 | MID2 | chrX | 107902765 | Intron | SBS | G | C |
| INDIA 8553 | SBSPON | chr8 | 73069709 | Intron | SBS | G | C |
| INDIA 8553 | Unknown | chr12 | 109323679 | IGR | SBS | G | C |
| INDIA 8553 | Unknown | chr1 | 178595706 | IGR | SBS | T | G |
| INDIA 8553 | Unknown | chr2 | 191547335 | IGR | SBS | G | C |
| INDIA 8553 | Unknown | chr4 | 155100309 | IGR | SBS | C | A |
| INDIA 8553 | ATXN1 | chr6 | 16485734 | Intron | SBS | G | C |
| INDIA 8553 | Unknown | chr7 | 53305261 | IGR | SBS | C | G |
| INDIA 8553 | ENST00000688408 | chr22 | 40673405 | Intron | SBS | G | C |
| INDIA 8553 | ATXN10 | chr22 | 45819339 | Intron | SBS | C | G |
| INDIA 8553 | LPIN1 | chr2 | 11711136 | Intron | SBS | C | G |
| INDIA 8553 | Unknown | chr13 | 68471418 | IGR | SBS | C | G |
| INDIA 8553 | LGR4 | chr11 | 27400936 | Intron | SBS | C | G |
| INDIA 8553 | DIRC3 | chr2 | 217431740 | Intron | SBS | C | G |
| INDIA 8553 | MAP4K3-DT | chr2 | 39590896 | Intron | DEL | A | - |
| INDIA 8553 | FUT8 | chr14 | 65488269 | Intron | DEL | T | - |
| INDIA 8553 | LINC01788 | chr1 | 70713469 | Intron | SBS | A | T |
| INDIA 8553 | MYOF | chr10 | 93364108 | Intron | SBS | G | C |
| INDIA 8553 | PRKCB | chr16 | 24203663 | Intron | SBS | C | G |
| INDIA 8553 | Unknown | chr17 | 52552055 | IGR | SBS | C | G |
| INDIA 8553 | Unknown | chr18 | 53985368 | IGR | SBS | G | C |
| INDIA 8553 | KCNJ6 | chr21 | 37682000 | Intron | SBS | G | T |
| INDIA 8554 | GALNT9 | chr12 | 132259081 | Intron | DEL | GAG | - |
| INDIA 8554 | PAPPA | chr9 | 116211336 | Intron | SBS | G | C |
| INDIA 8554 | TMEFF1 | chr9 | 100571095 | Intron | SBS | T | A |
| INDIA 8554 | LINC01010 | chr6 | 134358432 | Intron | SBS | C | G |
| INDIA 8554 | SOX3 | chrX | 140507540 | 5'Flank | SBS | C | G |
| INDIA 8554 | FGF13 | chrX | 139020688 | Intron | SBS | A | T |
| INDIA 8554 | EDA | chrX | 69746556 | Intron | DEL | CC | - |
| INDIA 8554 | IL1RAPL1 | chrX | 29747828 | Intron | SBS | T | A |
| INDIA 8554 | LINC00971 | chr3 | 84741111 | Intron | SBS | C | A |
| INDIA 8554 | Unknown | chr8 | 34920126 | IGR | SBS | T | A |
| INDIA 8554 | ELAVL4 | chr1 | 50047868 | 5'Flank | SBS | G | C |
| INDIA 8554 | Unknown | chr6 | 33751989 | IGR | SBS | C | G |
| INDIA 8554 | NAV2 | chr11 | 19642157 | Intron | SBS | T | G |
| INDIA 8554 | DNAJC10 | chr2 | 182795004 | 3'Flank | SBS | G | C |
| INDIA 8554 | TMEM132D | chr12 | 129819249 | Intron | SBS | A | T |
| INDIA 8554 | CLYBL | chr13 | 99724918 | Intron | SBS | G | C |
| INDIA 8554 | Unknown | chr2 | 60155012 | IGR | SBS | C | G |
| INDIA 8554 | Unknown | chr5 | 3772908 | IGR | SBS | T | A |
| INDIA 8554 | GRIK2 | chr6 | 101464995 | Intron | SBS | A | T |
| INDIA 8554 | NR1H4 | chr12 | 100491310 | Intron | SBS | G | T |
| INDIA 8554 | PLAC1 | chrX | 134562491 | 3'Flank | SBS | C | A |
| INDIA 8554 | LINC01358 | chr1 | 59112251 | Intron | DEL | A | - |
| INDIA 8554 | CSMD1 | chr8 | 3145186 | Intron | SBS | A | T |
| INDIA 8554 | ENST00000664407 | chr1 | 177830632 | Intron | SBS | G | C |
| INDIA 8554 | CDH12 | chr5 | 22227954 | Intron | DEL | C | - |
| INDIA 8554 | Unknown | chr4 | 180862867 | IGR | SBS | C | G |
| INDIA 8554 | Unknown | chr18 | 5355189 | IGR | DEL | G | - |
| INDIA 8554 | ENST00000647772 | chr6 | 92119138 | Intron | SBS | T | G |
| INDIA 8554 | Unknown | chr10 | 106254360 | IGR | SBS | C | G |
| INDIA 8554 | MRTFA | chr22 | 40441210 | Intron | DEL | G | - |
| INDIA 8554 | WWOX | chr16 | 78583741 | Intron | SBS | G | C |
| INDIA 8554 | COG3 | chr13 | 45529889 | Missense_Mutation | SBS | A | T |
| INDIA 8554 | PLCH1 | chr3 | 155747997 | 5'Flank | DEL | G | - |
| INDIA 8554 | RBFOX1 | chr16 | 6321540 | Intron | SBS | C | A |
| INDIA 8554 | Unknown | chr2 | 83645685 | IGR | SBS | A | T |
| INDIA 8554 | NALCN | chr13 | 101288531 | Intron | SBS | C | G |
| INDIA 8554 | CHTF8 | chr16 | 69116102 | 3'Flank | DEL | AAG | - |
| INDIA 8554 | KDM2B | chr12 | 121523355 | Intron | DEL | G | - |
| INDIA 8554 | APTX | chr9 | 32888509 | Intron | SBS | G | C |
| INDIA 8554 | PPARGC1B | chr5 | 149831814 | Intron | DEL | GA | - |

| | | | | | | | |
|---|---|---|---|---|---|---|---|
| INDIA 8554 | EPSTI1 | chr13 | 42906554 | Intron | SBS | C | G |
| INDIA 8554 | Unknown | chr8 | 34667839 | IGR | SBS | T | A |
| INDIA 8554 | Unknown | chr3 | 55430127 | IGR | SBS | G | T |
| INDIA 8554 | EPHA3 | chr3 | 89208697 | Intron | SBS | A | T |
| INDIA 8554 | Unknown | chr5 | 80372954 | IGR | SBS | A | T |
| INDIA 8554 | LINC02055 | chr8 | 135868688 | Intron | SBS | C | G |
| INDIA 8554 | Unknown | chr4 | 27828278 | IGR | SBS | C | G |
| INDIA 8554 | PLCE1 | chr10 | 94066544 | Intron | SBS | T | G |
| INDIA 8554 | Unknown | chr4 | 54550692 | IGR | SBS | G | C |
| INDIA 8554 | Unknown | chr19 | 32568494 | IGR | SBS | T | A |
| INDIA 8554 | ENST00000659397 | chr9 | 89994717 | Intron | DEL | A | - |
| INDIA 8554 | LINC02241 | chr5 | 20688159 | Intron | SBS | G | C |
| INDIA 8554 | MACROD2 | chr20 | 14915848 | Intron | DEL | TGA | - |
| INDIA 8554 | TAFA5 | chr22 | 48596341 | Intron | SBS | G | C |
| INDIA 8554 | Unknown | chr4 | 117893934 | IGR | SBS | A | C |
| INDIA 8554 | NRXN1 | chr2 | 51035431 | 5'Flank | SBS | G | C |
| INDIA 8554 | NRXN1 | chr2 | 51035433 | 5'Flank | SBS | T | A |
| INDIA 8554 | Unknown | chr4 | 59646046 | IGR | SBS | A | T |
| INDIA 8554 | MAGI1 | chr3 | 65969138 | Intron | SBS | A | C |
| INDIA 8554 | Unknown | chr5 | 19171113 | IGR | SBS | A | T |
| INDIA 8554 | LINC01519 | chr10 | 85199018 | 5'Flank | SBS | T | G |
| INDIA 8554 | Unknown | chr1 | 191456568 | IGR | SBS | T | A |
| INDIA 8554 | SLC35F3 | chr1 | 234231646 | Missense_Mutation | SBS | G | C |
| INDIA 8554 | Unknown | chr12 | 48541728 | IGR | SBS | C | A |
| INDIA 8554 | RBFOX3 | chr17 | 79323359 | Intron | SBS | G | C |
| INDIA 8554 | RBFOX3 | chr17 | 79323360 | Intron | SBS | G | T |
| INDIA 8554 | Unknown | chr15 | 97161767 | IGR | SBS | C | G |
| INDIA 8554 | AGA-DT | chr4 | 177561615 | Intron | DEL | T | - |
| INDIA 8554 | Unknown | chr13 | 64926781 | IGR | SBS | A | T |
| INDIA 8554 | Unknown | chr9 | 29912883 | IGR | SBS | T | G |
| INDIA 8554 | ENST00000669750 | chr1 | 170891952 | Intron | DEL | C | - |
| INDIA 8554 | DCX | chrX | 111363971 | Intron | SBS | G | C |
| INDIA 8554 | AAGAB | chr15 | 67215228 | Intron | SBS | G | C |
| INDIA 8554 | Unknown | chr7 | 118975242 | IGR | SBS | C | G |
| INDIA 8554 | PTF1A | chr10 | 23194741 | 3'Flank | SBS | G | C |
| INDIA 8554 | Unknown | chr3 | 193800600 | IGR | SBS | G | C |
| INDIA 8554 | Unknown | chr14 | 47825643 | IGR | SBS | G | C |
| INDIA 8554 | NTN1 | chr17 | 9246676 | 3'Flank | SBS | C | A |
| INDIA 8554 | MAGI2 | chr7 | 78437209 | Intron | SBS | C | A |
| INDIA 8554 | SORCS1 | chr10 | 107032630 | Intron | SBS | T | A |
| INDIA 8554 | Unknown | chr1 | 239334286 | IGR | SBS | C | A |
| INDIA 8554 | Unknown | chr10 | 36343878 | IGR | DEL | CCAAGCCAG | - |
| INDIA 8554 | VRK2 | chr2 | 58141855 | Intron | SBS | G | C |
| INDIA 8554 | Unknown | chr2 | 198838331 | IGR | SBS | G | C |
| INDIA 8554 | Unknown | chr3 | 117413870 | IGR | SBS | T | G |
| INDIA 8554 | CSMD3 | chr8 | 113321410 | Intron | SBS | C | G |
| INDIA 8554 | ENST00000646051 | chr10 | 71013355 | Intron | SBS | G | C |
| INDIA 8554 | CD300C | chr17 | 74543678 | Intron | SBS | G | T |
| INDIA 8554 | Unknown | chr3 | 167226403 | IGR | SBS | T | A |
| INDIA 8554 | NDST4 | chr4 | 114972695 | Intron | SBS | A | T |
| INDIA 8554 | ENST00000515232 | chr4 | 153059519 | Intron | DEL | TTC | - |
| INDIA 8555 | LINC01828 | chr2 | 67219895 | Intron | SBS | A | C |
| INDIA 8555 | ENST00000636261 | chr5 | 159536544 | Intron | SBS | G | C |
| INDIA 8555 | Unknown | chr1 | 208394623 | IGR | SBS | C | G |
| INDIA 8555 | Unknown | chr4 | 44514669 | IGR | SBS | G | C |
| INDIA 8555 | Unknown | chr15 | 96689008 | IGR | SBS | G | C |
| INDIA 8555 | TTLL11 | chr9 | 121813349 | 3'Flank | SBS | G | C |
| INDIA 8555 | Unknown | chr2 | 176182 | IGR | SBS | C | G |
| INDIA 8555 | Unknown | chr6 | 85402747 | IGR | SBS | G | C |
| INDIA 8555 | GORAB-AS1 | chr1 | 170471650 | Intron | SBS | C | A |
| INDIA 8555 | HEATR1 | chr1 | 236594410 | Intron | SBS | A | C |
| INDIA 8555 | LINC01010 | chr6 | 134411754 | Intron | SBS | C | G |
| INDIA 8555 | BTBD11 | chr12 | 107407875 | Intron | SBS | C | G |
| INDIA 8555 | RNU4ATAC7P | chr20 | 54859883 | 5'Flank | SBS | G | C |
| INDIA 8555 | RRP12 | chr10 | 97380558 | Intron | SBS | C | G |
| INDIA 8555 | TUT1 | chr11 | 62587986 | Intron | SBS | C | G |

| | | | | | | | |
|---|---|---|---|---|---|---|---|
| INDIA 8555 | SOX2-OT | chr3 | 181117129 | Intron | SBS | G | C |
| INDIA 8555 | NTM | chr11 | 132234555 | Intron | SBS | C | G |
| INDIA 8555 | MSI2 | chr17 | 57410391 | Intron | SBS | G | T |
| INDIA 8555 | ZIM2 | chr19 | 56776872 | Intron | SBS | G | C |
| INDIA 8555 | ADCY2 | chr5 | 7805642 | Intron | SBS | C | G |
| INDIA 8555 | FMN2 | chr1 | 240323612 | Intron | SBS | C | G |
| INDIA 8555 | NRG3 | chr10 | 82502092 | Intron | SBS | G | C |
| INDIA 8555 | ZBTB14 | chr18 | 5292991 | Intron | SBS | G | C |
| INDIA 8555 | PRRX2 | chr9 | 129696649 | Intron | SBS | G | C |
| INDIA 8555 | Unknown | chr5 | 32836511 | IGR | SBS | C | G |
| INDIA 8555 | Unknown | chr19 | 20050890 | IGR | SBS | G | C |
| INDIA 8555 | LHFPL3 | chr7 | 104851765 | Intron | SBS | G | C |
| INDIA 8555 | Unknown | chr7 | 109845280 | IGR | SBS | C | G |
| INDIA 8555 | TCF15 | chr20 | 605590 | Intron | SBS | C | G |
| INDIA 8555 | PAK3 | chrX | 111038380 | Intron | SBS | C | G |
| INDIA 8555 | CD226 | chr18 | 69895914 | Missense_Mutation | SBS | G | C |
| INDIA 8555 | Unknown | chr10 | 29189017 | IGR | SBS | G | C |
| INDIA 8555 | PDE1C | chr7 | 31897244 | Intron | SBS | C | G |
| INDIA 8555 | KPNB1 | chr17 | 47676161 | Intron | SBS | G | C |
| INDIA 8555 | Unknown | chr8 | 103104187 | IGR | SBS | C | G |
| INDIA 8555 | LINC01749 | chr20 | 63070454 | Intron | SBS | C | G |
| INDIA 8555 | EIPR1 | chr2 | 3269160 | Intron | SBS | C | G |
| INDIA 8555 | CSRNP3 | chr2 | 165533505 | Intron | SBS | C | G |
| INDIA 8555 | Unknown | chr20 | 56802062 | IGR | SBS | C | G |
| INDIA 8555 | SNX7 | chr1 | 98763916 | 3'Flank | SBS | G | C |
| INDIA 8555 | CACNA1E | chr1 | 181643953 | Intron | SBS | C | G |
| INDIA 8555 | PCDHGA6 | chr5 | 141376378 | Missense_Mutation | SBS | G | C |
| INDIA 8555 | CCDC120 | chrX | 49057186 | 5'Flank | SBS | C | G |
| INDIA 8555 | TUBB8B | chr18 | 53814 | 5'Flank | SBS | G | C |
| INDIA 8555 | Unknown | chr19 | 31638868 | IGR | SBS | C | G |
| INDIA 8555 | JAZF1 | chr7 | 27958609 | Intron | SBS | C | G |
| INDIA 8555 | LINC02730 | chr11 | 134017866 | Intron | SBS | C | G |
| INDIA 8555 | ENST00000649588 | chr3 | 135356170 | Intron | SBS | C | G |
| INDIA 8555 | CATSPERE | chr1 | 244622142 | Intron | SBS | G | C |
| INDIA 8555 | ENST00000512715 | chr4 | 143301153 | Intron | SBS | G | C |
| INDIA 8555 | ENST00000495817 | chr3 | 180828481 | Intron | DEL | A | - |
| INDIA 8555 | Unknown | chr6 | 103404230 | IGR | SBS | C | G |
| INDIA 8555 | KY | chr3 | 134619923 | Intron | SBS | C | G |
| INDIA 8555 | TSTD3 | chr6 | 99529794 | Intron | SBS | C | G |
| INDIA 8555 | MGAT5 | chr2 | 134426791 | Intron | SBS | G | C |
| INDIA 8555 | NMNAT3 | chr3 | 139661301 | Intron | SBS | C | G |
| INDIA 8555 | SHTN1 | chr10 | 116939661 | Intron | SBS | C | G |
| INDIA 8555 | TFEC | chr7 | 116023845 | Intron | SBS | G | C |
| INDIA 8555 | Unknown | chr11 | 124717876 | IGR | SBS | C | G |
| INDIA 8555 | GLIS3 | chr9 | 3929466 | Intron | SBS | C | G |
| INDIA 8555 | SUV39H2 | chr10 | 14877249 | 5'Flank | SBS | G | C |
| INDIA 8555 | FBLN5 | chr14 | 91872371 | Intron | SBS | G | C |
| INDIA 8555 | MAP3K13 | chr3 | 185423949 | Intron | SBS | C | G |
| INDIA 8555 | IKZF3 | chr17 | 39767508 | Intron | SBS | G | C |
| INDIA 8555 | ENST00000423712 | chr22 | 34698439 | 3'Flank | SBS | C | G |
| INDIA 8555 | TPM3 | chr1 | 154167385 | 3'UTR | SBS | G | C |
| INDIA 8555 | PARL | chr3 | 183829500 | 3'UTR | SBS | C | G |
| INDIA 8555 | Unknown | chr7 | 74000432 | IGR | SBS | C | G |
| INDIA 8555 | XKR4 | chr8 | 55172837 | Intron | SBS | C | G |
| INDIA 8555 | Unknown | chr11 | 128188966 | IGR | SBS | G | C |
| INDIA 8555 | HERC1 | chr15 | 63627089 | Intron | SBS | C | G |
| INDIA 8555 | SNTG2 | chr2 | 1278364 | Intron | SBS | G | C |
| INDIA 8555 | MAML3 | chr4 | 139923847 | Intron | SBS | G | C |
| INDIA 8555 | KCNMA1 | chr10 | 76989375 | Intron | SBS | C | G |
| INDIA 8555 | TMED10 | chr14 | 75158119 | Intron | SBS | G | C |
| INDIA 8555 | CEP192 | chr18 | 13032243 | Intron | SBS | G | C |
| INDIA 8555 | Unknown | chr15 | 50781929 | IGR | SBS | G | C |
| INDIA 8555 | Unknown | chr15 | 50781934 | IGR | SBS | G | C |
| INDIA 8555 | RAB4A | chr1 | 229268770 | 5'Flank | SBS | C | G |
| INDIA 8555 | MAP3K9 | chr14 | 70733250 | Missense_Mutation | SBS | C | G |
| INDIA 8555 | OTULIN | chr5 | 14679445 | Intron | SBS | C | G |

| | | | | | | | |
|---|---|---|---|---|---|---|---|
| INDIA 8555 | KIAA1671 | chr22 | 25114174 | Intron | SBS | C | G |
| INDIA 8555 | ANO4 | chr12 | 101101600 | Intron | SBS | G | C |
| INDIA 8555 | NFIA-AS2 | chr1 | 60972626 | Intron | SBS | C | G |
| INDIA 8555 | Unknown | chr7 | 106080229 | IGR | SBS | C | G |
| INDIA 8555 | FLI1 | chr11 | 128707359 | Intron | SBS | C | G |
| INDIA 8555 | Unknown | chr18 | 22106692 | IGR | SBS | G | C |
| INDIA 8555 | Unknown | chr20 | 30600080 | IGR | SBS | C | G |
| INDIA 8555 | INTS6L | chrX | 135551244 | Intron | SBS | C | G |
| INDIA 8555 | SENP2 | chr3 | 185582972 | 5'Flank | SBS | C | G |
| INDIA 8555 | ENST00000457577 | chr2 | 199862710 | 3'Flank | SBS | G | C |
| INDIA 8555 | PRKG1 | chr10 | 51412324 | Intron | SBS | C | G |
| INDIA 8556 | Unknown | chr6 | 18983986 | IGR | SBS | C | G |
| INDIA 8556 | EGFR | chr7 | 55113776 | Intron | SBS | C | G |
| INDIA 8556 | Unknown | chr11 | 20544538 | IGR | SBS | C | G |
| INDIA 8556 | CD74 | chr5 | 150397457 | 3'Flank | SBS | G | C |
| INDIA 8556 | BICDL1 | chr12 | 119991793 | Intron | SBS | A | C |
| INDIA 8556 | CAP2 | chr6 | 17419377 | Intron | SBS | T | A |
| INDIA 8556 | TLCD2 | chr17 | 1707224 | 3'UTR | SBS | A | T |
| INDIA 8556 | ENST00000527270 | chr11 | 33805407 | 5'Flank | SBS | G | C |
| INDIA 8556 | Unknown | chr5 | 71903649 | IGR | SBS | C | G |
| INDIA 8556 | DLGAP1 | chr18 | 3618326 | Intron | SBS | G | C |
| INDIA 8556 | PTPN13 | chr4 | 86642200 | Intron | SBS | C | G |
| INDIA 8556 | BBS9 | chr7 | 33308493 | Intron | INS | - | AA |
| INDIA 8556 | CDH12 | chr5 | 22305239 | Intron | SBS | C | G |
| INDIA 8556 | Unknown | chrX | 131982098 | IGR | SBS | A | T |
| INDIA 8556 | Unknown | chr6 | 151633694 | IGR | SBS | C | G |
| INDIA 8556 | P2RY12 | chr3 | 151351341 | Intron | DEL | T | - |
| INDIA 8556 | APCDD1L-DT | chr20 | 58618316 | Intron | SBS | C | G |
| INDIA 8556 | CASZ1 | chr1 | 10760764 | 5'UTR | SBS | G | C |
| INDIA 8556 | CACHD1 | chr1 | 64472090 | Intron | SBS | G | C |
| INDIA 8556 | ME3 | chr11 | 86490458 | Intron | SBS | C | G |
| INDIA 8556 | CLVS1 | chr8 | 61462083 | Intron | SBS | G | C |
| INDIA 8556 | GNG4 | chr1 | 235546158 | 3'Flank | SBS | C | G |
| INDIA 8556 | KAT6A | chr8 | 42054820 | 5'Flank | SBS | G | C |
| INDIA 8556 | NLRP12 | chr19 | 53795957 | Missense_Mutation | SBS | C | G |
| INDIA 8556 | LBR | chr1 | 225428967 | 5'Flank | SBS | G | C |
| INDIA 8556 | SPRED2 | chr2 | 65375488 | Intron | SBS | G | C |
| INDIA 8556 | RPL32 | chr3 | 12837492 | Intron | SBS | A | C |
| INDIA 8556 | ENST00000420648 | chr2 | 78593050 | 5'Flank | SBS | G | C |
| INDIA 8556 | LRRC4B | chr19 | 50531618 | Intron | DEL | C | - |
| INDIA 8556 | Unknown | chr2 | 234607940 | IGR | SBS | C | G |
| INDIA 8556 | Unknown | chr13 | 44287031 | IGR | SBS | G | C |
| INDIA 8556 | Unknown | chrX | 9308733 | IGR | SBS | C | G |
| INDIA 8556 | PAK3 | chrX | 111113386 | Intron | SBS | A | C |
| INDIA 8556 | DST | chr6 | 56765642 | Intron | SBS | G | C |
| INDIA 8556 | LINC01435 | chr10 | 108071112 | 5'Flank | DEL | CT | - |
| INDIA 8556 | NOL4L | chr20 | 32454125 | Intron | SBS | C | G |
| INDIA 8556 | Unknown | chr5 | 153374911 | IGR | SBS | C | G |
| INDIA 8556 | ENST00000669768 | chr11 | 112831177 | Intron | SBS | C | A |
| INDIA 8556 | THEM7P | chr11 | 32300334 | Intron | SBS | C | A |
| INDIA 8557 | Unknown | chr7 | 22369548 | IGR | SBS | C | G |
| INDIA 8557 | MIR548I4 | chrX | 84223017 | 3'Flank | SBS | T | A |
| INDIA 8557 | Unknown | chr7 | 46775764 | IGR | SBS | C | G |
| INDIA 8557 | CAMK2B | chr7 | 44231129 | Intron | SBS | G | C |
| INDIA 8557 | SOX6 | chr11 | 16460755 | Intron | SBS | G | C |
| INDIA 8557 | Unknown | chr14 | 87207434 | IGR | SBS | C | A |
| INDIA 8557 | Unknown | chr22 | 31919273 | IGR | SBS | T | G |
| INDIA 8557 | Unknown | chr6 | 106898622 | IGR | SBS | G | C |
| INDIA 8557 | PAPPA2 | chr1 | 176754978 | Intron | SBS | C | G |
| INDIA 8557 | Unknown | chr2 | 123492137 | IGR | DEL | C | - |
| INDIA 8557 | ELOVL6 | chr4 | 110068653 | Intron | SBS | A | C |
| INDIA 8557 | Unknown | chr10 | 23673309 | IGR | SBS | G | C |
| INDIA 8557 | NUP93 | chr16 | 56833499 | Intron | SBS | G | C |
| INDIA 8557 | ENST00000557745 | chr14 | 97120620 | Intron | SBS | C | G |
| INDIA 8557 | Unknown | chr8 | 34156155 | IGR | SBS | G | T |
| INDIA 8557 | TOMM34 | chr20 | 44939200 | 3'Flank | SBS | C | G |

| | | | | | | | |
|---|---|---|---|---|---|---|---|
| INDIA 8557 | Unknown | chr16 | 86628650 | IGR | SBS | T | A |
| INDIA 8557 | CHMP4C | chr8 | 81754002 | Intron | SBS | T | A |
| INDIA 8557 | AOAH | chr7 | 36522746 | Intron | SBS | C | G |
| INDIA 8557 | Unknown | chr7 | 57354501 | IGR | DEL | G | - |
| INDIA 8557 | CHIA | chr1 | 111289411 | 5'Flank | INS | - | G |
| INDIA 8557 | MARCHF5 | chr10 | 92356016 | 3'Flank | SBS | G | C |
| INDIA 8557 | Unknown | chr21 | 20216012 | IGR | SBS | A | T |
| INDIA 8557 | ANK2 | chr4 | 113226001 | Intron | SBS | A | T |
| INDIA 8557 | Unknown | chr6 | 143417408 | IGR | SBS | G | C |
| INDIA 8557 | Unknown | chr7 | 8758373 | IGR | SBS | G | C |
| INDIA 8557 | SSR1 | chr6 | 7277918 | 3'Flank | INS | - | T |
| INDIA 8557 | GALNTL6 | chr4 | 172173339 | Intron | SBS | G | T |
| INDIA 8557 | POU6F2 | chr7 | 39332998 | Intron | DEL | G | - |
| INDIA 8557 | Unknown | chr12 | 127857564 | IGR | DEL | A | - |
| INDIA 8557 | DAPP1 | chr4 | 99855323 | Intron | SBS | C | G |
| INDIA 8557 | Unknown | chr22 | 33996003 | IGR | SBS | T | A |
| INDIA 8557 | Unknown | chr3 | 5304880 | IGR | DEL | TG | - |
| INDIA 8557 | Unknown | chr6 | 103703656 | IGR | SBS | G | C |
| INDIA 8557 | SGMS1 | chr10 | 50516300 | Intron | SBS | G | C |
| INDIA 8557 | NRG3 | chr10 | 82275404 | Intron | SBS | T | A |
| INDIA 8557 | TMEM178A | chr2 | 39722684 | 3'Flank | SBS | A | C |
| INDIA 8557 | GPR157 | chr1 | 9109919 | Intron | SBS | G | C |
| INDIA 8557 | MED27 | chr9 | 131886322 | Intron | DEL | G | - |
| INDIA 8557 | Unknown | chr7 | 48999917 | IGR | SBS | C | A |
| INDIA 8557 | ENST00000637606 | chr9 | 82321056 | Intron | SBS | C | G |
| INDIA 8557 | DIAPH2 | chrX | 97152232 | Intron | SBS | G | C |
| INDIA 8557 | NTF3 | chr12 | 5476061 | Intron | INS | - | T |
| INDIA 8557 | Unknown | chr6 | 139889069 | IGR | SBS | C | G |
| INDIA 8557 | LINC02411 | chr12 | 127638002 | 5'Flank | SBS | G | C |
| INDIA 8557 | SLC8A1 | chr2 | 40169166 | Intron | DEL | A | - |
| INDIA 8557 | CFAP54 | chr12 | 96753862 | Frame_Shift_Ins | INS | - | A |
| INDIA 8557 | Unknown | chr10 | 113314704 | IGR | SBS | G | C |
| INDIA 8557 | ENST00000545853 | chr12 | 126637246 | Intron | SBS | C | G |
| INDIA 8557 | ENST00000659958 | chr17 | 33852056 | Intron | INS | - | T |
| INDIA 8557 | Unknown | chr8 | 59714795 | IGR | INS | - | T |
| INDIA 8557 | SLC8A3 | chr14 | 70094157 | Intron | SBS | C | G |
| INDIA 8557 | ETFBKMT | chr12 | 31658361 | 5'Flank | SBS | C | G |
| INDIA 8557 | Unknown | chr18 | 61133858 | IGR | SBS | G | C |
| INDIA 8557 | TMEM178B | chr7 | 141396045 | Intron | SBS | C | G |
| INDIA 8557 | ENST00000659524 | chr8 | 32967003 | Intron | SBS | G | T |
| INDIA 8557 | XXYLT1 | chr3 | 195076538 | Intron | SBS | C | G |
| INDIA 8557 | BBS9 | chr7 | 33308793 | Intron | SBS | C | G |
| INDIA 8557 | ATP6V0D2 | chr8 | 86148318 | Intron | INS | - | A |
| INDIA 8557 | Unknown | chr21 | 39963995 | IGR | SBS | C | G |
| INDIA 8557 | B4GALNT3 | chr12 | 510267 | Intron | SBS | C | G |
| INDIA 8557 | COPS7A | chr12 | 6725736 | Intron | SBS | G | C |
| INDIA 8557 | SLC8A3 | chr14 | 70155711 | Intron | SBS | C | A |
| INDIA 8557 | Unknown | chr11 | 130728572 | IGR | SBS | C | G |
| INDIA 8557 | DOCK2 | chr5 | 169912739 | Intron | SBS | T | A |
| INDIA 8557 | Unknown | chr20 | 12511702 | IGR | SBS | A | T |
| INDIA 8557 | LRRC8D | chr1 | 89842724 | Intron | SBS | A | C |
| INDIA 8557 | ESM1 | chr5 | 54994358 | Intron | SBS | G | C |
| INDIA 8557 | CDK5 | chr7 | 151055010 | Intron | SBS | G | C |
| INDIA 8557 | CCDC26 | chr8 | 129552761 | Intron | SBS | C | A |
| INDIA 8557 | LINC02183 | chr16 | 54547411 | Intron | SBS | C | G |
| INDIA 8557 | GLIS3 | chr9 | 3976523 | Intron | INS | - | A |
| INDIA 8557 | DEAF1 | chr11 | 691087 | Intron | SBS | C | G |
| INDIA 8557 | TRHDE | chr12 | 72564056 | Intron | SBS | A | C |
| INDIA 8557 | Unknown | chr4 | 129224187 | IGR | SBS | C | G |
| INDIA 8557 | Unknown | chr5 | 100308030 | IGR | INS | - | A |
| INDIA 8557 | SULF1 | chr8 | 69599072 | Intron | SBS | A | T |
| INDIA 8557 | ADAM12 | chr10 | 126391817 | 5'Flank | SBS | C | A |
| INDIA 8557 | ROBO2 | chr3 | 77168266 | Intron | SBS | C | G |
| INDIA 8557 | ATP13A5 | chr3 | 193332683 | Intron | SBS | G | C |
| INDIA 8557 | NBAT1 | chr6 | 22133920 | RNA | SBS | C | G |
| INDIA 8557 | PLCE1 | chr10 | 94324841 | Intron | SBS | G | C |

| | | | | | | | |
|---|---|---|---|---|---|---|---|
| INDIA 8557 | Unknown | chr16 | 64184457 | IGR | SBS | C | G |
| INDIA 8557 | SPOCK1 | chr5 | 137096537 | Intron | SBS | G | C |
| INDIA 8557 | FAM234B | chr12 | 13087818 | 3'Flank | SBS | C | G |
| INDIA 8557 | FOXF1 | chr16 | 86515657 | 3'Flank | SBS | C | A |
| INDIA 8557 | Unknown | chr14 | 86718390 | IGR | DEL | C | - |
| INDIA 8557 | POU2AF1 | chr11 | 111358077 | Intron | SBS | C | G |
| INDIA 8557 | SLC16A7 | chr12 | 59747545 | Intron | SBS | A | T |
| INDIA 8557 | EPHA7 | chr6 | 93366720 | Intron | SBS | C | G |
| INDIA 8557 | PKNOX2 | chr11 | 125193810 | Intron | DEL | G | - |
| INDIA 8557 | Unknown | chr5 | 123908884 | IGR | INS | - | A |
| INDIA 8557 | THOP1 | chr19 | 2791219 | Intron | INS | - | T |
| INDIA 8559 | Unknown | chr1 | 112083610 | IGR | SBS | T | A |
| INDIA 8559 | EGFLAM | chr5 | 38296102 | Intron | SBS | G | T |
| INDIA 8559 | HIVEP1 | chr6 | 12115656 | Intron | SBS | G | C |
| INDIA 8559 | Unknown | chr5 | 135435815 | IGR | SBS | C | G |
| INDIA 8559 | ROR1 | chr1 | 64080838 | Intron | SBS | C | A |
| INDIA 8559 | WNT5B | chr12 | 1527885 | 5'Flank | SBS | G | C |
| INDIA 8559 | Unknown | chr1 | 41411321 | IGR | SBS | G | C |
| INDIA 8559 | NTRK2 | chr9 | 84739763 | Intron | SBS | G | T |
| INDIA 8559 | SAE1 | chr19 | 47176650 | Intron | SBS | G | C |
| INDIA 8559 | GIMAP6 | chr7 | 150627051 | 3'UTR | DEL | T | - |
| INDIA 8559 | HRH1 | chr3 | 11155804 | Intron | SBS | C | G |
| INDIA 8559 | HCRTR2 | chr6 | 55251624 | Intron | SBS | C | G |
| INDIA 8559 | ZNF704 | chr8 | 80723973 | Intron | SBS | C | G |
| INDIA 8559 | Unknown | chr20 | 60997295 | IGR | SBS | C | G |
| INDIA 8559 | ASIC2 | chr17 | 33554645 | Intron | SBS | G | C |
| INDIA 8559 | ANO2 | chr12 | 5874231 | Intron | SBS | G | C |
| INDIA 8559 | ENST00000676095 | chr2 | 190840135 | Intron | SBS | A | C |
| INDIA 8559 | LINC00375 | chr13 | 85075107 | RNA | SBS | C | G |
| INDIA 8559 | Unknown | chr2 | 68879433 | IGR | SBS | C | G |
| INDIA 8559 | Unknown | chrX | 63921066 | IGR | SBS | T | G |
| INDIA 8559 | HBQ1 | chr16 | 183281 | 3'Flank | SBS | G | C |
| INDIA 8559 | Unknown | chr2 | 160893859 | IGR | SBS | G | C |
| INDIA 8559 | ENST00000559755 | chr15 | 100716275 | RNA | SBS | G | C |
| INDIA 8559 | SH3GL2 | chr9 | 17690606 | Intron | DEL | TG | - |
| INDIA 8559 | Unknown | chr1 | 214261770 | IGR | DEL | CCGG | - |
| INDIA 8559 | C1orf116 | chr1 | 207029158 | Intron | SBS | G | C |
| INDIA 8559 | CAMK2A | chr5 | 150218341 | 3'Flank | SBS | A | C |
| INDIA 8559 | PRICKLE1 | chr12 | 42561141 | Intron | SBS | A | T |
| INDIA 8559 | INHBB | chr2 | 120348436 | Intron | SBS | G | C |
| INDIA 8559 | MIR646HG | chr20 | 60589181 | Intron | SBS | C | G |
| INDIA 8559 | ST6GALNAC3 | chr1 | 76133341 | Intron | SBS | G | C |
| INDIA 8559 | VTI1A | chr10 | 112776428 | Intron | SBS | G | C |
| INDIA 8559 | TSHZ2 | chr20 | 53176418 | Intron | SBS | G | C |
| INDIA 8559 | MAD1L1 | chr7 | 2169117 | Intron | DEL | GGACAAGCG | - |
| INDIA 8559 | Unknown | chr1 | 115798280 | IGR | SBS | T | A |
| INDIA 8559 | Unknown | chr11 | 111186088 | IGR | SBS | G | C |
| INDIA 8559 | Unknown | chr3 | 99562646 | IGR | SBS | T | G |
| INDIA 8559 | GRM1 | chr6 | 146103771 | Intron | SBS | C | G |
| INDIA 8559 | ETV6 | chr12 | 11705800 | Intron | SBS | C | G |
| INDIA 8559 | ANKRD6 | chr6 | 89577236 | Intron | SBS | G | C |
| INDIA 8559 | GPA33 | chr1 | 167081435 | Intron | SBS | C | G |
| INDIA 8559 | TTC7B | chr14 | 90562357 | Intron | SBS | C | G |
| INDIA 8559 | KIRREL1 | chr1 | 157994215 | Intron | SBS | A | T |
| INDIA 8559 | C16orf96 | chr16 | 4561447 | Intron | SBS | G | C |
| INDIA 8559 | DNAJC16 | chr1 | 15525380 | 5'Flank | SBS | C | G |
| INDIA 8559 | AOAH | chr7 | 36594682 | Intron | SBS | C | G |
| INDIA 8559 | OR10G9 | chr11 | 124020529 | 5'Flank | SBS | G | C |
| INDIA 8559 | Unknown | chrX | 28030207 | IGR | INS | - | A |
| INDIA 8559 | Unknown | chrX | 140663219 | IGR | SBS | G | C |
| INDIA 8559 | PM20D1 | chr1 | 205844713 | Intron | SBS | G | C |
| INDIA 8559 | EPHB1 | chr3 | 135167035 | Intron | SBS | G | C |
| INDIA 8559 | Unknown | chr3 | 26761547 | IGR | SBS | C | A |
| INDIA 8559 | RAP1GAP2 | chr17 | 2903138 | Intron | SBS | C | G |
| INDIA 8559 | RPTOR | chr17 | 80948591 | Intron | SBS | G | C |
| INDIA 8559 | Unknown | chr1 | 178605398 | IGR | SBS | G | C |

| | | | | | | | |
|---|---|---|---|---|---|---|---|
| INDIA 8559 | VIT | chr2 | 36737825 | Intron | SBS | G | C |
| INDIA 8559 | Unknown | chr16 | 54753357 | IGR | DEL | C | - |
| INDIA 8559 | Unknown | chr4 | 45933529 | IGR | SBS | G | C |
| INDIA 8559 | Unknown | chr5 | 75993734 | IGR | SBS | C | G |
| INDIA 8559 | Unknown | chr9 | 32615235 | IGR | SBS | G | C |
| INDIA 8559 | CFAP57 | chr1 | 43227433 | Intron | SBS | T | G |
| INDIA 8559 | BTRC | chr10 | 101535451 | Missense_Mutation | SBS | G | C |
| INDIA 8559 | ACOXL | chr2 | 111047330 | Intron | SBS | G | C |
| INDIA 8559 | ENST00000634280 | chr20 | 22410873 | Intron | SBS | G | C |
| INDIA 8559 | FBXW11 | chr5 | 172002117 | Intron | SBS | G | T |
| INDIA 8559 | Unknown | chr9 | 29499165 | IGR | SBS | T | A |
| INDIA 8559 | BDNF | chr11 | 27672425 | Intron | SBS | G | C |
| INDIA 8559 | SMARCA4 | chr19 | 11030542 | Intron | SBS | G | C |
| INDIA 8559 | MAP3K7CL | chr21 | 29088091 | Intron | SBS | C | G |
| INDIA 8559 | ATN1 | chr12 | 6924447 | 5'Flank | SBS | A | T |
| INDIA 8559 | MASP1 | chr3 | 187220356 | Intron | SBS | A | T |
| INDIA 8559 | ENST00000487657 | chr4 | 128579115 | 3'Flank | SBS | A | T |
| INDIA 8559 | SPATA18 | chr4 | 52051647 | 5'UTR | SBS | C | G |
| INDIA 8559 | Unknown | chr9 | 1118889 | IGR | SBS | G | C |
| INDIA 8559 | Unknown | chr8 | 135082895 | IGR | SBS | C | G |
| INDIA 8559 | JARID2 | chr6 | 15399284 | Intron | SBS | T | A |
| INDIA 8559 | FLJ46284 | chr8 | 92862151 | Intron | SBS | C | G |
| INDIA 8559 | PVT1 | chr8 | 128142148 | Intron | SBS | G | C |
| INDIA 8559 | RBFOX1 | chr16 | 6968058 | Intron | SBS | G | C |
| INDIA 8559 | NFATC2 | chr20 | 51484773 | Intron | SBS | C | G |
| INDIA 8559 | KIRREL3 | chr11 | 126483656 | Intron | SBS | G | T |
| INDIA 8559 | DIP2A | chr21 | 46462091 | Intron | SBS | G | C |
| INDIA 8559 | UNC13B | chr9 | 35389272 | Intron | SBS | G | C |
| INDIA 8559 | Unknown | chr6 | 106872212 | IGR | SBS | G | C |
| INDIA 8559 | FRMD6-AS2 | chr14 | 51552562 | Intron | DEL | CC | - |
| INDIA 8559 | SUSD4 | chr1 | 223269932 | Intron | SBS | G | C |
| INDIA 8559 | Unknown | chr2 | 236523875 | IGR | SBS | G | C |
| INDIA 8559 | PRPF6 | chr20 | 64028473 | Splice_Region | SBS | C | G |
| INDIA 8559 | CTNNA2 | chr2 | 80016427 | Intron | SBS | C | G |
| INDIA 8559 | Unknown | chr7 | 31134979 | IGR | SBS | C | G |
| INDIA 8560 | Unknown | chr11 | 106524728 | IGR | SBS | G | C |
| INDIA 8560 | Unknown | chr18 | 38696045 | IGR | DEL | C | - |
| INDIA 8560 | PTPRD | chr9 | 10076927 | Intron | SBS | C | G |
| INDIA 8560 | Unknown | chr13 | 44755045 | IGR | SBS | G | C |
| INDIA 8560 | SHB | chr9 | 38065251 | Intron | DEL | C | - |
| INDIA 8560 | ANKRD34C-AS1 | chr15 | 79161107 | Intron | SBS | G | C |
| INDIA 8560 | ENST00000669419 | chr2 | 234870143 | Intron | SBS | G | C |
| INDIA 8560 | UNC13C | chr15 | 54015208 | Missense_Mutation | SBS | G | C |
| INDIA 8560 | ENST00000432993 | chr2 | 220105132 | 5'Flank | SBS | C | G |
| INDIA 8560 | Unknown | chr11 | 115120660 | IGR | SBS | G | C |
| INDIA 8560 | CHN2 | chr7 | 29403049 | Intron | DEL | T | - |
| INDIA 8560 | ENST00000652518 | chr11 | 121804764 | Intron | SBS | A | T |
| INDIA 8560 | TUB | chr11 | 8103248 | 3'UTR | SBS | G | C |
| INDIA 8560 | TP53 | chr17 | 7674241 | Missense_Mutation | SBS | G | C |
| INDIA 8560 | Unknown | chr7 | 24021765 | IGR | SBS | A | T |
| INDIA 8560 | Unknown | chr13 | 106810212 | IGR | SBS | G | C |
| INDIA 8560 | MED26 | chr19 | 16631012 | 5'Flank | SBS | G | C |
| INDIA 8560 | NF2 | chr22 | 29671075 | Intron | SBS | G | C |
| INDIA 8560 | Unknown | chr2 | 15758239 | IGR | SBS | T | A |
| INDIA 8560 | LRP2 | chr2 | 169201810 | Missense_Mutation | SBS | G | C |
| INDIA 8560 | NALF1 | chr13 | 107720147 | Intron | DEL | CA | - |
| INDIA 8560 | INPP4B | chr4 | 142108652 | Intron | INS | - | A |
| INDIA 8560 | CSMD2 | chr1 | 33605363 | Missense_Mutation | SBS | A | T |
| INDIA 8560 | TBC1D22A | chr22 | 47125905 | Intron | SBS | G | C |
| INDIA 8560 | ANKRD28 | chr3 | 15676232 | Intron | SBS | C | G |
| INDIA 8560 | GPR83 | chr11 | 94393136 | Intron | SBS | G | C |
| INDIA 8560 | TAF1 | chrX | 71459915 | Intron | SBS | G | C |
| INDIA 8560 | LINC01865 | chr2 | 315040 | 5'Flank | SBS | T | A |
| INDIA 8560 | CTNND2 | chr5 | 11241321 | Intron | SBS | C | G |
| INDIA 8560 | Unknown | chr13 | 69078701 | IGR | SBS | T | A |
| INDIA 8560 | ENST00000642330 | chr9 | 69090034 | Intron | SBS | C | A |

| | | | | | | | |
|---|---|---|---|---|---|---|---|
| INDIA 8560 | CARMIL2 | chr16 | 67645679 | Intron | DEL | AAGAG | - |
| INDIA 8560 | HRG-AS1 | chr3 | 186599418 | Intron | SBS | C | G |
| INDIA 8560 | KCNMB2 | chr3 | 178407512 | Intron | SBS | C | G |
| INDIA 8560 | GPATCH8 | chr17 | 44504110 | 5'Flank | SBS | G | C |
| INDIA 8560 | ADAMTS3 | chr4 | 72336410 | Intron | SBS | G | C |
| INDIA 8560 | SP140 | chr2 | 230296718 | Intron | SBS | G | C |
| INDIA 8560 | Unknown | chr15 | 99454442 | IGR | SBS | G | C |
| INDIA 8560 | ENST00000417485 | chr2 | 215604948 | Intron | SBS | G | C |
| INDIA 8560 | ENST00000671266 | chr8 | 117279404 | Intron | SBS | C | G |
| INDIA 8560 | ACVR2A | chr2 | 147884338 | Intron | SBS | G | C |
| INDIA 8560 | TBC1D8 | chr2 | 101112988 | Intron | DEL | T | - |
| INDIA 8560 | LAMC1 | chr1 | 183096462 | Intron | SBS | G | C |
| INDIA 8560 | Unknown | chr6 | 85041295 | IGR | SBS | G | C |
| INDIA 8560 | Unknown | chr1 | 106748941 | IGR | SBS | G | C |
| INDIA 8560 | WFIKKN1 | chr16 | 636634 | 3'Flank | SBS | G | C |
| INDIA 8560 | Unknown | chr12 | 115499152 | IGR | SBS | G | T |
| INDIA 8560 | Unknown | chr16 | 47041845 | IGR | SBS | C | G |
| INDIA 8560 | ENST00000653824 | chr1 | 166288233 | Intron | SBS | G | T |
| INDIA 8560 | ANXA2 | chr15 | 60376800 | Intron | SBS | C | G |
| INDIA 8560 | ENST00000690299 | chr20 | 61163954 | 5'Flank | SBS | G | T |
| INDIA 8560 | PRDM16 | chr1 | 3281596 | Intron | SBS | G | C |
| INDIA 8560 | Unknown | chr7 | 46640128 | IGR | SBS | G | C |
| INDIA 8560 | ZC3HC1 | chr7 | 130045459 | Intron | SBS | G | C |
| INDIA 8560 | Unknown | chr10 | 57531051 | IGR | SBS | G | C |
| INDIA 8560 | Unknown | chr11 | 119746411 | IGR | SBS | G | C |
| INDIA 8560 | LINC02366 | chr12 | 10750362 | Intron | SBS | C | G |
| INDIA 8560 | AKAP13 | chr15 | 85597615 | Intron | SBS | C | G |
| INDIA 8560 | UNC13A | chr19 | 17649518 | Silent | SBS | C | G |
| INDIA 8560 | Unknown | chr2 | 46885609 | IGR | SBS | G | C |
| INDIA 8560 | ENST00000554041 | chr14 | 100851773 | Intron | SBS | G | C |
| INDIA 8560 | Unknown | chr14 | 98246929 | IGR | SBS | C | G |
| INDIA 8560 | ENST00000664519 | chrX | 142255736 | Intron | SBS | G | T |
| INDIA 8560 | Unknown | chr1 | 209398102 | IGR | SBS | G | C |
| INDIA 8560 | GRID1 | chr10 | 85764975 | Intron | SBS | G | C |
| INDIA 8560 | MASP1 | chr3 | 187248943 | Intron | SBS | T | A |
| INDIA 8560 | GSG1L | chr16 | 28053256 | Intron | SBS | T | A |
| INDIA 8560 | RORA | chr15 | 60793951 | Intron | SBS | C | G |
| INDIA 8560 | SLC30A8 | chr8 | 116988420 | Intron | SBS | C | G |
| INDIA 8560 | ENST00000667627 | chr3 | 34150013 | 5'Flank | SBS | C | G |
| INDIA 8560 | Unknown | chr20 | 55386900 | IGR | SBS | C | G |
| INDIA 8560 | TMEM234 | chr1 | 32223125 | 5'Flank | SBS | G | C |
| INDIA 8560 | CLINT1 | chr5 | 157781325 | 3'Flank | SBS | C | G |
| INDIA 8560 | ZMYM4 | chr1 | 35269572 | Intron | SBS | C | G |
| INDIA 8560 | RNY4P14 | chr13 | 39781184 | 3'Flank | SBS | G | C |
| INDIA 8560 | SLC4A1 | chr17 | 44258334 | Intron | SBS | T | A |
| INDIA 8560 | Unknown | chr15 | 90375906 | IGR | SBS | C | G |
| INDIA 8560 | FHIT | chr3 | 59865835 | Intron | SBS | C | G |
| INDIA 8560 | LIMCH1 | chr4 | 41469193 | Intron | SBS | C | G |
| INDIA 8560 | LINC01901 | chr18 | 39853137 | Intron | SBS | G | C |
| INDIA 8560 | ZFPM1 | chr16 | 88464070 | Intron | SBS | C | G |
| INDIA 8560 | ST6GALNAC2 | chr17 | 76575116 | Intron | SBS | G | C |
| INDIA 8560 | TASP1 | chr20 | 13393384 | Intron | SBS | G | C |
| INDIA 8560 | Unknown | chr8 | 133765912 | IGR | SBS | C | G |
| INDIA 8560 | ELMO1 | chr7 | 37147431 | Intron | SBS | C | G |
| INDIA 8560 | GAREM1 | chr18 | 32468556 | Intron | SBS | C | G |
| INDIA 8560 | GNPAT | chr1 | 231267951 | Intron | SBS | G | C |
| INDIA 8560 | Unknown | chr17 | 13047153 | IGR | SBS | G | C |
| INDIA 8560 | RHBDL3 | chr17 | 32299006 | Intron | SBS | G | C |
| INDIA 8561 | MPPED2 | chr11 | 30567399 | Intron | SBS | C | G |
| INDIA 8561 | Unknown | chr4 | 121252375 | IGR | SBS | C | G |
| INDIA 8561 | LINC01283 | chrX | 39417216 | Intron | SBS | G | C |
| INDIA 8561 | ROBO2 | chr3 | 76764041 | Intron | SBS | T | G |
| INDIA 8561 | SMUG1 | chr12 | 54193937 | 5'Flank | SBS | G | T |
| INDIA 8561 | Unknown | chr16 | 63296006 | IGR | SBS | A | T |
| INDIA 8561 | Unknown | chr4 | 116612941 | IGR | SBS | C | G |
| INDIA 8561 | ENST00000518973 | chr8 | 137934646 | Intron | SBS | C | G |

| | | | | | | | |
|---|---|---|---|---|---|---|---|
| INDIA 8561 | Unknown | chr8 | 139478381 | IGR | SBS | T | A |
| INDIA 8561 | OLFML3 | chr1 | 113981940 | 3'UTR | SBS | C | G |
| INDIA 8561 | LINC01229 | chr16 | 79757332 | Intron | SBS | G | C |
| INDIA 8561 | DCT | chr13 | 94435275 | 3'Flank | SBS | G | C |
| INDIA 8561 | KLHL36 | chr16 | 84667207 | 3'UTR | SBS | C | G |
| INDIA 8561 | ENST00000521731 | chr8 | 28259115 | Intron | SBS | G | C |
| INDIA 8561 | ITM2B | chr13 | 48266145 | 3'UTR | SBS | G | C |
| INDIA 8561 | PER1 | chr17 | 8155526 | 5'Flank | SBS | G | C |
| INDIA 8561 | MUC16 | chr19 | 8969870 | Missense_Mutation | SBS | C | G |
| INDIA 8561 | CPED1 | chr7 | 121056527 | Intron | SBS | C | G |
| INDIA 8561 | ACSL5 | chr10 | 112403988 | Intron | SBS | C | G |
| INDIA 8561 | SEMA5A | chr5 | 9235712 | Intron | SBS | G | C |
| INDIA 8561 | ENST00000649122 | chr9 | 11650302 | Intron | SBS | G | C |
| INDIA 8561 | Unknown | chr11 | 93958040 | IGR | SBS | C | G |
| INDIA 8561 | TTC7B | chr14 | 90609363 | Intron | SBS | G | C |
| INDIA 8561 | PCOLCE2 | chr3 | 142831901 | Intron | SBS | G | C |
| INDIA 8561 | DNAJC25 | chr9 | 111644855 | Intron | SBS | C | G |
| INDIA 8561 | Unknown | chr15 | 66851331 | IGR | SBS | G | C |
| INDIA 8561 | DNAH9 | chr17 | 11808539 | Intron | SBS | C | G |
| INDIA 8561 | CHD6 | chr20 | 41421748 | Silent | SBS | G | T |
| INDIA 8561 | Unknown | chr10 | 7137243 | IGR | SBS | G | C |
| INDIA 8561 | LHFPL5 | chr6 | 35807428 | Intron | SBS | G | C |
| INDIA 8561 | CHST9 | chr18 | 27018944 | Intron | DEL | A | - |
| INDIA 8561 | PLCD1 | chr3 | 38030259 | 5'Flank | SBS | G | C |
| INDIA 8561 | MBNL3 | chrX | 132389867 | Intron | SBS | C | G |
| INDIA 8561 | PLPPR5-AS1 | chr1 | 99186191 | Intron | SBS | C | G |
| INDIA 8561 | NAV1 | chr1 | 201651357 | Intron | SBS | C | G |
| INDIA 8561 | Unknown | chr1 | 208015879 | IGR | SBS | G | C |
| INDIA 8561 | ENST00000453451 | chr2 | 34946804 | Intron | SBS | A | C |
| INDIA 8561 | PER2 | chr2 | 238274782 | Intron | SBS | G | C |
| INDIA 8561 | CLEC3B | chr3 | 45027803 | Intron | SBS | G | C |
| INDIA 8561 | Unknown | chr3 | 126374337 | IGR | SBS | C | G |
| INDIA 8561 | FHIP1A | chr4 | 151537938 | Intron | SBS | C | G |
| INDIA 8561 | SEMA5A | chr5 | 9038021 | 3'UTR | SBS | G | C |
| INDIA 8561 | CDH12 | chr5 | 22576599 | Intron | SBS | G | C |
| INDIA 8561 | IL13 | chr5 | 132662769 | 3'Flank | SBS | C | G |
| INDIA 8561 | ATXN1 | chr6 | 16553362 | Intron | SBS | C | G |
| INDIA 8561 | Unknown | chr6 | 112742939 | IGR | SBS | C | G |
| INDIA 8561 | PEX7 | chr6 | 136844860 | Intron | SBS | G | C |
| INDIA 8561 | FABP12 | chr8 | 81560295 | Intron | SBS | G | C |
| INDIA 8561 | MYO3A | chr10 | 26122117 | Intron | SBS | C | G |
| INDIA 8561 | Unknown | chr10 | 105514107 | IGR | SBS | G | C |
| INDIA 8561 | RBM14 | chr11 | 66619368 | Intron | SBS | C | G |
| INDIA 8561 | Unknown | chr13 | 21814707 | IGR | SBS | G | C |
| INDIA 8561 | SLC47A1 | chr17 | 19541556 | Intron | SBS | G | C |
| INDIA 8561 | ZNF536 | chr19 | 30569327 | Intron | SBS | C | G |
| INDIA 8561 | ENST00000446280 | chr20 | 48384848 | RNA | SBS | G | C |
| INDIA 8561 | Unknown | chr2 | 194993474 | IGR | SBS | G | T |
| INDIA 8561 | WSB1 | chr17 | 27309282 | Intron | SBS | G | C |
| INDIA 8561 | SPAG16 | chr2 | 214167230 | Intron | SBS | C | G |
| INDIA 8561 | LINC00670 | chr17 | 12566495 | Intron | SBS | G | C |
| INDIA 8561 | Unknown | chr2 | 99496036 | IGR | SBS | C | G |
| INDIA 8561 | Unknown | chr13 | 27500983 | IGR | SBS | G | C |
| INDIA 8561 | KY | chr3 | 134625821 | Intron | DEL | ACTC | - |
| INDIA 8561 | PSKH2 | chr8 | 86045147 | 3'Flank | SBS | G | C |
| INDIA 8561 | CCDC6 | chr10 | 59899940 | Intron | SBS | C | G |
| INDIA 8561 | LINC-PINT | chr7 | 130910186 | Intron | SBS | C | G |
| INDIA 8561 | Unknown | chr1 | 79737840 | IGR | SBS | G | T |
| INDIA 8561 | PDZRN3 | chr3 | 73401323 | Intron | SBS | C | G |
| INDIA 8561 | LINC02261 | chr4 | 27224014 | Intron | SBS | G | C |
| INDIA 8561 | LINC01942 | chr5 | 173690269 | Intron | SBS | G | C |
| INDIA 8561 | ITPRID1 | chr7 | 31652400 | Intron | SBS | C | G |
| INDIA 8561 | Unknown | chr7 | 83214068 | IGR | SBS | G | C |
| INDIA 8561 | Unknown | chr8 | 102629965 | IGR | SBS | G | C |
| INDIA 8561 | GLT1D1 | chr12 | 128932955 | Intron | SBS | A | T |
| INDIA 8561 | CTIF | chr18 | 48591002 | Intron | SBS | C | G |

| | | | | | | | |
|---|---|---|---|---|---|---|---|
| INDIA 8561 | LINC01591 | chr8 | 135288299 | Intron | SBS | G | T |
| INDIA 8561 | Unknown | chr18 | 76101037 | IGR | SBS | A | T |
| INDIA 8561 | GALNT9 | chr12 | 132251533 | Intron | SBS | C | G |
| INDIA 8561 | POU1F1 | chr3 | 87271317 | Intron | SBS | G | C |
| INDIA 8561 | CUX1 | chr7 | 101980617 | Intron | SBS | C | G |
| INDIA 8561 | RIN2 | chr20 | 19977326 | Intron | SBS | C | G |
| INDIA 8561 | PPIP5K2 | chr5 | 103130812 | Intron | SBS | G | C |
| INDIA 8561 | DAAM2 | chr6 | 39821072 | Intron | SBS | G | C |
| INDIA 8561 | ZNF467 | chr7 | 149767541 | Intron | SBS | C | G |
| INDIA 8561 | Unknown | chr1 | 240690881 | IGR | SBS | G | C |
| INDIA 8561 | Unknown | chr17 | 31724754 | IGR | SBS | C | G |
| INDIA 8561 | WDR4 | chr21 | 42859347 | Intron | SBS | C | G |
| INDIA 8561 | ROBO2 | chr3 | 77643141 | Intron | DEL | C | - |
| INDIA 8561 | ABLIM1 | chr10 | 114649612 | Intron | SBS | G | C |
| INDIA 8561 | TMEM218 | chr11 | 125096296 | 3'UTR | SBS | C | G |
| INDIA 8561 | Unknown | chr12 | 96050744 | IGR | SBS | G | C |
| INDIA 8561 | SLC25A21 | chr14 | 37172614 | 5'Flank | SBS | G | C |
| INDIA 8561 | LINC00907 | chr18 | 42428273 | Intron | SBS | C | G |
| INDIA 8561 | E2F5-DT | chr8 | 85168422 | 3'Flank | DEL | T | - |
| INDIA 8563 | LINC02930 | chr10 | 120650996 | Intron | SBS | G | C |
| INDIA 8563 | FAM135B | chr8 | 138490296 | Intron | SBS | C | G |
| INDIA 8563 | NOS1AP | chr1 | 162224851 | Intron | SBS | G | C |
| INDIA 8563 | Unknown | chr2 | 74817246 | IGR | SBS | A | T |
| INDIA 8563 | DYNLT5 | chr1 | 66774350 | Intron | SBS | C | G |
| INDIA 8563 | ENST00000650577 | chr17 | 53942014 | Intron | SBS | T | A |
| INDIA 8563 | DHFR | chr5 | 80631775 | Intron | SBS | C | G |
| INDIA 8563 | SEMA6A | chr5 | 116549978 | Intron | SBS | C | G |
| INDIA 8563 | Unknown | chr7 | 38674487 | IGR | SBS | T | A |
| INDIA 8563 | GRID1 | chr10 | 85601542 | 3'UTR | SBS | G | C |
| INDIA 8563 | EMX2OS | chr10 | 117505680 | Intron | SBS | C | A |
| INDIA 8563 | MINAR2 | chr5 | 129771415 | 3'Flank | SBS | C | G |
| INDIA 8563 | INHCAP | chr3 | 133670508 | Intron | SBS | G | C |
| INDIA 8563 | TARS3 | chr15 | 101711015 | Intron | SBS | C | G |
| INDIA 8563 | TGFBR3 | chr1 | 91795799 | Intron | SBS | C | G |
| INDIA 8563 | ADGRV1 | chr5 | 90731603 | Intron | SBS | G | C |
| INDIA 8563 | SYNJ2 | chr6 | 157994326 | Intron | SBS | G | C |
| INDIA 8563 | Unknown | chr7 | 41911626 | IGR | DEL | A | - |
| INDIA 8563 | Unknown | chr7 | 125952886 | IGR | SBS | C | G |
| INDIA 8563 | ENST00000665012 | chr8 | 90318330 | Intron | SBS | C | G |
| INDIA 8563 | PTK2 | chr8 | 140704416 | Intron | SBS | C | G |
| INDIA 8563 | ENST00000421324 | chr10 | 78314896 | Intron | SBS | C | A |
| INDIA 8563 | TMCC3 | chr12 | 94622115 | Intron | SBS | C | G |
| INDIA 8563 | VAT1L | chr16 | 77825721 | Intron | SBS | G | C |
| INDIA 8563 | Unknown | chr18 | 63518771 | IGR | SBS | C | G |
| INDIA 8563 | ZNF233 | chr19 | 44268716 | Intron | SBS | G | C |
| INDIA 8563 | Unknown | chr2 | 112794941 | IGR | SBS | C | G |
| INDIA 8563 | Unknown | chr2 | 41998389 | IGR | SBS | G | C |
| INDIA 8563 | ENST00000552778 | chr12 | 89566887 | Intron | SBS | C | G |
| INDIA 8563 | Unknown | chr1 | 40929819 | IGR | DEL | G | - |
| INDIA 8563 | SLC8A1 | chr2 | 40199289 | Intron | SBS | G | C |
| INDIA 8563 | CACNA2D3 | chr3 | 54276729 | Intron | SBS | G | C |
| INDIA 8563 | Unknown | chr9 | 12248504 | IGR | SBS | C | A |
| INDIA 8563 | MARCHF7 | chr2 | 159748450 | Missense_Mutation | SBS | C | G |
| INDIA 8563 | CDH4 | chr20 | 61746146 | Intron | SBS | G | C |
| INDIA 8563 | ENST00000556361 | chr14 | 66415083 | Intron | SBS | G | T |
| INDIA 8563 | Unknown | chr5 | 58349392 | IGR | SBS | C | G |
| INDIA 8563 | Unknown | chr9 | 108638416 | IGR | SBS | G | C |
| INDIA 8563 | KIF6 | chr6 | 39445581 | Intron | SBS | T | A |
| INDIA 8563 | AMBRA1 | chr11 | 46415324 | Intron | SBS | C | G |
| INDIA 8563 | ENST00000665334 | chr14 | 48205095 | Intron | DEL | T | - |
| INDIA 8563 | SPOCK1 | chr5 | 137155517 | Intron | SBS | G | C |
| INDIA 8563 | DOCK8 | chr9 | 346629 | Intron | DEL | TGATT | - |
| INDIA 8563 | PMF1 | chr1 | 156223134 | Intron | SBS | G | C |
| INDIA 8563 | SPTBN1 | chr2 | 54591769 | Intron | SBS | C | G |
| INDIA 8563 | NAV2 | chr11 | 19873695 | Intron | SBS | C | G |
| INDIA 8563 | PLCB1 | chr20 | 8251920 | Intron | SBS | G | C |

| | | | | | | | |
|---|---|---|---|---|---|---|---|
| INDIA 8563 | LINC02810 | chr1 | 17733919 | Intron | SBS | C | G |
| INDIA 8563 | Unknown | chr9 | 98923212 | IGR | DEL | C | - |
| INDIA 8563 | ENST00000658872 | chr10 | 31118219 | Intron | SBS | T | A |
| INDIA 8563 | SLIT2 | chr4 | 20277036 | Intron | SBS | G | C |
| INDIA 8563 | Unknown | chr6 | 40659322 | IGR | SBS | C | G |
| INDIA 8563 | Unknown | chr13 | 102719166 | IGR | SBS | A | T |
| INDIA 8563 | ENST00000580658 | chr17 | 34078954 | 3'Flank | SBS | G | C |
| INDIA 8563 | CACNA1S | chr1 | 201048954 | Intron | SBS | G | T |
| INDIA 8563 | CUL1 | chr7 | 148783885 | Missense_Mutation | SBS | G | C |
| INDIA 8563 | Unknown | chr16 | 49365198 | IGR | SBS | G | C |
| INDIA 8563 | Unknown | chr2 | 7803019 | IGR | SBS | G | C |
| INDIA 8563 | SV2A | chr1 | 149916958 | Intron | SBS | G | C |
| INDIA 8563 | GBE1 | chr3 | 81758774 | Intron | SBS | C | G |
| INDIA 8563 | NHLRC1 | chr6 | 18121480 | Missense_Mutation | SBS | G | C |
| INDIA 8563 | C9orf24 | chr9 | 34382679 | Intron | SBS | A | T |
| INDIA 8563 | SHANK2 | chr11 | 70897281 | Intron | SBS | G | C |
| INDIA 8563 | MAP4K3-DT | chr2 | 39609259 | Intron | INS | - | T |
| INDIA 8563 | POU6F2 | chr7 | 39043928 | Intron | SBS | C | A |
| INDIA 8563 | Unknown | chr8 | 34045501 | IGR | SBS | C | G |
| INDIA 8563 | Unknown | chr10 | 110848329 | IGR | SBS | G | C |
| INDIA 8563 | ENST00000553754 | chr14 | 65318210 | Intron | SBS | C | G |
| INDIA 8563 | Unknown | chr2 | 40869143 | IGR | DEL | A | - |
| INDIA 8563 | ZBTB8OS | chr1 | 32651252 | 5'Flank | SBS | C | G |
| INDIA 8563 | FYCO1 | chr3 | 45928365 | Intron | SBS | G | C |
| INDIA 8563 | Unknown | chr10 | 16331286 | IGR | SBS | C | G |
| INDIA 8563 | Unknown | chr4 | 10977177 | IGR | SBS | C | G |
| INDIA 8563 | ENST00000671933 | chr4 | 25560481 | Intron | SBS | G | C |
| INDIA 8563 | DAB2IP | chr9 | 121693538 | Intron | SBS | C | G |
| INDIA 8563 | Unknown | chr7 | 95670315 | IGR | SBS | C | G |
| INDIA 8563 | LRRC59 | chr17 | 50394943 | Nonsense_Mutation | INS | - | TTTA |
| INDIA 8563 | LINC01818 | chr2 | 150327895 | Intron | SBS | G | C |
| INDIA 8563 | Unknown | chr12 | 127093593 | IGR | SBS | C | G |
| INDIA 8563 | WDR59 | chr16 | 74947115 | Intron | SBS | G | C |
| INDIA 8563 | SPHK1 | chr17 | 76377347 | Intron | SBS | C | G |
| INDIA 8563 | STK17A | chr7 | 43609884 | Intron | SBS | C | G |
| INDIA 8563 | Unknown | chr7 | 84831914 | IGR | SBS | A | T |
| INDIA 8563 | PKHD1L1 | chr8 | 109464597 | Missense_Mutation | SBS | A | T |
| INDIA 8563 | MICU1 | chr10 | 72389093 | Intron | SBS | G | C |
| INDIA 8563 | DAB2 | chr5 | 39464303 | 5'Flank | SBS | G | C |
| INDIA 8563 | Unknown | chr1 | 172499753 | IGR | SBS | G | T |
| INDIA 8563 | LRIG1 | chr3 | 66384511 | Intron | SBS | C | G |
| INDIA 8563 | MAPK8IP3 | chr16 | 1729514 | Missense_Mutation | SBS | G | C |
| INDIA 8563 | PAX1 | chr20 | 21713924 | Intron | SBS | G | C |
| INDIA 8563 | ENST00000660981 | chr16 | 54494651 | 5'Flank | SBS | G | C |
| INDIA 8563 | WDR59 | chr16 | 74872678 | 3'UTR | SBS | G | C |
| INDIA 8563 | ENST00000665984 | chr1 | 72560797 | Intron | SBS | G | C |
| INDIA 8563 | TEAD4 | chr12 | 2969080 | Intron | SBS | G | C |
| INDIA 8564 | HS3ST2 | chr16 | 22874643 | Intron | SBS | G | C |
| INDIA 8564 | Unknown | chrX | 104049173 | IGR | SBS | G | C |
| INDIA 8564 | Unknown | chr9 | 2335501 | IGR | SBS | C | G |
| INDIA 8564 | NLGN4X | chrX | 6227618 | Intron | SBS | C | G |
| INDIA 8564 | ERBB4 | chr2 | 211439220 | Intron | SBS | T | G |
| INDIA 8564 | Unknown | chrX | 153431883 | IGR | SBS | C | G |
| INDIA 8564 | RASSF1 | chr3 | 50331080 | Intron | SBS | C | G |
| INDIA 8564 | ITGAL | chr16 | 30526080 | 3'Flank | SBS | G | C |
| INDIA 8564 | ERCC4 | chr16 | 13940966 | Intron | SBS | G | C |
| INDIA 8564 | NR3C1 | chr5 | 143348075 | Intron | SBS | C | G |
| INDIA 8564 | SMARCA2 | chr9 | 2100019 | Intron | SBS | G | C |
| INDIA 8564 | Unknown | chr11 | 7209027 | IGR | SBS | G | C |
| INDIA 8564 | SVEP1 | chr9 | 110521219 | Intron | SBS | C | G |
| INDIA 8564 | CADM1 | chr11 | 115422476 | Intron | SBS | G | C |
| INDIA 8564 | XYLT1 | chr16 | 17117593 | Intron | SBS | C | G |
| INDIA 8564 | MAP7D2 | chrX | 20019799 | Intron | SBS | C | G |
| INDIA 8564 | PRELID2 | chr5 | 145595705 | Intron | SBS | C | G |
| INDIA 8564 | IGF2R | chr6 | 159991273 | Nonsense_Mutation | SBS | C | G |
| INDIA 8564 | SLC24A2 | chr9 | 19646892 | Intron | SBS | C | G |

| | | | | | | | |
|---|---|---|---|---|---|---|---|
| INDIA 8564 | ENST00000640003 | chr9 | 23138629 | Intron | SBS | C | G |
| INDIA 8564 | ENST00000650453 | chr9 | 84434387 | Intron | SBS | C | G |
| INDIA 8564 | ZNF41 | chrX | 47456215 | Intron | SBS | C | G |
| INDIA 8564 | Unknown | chr2 | 155208846 | IGR | SBS | C | G |
| INDIA 8564 | Unknown | chr2 | 155208843 | IGR | SBS | G | C |
| INDIA 8564 | PTBP3 | chr9 | 112261116 | Intron | SBS | C | G |
| INDIA 8564 | NTM | chr11 | 132012829 | Intron | SBS | G | C |
| INDIA 8564 | TRPM3 | chr9 | 70812928 | Intron | SBS | C | G |
| INDIA 8564 | PSD3 | chr8 | 18729407 | Intron | SBS | C | G |
| INDIA 8564 | PRAG1 | chr8 | 8348439 | Intron | SBS | A | T |
| INDIA 8564 | PLXNB3 | chrX | 153778552 | Intron | SBS | G | C |
| INDIA 8564 | ADRA1A | chr8 | 26842940 | Intron | SBS | G | C |
| INDIA 8564 | NAV2 | chr11 | 20077548 | Splice_Region | SBS | C | G |
| INDIA 8564 | Unknown | chrX | 4606960 | IGR | SBS | G | C |
| INDIA 8564 | Unknown | chr2 | 179891697 | IGR | SBS | G | C |
| INDIA 8564 | Unknown | chr10 | 20688097 | IGR | SBS | G | C |
| INDIA 8564 | Unknown | chrX | 49677426 | IGR | SBS | C | G |
| INDIA 8564 | CPO | chr2 | 206943887 | Intron | SBS | G | C |
| INDIA 8564 | VPS26C | chr21 | 37220420 | 3'Flank | SBS | C | G |
| INDIA 8564 | SNX29 | chr16 | 12510080 | Intron | SBS | C | G |
| INDIA 8564 | LRP2 | chr2 | 169338711 | Intron | SBS | G | C |
| INDIA 8564 | CDIPT | chr16 | 29858866 | 3'UTR | SBS | C | A |
| INDIA 8564 | SGCD | chr5 | 156326736 | 5'Flank | SBS | G | C |
| INDIA 8564 | AFF3 | chr2 | 99859740 | Intron | SBS | G | C |
| INDIA 8564 | TBR1 | chr2 | 161423194 | Intron | SBS | G | C |
| INDIA 8564 | MYO3A | chr10 | 26111746 | Intron | SBS | G | C |
| INDIA 8564 | ZBTB16 | chr11 | 114205527 | Intron | SBS | G | C |
| INDIA 8564 | ACSM1 | chr16 | 20660382 | Intron | SBS | C | G |
| INDIA 8564 | Unknown | chr11 | 2187163 | IGR | SBS | C | G |
| INDIA 8564 | ZMYM3 | chrX | 71235400 | 3'Flank | SBS | C | G |
| INDIA 8564 | Unknown | chr11 | 119772533 | IGR | SBS | G | C |
| INDIA 8564 | Unknown | chr2 | 153413147 | IGR | SBS | G | T |
| INDIA 8564 | TNC | chr9 | 115018744 | 3'Flank | SBS | C | G |
| INDIA 8564 | ADGRG2 | chrX | 19070596 | Intron | SBS | G | C |
| INDIA 8564 | LRRC4C | chr11 | 40296242 | Intron | SBS | G | C |
| INDIA 8564 | AOPEP | chr9 | 95057262 | Intron | SBS | C | G |
| INDIA 8564 | B4GALT4 | chr3 | 119213698 | Intron | SBS | C | G |
| INDIA 8564 | DUSP4 | chr8 | 29341448 | Intron | SBS | C | G |
| INDIA 8564 | Unknown | chr16 | 86790045 | IGR | SBS | G | C |
| INDIA 8564 | ASTN2 | chr9 | 116928020 | Intron | SBS | G | C |
| INDIA 8564 | LINC00703 | chr10 | 4409715 | RNA | SBS | G | C |
| INDIA 8564 | NAV2 | chr11 | 19821511 | Intron | SBS | C | G |
| INDIA 8564 | ARPP21 | chr3 | 35663981 | Intron | SBS | G | C |
| INDIA 8564 | Unknown | chr8 | 8528612 | IGR | SBS | G | C |
| INDIA 8564 | Unknown | chr2 | 203322989 | IGR | SBS | C | G |
| INDIA 8564 | ERP44 | chr9 | 100020225 | Intron | SBS | G | C |
| INDIA 8564 | Unknown | chr15 | 76964791 | IGR | SBS | G | C |
| INDIA 8564 | Unknown | chrX | 68977911 | IGR | SBS | G | T |
| INDIA 8564 | RCSD1 | chr1 | 167644644 | Intron | SBS | G | C |
| INDIA 8564 | Unknown | chrX | 85429620 | IGR | SBS | G | C |
| INDIA 8564 | ITGB1-DT | chr10 | 33069720 | Intron | SBS | C | G |
| INDIA 8564 | ABHD2 | chr15 | 89176589 | Intron | SBS | G | C |
| INDIA 8564 | ENST00000669987 | chr9 | 36794496 | Intron | SBS | C | G |
| INDIA 8564 | Unknown | chr1 | 162944651 | IGR | SBS | C | G |
| INDIA 8564 | Unknown | chr1 | 159258594 | IGR | SBS | C | G |
| INDIA 8564 | DIRC3-AS1 | chr2 | 216937362 | Intron | SBS | G | C |
| INDIA 8564 | ENST00000636095 | chrX | 137871438 | Intron | SBS | T | A |
| INDIA 8564 | SLC9A3R2 | chr16 | 2043540 | 3'Flank | SBS | G | C |
| INDIA 8564 | LINC01127 | chr2 | 101960293 | 5'Flank | SBS | C | G |
| INDIA 8564 | COL4A3 | chr2 | 227202240 | Intron | SBS | G | C |
| INDIA 8564 | GAP43 | chr3 | 115655876 | Intron | SBS | G | C |
| INDIA 8564 | GRID2 | chr4 | 93308457 | Intron | SBS | C | G |
| INDIA 8564 | UGT8 | chr4 | 114624640 | Intron | SBS | G | C |
| INDIA 8564 | Unknown | chr9 | 75446259 | IGR | INS | - | G |
| INDIA 8564 | WHRN | chr9 | 114407078 | Intron | SBS | G | C |
| INDIA 8564 | PARD3 | chr10 | 34365550 | Intron | SBS | G | C |

| | | | | | | | |
|---|---|---|---|---|---|---|---|
| INDIA 8564 | Unknown | chr11 | 23224123 | IGR | SBS | G | C |
| INDIA 8564 | SLC1A2 | chr11 | 35288250 | Intron | SBS | C | G |
| INDIA 8564 | CENPV | chr17 | 16348736 | Intron | SBS | C | G |
| INDIA 8564 | Unknown | chrX | 45958763 | IGR | SBS | G | C |
| INDIA 8564 | KIAA1217 | chr10 | 24470162 | Intron | SBS | G | C |
| INDIA 8565 | EHBP1 | chr2 | 62991693 | Intron | SBS | G | T |
| INDIA 8565 | Unknown | chr1 | 73958198 | IGR | DEL | G | - |
| INDIA 8565 | E2F2 | chr1 | 23506779 | 3'UTR | DEL | ACA | - |
| INDIA 8565 | DCC | chr18 | 52936722 | Intron | SBS | G | T |
| INDIA 8565 | DYNC1H1 | chr14 | 101992570 | Intron | SBS | C | G |
| INDIA 8565 | NELL1 | chr11 | 20747987 | Intron | DEL | G | - |
| INDIA 8565 | Unknown | chr18 | 65372903 | IGR | SBS | C | G |
| INDIA 8565 | SPOCD1 | chr1 | 31786467 | 3'Flank | SBS | T | A |
| INDIA 8565 | Unknown | chr6 | 17046874 | IGR | DEL | A | - |
| INDIA 8565 | MTCL1 | chr18 | 8786728 | Intron | SBS | G | C |
| INDIA 8565 | LINC02119 | chr5 | 38093602 | Intron | SBS | C | G |
| INDIA 8565 | SORCS3 | chr10 | 104899422 | Intron | SBS | T | A |
| INDIA 8565 | Unknown | chr10 | 2671067 | IGR | DEL | C | - |
| INDIA 8565 | TENM3-AS1 | chr4 | 182020064 | Intron | SBS | T | A |
| INDIA 8565 | MYO9A | chr15 | 72019049 | Missense_Mutation | SBS | T | A |
| INDIA 8565 | Unknown | chr11 | 27911389 | IGR | SBS | G | C |
| INDIA 8565 | Unknown | chr2 | 155207936 | IGR | SBS | C | G |
| INDIA 8565 | Unknown | chr6 | 99864103 | IGR | DEL | AC | - |
| INDIA 8565 | Unknown | chr18 | 22297917 | IGR | DEL | AT | - |
| INDIA 8565 | Unknown | chr12 | 23339112 | IGR | SBS | G | T |
| INDIA 8565 | LRRC7 | chr1 | 69880439 | Intron | DEL | C | - |
| INDIA 8565 | Unknown | chr5 | 2301944 | IGR | SBS | A | T |
| INDIA 8565 | XIRP2 | chr2 | 167195776 | Intron | SBS | C | A |
| INDIA 8565 | Unknown | chr9 | 129398404 | IGR | SBS | T | A |
| INDIA 8565 | ITM2C | chr2 | 230871194 | Intron | DEL | C | - |
| INDIA 8565 | PRKN | chr6 | 161999147 | Intron | SBS | C | A |
| INDIA 8565 | DPP9 | chr19 | 4714105 | Missense_Mutation | SBS | G | C |
| INDIA 8565 | ENST00000668062 | chr7 | 136340962 | Intron | SBS | C | A |
| INDIA 8565 | Unknown | chr6 | 65824156 | IGR | DEL | C | - |
| INDIA 8565 | CD244 | chr1 | 160836434 | Intron | DEL | G | - |
| INDIA 8565 | Unknown | chr16 | 51535718 | IGR | SBS | G | T |
| INDIA 8565 | ENST00000657278 | chr13 | 105982160 | Intron | SBS | C | G |
| INDIA 8565 | ENST00000450290 | chr2 | 80861799 | Intron | SBS | A | T |
| INDIA 8565 | PLCL1 | chr2 | 198085564 | Missense_Mutation | SBS | G | T |
| INDIA 8565 | Unknown | chr8 | 126387408 | IGR | SBS | C | A |
| INDIA 8565 | BNC2 | chr9 | 16813989 | Intron | DEL | G | - |
| INDIA 8565 | DENND1A | chr9 | 123569959 | Intron | SBS | T | A |
| INDIA 8565 | CFAP54 | chr12 | 96741083 | Intron | SBS | C | G |
| INDIA 8565 | Unknown | chr13 | 90263416 | IGR | SBS | G | C |
| INDIA 8565 | LINC02126 | chr16 | 65170095 | Intron | SBS | A | T |
| INDIA 8565 | Unknown | chr18 | 25668251 | IGR | SBS | G | C |
| INDIA 8565 | LINC01428 | chr20 | 7189956 | Intron | SBS | T | A |
| INDIA 8565 | TM9SF4 | chr20 | 32169724 | 3'Flank | SBS | A | T |
| INDIA 8565 | MYLK | chr3 | 123864899 | Intron | SBS | T | A |
| INDIA 8565 | ENST00000685457 | chr2 | 203232682 | 3'Flank | SBS | T | G |
| INDIA 8565 | Unknown | chr7 | 31460565 | IGR | SBS | C | G |
| INDIA 8565 | CCDC149 | chr4 | 24903418 | Intron | SBS | G | C |
| INDIA 8565 | NEGR1 | chr1 | 72282094 | Intron | SBS | G | C |
| INDIA 8565 | RAB8B | chr15 | 63240113 | Intron | SBS | G | T |
| INDIA 8565 | Unknown | chr2 | 235417661 | IGR | SBS | G | T |
| INDIA 8565 | LINC01705 | chr1 | 222055689 | Intron | SBS | C | G |
| INDIA 8565 | Unknown | chr18 | 25661584 | IGR | SBS | C | A |
| INDIA 8565 | SORCS2 | chr4 | 7699555 | Intron | SBS | A | T |
| INDIA 8565 | Unknown | chr6 | 39775770 | IGR | SBS | G | C |
| INDIA 8565 | ENST00000620729 | chr17 | 32404447 | 5'Flank | SBS | C | G |
| INDIA 8565 | ENST00000523584 | chr5 | 31089193 | 3'Flank | SBS | C | A |
| INDIA 8565 | Unknown | chr3 | 80898657 | IGR | SBS | G | T |
| INDIA 8565 | SPATS2L | chr2 | 200379651 | Intron | SBS | C | A |
| INDIA 8565 | Unknown | chr3 | 1865331 | IGR | SBS | G | C |
| INDIA 8565 | NTRK3 | chr15 | 88082680 | Intron | SBS | G | C |
| INDIA 8565 | MECOM | chr3 | 169089074 | Missense_Mutation | SBS | C | G |

| | | | | | | | |
|---|---|---|---|---|---|---|---|
| INDIA 8565 | Unknown | chr20 | 18251872 | IGR | DEL | G | - |
| INDIA 8565 | ENST00000673993 | chr22 | 48336404 | 5'Flank | INS | - | T |
| INDIA 8565 | TPK1 | chr7 | 144636031 | Intron | SBS | A | T |
| INDIA 8565 | OSBPL2 | chr20 | 62294199 | 3'UTR | SBS | C | G |
| INDIA 8565 | BMPER | chr7 | 34145532 | Intron | SBS | G | C |
| INDIA 8565 | Unknown | chr5 | 61816362 | IGR | SBS | G | C |
| INDIA 8565 | Unknown | chr5 | 165296564 | IGR | SBS | G | C |
| INDIA 8565 | OTX1 | chr2 | 63055368 | Intron | SBS | G | C |
| INDIA 8565 | LSAMP | chr3 | 116637419 | Intron | SBS | C | A |
| INDIA 8565 | ENST00000589276 | chr19 | 5860293 | 3'Flank | SBS | G | T |
| INDIA 8565 | Unknown | chr22 | 42545447 | IGR | SBS | C | G |
| INDIA 8565 | FGF12 | chr3 | 192739100 | Intron | SBS | T | A |
| INDIA 8565 | ENST00000412811 | chr3 | 40834121 | Intron | DEL | C | - |
| INDIA 8565 | FAM124B | chr2 | 224398638 | Intron | SBS | G | C |
| INDIA 8565 | IQCE | chr7 | 2584449 | Intron | SBS | C | G |
| INDIA 8565 | CTU1 | chr19 | 51107054 | Intron | SBS | C | G |
| INDIA 8565 | GRHL2 | chr8 | 101502615 | Intron | DEL | G | - |
| INDIA 8565 | MIR3681HG | chr2 | 12498234 | Intron | SBS | A | T |
| INDIA 8565 | FAT2 | chr5 | 151500059 | 3'Flank | SBS | G | T |
| INDIA 8565 | ELMO1 | chr7 | 37331739 | Intron | SBS | G | C |
| INDIA 8565 | Unknown | chr6 | 143526112 | IGR | DEL | C | - |
| INDIA 8565 | ENST00000563114 | chr16 | 78054186 | Intron | SBS | T | A |
| INDIA 8565 | DENND1A | chr9 | 123570089 | Intron | SBS | C | G |
| INDIA 8565 | CACNA1C | chr12 | 2534049 | Intron | SBS | T | A |
| INDIA 8565 | ENST00000654812 | chr20 | 12291135 | Intron | SBS | G | C |
| INDIA 8565 | CNTNAP2 | chr7 | 147462159 | Intron | SBS | G | C |
| INDIA 8565 | Unknown | chr6 | 138031065 | IGR | SBS | G | C |
| INDIA 8565 | CCDC42 | chr17 | 8726928 | 3'Flank | DEL | GT | - |
| INDIA 8565 | ADGRV1 | chr5 | 91027467 | Intron | SBS | G | C |
| INDIA 8565 | AHRR | chr5 | 392831 | Intron | SBS | G | T |
| INDIA 8565 | ENST00000599640 | chr1 | 148161246 | RNA | SBS | C | G |
| INDIA 8565 | PITPNM2 | chr12 | 123079115 | Intron | SBS | C | G |
| INDIA 8565 | PSD3 | chr8 | 18975089 | Intron | SBS | C | A |
| INDIA 8566 | ADAMTS16 | chr5 | 5277466 | Intron | SBS | G | C |
| INDIA 8566 | RBM10 | chrX | 47181332 | Frame_Shift_Del | DEL | GCCCATG | - |
| INDIA 8566 | DNAH7 | chr2 | 195962115 | Intron | SBS | G | C |
| INDIA 8566 | ZFHX3 | chr16 | 72911072 | Intron | SBS | C | A |
| INDIA 8566 | IP6K3 | chr6 | 33722977 | Missense_Mutation | SBS | G | C |
| INDIA 8566 | GPAM | chr10 | 112191199 | Intron | SBS | G | C |
| INDIA 8566 | PCDHB9 | chr5 | 141187198 | 5'UTR | DEL | C | - |
| INDIA 8566 | Unknown | chr1 | 98781046 | IGR | SBS | G | C |
| INDIA 8566 | ENST00000507435 | chr5 | 4533329 | Intron | SBS | C | G |
| INDIA 8566 | Unknown | chr6 | 139918687 | IGR | SBS | C | G |
| INDIA 8566 | ENST00000668718 | chr5 | 25994993 | 3'Flank | SBS | G | C |
| INDIA 8566 | LINC02240 | chr5 | 125126769 | Intron | SBS | C | G |
| INDIA 8566 | VIPAS39 | chr14 | 77448361 | Intron | SBS | C | G |
| INDIA 8566 | RIMS1 | chr6 | 72029679 | Intron | SBS | A | T |
| INDIA 8566 | ENST00000640504 | chr8 | 102685802 | RNA | SBS | G | C |
| INDIA 8566 | PTPRD | chr9 | 8591721 | Intron | SBS | G | C |
| INDIA 8566 | RUNX1 | chr21 | 35393068 | Intron | SBS | A | C |
| INDIA 8566 | ACOT11 | chr1 | 54580829 | Intron | SBS | G | C |
| INDIA 8566 | Unknown | chr12 | 5166173 | IGR | SBS | A | T |
| INDIA 8566 | ZMIZ1 | chr10 | 79223675 | Intron | DEL | AC | - |
| INDIA 8566 | Unknown | chr15 | 26513852 | IGR | SBS | G | C |
| INDIA 8566 | THSD4 | chr15 | 71495599 | Intron | SBS | C | G |
| INDIA 8566 | Unknown | chr17 | 53236679 | IGR | SBS | A | T |
| INDIA 8566 | SPOCK1 | chr5 | 137073036 | Intron | SBS | G | C |
| INDIA 8566 | DMTF1 | chr7 | 87154749 | Intron | SBS | C | G |
| INDIA 8566 | OTOGL | chr12 | 80257205 | Intron | SBS | C | G |
| INDIA 8566 | Unknown | chr13 | 47182439 | IGR | SBS | C | G |
| INDIA 8566 | XPR1 | chr1 | 180892925 | 3'Flank | SBS | C | G |
| INDIA 8566 | CDH4 | chr20 | 61313541 | Intron | SBS | G | C |
| INDIA 8566 | ENST00000656098 | chr6 | 98271458 | Intron | SBS | T | G |
| INDIA 8566 | UVRAG | chr11 | 76131401 | Intron | SBS | G | C |
| INDIA 8566 | KANK4 | chr1 | 62312585 | Intron | SBS | C | G |
| INDIA 8566 | Unknown | chr1 | 91582361 | IGR | SBS | C | G |

| | | | | | | | |
|---|---|---|---|---|---|---|---|
| INDIA 8566 | S100A7 | chr1 | 153459930 | Intron | DEL | G | - |
| INDIA 8566 | GPATCH2 | chr1 | 217538938 | Intron | SBS | C | G |
| INDIA 8566 | PLD5 | chr1 | 242432634 | Intron | SBS | C | G |
| INDIA 8566 | Unknown | chr2 | 71243815 | IGR | SBS | C | G |
| INDIA 8566 | Unknown | chr5 | 171580039 | IGR | SBS | C | G |
| INDIA 8566 | ENST00000438818 | chr6 | 40712730 | 3'Flank | SBS | A | T |
| INDIA 8566 | Unknown | chr10 | 3703078 | IGR | SBS | C | G |
| INDIA 8566 | PCDH15 | chr10 | 55632488 | 5'Flank | SBS | G | C |
| INDIA 8566 | PYROXD2 | chr10 | 98390382 | Intron | SBS | C | G |
| INDIA 8566 | OR9I1 | chr11 | 58122257 | Intron | SBS | C | G |
| INDIA 8566 | Unknown | chr11 | 95363814 | IGR | SBS | C | G |
| INDIA 8566 | Unknown | chr11 | 128018852 | IGR | SBS | G | C |
| INDIA 8566 | Unknown | chr12 | 91077851 | IGR | SBS | C | G |
| INDIA 8566 | MYO16 | chr13 | 108922643 | Intron | SBS | C | G |
| INDIA 8566 | TMEM260 | chr14 | 56621585 | Silent | SBS | C | G |
| INDIA 8566 | Unknown | chr20 | 16810185 | IGR | SBS | C | G |
| INDIA 8566 | LINC01588 | chr14 | 50102170 | Intron | SBS | C | G |
| INDIA 8566 | LINC00504 | chr4 | 14864490 | Intron | SBS | G | C |
| INDIA 8566 | PALM2AKAP2 | chr9 | 109801482 | Intron | SBS | G | C |
| INDIA 8566 | Unknown | chr7 | 54013708 | IGR | SBS | G | C |
| INDIA 8566 | CHST1 | chr11 | 45661827 | Intron | SBS | G | C |
| INDIA 8566 | CCDC33 | chr15 | 74267987 | Intron | SBS | G | C |
| INDIA 8566 | Unknown | chr12 | 114911584 | IGR | SBS | G | C |
| INDIA 8566 | Unknown | chr20 | 6707742 | IGR | SBS | C | G |
| INDIA 8566 | RBMS3 | chr3 | 30006567 | 3'UTR | SBS | G | C |
| INDIA 8566 | FNIP1 | chr5 | 131750333 | Intron | SBS | G | C |
| INDIA 8566 | Unknown | chr5 | 171596343 | IGR | SBS | C | G |
| INDIA 8566 | ENST00000659436 | chr11 | 15865720 | RNA | SBS | G | C |
| INDIA 8566 | HMGA2 | chr12 | 65859191 | Intron | SBS | C | G |
| INDIA 8566 | Unknown | chr13 | 22422649 | IGR | SBS | C | G |
| INDIA 8566 | PTPRT | chr20 | 42150008 | Intron | SBS | C | G |
| INDIA 8566 | ENST00000652038 | chr20 | 57115917 | Intron | SBS | C | G |
| INDIA 8566 | Unknown | chr4 | 8940442 | IGR | DEL | A | - |
| INDIA 8566 | KLHL29 | chr2 | 23589350 | Intron | DEL | A | - |
| INDIA 8566 | TENM3 | chr4 | 182691219 | Intron | SBS | T | A |
| INDIA 8566 | LINC00293 | chr8 | 46905267 | Intron | SBS | C | G |
| INDIA 8566 | LINC01505 | chr9 | 106615666 | Intron | SBS | G | C |
| INDIA 8566 | GYS2 | chr12 | 21558286 | Frame_Shift_Del | DEL | G | - |
| INDIA 8566 | ENST00000690299 | chr20 | 61159210 | 3'Flank | SBS | G | C |
| INDIA 8566 | ENST00000657880 | chr2 | 122512239 | Intron | SBS | G | C |
| INDIA 8566 | PID1 | chr2 | 229110419 | Intron | SBS | A | T |
| INDIA 8566 | HGFAC | chr4 | 3437375 | 5'Flank | SBS | G | C |
| INDIA 8566 | ROPN1L | chr5 | 10451015 | Intron | SBS | G | C |
| INDIA 8566 | LINC01033 | chr5 | 54326826 | Intron | DEL | C | - |
| INDIA 8566 | Unknown | chr11 | 128310470 | IGR | SBS | G | T |
| INDIA 8566 | MIR4432HG | chr2 | 60378508 | Intron | DEL | C | - |
| INDIA 8566 | ARHGEF3 | chr3 | 56930831 | Intron | SBS | G | C |
| INDIA 8566 | ANKUB1 | chr3 | 149773394 | Intron | SBS | C | G |
| INDIA 8566 | Unknown | chr4 | 34063615 | IGR | SBS | G | C |
| INDIA 8566 | ADGRV1 | chr5 | 90701452 | Intron | SBS | G | C |
| INDIA 8566 | ENST00000523604 | chr8 | 141322262 | 5'Flank | SBS | C | G |
| INDIA 8566 | NFATC2 | chr20 | 51390947 | 3'UTR | SBS | G | C |
| INDIA 8566 | ENST00000438247 | chr2 | 1557590 | Intron | DEL | G | - |
| INDIA 8566 | Unknown | chr11 | 69537350 | IGR | SBS | G | C |
| INDIA 8566 | NEK9 | chr14 | 75082231 | 3'UTR | SBS | G | C |
| INDIA 8566 | Unknown | chr4 | 152558450 | IGR | SBS | G | T |
| INDIA 8566 | COL5A1 | chr9 | 134741118 | Intron | DEL | C | - |
| INDIA 8566 | CPM | chr12 | 68870096 | Intron | SBS | C | G |
| INDIA 8566 | ENST00000643167 | chr1 | 55722354 | Intron | SBS | C | G |
| INDIA 8566 | Unknown | chr5 | 61599489 | IGR | SBS | G | C |
| INDIA 8568 | GID4 | chr17 | 18067483 | 3'UTR | SBS | G | C |
| INDIA 8568 | RSU1 | chr10 | 16789650 | Intron | SBS | C | A |
| INDIA 8568 | ENST00000664549 | chr10 | 7074404 | Intron | SBS | A | T |
| INDIA 8568 | RASGEF1B | chr4 | 81883960 | Intron | SBS | G | C |
| INDIA 8568 | DMD | chrX | 31779435 | Intron | SBS | C | G |
| INDIA 8568 | ITFG1 | chr16 | 47247822 | Intron | INS | - | A |

| | | | | | | | |
|---|---|---|---|---|---|---|---|
| INDIA 8568 | FREM2 | chr13 | 38842467 | Intron | SBS | G | C |
| INDIA 8568 | ST8SIA2 | chr15 | 92448207 | Intron | SBS | G | C |
| INDIA 8568 | MID1 | chrX | 10541169 | Intron | SBS | C | G |
| INDIA 8568 | ENST00000419809 | chr2 | 67663928 | Intron | SBS | G | C |
| INDIA 8568 | EYA1 | chr8 | 71502870 | Intron | SBS | C | G |
| INDIA 8568 | Unknown | chr16 | 51817948 | IGR | SBS | C | A |
| INDIA 8568 | DLGAP2 | chr8 | 759751 | Intron | SBS | C | G |
| INDIA 8568 | ST8SIA2 | chr15 | 92448232 | Intron | SBS | G | C |
| INDIA 8568 | Unknown | chr4 | 28927132 | IGR | SBS | G | C |
| INDIA 8568 | GPC5 | chr13 | 92740732 | Intron | INS | - | A |
| INDIA 8568 | LAMA1 | chr18 | 7042159 | Missense_Mutation | SBS | G | C |
| INDIA 8568 | ENST00000454530 | chr2 | 3158978 | 3'Flank | SBS | C | G |
| INDIA 8568 | Unknown | chr2 | 83210231 | IGR | SBS | G | C |
| INDIA 8568 | UNC5C | chr4 | 95465105 | Intron | SBS | C | A |
| INDIA 8568 | CCDC22 | chrX | 49235469 | 5'Flank | SBS | C | G |
| INDIA 8568 | ENST00000636252 | chr13 | 108360477 | Intron | SBS | C | G |
| INDIA 8568 | C10orf143 | chr10 | 130050804 | Intron | SBS | G | C |
| INDIA 8568 | ENST00000666857 | chr6 | 14505802 | 3'Flank | SBS | G | C |
| INDIA 8568 | MKRN3 | chr15 | 23565445 | 5'Flank | SBS | G | C |
| INDIA 8568 | Unknown | chr4 | 170833683 | IGR | SBS | A | T |
| INDIA 8568 | Unknown | chr11 | 104630500 | IGR | SBS | A | T |
| INDIA 8568 | SNAP29 | chr22 | 20882381 | Intron | SBS | G | C |
| INDIA 8568 | Unknown | chr9 | 87377430 | IGR | SBS | T | G |
| INDIA 8568 | CNPPD1 | chr2 | 219168917 | 3'Flank | SBS | G | C |
| INDIA 8568 | PAX7 | chr1 | 18661099 | Intron | SBS | G | C |
| INDIA 8568 | Unknown | chr6 | 62513274 | IGR | SBS | T | G |
| INDIA 8568 | LRRC41 | chr1 | 46289112 | Intron | SBS | G | C |
| INDIA 8568 | THSD4 | chr15 | 71186324 | Intron | SBS | C | G |
| INDIA 8568 | KLF7 | chr2 | 207071326 | 3'Flank | SBS | C | G |
| INDIA 8568 | CCDC178 | chr18 | 33047882 | Intron | SBS | C | G |
| INDIA 8568 | NDUFB2 | chr7 | 140720186 | Intron | SBS | G | C |
| INDIA 8568 | FBXO36 | chr2 | 229947474 | Intron | SBS | C | G |
| INDIA 8568 | GRM3 | chr7 | 86839311 | Missense_Mutation | SBS | C | G |
| INDIA 8568 | ENST00000641428 | chr4 | 144178206 | Intron | SBS | G | C |
| INDIA 8568 | Unknown | chr1 | 228666712 | IGR | SBS | C | G |
| INDIA 8568 | ADGRD1 | chr12 | 131011333 | Intron | SBS | G | C |
| INDIA 8568 | ZFAND3 | chr6 | 38073010 | Intron | SBS | C | G |
| INDIA 8568 | ZIC1 | chr3 | 147498446 | Intron | SBS | T | A |
| INDIA 8568 | Unknown | chr22 | 49748468 | IGR | SBS | G | C |
| INDIA 8568 | IFT140 | chr16 | 1605092 | Intron | SBS | C | G |
| INDIA 8568 | Unknown | chr1 | 79903448 | IGR | SBS | C | G |
| INDIA 8568 | ENST00000496674 | chr3 | 78275623 | Intron | SBS | C | G |
| INDIA 8568 | Unknown | chr7 | 66969577 | IGR | DEL | GGACAG | - |
| INDIA 8568 | MAP3K21 | chr1 | 233375959 | Silent | SBS | C | G |
| INDIA 8568 | ENST00000420237 | chr1 | 219219607 | 3'Flank | SBS | C | A |
| INDIA 8568 | DIS3L2 | chr2 | 232235774 | Intron | DEL | TTGGT | - |
| INDIA 8568 | PTCHD4 | chr6 | 47931524 | Intron | SBS | C | G |
| INDIA 8568 | ENST00000661028 | chr18 | 68492694 | Intron | SBS | A | C |
| INDIA 8568 | NR6A1 | chr9 | 124558236 | Intron | SBS | G | C |
| INDIA 8568 | CIT | chr12 | 119817126 | Intron | SBS | G | C |
| INDIA 8568 | HHLA2 | chr3 | 108296073 | 5'Flank | SBS | T | A |
| INDIA 8568 | ENST00000442996 | chr2 | 174741890 | Intron | SBS | C | G |
| INDIA 8568 | ENST00000609699 | chr5 | 55041816 | Intron | SBS | G | C |
| INDIA 8568 | PGAP4 | chr9 | 101480198 | Intron | SBS | A | C |
| INDIA 8568 | TAFA2 | chr12 | 61908966 | Intron | SBS | G | C |
| INDIA 8568 | PENK | chr8 | 56447915 | 5'Flank | SBS | C | G |
| INDIA 8568 | SCUBE1 | chr22 | 43321518 | Intron | SBS | G | C |
| INDIA 8568 | CITED2 | chr6 | 139368206 | 3'Flank | SBS | C | G |
| INDIA 8568 | TENM3 | chr4 | 182424677 | Intron | SBS | G | C |
| INDIA 8568 | TDRD6 | chr6 | 46692977 | Missense_Mutation | SBS | C | G |
| INDIA 8568 | FSTL1 | chr3 | 120428161 | Intron | INS | - | T |
| INDIA 8568 | LINC02818 | chr1 | 179824900 | 3'Flank | SBS | C | A |
| INDIA 8568 | KDM5B | chr1 | 202739110 | Intron | DEL | TGTC | - |
| INDIA 8568 | LRRN2 | chr1 | 204643658 | Intron | SBS | C | G |
| INDIA 8568 | Unknown | chr1 | 209815214 | IGR | SBS | C | G |
| INDIA 8568 | SLC24A2 | chr9 | 19530709 | Intron | DEL | AATC | - |

| | | | | | | | |
|---|---|---|---|---|---|---|---|
| INDIA 8568 | Unknown | chr12 | 115354287 | IGR | INS | - | TA |
| INDIA 8568 | Unknown | chr13 | 105212386 | IGR | SBS | G | C |
| INDIA 8568 | NAB1 | chr2 | 190679760 | Intron | SBS | C | G |
| INDIA 8568 | Unknown | chr2 | 238842241 | IGR | SBS | C | G |
| INDIA 8568 | MTCH1 | chr6 | 36986792 | 5'Flank | SBS | C | G |
| INDIA 8568 | RARB | chr3 | 25382074 | Intron | SBS | C | G |
| INDIA 8568 | Unknown | chr16 | 35794863 | IGR | DEL | A | - |
| INDIA 8568 | IL1RAP | chr3 | 190588618 | Intron | SBS | C | G |
| INDIA 8568 | Unknown | chr7 | 67989394 | IGR | SBS | T | A |
| INDIA 8568 | H3C8 | chr6 | 26271550 | 5'Flank | SBS | G | C |
| INDIA 8568 | Unknown | chr11 | 69862351 | IGR | SBS | G | C |
| INDIA 8568 | THRB | chr3 | 24227370 | Intron | SBS | G | C |
| INDIA 8568 | ENST00000435967 | chr7 | 8323578 | Intron | SBS | G | C |
| INDIA 8568 | RFTN1 | chr3 | 16471962 | Intron | SBS | C | G |
| INDIA 8568 | LINGO2 | chr9 | 28940749 | Intron | SBS | A | T |
| INDIA 8568 | Unknown | chr1 | 172630102 | IGR | SBS | G | C |
| INDIA 8568 | TANC1 | chr2 | 159155462 | Intron | DEL | TGGAG | - |
| INDIA 8568 | Unknown | chr6 | 23871755 | IGR | SBS | G | C |
| INDIA 8568 | Unknown | chr4 | 137392016 | IGR | SBS | T | A |
| INDIA 8568 | CRAMP1 | chr16 | 1648018 | Intron | SBS | C | G |
| INDIA 8568 | Unknown | chr8 | 76151316 | IGR | SBS | G | C |
| INDIA 8568 | STPG2 | chr4 | 97300296 | Intron | SBS | C | G |
| INDIA 8568 | ARMC2 | chr6 | 108957643 | Intron | SBS | C | G |
| INDIA 8568 | Unknown | chr17 | 40612977 | IGR | SBS | C | G |
| INDIA 8569 | PYCR2 | chr1 | 225921476 | Intron | SBS | C | G |
| INDIA 8569 | STK32B | chr4 | 5367432 | Intron | SBS | C | G |
| INDIA 8569 | LINC00457 | chr13 | 34557443 | Intron | SBS | T | G |
| INDIA 8569 | CATSPERE | chr1 | 244593116 | Intron | SBS | T | G |
| INDIA 8569 | LINGO2 | chr9 | 29032186 | Intron | SBS | G | C |
| INDIA 8569 | CDH12 | chr5 | 21896043 | Intron | SBS | T | G |
| INDIA 8569 | PYCR2 | chr1 | 225923644 | Intron | SBS | C | G |
| INDIA 8569 | FHIT | chr3 | 60439727 | Intron | SBS | C | G |
| INDIA 8569 | Unknown | chr10 | 33670067 | IGR | SBS | G | C |
| INDIA 8569 | KCTD1 | chr18 | 26468050 | Intron | DEL | C | - |
| INDIA 8569 | VGLL4 | chr3 | 11641856 | Intron | SBS | C | A |
| INDIA 8569 | Unknown | chr22 | 45123665 | IGR | SBS | C | G |
| INDIA 8569 | EYS | chr6 | 64834086 | Intron | SBS | C | G |
| INDIA 8569 | TAF4 | chr20 | 61968216 | Intron | SBS | T | G |
| INDIA 8569 | LINC01426 | chr21 | 34774969 | Intron | SBS | G | C |
| INDIA 8569 | Unknown | chr6 | 91228597 | IGR | SBS | A | C |
| INDIA 8569 | Unknown | chr14 | 45699149 | IGR | SBS | C | G |
| INDIA 8569 | ENST00000655430 | chr16 | 49123948 | Intron | SBS | G | C |
| INDIA 8569 | FRMD3 | chr9 | 83333655 | Intron | SBS | A | T |
| INDIA 8569 | DUSP10 | chr1 | 221739253 | Missense_Mutation | SBS | C | G |
| INDIA 8569 | CDC42BPA | chr1 | 227073293 | Intron | SBS | G | C |
| INDIA 8569 | ADCY10 | chr1 | 167902327 | Intron | SBS | C | A |
| INDIA 8569 | VANGL2 | chr1 | 160406643 | Intron | SBS | C | G |
| INDIA 8569 | CCDC88C | chr14 | 91295711 | Intron | SBS | C | G |
| INDIA 8569 | ARF1 | chr1 | 228093472 | Intron | DEL | CAGGGC | - |
| INDIA 8569 | ENST00000452528 | chr1 | 157938053 | Intron | SBS | C | G |
| INDIA 8569 | ACAD11 | chr3 | 132616033 | Intron | SBS | C | G |
| INDIA 8569 | ENST00000519555 | chr8 | 6072605 | Intron | SBS | T | G |
| INDIA 8569 | Unknown | chr14 | 83759359 | IGR | SBS | G | C |
| INDIA 8569 | ALDH1A1 | chr9 | 73021154 | Intron | SBS | G | T |
| INDIA 8569 | MACROD2 | chr20 | 14804939 | Intron | DEL | GTA | - |
| INDIA 8569 | Unknown | chr18 | 26415037 | IGR | SBS | C | G |
| INDIA 8569 | Unknown | chr3 | 138968812 | IGR | SBS | G | C |
| INDIA 8569 | ZNF516 | chr18 | 76401444 | Intron | DEL | C | - |
| INDIA 8569 | DGKI | chr7 | 137794707 | Intron | DEL | G | - |
| INDIA 8569 | ESRRG | chr1 | 216542826 | Intron | SBS | C | G |
| INDIA 8569 | CLDN10-AS1 | chr13 | 95530459 | Intron | SBS | G | T |
| INDIA 8569 | OTX2-AS1 | chr14 | 57002973 | Intron | DEL | C | - |
| INDIA 8569 | STAC | chr3 | 36409302 | Intron | SBS | C | G |
| INDIA 8569 | PRRX1 | chr1 | 170714775 | Intron | SBS | C | G |
| INDIA 8569 | FBXO6 | chr1 | 11673683 | Missense_Mutation | SBS | C | A |
| INDIA 8569 | DDX56 | chr7 | 44570282 | Intron | SBS | G | C |

| | | | | | | | |
|---|---|---|---|---|---|---|---|
| INDIA 8569 | CFAP161 | chr15 | 81096814 | 5'Flank | SBS | C | G |
| INDIA 8569 | SSUH2 | chr3 | 8669074 | Intron | SBS | C | G |
| INDIA 8569 | ERG | chr21 | 38649729 | Intron | SBS | T | G |
| INDIA 8569 | CACNA1I | chr22 | 39616008 | Intron | DEL | G | - |
| INDIA 8569 | GAPDHS | chr19 | 35543209 | Intron | SBS | C | G |
| INDIA 8569 | TOR1AIP2 | chr1 | 179867243 | Intron | SBS | G | C |
| INDIA 8569 | HIP1 | chr7 | 75689031 | Intron | SBS | G | C |
| INDIA 8569 | TEX22 | chr14 | 105411814 | Missense_Mutation | SBS | C | G |
| INDIA 8569 | DOCK8-AS2 | chr9 | 471890 | Intron | SBS | G | C |
| INDIA 8569 | TNRC6B | chr22 | 40307969 | Intron | SBS | C | G |
| INDIA 8569 | Unknown | chr2 | 62526080 | IGR | SBS | C | G |
| INDIA 8569 | Unknown | chr10 | 60771252 | IGR | SBS | C | G |
| INDIA 8569 | ENST00000657880 | chr2 | 122856471 | Intron | SBS | G | C |
| INDIA 8569 | Unknown | chr5 | 62168643 | IGR | SBS | G | C |
| INDIA 8569 | NOTCH1 | chr9 | 136521628 | Intron | SBS | C | G |
| INDIA 8569 | Unknown | chr16 | 54356452 | IGR | SBS | G | C |
| INDIA 8569 | Unknown | chrX | 149322268 | IGR | SBS | C | A |
| INDIA 8569 | DISP1 | chr1 | 223004400 | Silent | SBS | G | C |
| INDIA 8569 | Unknown | chr2 | 41545816 | IGR | INS | - | C |
| INDIA 8569 | NKRF | chrX | 119603743 | Intron | SBS | G | C |
| INDIA 8569 | MIR3681HG | chr2 | 12516520 | Intron | SBS | C | G |
| INDIA 8569 | USH2A | chr1 | 216358369 | Intron | SBS | G | C |
| INDIA 8569 | EDNRA | chr4 | 147482914 | Intron | SBS | G | C |
| INDIA 8569 | Unknown | chr9 | 117693130 | IGR | SBS | G | C |
| INDIA 8569 | GABRB3 | chr15 | 26661582 | Intron | SBS | G | C |
| INDIA 8569 | SAMD12 | chr8 | 118261527 | Intron | SBS | C | A |
| INDIA 8569 | Unknown | chr13 | 111911338 | IGR | SBS | G | T |
| INDIA 8569 | ARHGAP26 | chr5 | 143206011 | Intron | INS | - | A |
| INDIA 8569 | ALDH8A1 | chr6 | 134932556 | Intron | SBS | C | G |
| INDIA 8569 | Unknown | chr10 | 128744392 | IGR | SBS | C | G |
| INDIA 8569 | KCNQ1 | chr11 | 2459090 | Intron | DEL | GGG | - |
| INDIA 8569 | ENST00000557467 | chr14 | 56846754 | Intron | SBS | G | C |
| INDIA 8569 | Unknown | chr3 | 94217146 | IGR | SBS | C | G |
| INDIA 8569 | SLC35G1 | chr10 | 93903480 | 3'UTR | SBS | C | G |
| INDIA 8569 | Unknown | chr13 | 67950725 | IGR | SBS | C | G |
| INDIA 8569 | Unknown | chr18 | 65087913 | IGR | SBS | C | G |
| INDIA 8569 | Unknown | chr2 | 139291218 | IGR | SBS | C | A |
| INDIA 8569 | ENST00000613161 | chr15 | 50863367 | Intron | SBS | G | C |
| INDIA 8569 | CNTN4 | chr3 | 2402021 | Intron | SBS | C | G |
| INDIA 8569 | MROH1 | chr8 | 144150304 | Intron | SBS | G | C |
| INDIA 8569 | Unknown | chr2 | 234177416 | IGR | SBS | C | G |
| INDIA 8569 | MYRIP | chr3 | 40185781 | Intron | SBS | G | C |
| INDIA 8569 | TLN2 | chr15 | 62734909 | Intron | SBS | C | G |
| INDIA 8569 | Unknown | chr18 | 40254745 | IGR | SBS | G | C |
| INDIA 8569 | MIR9-3HG | chr15 | 89419539 | Intron | SBS | G | T |
| INDIA 8569 | Unknown | chr2 | 67859836 | IGR | SBS | G | C |
| INDIA 8569 | SLC5A12 | chr11 | 26701136 | Intron | SBS | G | C |
| INDIA 8569 | Unknown | chr16 | 17752401 | IGR | SBS | C | G |
| INDIA 8569 | DNAAF5 | chr7 | 774038 | Intron | DEL | C | - |
| INDIA 8569 | Unknown | chr6 | 257356 | IGR | SBS | C | G |
| INDIA 8569 | Unknown | chr10 | 36263109 | IGR | SBS | T | A |
| INDIA 8570 | Unknown | chr12 | 90211130 | IGR | SBS | C | G |
| INDIA 8570 | ELF3 | chr1 | 202012977 | Nonsense_Mutation | SBS | C | G |
| INDIA 8570 | Unknown | chr1 | 212454835 | IGR | SBS | G | C |
| INDIA 8570 | ADORA1 | chr1 | 203151568 | Intron | SBS | G | C |
| INDIA 8570 | Unknown | chr13 | 106391761 | IGR | SBS | G | C |
| INDIA 8570 | CTPS2 | chrX | 16678951 | Intron | SBS | G | C |
| INDIA 8570 | Unknown | chrX | 147066840 | IGR | SBS | G | C |
| INDIA 8570 | Unknown | chr1 | 172501999 | IGR | SBS | G | C |
| INDIA 8570 | Unknown | chr5 | 67936068 | IGR | SBS | G | C |
| INDIA 8570 | Unknown | chr5 | 75735311 | IGR | SBS | G | C |
| INDIA 8570 | GABBR2 | chr9 | 98477000 | Intron | SBS | C | G |
| INDIA 8570 | COL4A6 | chrX | 108396458 | Intron | SBS | C | G |
| INDIA 8570 | STIM1 | chr11 | 3889717 | Intron | SBS | C | G |
| INDIA 8570 | Unknown | chr3 | 188930596 | IGR | SBS | G | C |
| INDIA 8570 | Unknown | chr8 | 37301790 | IGR | SBS | G | C |

| | | | | | | | |
|---|---|---|---|---|---|---|---|
| INDIA 8570 | GPBP1 | chr5 | 57245353 | Intron | SBS | G | C |
| INDIA 8570 | IL2RA | chr10 | 6063686 | 5'Flank | SBS | C | G |
| INDIA 8570 | Unknown | chr18 | 40647666 | IGR | DEL | T | - |
| INDIA 8570 | DIRC3 | chr2 | 217736593 | Intron | SBS | G | C |
| INDIA 8570 | SLC35E3 | chr12 | 68751985 | Intron | SBS | G | C |
| INDIA 8570 | TSHZ3-AS1 | chr19 | 31365677 | Intron | DEL | A | - |
| INDIA 8570 | ENST00000670789 | chr5 | 52670539 | 3'Flank | SBS | G | C |
| INDIA 8570 | IGSF6 | chr16 | 21645701 | Intron | SBS | C | G |
| INDIA 8570 | Unknown | chr21 | 23039895 | IGR | SBS | C | G |
| INDIA 8570 | MTSS1 | chr8 | 124710936 | Intron | SBS | C | G |
| INDIA 8570 | Unknown | chr2 | 75321879 | IGR | SBS | G | C |
| INDIA 8570 | FOXP1 | chr3 | 71304884 | Intron | SBS | C | G |
| INDIA 8570 | LINC01170 | chr5 | 124240855 | Intron | SBS | G | C |
| INDIA 8570 | SPOCK1 | chr5 | 137332872 | Intron | SBS | G | C |
| INDIA 8570 | SH3TC2 | chr5 | 149060545 | Intron | SBS | C | G |
| INDIA 8570 | RAPGEF5 | chr7 | 22235019 | Intron | SBS | C | G |
| INDIA 8570 | KC6 | chr18 | 41592499 | Intron | SBS | T | A |
| INDIA 8570 | Unknown | chr19 | 15594409 | IGR | SBS | A | C |
| INDIA 8570 | DAB1 | chr1 | 57460429 | Intron | SBS | G | C |
| INDIA 8570 | RRAS | chr19 | 49636361 | Intron | SBS | G | C |
| INDIA 8570 | Unknown | chr4 | 33446914 | IGR | SBS | T | A |
| INDIA 8570 | MRPS9 | chr2 | 105059355 | Intron | SBS | G | C |
| INDIA 8570 | PLCL2 | chr3 | 16906857 | Intron | SBS | G | C |
| INDIA 8570 | Unknown | chr18 | 78207603 | IGR | SBS | C | G |
| INDIA 8570 | ENST00000554029 | chr14 | 38003960 | Intron | SBS | G | C |
| INDIA 8570 | ENST00000669923 | chr16 | 52237587 | 5'Flank | SBS | G | C |
| INDIA 8570 | PRKCA | chr17 | 66589139 | Intron | SBS | A | C |
| INDIA 8570 | LINC02119 | chr5 | 38094400 | Intron | SBS | G | C |
| INDIA 8570 | MSRB3 | chr12 | 65432539 | Intron | SBS | C | G |
| INDIA 8570 | Unknown | chr7 | 129540500 | IGR | SBS | C | G |
| INDIA 8570 | GRHL2 | chr8 | 101632393 | Intron | SBS | C | G |
| INDIA 8570 | LINC01568 | chr16 | 73387299 | Intron | SBS | C | G |
| INDIA 8570 | DFFA | chr1 | 10461557 | Nonsense_Mutation | SBS | G | C |
| INDIA 8570 | Unknown | chr4 | 31234262 | IGR | SBS | C | G |
| INDIA 8570 | LEF1 | chr4 | 108103350 | Intron | SBS | C | G |
| INDIA 8570 | RXFP1 | chr4 | 158406900 | Intron | SBS | C | G |
| INDIA 8570 | Unknown | chr6 | 18751133 | IGR | SBS | C | G |
| INDIA 8570 | Unknown | chr6 | 55964263 | IGR | SBS | G | C |
| INDIA 8570 | CCNJ | chr10 | 96044596 | Intron | SBS | G | C |
| INDIA 8570 | Unknown | chr11 | 38784442 | IGR | SBS | C | G |
| INDIA 8570 | NRXN3 | chr14 | 78422839 | Intron | SBS | G | C |
| INDIA 8570 | SLC13A2 | chr17 | 28493175 | Intron | SBS | C | G |
| INDIA 8570 | ATP9B | chr18 | 79113757 | Intron | SBS | C | G |
| INDIA 8570 | C2orf88 | chr2 | 190181819 | Intron | INS | - | T |
| INDIA 8570 | RN7SKP293 | chr6 | 12408713 | 5'Flank | SBS | C | G |
| INDIA 8570 | LINC01010 | chr6 | 134405643 | Intron | SBS | G | C |
| INDIA 8570 | ENST00000643090 | chr7 | 17016503 | Intron | SBS | C | G |
| INDIA 8570 | ENST00000667254 | chr8 | 37561974 | Intron | SBS | G | C |
| INDIA 8570 | PHACTR1 | chr6 | 13269403 | Intron | SBS | G | C |
| INDIA 8570 | LINC00964 | chr8 | 124860483 | Intron | SBS | G | C |
| INDIA 8570 | CCNF | chr16 | 2446496 | Intron | SBS | G | C |
| INDIA 8570 | CASS4 | chr20 | 56450646 | Frame_Shift_Del | DEL | G | - |
| INDIA 8570 | Unknown | chr2 | 133635252 | IGR | SBS | G | C |
| INDIA 8570 | PMFBP1 | chr16 | 72125903 | Intron | SBS | T | A |
| INDIA 8570 | FHL3 | chr1 | 37995048 | 3'Flank | SBS | C | G |
| INDIA 8570 | ANKRD35 | chr1 | 145866964 | Intron | SBS | G | C |
| INDIA 8570 | Unknown | chr2 | 4500545 | IGR | SBS | C | A |
| INDIA 8570 | Unknown | chr2 | 4500546 | IGR | SBS | C | A |
| INDIA 8570 | ENST00000647894 | chr2 | 13294147 | Intron | SBS | G | C |
| INDIA 8570 | AAK1 | chr2 | 69454164 | 3'Flank | SBS | C | G |
| INDIA 8570 | CLSTN2 | chr3 | 140442953 | Intron | SBS | G | C |
| INDIA 8570 | KIT | chr4 | 54667024 | Intron | SBS | C | G |
| INDIA 8570 | STAG3 | chr7 | 100204551 | Intron | SBS | G | C |
| INDIA 8570 | TOX | chr8 | 58865117 | Intron | SBS | G | C |
| INDIA 8570 | TG | chr8 | 133004658 | Intron | SBS | G | C |
| INDIA 8570 | PPFIBP2 | chr11 | 7537958 | Intron | SBS | G | C |

| | | | | | | | |
|---|---|---|---|---|---|---|---|
| INDIA 8570 | NEAT1 | chr11 | 65424000 | RNA | SBS | G | C |
| INDIA 8570 | SOX5-AS1 | chr12 | 24222919 | 5'Flank | SBS | G | C |
| INDIA 8570 | PPM1H | chr12 | 62808506 | Intron | SBS | G | C |
| INDIA 8570 | EML1 | chr14 | 99860216 | Intron | SBS | C | G |
| INDIA 8570 | PLCB2 | chr15 | 40293534 | Intron | SBS | C | G |
| INDIA 8570 | PEPD | chr19 | 33386488 | 3'Flank | SBS | C | G |
| INDIA 8570 | SIPA1L3 | chr19 | 38148459 | Intron | SBS | C | G |
| INDIA 8570 | BZW2 | chr7 | 16668069 | Intron | SBS | G | C |
| INDIA 8570 | TMEM125 | chr1 | 43274218 | 3'Flank | SBS | G | C |
| INDIA 8570 | FLVCR1 | chr1 | 212856957 | 5'Flank | SBS | C | G |
| INDIA 8570 | PCDH9 | chr13 | 66926770 | Intron | SBS | C | G |
| INDIA 8571 | TSTD2 | chr9 | 97614854 | Intron | SBS | G | C |
| INDIA 8571 | LHX4 | chr1 | 180254162 | Intron | SBS | G | C |
| INDIA 8571 | MTTP | chr4 | 99584716 | Intron | SBS | C | G |
| INDIA 8571 | Unknown | chr15 | 80605604 | IGR | SBS | G | C |
| INDIA 8571 | UAP1L1 | chr9 | 137083381 | 3'UTR | SBS | G | C |
| INDIA 8571 | MAILR | chr8 | 102921940 | Intron | SBS | C | G |
| INDIA 8571 | TMEM246-AS1 | chr9 | 101466702 | 5'Flank | SBS | T | G |
| INDIA 8571 | LRRC20 | chr10 | 70297523 | 3'Flank | SBS | G | C |
| INDIA 8571 | ENST00000522743 | chr8 | 94099502 | Intron | SBS | C | G |
| INDIA 8571 | Unknown | chr9 | 107698736 | IGR | SBS | A | C |
| INDIA 8571 | SLC41A1 | chr1 | 205799940 | Intron | SBS | G | C |
| INDIA 8571 | Unknown | chr1 | 208004026 | IGR | SBS | C | G |
| INDIA 8571 | MYO1D | chr17 | 32874599 | Intron | SBS | C | G |
| INDIA 8571 | GPAM | chr10 | 112151314 | 3'UTR | SBS | G | C |
| INDIA 8571 | SCAMP1 | chr5 | 78359921 | 5'Flank | SBS | C | G |
| INDIA 8571 | MIR4290HG | chr9 | 90040611 | Intron | SBS | G | C |
| INDIA 8571 | COLEC12 | chr18 | 314376 | 3'Flank | SBS | G | C |
| INDIA 8571 | HECW1 | chr7 | 43121276 | Intron | SBS | G | C |
| INDIA 8571 | SLC19A2 | chr1 | 169486381 | 5'Flank | SBS | G | C |
| INDIA 8571 | CASC2 | chr10 | 118117720 | Intron | SBS | G | C |
| INDIA 8571 | Unknown | chr1 | 242695910 | IGR | SBS | C | G |
| INDIA 8571 | VAV1 | chr19 | 6791313 | Intron | SBS | G | C |
| INDIA 8571 | POU2F1 | chr1 | 167275640 | Intron | SBS | T | A |
| INDIA 8571 | Unknown | chr22 | 49698499 | IGR | SBS | G | C |
| INDIA 8571 | RNF180 | chr5 | 64241194 | Intron | SBS | C | A |
| INDIA 8571 | ARHGEF6 | chrX | 136689670 | Intron | SBS | C | G |
| INDIA 8571 | PDS5B | chr13 | 32585894 | 5'Flank | SBS | C | G |
| INDIA 8571 | SDK1 | chr7 | 3374410 | Intron | SBS | T | G |
| INDIA 8571 | ST8SIA4 | chr5 | 100867993 | Intron | SBS | G | C |
| INDIA 8571 | SOX9 | chr17 | 72122273 | Intron | SBS | G | C |
| INDIA 8571 | Unknown | chr5 | 25606856 | IGR | SBS | G | C |
| INDIA 8571 | HIVEP3 | chr1 | 41821224 | Intron | SBS | C | G |
| INDIA 8571 | C6orf62 | chr6 | 24715290 | Intron | SBS | C | G |
| INDIA 8571 | Unknown | chr10 | 83694530 | IGR | SBS | G | T |
| INDIA 8571 | LINC01804 | chr2 | 15695218 | Intron | SBS | G | C |
| INDIA 8571 | Unknown | chr7 | 52205886 | IGR | SBS | C | G |
| INDIA 8571 | Unknown | chr7 | 97047897 | IGR | SBS | G | C |
| INDIA 8571 | UNC5D | chr8 | 35497485 | Intron | SBS | G | C |
| INDIA 8571 | Unknown | chr2 | 65146903 | IGR | DEL | G | - |
| INDIA 8571 | ETV6 | chr12 | 11837453 | Intron | SBS | C | G |
| INDIA 8571 | Unknown | chrX | 126614282 | IGR | SBS | G | C |
| INDIA 8571 | Unknown | chr11 | 86098080 | IGR | SBS | C | G |
| INDIA 8571 | MSI2 | chr17 | 57461127 | Intron | SBS | G | C |
| INDIA 8571 | LINC01004 | chr7 | 104947997 | 3'Flank | SBS | G | C |
| INDIA 8571 | DEDD | chr1 | 161129240 | Intron | SBS | C | A |
| INDIA 8571 | Unknown | chr20 | 63225021 | IGR | SBS | G | C |
| INDIA 8571 | COL4A6 | chrX | 108175423 | Intron | SBS | C | G |
| INDIA 8571 | GABRA4 | chr4 | 46993109 | Intron | SBS | A | T |
| INDIA 8571 | ENST00000542489 | chr12 | 21756761 | 5'Flank | INS | - | TTCA |
| INDIA 8571 | ENST00000643262 | chr16 | 51416903 | Intron | SBS | C | G |
| INDIA 8571 | Unknown | chr18 | 22234365 | IGR | SBS | C | G |
| INDIA 8571 | ENST00000660753 | chr4 | 13519581 | 3'Flank | SBS | C | G |
| INDIA 8571 | Unknown | chr4 | 67141678 | IGR | SBS | G | C |
| INDIA 8571 | Unknown | chr14 | 86310909 | IGR | SBS | G | C |
| INDIA 8571 | RSF1 | chr11 | 77705669 | Intron | SBS | G | C |

| | | | | | | | |
|---|---|---|---|---|---|---|---|
| INDIA 8571 | Unknown | chr4 | 27593428 | IGR | SBS | G | C |
| INDIA 8571 | NUP210L | chr1 | 154153978 | Intron | SBS | C | G |
| INDIA 8571 | CDH4 | chr20 | 61473883 | Intron | SBS | G | C |
| INDIA 8571 | LINC01709 | chr1 | 101706341 | Intron | INS | - | CT |
| INDIA 8571 | ENST00000655120 | chr16 | 85694891 | 5'Flank | SBS | A | T |
| INDIA 8571 | Unknown | chr1 | 191320210 | IGR | SBS | G | C |
| INDIA 8571 | PLEKHG4B | chr5 | 173581 | Intron | SBS | C | G |
| INDIA 8571 | HECW1 | chr7 | 43244514 | Intron | SBS | C | G |
| INDIA 8571 | PKIA | chr8 | 78568883 | Intron | SBS | G | C |
| INDIA 8571 | SORCS3 | chr10 | 104870520 | Intron | SBS | C | G |
| INDIA 8571 | PRMT3 | chr11 | 20508407 | Silent | SBS | C | G |
| INDIA 8571 | TRPC4 | chr13 | 37752468 | Intron | SBS | C | G |
| INDIA 8571 | VSX2 | chr14 | 74246854 | Intron | SBS | G | C |
| INDIA 8571 | DRAIC | chr15 | 69696119 | 3'Flank | SBS | C | G |
| INDIA 8571 | HIVEP3 | chr1 | 41821322 | Intron | SBS | C | G |
| INDIA 8571 | LINC00709 | chr10 | 9284621 | Intron | SBS | C | G |
| INDIA 8571 | MAP4K3 | chr2 | 39369832 | Intron | SBS | C | G |
| INDIA 8571 | BCL2L1 | chr20 | 31717745 | Intron | SBS | C | G |
| INDIA 8571 | STEAP4 | chr7 | 88301235 | Intron | SBS | G | C |
| INDIA 8571 | SLC25A47 | chr14 | 100326045 | Intron | SBS | C | A |
| INDIA 8571 | ENST00000654998 | chr1 | 227796149 | 3'Flank | SBS | G | C |
| INDIA 8571 | FMN2 | chr1 | 240370531 | Intron | SBS | C | G |
| INDIA 8571 | DNAJC13 | chr3 | 132465976 | Intron | SBS | C | G |
| INDIA 8571 | Unknown | chr21 | 23197802 | IGR | SBS | G | C |
| INDIA 8571 | EPHB1 | chr3 | 134823066 | Intron | SBS | C | G |
| INDIA 8571 | WWTR1 | chr3 | 149531979 | Intron | SBS | C | G |
| INDIA 8571 | Unknown | chr20 | 33697126 | IGR | SBS | G | C |
| INDIA 8571 | ZEB2 | chr2 | 144362417 | 3'Flank | SBS | C | G |
| INDIA 8571 | MORC1 | chr3 | 108993966 | Intron | SBS | C | G |
| INDIA 8571 | AIG1 | chr6 | 143181452 | Intron | SBS | C | G |
| INDIA 8571 | Unknown | chr13 | 68443886 | IGR | DEL | A | - |
| INDIA 8571 | PCNA | chr20 | 5112877 | 3'Flank | SBS | G | C |
| INDIA 8571 | ENST00000663196 | chr20 | 53738633 | 5'Flank | DEL | AGGAAAGTA/ | - |
| INDIA 8571 | KAZN | chr1 | 13895601 | Intron | SBS | C | G |
| INDIA 8571 | ZMAT4 | chr8 | 40562641 | Intron | SBS | C | G |
| INDIA 8571 | Unknown | chr2 | 136174651 | IGR | SBS | C | G |
| INDIA 8571 | Unknown | chr20 | 21910671 | IGR | SBS | C | G |
| INDIA 8572 | LRCH1 | chr13 | 46595368 | Intron | SBS | C | G |
| INDIA 8572 | ENST00000446028 | chrX | 56618473 | RNA | SBS | G | C |
| INDIA 8572 | DUS2 | chr16 | 68079013 | 3'UTR | SBS | C | G |
| INDIA 8572 | KCNJ6 | chr21 | 37686237 | Intron | SBS | C | G |
| INDIA 8572 | RBM10 | chrX | 47179957 | In_Frame_Del | DEL | TCC | - |
| INDIA 8572 | Unknown | chr4 | 3731804 | IGR | SBS | C | G |
| INDIA 8572 | CENPP | chr9 | 92324101 | 5'Flank | SBS | C | G |
| INDIA 8572 | ENST00000701975 | chr5 | 57689566 | Intron | SBS | C | G |
| INDIA 8572 | PPP1R12A | chr12 | 79933308 | Intron | SBS | G | C |
| INDIA 8572 | LIN7B | chr19 | 49118521 | 3'Flank | SBS | G | C |
| INDIA 8572 | ENST00000559600 | chr15 | 45709142 | Intron | SBS | C | G |
| INDIA 8572 | ENST00000643626 | chr16 | 51321406 | Intron | SBS | C | G |
| INDIA 8572 | Unknown | chrX | 35913076 | IGR | SBS | C | G |
| INDIA 8572 | ENST00000653849 | chrX | 129259488 | Intron | SBS | C | G |
| INDIA 8572 | GALNT9 | chr12 | 132297121 | Intron | SBS | C | G |
| INDIA 8572 | TACC2 | chr10 | 121994772 | Intron | SBS | G | C |
| INDIA 8572 | Unknown | chr7 | 12126995 | IGR | SBS | G | C |
| INDIA 8572 | XKR6 | chr8 | 10957257 | Intron | SBS | C | G |
| INDIA 8572 | LINC01435 | chr10 | 108052774 | Intron | SBS | C | G |
| INDIA 8572 | ADAM12 | chr10 | 126101125 | Missense_Mutation | SBS | C | G |
| INDIA 8572 | TTLL1 | chr22 | 43068906 | Intron | SBS | C | G |
| INDIA 8572 | Unknown | chrX | 133120199 | IGR | SBS | G | C |
| INDIA 8572 | ENST00000450811 | chr1 | 62905946 | 3'Flank | SBS | C | G |
| INDIA 8572 | Unknown | chr11 | 41962906 | IGR | SBS | G | C |
| INDIA 8572 | Unknown | chr12 | 104152599 | IGR | SBS | C | G |
| INDIA 8572 | TPK1 | chr7 | 144695320 | Intron | SBS | C | G |
| INDIA 8572 | RBM20 | chr10 | 110684980 | Intron | SBS | G | C |
| INDIA 8572 | Unknown | chr6 | 148617356 | IGR | SBS | G | C |
| INDIA 8572 | CENPP | chr9 | 92324784 | 5'Flank | SBS | C | G |

| | | | | | | | |
|---|---|---|---|---|---|---|---|
| INDIA 8572 | RCAN1 | chr21 | 34611763 | Intron | SBS | G | C |
| INDIA 8572 | SORBS2 | chr4 | 185951057 | Intron | SBS | C | G |
| INDIA 8572 | Unknown | chr21 | 17420595 | IGR | SBS | G | C |
| INDIA 8572 | RBM20 | chr10 | 110685044 | Intron | SBS | G | C |
| INDIA 8572 | Unknown | chr9 | 110840611 | IGR | SBS | G | C |
| INDIA 8572 | ENST00000443714 | chr7 | 45456588 | 5'Flank | SBS | G | C |
| INDIA 8572 | SULT1B1 | chr4 | 69731855 | Intron | SBS | C | G |
| INDIA 8572 | ENST00000522129 | chr8 | 9383565 | Intron | SBS | C | G |
| INDIA 8572 | Unknown | chr8 | 64480813 | IGR | SBS | G | T |
| INDIA 8572 | BAALC | chr8 | 103192455 | Intron | SBS | C | G |
| INDIA 8572 | LINC02625 | chr10 | 61782147 | Intron | SBS | G | C |
| INDIA 8572 | ENST00000568150 | chr16 | 48631423 | Intron | DEL | C | - |
| INDIA 8572 | KCNQ2 | chr20 | 63402154 | 3'UTR | SBS | G | C |
| INDIA 8572 | KRT72 | chr12 | 52602997 | 5'Flank | SBS | C | G |
| INDIA 8572 | GS1-279B7.1 | chr1 | 185339251 | 5'Flank | SBS | C | G |
| INDIA 8572 | NBEAL2 | chr3 | 46997002 | Missense_Mutation | SBS | G | C |
| INDIA 8572 | ENST00000667321 | chr7 | 4642762 | 3'Flank | SBS | G | C |
| INDIA 8572 | Unknown | chr22 | 26154854 | IGR | SBS | G | C |
| INDIA 8572 | Unknown | chr7 | 130713076 | IGR | SBS | C | G |
| INDIA 8572 | GCLM | chr1 | 93907973 | Intron | SBS | C | G |
| INDIA 8572 | FAM118A | chr22 | 45318361 | Intron | SBS | C | G |
| INDIA 8572 | DOCK5 | chr8 | 25276802 | Intron | SBS | C | G |
| INDIA 8572 | Unknown | chr9 | 74356200 | IGR | SBS | C | G |
| INDIA 8572 | SLC16A13 | chr17 | 7040639 | 3'Flank | SBS | G | C |
| INDIA 8572 | ENST00000529278 | chr11 | 102466843 | 5'Flank | SBS | G | C |
| INDIA 8572 | ABLIM3 | chr5 | 149199710 | Intron | SBS | C | G |
| INDIA 8572 | Unknown | chr6 | 78022445 | IGR | SBS | C | G |
| INDIA 8572 | SLC14A2-AS1 | chr18 | 45427036 | Intron | SBS | G | C |
| INDIA 8572 | Unknown | chr1 | 105245286 | IGR | SBS | C | G |
| INDIA 8572 | PRKCE | chr2 | 45861101 | Intron | SBS | C | G |
| INDIA 8572 | DDX47 | chr12 | 12821352 | Nonsense_Mutation | SBS | C | G |
| INDIA 8572 | ENST00000702769 | chr12 | 88414602 | Intron | SBS | C | G |
| INDIA 8572 | NUP50 | chr22 | 45176416 | Intron | SBS | G | C |
| INDIA 8572 | LINC01208 | chr3 | 176523519 | Intron | SBS | G | C |
| INDIA 8572 | GALNTL6 | chr4 | 172993570 | Intron | SBS | G | C |
| INDIA 8572 | IKZF4 | chr12 | 56022133 | Intron | SBS | G | C |
| INDIA 8572 | DAP | chr5 | 10730043 | Intron | SBS | T | G |
| INDIA 8572 | LINC01396 | chr4 | 4844149 | 5'Flank | SBS | C | G |
| INDIA 8572 | AGBL4 | chr1 | 49243940 | Intron | SBS | G | C |
| INDIA 8572 | KCNK12 | chr2 | 47566201 | Intron | SBS | C | G |
| INDIA 8572 | Unknown | chr7 | 50181225 | IGR | SBS | C | G |
| INDIA 8572 | CABCOCO1 | chr10 | 61698519 | Intron | SBS | G | C |
| INDIA 8572 | FHOD3 | chr18 | 36699369 | Intron | SBS | C | G |
| INDIA 8572 | RASSF5 | chr1 | 206538093 | Intron | SBS | C | G |
| INDIA 8572 | TCF7L1 | chr2 | 85183548 | Intron | SBS | G | C |
| INDIA 8572 | LSAMP | chr3 | 116138616 | Intron | SBS | G | T |
| INDIA 8572 | Unknown | chr5 | 35277007 | IGR | SBS | C | G |
| INDIA 8572 | Unknown | chr5 | 134593605 | IGR | SBS | A | C |
| INDIA 8572 | PCDHGA4 | chr5 | 141356215 | Missense_Mutation | SBS | G | C |
| INDIA 8572 | CTNNAL1 | chr9 | 108962997 | Intron | SBS | G | C |
| INDIA 8572 | LINC02664 | chr10 | 31243786 | Intron | SBS | G | C |
| INDIA 8572 | LINC02306 | chr14 | 25819210 | Intron | SBS | G | C |
| INDIA 8572 | Unknown | chr21 | 22615363 | IGR | SBS | G | C |
| INDIA 8572 | Unknown | chr8 | 33217866 | IGR | SBS | G | C |
| INDIA 8572 | ACSM3 | chr16 | 20790630 | Missense_Mutation | SBS | G | C |
| INDIA 8572 | PSMA4 | chr15 | 78552829 | 3'Flank | SBS | G | C |
| INDIA 8572 | CASZ1 | chr1 | 10723744 | Intron | SBS | C | G |
| INDIA 8572 | ZNF219 | chr14 | 21098836 | 5'Flank | SBS | C | G |
| INDIA 8572 | LINC02074 | chr17 | 74044723 | Intron | SBS | G | C |
| INDIA 8572 | ENST00000671547 | chr13 | 110139935 | 5'Flank | SBS | G | T |
| INDIA 8572 | HIVEP3 | chr1 | 41828207 | Intron | SBS | C | G |
| INDIA 8572 | ENST00000652067 | chr1 | 222251708 | Intron | SBS | G | C |
| INDIA 8572 | ENST00000690551 | chr17 | 55619983 | Intron | SBS | G | C |
| INDIA 8572 | NRG2 | chr5 | 140026383 | Intron | SBS | G | C |
| INDIA 8573 | ME3 | chr11 | 86677265 | 5'Flank | SBS | C | G |
| INDIA 8573 | ENST00000670132 | chr8 | 119373342 | Intron | SBS | C | G |

| | | | | | | | |
|---|---|---|---|---|---|---|---|
| INDIA 8573 | Unknown | chr11 | 34720797 | IGR | SBS | G | C |
| INDIA 8573 | Unknown | chr8 | 139157723 | IGR | DEL | TGCAA | - |
| INDIA 8573 | Unknown | chr6 | 103925306 | IGR | SBS | C | G |
| INDIA 8573 | LINC02698 | chr11 | 115826745 | Intron | SBS | C | G |
| INDIA 8573 | ENST00000505955 | chr5 | 73392882 | Intron | SBS | T | A |
| INDIA 8573 | AFF2 | chrX | 148851641 | Intron | SBS | G | T |
| INDIA 8573 | Unknown | chr5 | 3927990 | IGR | SBS | G | C |
| INDIA 8573 | TRAPPC9 | chr8 | 139818274 | Intron | DEL | GAG | - |
| INDIA 8573 | DOCK2 | chr5 | 169853435 | Intron | SBS | C | G |
| INDIA 8573 | KIF6 | chr6 | 39426310 | Intron | SBS | C | G |
| INDIA 8573 | Unknown | chrX | 84243880 | IGR | SBS | T | A |
| INDIA 8573 | CSMD1 | chr8 | 3506295 | Intron | SBS | G | C |
| INDIA 8573 | PLCE1 | chr10 | 94115837 | Intron | DEL | AGC | - |
| INDIA 8573 | CEBPZ | chr2 | 37228128 | Frame_Shift_Del | DEL | A | - |
| INDIA 8573 | ENST00000621575 | chr13 | 33843831 | 5'Flank | DEL | T | - |
| INDIA 8573 | CADPS | chr3 | 62777913 | Intron | SBS | C | G |
| INDIA 8573 | GRM1 | chr6 | 146038248 | Intron | SBS | C | G |
| INDIA 8573 | ABL2 | chr1 | 179157687 | Intron | SBS | C | G |
| INDIA 8573 | RPS6KB2 | chr11 | 67432703 | Intron | SBS | C | G |
| INDIA 8573 | ENST00000559783 | chr15 | 61378560 | Intron | DEL | TG | - |
| INDIA 8573 | Unknown | chr3 | 36373272 | IGR | SBS | G | C |
| INDIA 8573 | MAGI2 | chr7 | 79028508 | Intron | SBS | G | T |
| INDIA 8573 | GTF2IRD1 | chr7 | 74519734 | Intron | DEL | G | - |
| INDIA 8573 | ABCA9 | chr17 | 69020462 | Missense_Mutation | SBS | C | G |
| INDIA 8573 | SPOCK1 | chr5 | 137569427 | Intron | SBS | C | G |
| INDIA 8573 | SOX5 | chr12 | 24081106 | Intron | SBS | C | G |
| INDIA 8573 | TENT4A | chr5 | 6755283 | 3'UTR | DEL | T | - |
| INDIA 8573 | NRG3 | chr10 | 82460832 | Intron | DEL | TCC | - |
| INDIA 8573 | HOXC11 | chr12 | 53972109 | 5'Flank | SBS | C | G |
| INDIA 8573 | Unknown | chr3 | 147682081 | IGR | SBS | C | G |
| INDIA 8573 | TXK | chr4 | 48129608 | Intron | SBS | G | C |
| INDIA 8573 | DLG2 | chr11 | 83540876 | Intron | SBS | T | A |
| INDIA 8573 | LAMA2 | chr6 | 129279904 | Intron | SBS | C | G |
| INDIA 8573 | Unknown | chr20 | 22860795 | IGR | DEL | TCTGATT | - |
| INDIA 8573 | ALKAL2 | chr2 | 275818 | 3'Flank | DEL | TC | - |
| INDIA 8573 | Unknown | chr8 | 26125599 | IGR | SBS | G | T |
| INDIA 8573 | Unknown | chr10 | 61223154 | IGR | SBS | C | G |
| INDIA 8573 | CD5 | chr11 | 61126565 | 3'UTR | SBS | C | G |
| INDIA 8573 | CACNG2 | chr22 | 36663609 | Intron | SBS | C | G |
| INDIA 8573 | CELA2A | chr1 | 15467710 | Intron | INS | - | A |
| INDIA 8573 | DPH6 | chr15 | 35268310 | Intron | DEL | TTGAAGC | - |
| INDIA 8573 | LINC02227 | chr5 | 158321686 | RNA | SBS | T | A |
| INDIA 8573 | Unknown | chr1 | 39980601 | IGR | DEL | G | - |
| INDIA 8573 | Unknown | chr3 | 41066692 | IGR | SBS | C | G |
| INDIA 8573 | PREX2 | chr8 | 68031828 | Intron | SBS | T | G |
| INDIA 8573 | MEGF8 | chr19 | 42353222 | Intron | DEL | C | - |
| INDIA 8573 | ATRX | chrX | 77547281 | Intron | SBS | C | A |
| INDIA 8573 | MEIS1 | chr2 | 66476054 | Intron | SBS | T | G |
| INDIA 8573 | SLIT2 | chr4 | 20367780 | Intron | DEL | T | - |
| INDIA 8573 | LINC01455 | chr5 | 79962030 | Intron | SBS | C | G |
| INDIA 8573 | FOXN3 | chr14 | 89456304 | Intron | SBS | G | C |
| INDIA 8573 | FXN | chr9 | 69046958 | Intron | INS | - | A |
| INDIA 8573 | ERBB2 | chr17 | 39712185 | Intron | SBS | C | G |
| INDIA 8573 | ENST00000546558 | chr12 | 30322785 | 3'Flank | SBS | G | C |
| INDIA 8573 | RASA3 | chr13 | 114078089 | Intron | SBS | C | G |
| INDIA 8573 | ENST00000652126 | chr14 | 39878097 | Intron | SBS | C | G |
| INDIA 8573 | MTUS2 | chr13 | 28856292 | Intron | SBS | G | C |
| INDIA 8573 | Unknown | chr5 | 123644638 | IGR | SBS | G | C |
| INDIA 8573 | LGR4 | chr11 | 27401860 | Intron | DEL | GAGGG | - |
| INDIA 8573 | Unknown | chr3 | 185986049 | IGR | SBS | C | G |
| INDIA 8573 | Unknown | chr5 | 133803532 | IGR | SBS | G | C |
| INDIA 8573 | MGP | chr12 | 14878759 | 3'Flank | INS | - | C |
| INDIA 8573 | ENST00000653849 | chrX | 129234255 | Intron | SBS | T | A |
| INDIA 8573 | ENST00000634305 | chr14 | 36439156 | Intron | SBS | C | G |
| INDIA 8573 | Unknown | chr13 | 27799850 | IGR | SBS | G | C |
| INDIA 8573 | ENST00000702609 | chr3 | 16125601 | Intron | SBS | C | G |

| | | | | | | | |
|---|---|---|---|---|---|---|---|
| INDIA 8573 | CACNA2D4 | chr12 | 1918826 | 5'Flank | SBS | C | A |
| INDIA 8573 | RERGL | chr12 | 18226399 | Intron | SBS | G | C |
| INDIA 8573 | DRAIC | chr15 | 69797402 | Intron | DEL | A | - |
| INDIA 8573 | BCR | chr22 | 23255669 | Intron | DEL | C | - |
| INDIA 8573 | Unknown | chr2 | 147644734 | IGR | SBS | G | C |
| INDIA 8573 | ENST00000640877 | chr12 | 4949646 | Intron | DEL | GGATGTT | - |
| INDIA 8573 | MIR4527HG | chr18 | 47587784 | Intron | DEL | A | - |
| INDIA 8573 | CFAP20 | chr16 | 58110304 | 3'Flank | INS | - | T |
| INDIA 8573 | Unknown | chr2 | 226065400 | IGR | DEL | AATGC | - |
| INDIA 8573 | UNC13C | chr15 | 54604750 | Intron | SBS | G | C |
| INDIA 8573 | LINC00426 | chr13 | 30364016 | Intron | SBS | G | C |
| INDIA 8573 | Unknown | chr5 | 179356460 | IGR | SBS | G | C |
| INDIA 8573 | Unknown | chr7 | 68474698 | IGR | SBS | T | G |
| INDIA 8573 | STIM1 | chr11 | 3942812 | Intron | SBS | C | G |
| INDIA 8573 | GRIN2B | chr12 | 13616311 | Intron | SBS | C | G |
| INDIA 8573 | DRAIC | chr15 | 69730384 | Intron | SBS | C | A |
| INDIA 8573 | NRXN3 | chr14 | 79626344 | Intron | SBS | G | T |
| INDIA 8573 | HELZ | chr17 | 67109344 | Missense_Mutation | SBS | G | C |
| INDIA 8573 | Unknown | chr6 | 381107 | IGR | SBS | G | C |
| INDIA 8573 | MKRN2 | chr3 | 12582895 | 3'UTR | SBS | G | C |
| INDIA 8573 | MKRN2 | chr3 | 12582363 | 3'UTR | SBS | G | C |
| INDIA 8574 | TRDN | chr6 | 123423437 | Intron | SBS | T | A |
| INDIA 8574 | HECTD4 | chr12 | 112197295 | Intron | SBS | C | G |
| INDIA 8574 | ENST00000512859 | chr5 | 81851925 | RNA | SBS | G | C |
| INDIA 8574 | DCC | chr18 | 52749930 | Intron | SBS | A | T |
| INDIA 8574 | CCDC26 | chr8 | 129134173 | Intron | SBS | G | T |
| INDIA 8574 | PLAC8L1 | chr5 | 146079900 | 3'Flank | SBS | A | T |
| INDIA 8574 | RBFOX1 | chr16 | 7565086 | Intron | SBS | C | A |
| INDIA 8574 | Unknown | chr2 | 209112952 | IGR | SBS | C | A |
| INDIA 8574 | Unknown | chr5 | 25060305 | IGR | SBS | T | A |
| INDIA 8574 | CHST11 | chr12 | 104635741 | Intron | DEL | G | - |
| INDIA 8574 | TENM3 | chr4 | 182309716 | Intron | SBS | C | G |
| INDIA 8574 | Unknown | chr5 | 166598176 | IGR | SBS | C | A |
| INDIA 8574 | LINC01060 | chr4 | 188657461 | Intron | SBS | C | A |
| INDIA 8574 | ENST00000657278 | chr13 | 106046851 | Intron | SBS | A | T |
| INDIA 8574 | SLC7A14 | chr3 | 170589866 | 5'Flank | SBS | A | T |
| INDIA 8574 | FBXW4 | chr10 | 101688005 | Intron | SBS | C | A |
| INDIA 8574 | NCOA2 | chr8 | 70162028 | Intron | DEL | AG | - |
| INDIA 8574 | Unknown | chr11 | 104771914 | IGR | DEL | T | - |
| INDIA 8574 | ROBO1 | chr3 | 79004018 | Intron | SBS | C | G |
| INDIA 8574 | ENST00000518570 | chr6 | 119140893 | Intron | SBS | A | T |
| INDIA 8574 | ELMO1 | chr7 | 37333776 | Intron | SBS | C | G |
| INDIA 8574 | NRG3 | chr10 | 82482957 | Intron | SBS | C | G |
| INDIA 8574 | Unknown | chr14 | 87041312 | IGR | SBS | G | C |
| INDIA 8574 | Unknown | chr16 | 35182740 | IGR | SBS | C | A |
| INDIA 8574 | Unknown | chr18 | 38191668 | IGR | SBS | G | C |
| INDIA 8574 | DGKI | chr7 | 137569082 | Intron | SBS | C | A |
| INDIA 8574 | ENST00000511735 | chr4 | 17052274 | Intron | SBS | C | G |
| INDIA 8574 | Unknown | chr3 | 99025995 | IGR | SBS | C | G |
| INDIA 8574 | ENST00000642132 | chr1 | 50255484 | Intron | SBS | T | A |
| INDIA 8574 | NLGN1 | chr3 | 173413017 | Intron | SBS | T | A |
| INDIA 8574 | ENST00000457975 | chr10 | 55343383 | Intron | SBS | G | C |
| INDIA 8574 | ARHGEF3 | chr3 | 57019554 | Intron | SBS | T | A |
| INDIA 8574 | RAP1GAP2 | chr17 | 2882732 | Intron | SBS | G | T |
| INDIA 8574 | PROM1 | chr4 | 16006772 | Intron | SBS | C | A |
| INDIA 8574 | Unknown | chr7 | 81801801 | IGR | SBS | C | G |
| INDIA 8574 | Unknown | chr9 | 24380232 | IGR | SBS | G | T |
| INDIA 8574 | LRMDA | chr10 | 75942343 | Intron | SBS | T | A |
| INDIA 8574 | MYADM-AS1 | chr19 | 53860234 | Intron | SBS | C | G |
| INDIA 8574 | LRRC3B | chr3 | 26662787 | Intron | SBS | T | A |
| INDIA 8574 | DHRS3 | chr1 | 12598105 | Intron | INS | - | CC |
| INDIA 8574 | KCNU1 | chr8 | 36922998 | Intron | SBS | G | C |
| INDIA 8574 | Unknown | chr5 | 163831462 | IGR | SBS | A | T |
| INDIA 8574 | Unknown | chr2 | 116773845 | IGR | SBS | G | C |
| INDIA 8574 | ARL4C | chr2 | 234495325 | 3'UTR | SBS | C | A |
| INDIA 8574 | Unknown | chr8 | 48859891 | IGR | SBS | T | A |

| | | | | | | | |
|---|---|---|---|---|---|---|---|
| INDIA 8574 | ENST00000426204 | chr9 | 111272825 | RNA | DEL | A | - |
| INDIA 8574 | MEGF8 | chr19 | 42344087 | Intron | SBS | C | G |
| INDIA 8574 | MCTP1 | chr5 | 94975195 | Intron | SBS | G | C |
| INDIA 8574 | Unknown | chr20 | 60885320 | IGR | SBS | G | C |
| INDIA 8574 | NEGR1 | chr1 | 72275817 | Intron | SBS | G | C |
| INDIA 8574 | Unknown | chr12 | 127835210 | IGR | SBS | G | C |
| INDIA 8574 | TARID | chr6 | 133788118 | Intron | SBS | T | A |
| INDIA 8574 | Unknown | chr9 | 71485715 | IGR | SBS | C | G |
| INDIA 8574 | ENST00000657272 | chr7 | 9681056 | Intron | SBS | C | G |
| INDIA 8574 | Unknown | chr2 | 60125854 | IGR | SBS | G | C |
| INDIA 8574 | LINC00299 | chr2 | 8096694 | Intron | SBS | C | A |
| INDIA 8574 | SCP2D1 | chr20 | 18814720 | 3'Flank | SBS | G | C |
| INDIA 8574 | PDLIM5 | chr4 | 94518927 | Intron | DEL | CT | - |
| INDIA 8574 | Unknown | chr16 | 26951623 | IGR | DEL | C | - |
| INDIA 8574 | ANK3 | chr10 | 60293920 | Intron | SBS | A | C |
| INDIA 8574 | NCAM2 | chr21 | 21078441 | Intron | SBS | G | C |
| INDIA 8574 | Unknown | chr15 | 48384815 | IGR | DEL | A | - |
| INDIA 8574 | SAMD12-AS1 | chr8 | 118783022 | Intron | SBS | C | A |
| INDIA 8574 | SNHG20 | chr17 | 77097141 | Intron | DEL | A | - |
| INDIA 8574 | ENST00000427015 | chr6 | 147900488 | Intron | SBS | C | A |
| INDIA 8574 | NRG3 | chr10 | 82397849 | Intron | SBS | A | T |
| INDIA 8574 | Unknown | chr20 | 59561060 | IGR | DEL | C | - |
| INDIA 8574 | Unknown | chr2 | 14577898 | IGR | SBS | G | T |
| INDIA 8574 | IGLV9-49 | chr22 | 22345867 | 3'Flank | DEL | C | - |
| INDIA 8574 | Unknown | chr12 | 121619874 | IGR | SBS | C | G |
| INDIA 8574 | RTN4 | chr2 | 55049495 | Intron | SBS | G | C |
| INDIA 8574 | TANC1 | chr2 | 159174296 | Intron | SBS | C | G |
| INDIA 8574 | TPD52L1 | chr6 | 125198140 | Intron | SBS | C | G |
| INDIA 8574 | PRR14 | chr16 | 30653267 | Intron | SBS | G | T |
| INDIA 8574 | LINC00334 | chr21 | 45252138 | Intron | SBS | C | G |
| INDIA 8574 | DNAJC17 | chr15 | 40770402 | Intron | SBS | G | C |
| INDIA 8574 | MIR422A | chr15 | 63869184 | 3'Flank | SBS | G | C |
| INDIA 8574 | LY86-AS1 | chr6 | 6416351 | Intron | SBS | G | C |
| INDIA 8574 | PRKAR1B | chr7 | 576541 | Intron | SBS | G | C |
| INDIA 8574 | VAC14 | chr16 | 70700827 | Intron | SBS | C | G |
| INDIA 8574 | BCL2L14 | chr12 | 12099087 | 3'UTR | SBS | G | C |
| INDIA 8574 | Unknown | chr2 | 22893549 | IGR | SBS | T | A |
| INDIA 8574 | EPHB2 | chr1 | 22868207 | Intron | SBS | G | C |
| INDIA 8574 | IGF1R | chr15 | 98778818 | Intron | DEL | ACC | - |
| INDIA 8574 | P4HTM | chr3 | 49002932 | Intron | SBS | G | C |
| INDIA 8574 | Unknown | chr1 | 43135316 | IGR | SBS | C | G |
| INDIA 8574 | MYO1E | chr15 | 59191017 | Intron | SBS | C | G |
| INDIA 8574 | NSUN7 | chr4 | 40812519 | 3'Flank | SBS | G | C |
| INDIA 8574 | Unknown | chr11 | 69940333 | IGR | SBS | A | T |
| INDIA 8574 | ENST00000457815 | chr3 | 194098212 | Intron | SBS | C | G |
| INDIA 8574 | LACTBL1 | chr1 | 22954763 | Intron | SBS | G | C |
| INDIA 8575 | Unknown | chr6 | 125105716 | IGR | SBS | C | G |
| INDIA 8575 | Unknown | chrX | 117710536 | IGR | SBS | C | G |
| INDIA 8575 | RGS22 | chr8 | 100026525 | Intron | SBS | G | C |
| INDIA 8575 | Unknown | chr16 | 64779595 | IGR | SBS | C | G |
| INDIA 8575 | Unknown | chrX | 4254863 | IGR | SBS | C | G |
| INDIA 8575 | BCOR | chrX | 40146990 | Intron | SBS | G | C |
| INDIA 8575 | PUDP | chrX | 7045509 | 3'Flank | SBS | C | G |
| INDIA 8575 | ENST00000422048 | chrX | 27224309 | Intron | DEL | C | - |
| INDIA 8575 | Unknown | chrX | 25270349 | IGR | SBS | C | G |
| INDIA 8575 | PCP4 | chr21 | 39904647 | Intron | SBS | G | C |
| INDIA 8575 | METAP1D | chr2 | 171998078 | 5'Flank | SBS | G | C |
| INDIA 8575 | Unknown | chr11 | 131045112 | IGR | SBS | C | G |
| INDIA 8575 | ENST00000647485 | chr16 | 49291248 | Intron | SBS | G | C |
| INDIA 8575 | CDH11 | chr16 | 64982239 | Missense_Mutation | SBS | G | C |
| INDIA 8575 | Unknown | chr10 | 74754051 | IGR | SBS | G | C |
| INDIA 8575 | LINC01339 | chr5 | 90153330 | RNA | SBS | G | C |
| INDIA 8575 | Unknown | chr20 | 39852911 | IGR | DEL | G | - |
| INDIA 8575 | CPNE5 | chr6 | 36813013 | Intron | SBS | C | G |
| INDIA 8575 | PEX16 | chr11 | 45913869 | Missense_Mutation | SBS | G | C |
| INDIA 8575 | Unknown | chr7 | 19307196 | IGR | SBS | C | G |

| | | | | | | | |
|---|---|---|---|---|---|---|---|
| INDIA 8575 | Unknown | chr22 | 26076846 | IGR | SBS | C | G |
| INDIA 8575 | DEFB1 | chr8 | 6882543 | 5'Flank | SBS | G | C |
| INDIA 8575 | ENST00000652090 | chr10 | 122680359 | 3'Flank | SBS | C | G |
| INDIA 8575 | Unknown | chr13 | 45945068 | IGR | SBS | G | C |
| INDIA 8575 | FAM124A | chr13 | 51271934 | Intron | SBS | G | C |
| INDIA 8575 | MIR100HG | chr11 | 122128268 | Intron | SBS | C | G |
| INDIA 8575 | ENST00000565564 | chr21 | 30094859 | Intron | SBS | A | C |
| INDIA 8575 | TIAM1 | chr21 | 31401437 | Intron | SBS | G | C |
| INDIA 8575 | PRKCE | chr2 | 45671144 | Intron | SBS | C | G |
| INDIA 8575 | Unknown | chr6 | 92945682 | IGR | SBS | G | C |
| INDIA 8575 | TAFA4 | chr3 | 68747703 | Intron | SBS | G | T |
| INDIA 8575 | NCR2 | chr6 | 41331260 | 5'Flank | SBS | G | C |
| INDIA 8575 | NRG1 | chr8 | 32171000 | Intron | SBS | C | G |
| INDIA 8575 | Unknown | chr10 | 29102489 | IGR | SBS | C | G |
| INDIA 8575 | Unknown | chr10 | 117942103 | IGR | SBS | C | G |
| INDIA 8575 | Unknown | chr14 | 97383571 | IGR | SBS | C | G |
| INDIA 8575 | Unknown | chrX | 137494180 | IGR | SBS | C | G |
| INDIA 8575 | ENST00000654062 | chr7 | 13858994 | RNA | SBS | C | G |
| INDIA 8575 | DLGAP1 | chr18 | 3595422 | Intron | DEL | ATGTAA | - |
| INDIA 8575 | Unknown | chr13 | 47588924 | IGR | SBS | G | C |
| INDIA 8575 | COLEC12 | chr18 | 357202 | Intron | SBS | C | G |
| INDIA 8575 | LINC02060 | chr5 | 88415328 | Intron | SBS | G | C |
| INDIA 8575 | HPS4 | chr22 | 26464344 | Missense_Mutation | SBS | G | C |
| INDIA 8575 | ENST00000407524 | chr2 | 239806495 | 3'Flank | SBS | G | T |
| INDIA 8575 | THSD4 | chr15 | 71280920 | Intron | SBS | G | C |
| INDIA 8575 | Unknown | chr21 | 41114047 | IGR | SBS | G | C |
| INDIA 8575 | Unknown | chr6 | 114587257 | IGR | SBS | C | G |
| INDIA 8575 | ENST00000427460 | chr6 | 159351947 | 3'Flank | DEL | T | - |
| INDIA 8575 | TPK1 | chr7 | 144735367 | Intron | SBS | C | G |
| INDIA 8575 | RAPGEF2 | chr4 | 159210876 | Intron | SBS | C | G |
| INDIA 8575 | Unknown | chr2 | 146512216 | IGR | SBS | G | C |
| INDIA 8575 | Unknown | chr5 | 116631660 | IGR | SBS | C | G |
| INDIA 8575 | PODXL | chr7 | 131502666 | 3'UTR | SBS | G | C |
| INDIA 8575 | Unknown | chr8 | 101027779 | IGR | SBS | G | C |
| INDIA 8575 | Unknown | chr17 | 55714544 | IGR | SBS | G | C |
| INDIA 8575 | Unknown | chr10 | 74738428 | IGR | SBS | G | C |
| INDIA 8575 | FOXP1 | chr3 | 71041603 | Intron | SBS | C | G |
| INDIA 8575 | QSOX2 | chr9 | 136221850 | Nonsense_Mutation | SBS | G | C |
| INDIA 8575 | NPTX2 | chr7 | 98631205 | 3'Flank | SBS | C | G |
| INDIA 8575 | Unknown | chr1 | 108099391 | IGR | SBS | C | G |
| INDIA 8575 | PRMT7 | chr16 | 68346133 | Intron | SBS | C | G |
| INDIA 8575 | RN7SKP148 | chr5 | 175629796 | 3'Flank | SBS | G | C |
| INDIA 8575 | Unknown | chr10 | 128702294 | IGR | SBS | G | C |
| INDIA 8575 | SV2C | chr5 | 76169763 | Intron | SBS | G | C |
| INDIA 8575 | ENST00000659636 | chr2 | 239469738 | Intron | SBS | C | G |
| INDIA 8575 | ABCB5 | chr7 | 20732908 | Intron | SBS | C | G |
| INDIA 8575 | MAGI2 | chr7 | 78903503 | Intron | SBS | G | C |
| INDIA 8575 | SOGA1 | chr20 | 36839372 | Missense_Mutation | SBS | G | C |
| INDIA 8575 | RNF144A | chr2 | 6998267 | Intron | SBS | C | G |
| INDIA 8575 | BCR | chr22 | 23251071 | Intron | SBS | G | C |
| INDIA 8575 | ATG10 | chr5 | 82175153 | Intron | SBS | C | G |
| INDIA 8575 | Unknown | chr7 | 158837186 | IGR | SBS | G | C |
| INDIA 8575 | OCA2 | chr15 | 27798898 | Intron | SBS | G | C |
| INDIA 8575 | PHACTR4 | chr1 | 28432706 | Intron | SBS | G | C |
| INDIA 8575 | Unknown | chr13 | 48109849 | IGR | SBS | C | G |
| INDIA 8575 | ENST00000449215 | chr1 | 14259682 | Intron | SBS | G | C |
| INDIA 8575 | ZNF394 | chr7 | 99500240 | 5'UTR | SBS | C | G |
| INDIA 8575 | ENST00000652090 | chr10 | 122680273 | 3'Flank | SBS | C | G |
| INDIA 8575 | LINC01588 | chr14 | 49995408 | Intron | SBS | C | G |
| INDIA 8575 | RUNX1 | chr21 | 35589833 | Intron | SBS | C | G |
| INDIA 8575 | CSMD3 | chr8 | 113368276 | Intron | SBS | A | T |
| INDIA 8575 | LINC01687 | chr21 | 22084610 | Intron | SBS | G | C |
| INDIA 8575 | UBTD2 | chr5 | 172254503 | Intron | SBS | C | G |
| INDIA 8575 | BCKDHB | chr6 | 80204963 | Intron | SBS | G | C |
| INDIA 8575 | Unknown | chr7 | 56169429 | IGR | SBS | G | C |
| INDIA 8575 | RNU6-470P | chr17 | 43409876 | 5'Flank | SBS | C | G |

| | | | | | | | |
|---|---|---|---|---|---|---|---|
| INDIA 8575 | Unknown | chr17 | 76510292 | IGR | SBS | C | G |
| INDIA 8575 | ENST00000673514 | chr4 | 33482447 | Intron | SBS | A | C |
| INDIA 8575 | Unknown | chr5 | 147791080 | IGR | SBS | C | G |
| INDIA 8575 | Unknown | chr2 | 222784945 | IGR | DEL | G | - |
| INDIA 8575 | Unknown | chr5 | 61577502 | IGR | SBS | C | G |
| INDIA 8575 | EYS | chr6 | 64768878 | Intron | SBS | G | C |
| INDIA 8575 | ENST00000655430 | chr16 | 49243284 | Intron | SBS | G | C |
| INDIA 8576 | TENM1 | chrX | 124496787 | Intron | SBS | A | T |
| INDIA 8576 | WWOX | chr16 | 78844005 | Intron | SBS | C | G |
| INDIA 8576 | ENST00000624062 | chr10 | 112831794 | RNA | SBS | C | G |
| INDIA 8576 | Unknown | chr8 | 100470000 | IGR | SBS | C | G |
| INDIA 8576 | Unknown | chr10 | 120327035 | IGR | SBS | C | G |
| INDIA 8576 | VAV3 | chr1 | 107657576 | Intron | SBS | G | C |
| INDIA 8576 | PER1 | chr17 | 8150305 | Missense_Mutation | SBS | C | G |
| INDIA 8576 | Unknown | chr3 | 32381786 | IGR | SBS | G | C |
| INDIA 8576 | GNAT2 | chr1 | 109610689 | Intron | SBS | G | C |
| INDIA 8576 | PCDH9 | chr13 | 66387244 | Intron | SBS | G | C |
| INDIA 8576 | Unknown | chr4 | 64440975 | IGR | DEL | C | - |
| INDIA 8576 | ADAM12 | chr10 | 126109867 | Intron | SBS | C | G |
| INDIA 8576 | C16orf78 | chr16 | 49378483 | Missense_Mutation | SBS | G | C |
| INDIA 8576 | SMIM2-AS1 | chr13 | 44191775 | Intron | SBS | G | C |
| INDIA 8576 | E2F6 | chr2 | 11460040 | Intron | SBS | C | G |
| INDIA 8576 | ALK | chr2 | 29913523 | Intron | SBS | G | C |
| INDIA 8576 | DCP1A | chr3 | 53298783 | Intron | SBS | C | G |
| INDIA 8576 | DOCK4 | chr7 | 111853704 | Intron | SBS | C | G |
| INDIA 8576 | DPP6 | chr7 | 154192589 | Intron | SBS | C | G |
| INDIA 8576 | CCDC3 | chr10 | 12973184 | Intron | SBS | C | G |
| INDIA 8576 | Unknown | chr12 | 66071406 | IGR | SBS | G | T |
| INDIA 8576 | ENST00000592429 | chr19 | 5174774 | 5'Flank | SBS | G | C |
| INDIA 8576 | ENST00000416173 | chr2 | 14910630 | 5'Flank | SBS | G | C |
| INDIA 8576 | Unknown | chr8 | 122995502 | IGR | SBS | A | T |
| INDIA 8576 | ME1 | chr6 | 83212810 | Intron | SBS | C | G |
| INDIA 8576 | LINC02465 | chr4 | 129901650 | Intron | SBS | T | A |
| INDIA 8576 | ENST00000665012 | chr8 | 90321963 | Intron | SBS | C | G |
| INDIA 8576 | NSG2 | chr5 | 174045693 | 5'Flank | SBS | C | G |
| INDIA 8576 | OR52L1 | chr11 | 5990844 | 5'Flank | SBS | G | C |
| INDIA 8576 | KIRREL1 | chr1 | 158019120 | Intron | SBS | C | A |
| INDIA 8576 | ENST00000425137 | chr10 | 28010505 | Intron | SBS | G | C |
| INDIA 8576 | Unknown | chr5 | 39482160 | IGR | INS | - | AGAG |
| INDIA 8576 | BRINP3 | chr1 | 190414757 | Intron | SBS | G | C |
| INDIA 8576 | Unknown | chr21 | 28833335 | IGR | SBS | G | C |
| INDIA 8576 | MERTK | chr2 | 112004538 | Intron | SBS | G | C |
| INDIA 8576 | LINC01194 | chr5 | 12598508 | Intron | SBS | G | C |
| INDIA 8576 | ZMYND11 | chr10 | 191855 | Intron | INS | - | T |
| INDIA 8576 | MAP3K13 | chr3 | 185329700 | Intron | SBS | G | C |
| INDIA 8576 | Unknown | chr5 | 153832987 | IGR | SBS | G | C |
| INDIA 8576 | Unknown | chr9 | 114126905 | IGR | SBS | C | G |
| INDIA 8576 | HNRNPA1L3 | chr16 | 51588786 | Intron | SBS | T | A |
| INDIA 8576 | DNAH9 | chr17 | 11923891 | Missense_Mutation | SBS | G | C |
| INDIA 8576 | CLSTN2 | chr3 | 140243392 | Intron | DEL | A | - |
| INDIA 8576 | ELAVL1 | chr19 | 7965819 | Intron | INS | - | TCCT |
| INDIA 8576 | DTWD2 | chr5 | 118920420 | Intron | SBS | C | G |
| INDIA 8576 | Unknown | chr5 | 121697445 | IGR | SBS | C | G |
| INDIA 8576 | PITPNM2 | chr12 | 123043789 | Intron | SBS | C | G |
| INDIA 8576 | PPIE | chr1 | 39744220 | Intron | SBS | G | C |
| INDIA 8576 | HEXB | chr5 | 74684543 | 5'Flank | SBS | C | G |
| INDIA 8576 | ABCC4 | chr13 | 95111938 | Intron | SBS | C | G |
| INDIA 8576 | INTS2 | chr17 | 61869876 | Missense_Mutation | SBS | G | C |
| INDIA 8576 | ENST00000510095 | chr4 | 11703264 | Intron | SBS | G | C |
| INDIA 8576 | NPNT | chr4 | 105897614 | Intron | SBS | T | A |
| INDIA 8576 | SUGCT | chr7 | 40677291 | Intron | SBS | C | G |
| INDIA 8576 | Unknown | chr8 | 125452138 | IGR | INS | - | CTTA |
| INDIA 8576 | CAMTA1 | chr1 | 7658037 | Intron | SBS | C | G |
| INDIA 8576 | LRRN2 | chr1 | 204663099 | Intron | SBS | C | G |
| INDIA 8576 | OBI1-AS1 | chr13 | 78339960 | Intron | SBS | G | C |
| INDIA 8576 | WFDC6 | chr20 | 45537449 | Intron | SBS | G | C |

| | | | | | | | |
|---|---|---|---|---|---|---|---|
| INDIA 8576 | R3HDM1 | chr2 | 135631957 | Missense_Mutation | SBS | G | C |
| INDIA 8576 | TMEM131L | chr4 | 153621527 | Intron | SBS | C | G |
| INDIA 8576 | MTCL1 | chr18 | 8796258 | Missense_Mutation | SBS | G | C |
| INDIA 8576 | DDX23 | chr12 | 48829674 | 3'Flank | SBS | C | G |
| INDIA 8576 | PPIE | chr1 | 39743225 | Missense_Mutation | SBS | G | C |
| INDIA 8576 | LEPR | chr1 | 65578485 | Intron | SBS | C | G |
| INDIA 8576 | Unknown | chr1 | 5433396 | IGR | SBS | G | C |
| INDIA 8576 | SLC1A3 | chr5 | 36661583 | Intron | SBS | G | C |
| INDIA 8576 | TCF7 | chr5 | 134143655 | Intron | SBS | C | G |
| INDIA 8576 | KIAA2012 | chr2 | 202109428 | Intron | SBS | G | C |
| INDIA 8576 | PLXNA4 | chr7 | 132508558 | Missense_Mutation | SBS | C | G |
| INDIA 8576 | ENST00000493899 | chr10 | 62878758 | Intron | SBS | C | G |
| INDIA 8576 | OR4S1 | chr11 | 48305671 | 5'Flank | SBS | C | G |
| INDIA 8576 | VWA5B1 | chr1 | 20350714 | Intron | SBS | C | G |
| INDIA 8576 | RERE | chr1 | 8793456 | Intron | SBS | C | G |
| INDIA 8576 | ENST00000660954 | chr3 | 99622407 | Intron | SBS | G | C |
| INDIA 8576 | NDUFAF2 | chr5 | 61028980 | Intron | SBS | C | G |
| INDIA 8576 | RN7SKP102 | chr2 | 123869059 | 3'Flank | SBS | C | G |
| INDIA 8576 | ENST00000664654 | chr10 | 5571726 | 5'Flank | SBS | C | G |
| INDIA 8576 | LINC02641 | chr10 | 123459376 | Intron | SBS | C | G |
| INDIA 8576 | TMEM132D | chr12 | 129176834 | Intron | DEL | G | - |
| INDIA 8576 | ADAMTS17 | chr15 | 100203092 | Intron | DEL | G | - |
| INDIA 8576 | CHIT1 | chr1 | 203225697 | Frame_Shift_Del | DEL | AG | - |
| INDIA 8576 | Unknown | chr3 | 7925607 | IGR | SBS | G | C |
| INDIA 8576 | CDH13 | chr16 | 83214192 | Intron | SBS | G | C |
| INDIA 8576 | FERMT1 | chr20 | 6096640 | Intron | SBS | C | G |
| INDIA 8576 | LINC02934 | chr2 | 65985385 | Intron | SBS | C | G |
| INDIA 8576 | HOOK3 | chr8 | 43024266 | 3'UTR | SBS | C | G |
| INDIA 8576 | ENST00000649194 | chr18 | 37730014 | Intron | SBS | C | A |
| INDIA 8576 | Unknown | chr6 | 45710913 | IGR | SBS | C | A |
| INDIA 8576 | Unknown | chr6 | 119716246 | IGR | SBS | T | A |
| INDIA 8577 | PDK3 | chrX | 24546731 | 3'UTR | SBS | G | C |
| INDIA 8577 | ENST00000643349 | chr11 | 2132354 | 3'UTR | SBS | G | C |
| INDIA 8577 | TAF1 | chrX | 71368007 | Intron | SBS | G | C |
| INDIA 8577 | ADAMTS15 | chr11 | 130467408 | Intron | SBS | G | C |
| INDIA 8577 | SH3KBP1 | chrX | 19652007 | Intron | SBS | G | C |
| INDIA 8577 | LINC01791 | chr19 | 31181991 | Intron | SBS | C | G |
| INDIA 8577 | Unknown | chrX | 109930809 | IGR | SBS | G | C |
| INDIA 8577 | ENST00000662653 | chr11 | 119888228 | 5'Flank | SBS | G | C |
| INDIA 8577 | PLEKHB1 | chr11 | 73664139 | 3'Flank | SBS | G | C |
| INDIA 8577 | PPFIA1 | chr11 | 70332905 | Intron | SBS | C | G |
| INDIA 8577 | NRXN2 | chr11 | 64690298 | Intron | SBS | G | C |
| INDIA 8577 | ZNF710 | chr15 | 90073483 | Intron | SBS | G | C |
| INDIA 8577 | Unknown | chr11 | 50070854 | IGR | SBS | G | C |
| INDIA 8577 | P2RY6 | chr11 | 73281960 | Intron | SBS | G | C |
| INDIA 8577 | CNTN5 | chr11 | 100105944 | Intron | SBS | G | C |
| INDIA 8577 | GNAO1 | chr16 | 56242139 | Intron | SBS | G | C |
| INDIA 8577 | PTPRS | chr19 | 5216093 | Intron | SBS | G | C |
| INDIA 8577 | NLGN4X | chrX | 6132807 | Intron | SBS | C | G |
| INDIA 8577 | ENST00000432359 | chrX | 38774806 | Intron | SBS | C | G |
| INDIA 8577 | CMIP | chr16 | 81546323 | Intron | SBS | G | C |
| INDIA 8577 | TUBGCP3 | chr13 | 112523071 | Intron | SBS | G | C |
| INDIA 8577 | ELAVL2 | chr9 | 23724448 | Intron | SBS | C | G |
| INDIA 8577 | ALG8 | chr11 | 78138576 | Intron | SBS | C | G |
| INDIA 8577 | RYR1 | chr19 | 38529576 | Intron | SBS | G | C |
| INDIA 8577 | TRIM32 | chr9 | 116701797 | 3'Flank | SBS | G | C |
| INDIA 8577 | LINC00923 | chr15 | 97750263 | Intron | SBS | G | T |
| INDIA 8577 | LINC01533 | chr19 | 32023164 | 5'Flank | SBS | G | C |
| INDIA 8577 | NHS | chrX | 17723433 | Intron | SBS | G | C |
| INDIA 8577 | Unknown | chr10 | 81339794 | IGR | SBS | C | G |
| INDIA 8577 | ZNF536 | chr19 | 30388052 | Intron | SBS | G | C |
| INDIA 8577 | MCTS1 | chrX | 120603961 | 5'Flank | SBS | C | G |
| INDIA 8577 | GABBR2 | chr9 | 98426374 | Intron | SBS | G | C |
| INDIA 8577 | UPF3B | chrX | 119852608 | Intron | SBS | C | G |
| INDIA 8577 | CDH8 | chr16 | 61836714 | Intron | SBS | C | G |
| INDIA 8577 | ALDH1L1 | chr3 | 126151004 | Intron | SBS | G | C |

| | | | | | | | |
|---|---|---|---|---|---|---|---|
| INDIA 8577 | ITPKC | chr19 | 40718030 | Missense_Mutation | SBS | G | C |
| INDIA 8577 | GPR4 | chr19 | 45584842 | 3'Flank | SBS | C | G |
| INDIA 8577 | Unknown | chr11 | 128341449 | IGR | SBS | C | G |
| INDIA 8577 | SYMPK | chr19 | 45852892 | Intron | SBS | C | G |
| INDIA 8577 | COL27A1 | chr9 | 114214784 | Intron | SBS | G | C |
| INDIA 8577 | GRM4 | chr6 | 34122307 | Intron | SBS | C | G |
| INDIA 8577 | GTF3C1 | chr16 | 27514046 | Intron | SBS | G | C |
| INDIA 8577 | IGFL4 | chr19 | 46042383 | 5'Flank | SBS | C | G |
| INDIA 8577 | KIAA0040 | chr1 | 175159643 | 3'UTR | SBS | G | C |
| INDIA 8577 | BBOX1 | chr11 | 27127376 | Missense_Mutation | SBS | G | C |
| INDIA 8577 | VGLL4 | chr3 | 11643699 | 5'UTR | SBS | C | G |
| INDIA 8577 | MARK2 | chr11 | 63866584 | Intron | SBS | G | C |
| INDIA 8577 | Unknown | chr2 | 145990173 | IGR | SBS | G | C |
| INDIA 8577 | TRMO | chr9 | 97922068 | Intron | SBS | C | G |
| INDIA 8577 | KCTD21-AS1 | chr11 | 78151813 | Intron | SBS | C | G |
| INDIA 8577 | FAM20A | chr17 | 68596620 | Intron | SBS | C | G |
| INDIA 8577 | PPARGC1B | chr5 | 149822252 | Intron | SBS | C | G |
| INDIA 8577 | CHRDL2 | chr11 | 74722038 | Intron | SBS | C | G |
| INDIA 8577 | Unknown | chr18 | 67357014 | IGR | SBS | C | G |
| INDIA 8577 | ENST00000643090 | chr7 | 17023982 | Intron | SBS | G | C |
| INDIA 8577 | SLCO2A1 | chr3 | 133933587 | 3'UTR | SBS | G | C |
| INDIA 8577 | SHANK2 | chr11 | 70505746 | Intron | SBS | C | G |
| INDIA 8577 | SIPA1L3 | chr19 | 37994371 | Intron | SBS | G | C |
| INDIA 8577 | PLEKHG1 | chr6 | 150803606 | Intron | SBS | C | G |
| INDIA 8577 | Unknown | chr10 | 8123495 | IGR | SBS | C | G |
| INDIA 8577 | SLC25A21 | chr14 | 37077558 | Intron | SBS | G | C |
| INDIA 8577 | GRAMD1A | chr19 | 35005949 | Intron | SBS | G | C |
| INDIA 8577 | ERBIN | chr5 | 66044322 | Intron | SBS | C | G |
| INDIA 8577 | ATP2C2 | chr16 | 84416610 | Intron | SBS | G | C |
| INDIA 8577 | ENST00000490856 | chr7 | 106492770 | Intron | SBS | C | G |
| INDIA 8577 | Unknown | chr10 | 71094613 | IGR | SBS | C | G |
| INDIA 8577 | MRPS18A | chr6 | 43681892 | Intron | SBS | G | C |
| INDIA 8577 | Unknown | chr15 | 96968192 | IGR | SBS | C | G |
| INDIA 8577 | ADAMTS17 | chr15 | 100157739 | Intron | SBS | G | C |
| INDIA 8577 | CACNA1E | chr1 | 181449567 | Intron | SBS | C | G |
| INDIA 8577 | LINC01117 | chr2 | 176791714 | Intron | SBS | C | G |
| INDIA 8577 | PARD3B | chr2 | 205563900 | Intron | SBS | C | G |
| INDIA 8577 | ERBB4 | chr2 | 212227397 | Intron | SBS | G | C |
| INDIA 8577 | Unknown | chr7 | 41043405 | IGR | SBS | C | G |
| INDIA 8577 | ENST00000673717 | chr8 | 141658713 | Intron | SBS | C | G |
| INDIA 8577 | SPTLC1 | chr9 | 92037212 | Intron | SBS | G | C |
| INDIA 8577 | Unknown | chr9 | 108620614 | IGR | SBS | G | C |
| INDIA 8577 | Unknown | chr13 | 97818239 | IGR | SBS | G | C |
| INDIA 8577 | CDYL2 | chr16 | 80680427 | Intron | SBS | C | G |
| INDIA 8577 | MAGIX | chrX | 49166130 | Missense_Mutation | SBS | C | G |
| INDIA 8577 | AR | chrX | 67713178 | Intron | SBS | G | C |
| INDIA 8577 | Unknown | chr15 | 68501544 | IGR | SBS | C | G |
| INDIA 8577 | UBE2D3-AS1 | chr4 | 102833927 | Intron | SBS | C | G |
| INDIA 8577 | Unknown | chr4 | 152255595 | IGR | SBS | G | C |
| INDIA 8577 | ENST00000417838 | chr6 | 148134385 | Intron | SBS | G | C |
| INDIA 8577 | PAX5 | chr9 | 36886416 | Intron | SBS | C | G |
| INDIA 8577 | ENST00000552639 | chr12 | 38547470 | Intron | SBS | C | G |
| INDIA 8577 | Unknown | chr2 | 70862584 | IGR | SBS | G | C |
| INDIA 8577 | CNTNAP5 | chr2 | 124635539 | Intron | SBS | C | G |
| INDIA 8577 | ENST00000509057 | chr5 | 5037851 | 5'Flank | SBS | G | C |
| INDIA 8577 | SLC12A3 | chr16 | 56892192 | Intron | SBS | C | G |
| INDIA 8577 | YPEL1 | chr22 | 21727175 | Intron | SBS | G | C |
| INDIA 8577 | ENST00000515598 | chr5 | 146305096 | 3'Flank | SBS | G | C |
| INDIA 8577 | Unknown | chr11 | 81736651 | IGR | SBS | C | G |
| INDIA 8577 | Unknown | chr1 | 238120155 | IGR | SBS | C | G |
| INDIA 8577 | NCOR2 | chr12 | 124330060 | Intron | SBS | C | G |
| INDIA 8578 | C13orf42 | chr13 | 51152904 | Intron | SBS | A | T |
| INDIA 8578 | RAD51B | chr14 | 68478528 | 3'Flank | SBS | C | G |
| INDIA 8578 | ENST00000666115 | chr13 | 59225146 | Intron | SBS | A | T |
| INDIA 8578 | KCNU1 | chr8 | 36909523 | Silent | SBS | A | T |
| INDIA 8578 | COG2 | chr1 | 230644461 | Intron | SBS | A | T |

| | | | | | | | |
|---|---|---|---|---|---|---|---|
| INDIA 8578 | Unknown | chr13 | 97627103 | IGR | SBS | C | G |
| INDIA 8578 | Unknown | chr19 | 56974961 | IGR | SBS | C | A |
| INDIA 8578 | Unknown | chr3 | 74201044 | IGR | SBS | G | C |
| INDIA 8578 | ARRDC3 | chr5 | 91384140 | 5'Flank | SBS | C | G |
| INDIA 8578 | KCNMA1 | chr10 | 77288141 | Intron | SBS | T | G |
| INDIA 8578 | DCLK1 | chr13 | 35943672 | Intron | SBS | G | C |
| INDIA 8578 | ENST00000640003 | chr9 | 23176187 | Intron | SBS | T | A |
| INDIA 8578 | Unknown | chr1 | 182781742 | IGR | SBS | T | A |
| INDIA 8578 | Unknown | chr3 | 176932241 | IGR | SBS | C | A |
| INDIA 8578 | KIAA0895 | chr7 | 36366702 | Intron | SBS | A | C |
| INDIA 8578 | LINC00907 | chr18 | 42533912 | Intron | SBS | G | C |
| INDIA 8578 | ZCCHC3 | chr20 | 301393 | 3'Flank | SBS | G | T |
| INDIA 8578 | Unknown | chr5 | 6554076 | IGR | SBS | C | G |
| INDIA 8578 | CADPS | chr3 | 62755410 | Intron | SBS | G | C |
| INDIA 8578 | DLG2 | chr11 | 84930343 | Intron | SBS | G | C |
| INDIA 8578 | Unknown | chr12 | 93247459 | IGR | SBS | C | G |
| INDIA 8578 | SLCO3A1 | chr15 | 91984127 | Intron | DEL | GGTT | - |
| INDIA 8578 | SLC39A11 | chr17 | 72657950 | Intron | SBS | G | C |
| INDIA 8578 | GUCA2A | chr1 | 42162168 | 3'Flank | SBS | G | C |
| INDIA 8578 | Unknown | chr21 | 17488151 | IGR | SBS | G | T |
| INDIA 8578 | CCDC93 | chr2 | 117961644 | Intron | SBS | A | T |
| INDIA 8578 | NRP1 | chr10 | 33216943 | Intron | SBS | C | G |
| INDIA 8578 | CCNY | chr10 | 35547117 | Intron | SBS | C | G |
| INDIA 8578 | SLC30A7 | chr1 | 100894929 | 5'Flank | SBS | C | G |
| INDIA 8578 | KCNT1 | chr9 | 135712229 | Intron | SBS | G | C |
| INDIA 8578 | XKR4 | chr8 | 55422797 | Intron | SBS | G | C |
| INDIA 8578 | KIF13B | chr8 | 29100813 | Intron | SBS | G | C |
| INDIA 8578 | FAM13A | chr4 | 88851023 | Missense_Mutation | SBS | G | C |
| INDIA 8578 | LINC01814 | chr2 | 8458944 | 3'Flank | DEL | TTTCAAA | - |
| INDIA 8578 | Unknown | chrX | 54715948 | IGR | SBS | G | C |
| INDIA 8578 | FBLN7 | chr2 | 112174616 | Intron | SBS | G | C |
| INDIA 8578 | XIRP2 | chr2 | 167201624 | Intron | SBS | T | A |
| INDIA 8578 | STXBP5L | chr3 | 120933575 | Intron | SBS | G | C |
| INDIA 8578 | Unknown | chrX | 79099677 | IGR | SBS | C | G |
| INDIA 8578 | ENST00000431589 | chr20 | 42034996 | Intron | SBS | G | C |
| INDIA 8578 | Unknown | chrX | 36669833 | IGR | SBS | G | C |
| INDIA 8578 | C4orf19 | chr4 | 37464002 | Intron | INS | - | CT |
| INDIA 8578 | ENST00000523584 | chr5 | 31101357 | Intron | SBS | G | C |
| INDIA 8578 | SLC26A3 | chr7 | 107801420 | Intron | SBS | G | C |
| INDIA 8578 | CFAP58 | chr10 | 104453567 | Intron | SBS | C | G |
| INDIA 8578 | DDX56 | chr7 | 44574230 | 5'Flank | SBS | G | C |
| INDIA 8578 | ENST00000687248 | chrX | 22392602 | Intron | SBS | G | C |
| INDIA 8578 | CARMIL3 | chr14 | 24048117 | 5'Flank | SBS | C | G |
| INDIA 8578 | Unknown | chr3 | 106436463 | IGR | INS | - | T |
| INDIA 8578 | MAP3K20 | chr2 | 173193804 | Intron | SBS | T | A |
| INDIA 8578 | HOXA13 | chr7 | 27199399 | Missense_Mutation | SBS | C | G |
| INDIA 8578 | ENST00000681682 | chr4 | 111058865 | Intron | SBS | C | G |
| INDIA 8578 | PTPRD | chr9 | 9760065 | Intron | SBS | A | C |
| INDIA 8578 | Unknown | chr13 | 32796370 | IGR | SBS | C | G |
| INDIA 8578 | CACNA1D | chr3 | 53747396 | Frame_Shift_Del | DEL | CT | - |
| INDIA 8578 | Unknown | chr5 | 79876461 | IGR | SBS | T | A |
| INDIA 8578 | MYH14 | chr19 | 50210188 | Intron | SBS | G | C |
| INDIA 8578 | MEIS2 | chr15 | 36989614 | Intron | SBS | C | G |
| INDIA 8578 | Unknown | chr2 | 17161879 | IGR | SBS | G | C |
| INDIA 8578 | Unknown | chr14 | 82869331 | IGR | SBS | A | T |
| INDIA 8578 | Unknown | chr3 | 180060798 | IGR | SBS | C | G |
| INDIA 8578 | SAXO1 | chr9 | 18948807 | Intron | SBS | G | C |
| INDIA 8578 | Unknown | chr20 | 61200419 | IGR | SBS | G | C |
| INDIA 8578 | VPS41 | chr7 | 38725532 | 3'UTR | SBS | C | G |
| INDIA 8578 | Unknown | chr7 | 24461928 | IGR | SBS | G | C |
| INDIA 8578 | ENST00000681682 | chr4 | 110958910 | Intron | SBS | C | A |
| INDIA 8578 | Unknown | chrX | 79093147 | IGR | SBS | A | C |
| INDIA 8578 | FNDC3A | chr13 | 49033282 | Intron | SBS | G | C |
| INDIA 8578 | Unknown | chr6 | 82802963 | IGR | SBS | G | C |
| INDIA 8578 | Unknown | chr16 | 64710149 | IGR | SBS | C | G |
| INDIA 8578 | LINC00645 | chr14 | 27640145 | 3'Flank | SBS | G | C |

| | | | | | | | |
|---|---|---|---|---|---|---|---|
| INDIA 8578 | ENST00000656455 | chr8 | 36752890 | Intron | SBS | C | G |
| INDIA 8578 | Unknown | chr18 | 56359661 | IGR | SBS | G | C |
| INDIA 8578 | Unknown | chr2 | 140201496 | IGR | SBS | A | C |
| INDIA 8578 | Unknown | chr7 | 68065581 | IGR | SBS | C | G |
| INDIA 8578 | LINC01645 | chr1 | 177361379 | Intron | SBS | C | G |
| INDIA 8578 | Unknown | chr16 | 1027508 | IGR | INS | - | T |
| INDIA 8578 | ENST00000657889 | chr7 | 54181742 | 3'Flank | SBS | G | C |
| INDIA 8578 | TSC22D3 | chrX | 107722471 | Intron | SBS | G | C |
| INDIA 8578 | BPTF | chr17 | 67876329 | Intron | SBS | G | C |
| INDIA 8578 | ESRRB | chr14 | 76409846 | Intron | SBS | T | A |
| INDIA 8578 | TNIK | chr3 | 171253737 | Intron | SBS | C | G |
| INDIA 8578 | KLHL6 | chr3 | 183505301 | Intron | SBS | G | C |
| INDIA 8578 | NTRK3 | chr15 | 87981541 | Intron | DEL | AG | - |
| INDIA 8578 | GLIS1 | chr1 | 53664266 | Intron | SBS | G | C |
| INDIA 8578 | JAM3 | chr11 | 134065608 | 5'Flank | SBS | G | C |
| INDIA 8578 | COL22A1 | chr8 | 138744866 | Intron | SBS | G | C |
| INDIA 8578 | Unknown | chr14 | 55854820 | IGR | SBS | C | G |
| INDIA 8578 | SCAMP2 | chr15 | 74873377 | 5'Flank | SBS | G | C |
| INDIA 8578 | ENST00000640770 | chr2 | 497084 | 3'Flank | SBS | G | C |
| INDIA 8578 | SNED1 | chr2 | 241049277 | Intron | SBS | C | G |
| INDIA 8578 | PDGFC | chr4 | 156860079 | Intron | SBS | G | C |
| INDIA 8578 | ENST00000419326 | chr7 | 36766757 | 3'Flank | SBS | C | G |
| INDIA 8578 | SHKBP1 | chr19 | 40595575 | 3'Flank | SBS | C | G |
| INDIA 8578 | PDE11A | chr2 | 177762607 | Intron | SBS | C | G |
| INDIA 8578 | DAAM1 | chr14 | 59274902 | Intron | SBS | T | G |
| INDIA 8579 | Unknown | chrX | 49864514 | IGR | SBS | C | G |
| INDIA 8579 | RASEF | chr9 | 83016259 | Intron | SBS | C | G |
| INDIA 8579 | BRINP1 | chr9 | 119320083 | Intron | SBS | C | G |
| INDIA 8579 | ADAMTSL1 | chr9 | 18250877 | Intron | SBS | C | G |
| INDIA 8579 | PCDH1 | chr5 | 141868378 | Intron | SBS | G | C |
| INDIA 8579 | CTNNA3 | chr10 | 66768363 | Intron | SBS | G | C |
| INDIA 8579 | Unknown | chrX | 20602954 | IGR | SBS | C | G |
| INDIA 8579 | Unknown | chr1 | 221344393 | IGR | SBS | A | C |
| INDIA 8579 | ARMC3 | chr10 | 22926644 | 5'Flank | SBS | C | G |
| INDIA 8579 | ITGA9 | chr3 | 37752078 | Intron | SBS | G | C |
| INDIA 8579 | KPNA7 | chr7 | 99193204 | Intron | SBS | C | G |
| INDIA 8579 | ENST00000563811 | chr16 | 65610195 | Intron | SBS | C | G |
| INDIA 8579 | DNMBP | chr10 | 99873346 | 3'Flank | SBS | A | C |
| INDIA 8579 | RGS6 | chr14 | 72393619 | Intron | SBS | G | C |
| INDIA 8579 | PIWIL4-AS1 | chr11 | 94705704 | Intron | SBS | T | G |
| INDIA 8579 | DDAH1 | chr1 | 85346285 | Intron | SBS | C | G |
| INDIA 8579 | Unknown | chr2 | 138301153 | IGR | SBS | C | G |
| INDIA 8579 | SEMA3E | chr7 | 83592934 | Intron | SBS | C | G |
| INDIA 8579 | ENST00000559783 | chr15 | 61478502 | Intron | SBS | G | C |
| INDIA 8579 | PREB | chr2 | 27130998 | 3'UTR | SBS | G | T |
| INDIA 8579 | Unknown | chr12 | 63234596 | IGR | SBS | C | G |
| INDIA 8579 | PICALM | chr11 | 85976254 | Intron | SBS | C | G |
| INDIA 8579 | COG4 | chr16 | 70512362 | Missense_Mutation | SBS | C | G |
| INDIA 8579 | ASTN1 | chr1 | 177090254 | Intron | SBS | G | C |
| INDIA 8579 | PARD3B | chr2 | 205616095 | 3'UTR | SBS | C | A |
| INDIA 8579 | IGFBP5 | chr2 | 216691587 | Intron | SBS | C | G |
| INDIA 8579 | Unknown | chr16 | 66161272 | IGR | DEL | G | - |
| INDIA 8579 | GRM5 | chr11 | 88808955 | Intron | SBS | C | G |
| INDIA 8579 | ZBTB20 | chr3 | 114870066 | Intron | SBS | C | G |
| INDIA 8579 | MAP9 | chr4 | 155376975 | 5'Flank | SBS | T | G |
| INDIA 8579 | MEF2C-AS1 | chr5 | 89426826 | Intron | SBS | T | G |
| INDIA 8579 | CAMK4 | chr5 | 111350565 | Intron | SBS | C | G |
| INDIA 8579 | LINC00326 | chr6 | 132996972 | Intron | SBS | G | C |
| INDIA 8579 | TRAPPC9 | chr8 | 140262090 | Intron | SBS | C | G |
| INDIA 8579 | AFAP1L2 | chr10 | 114359501 | Intron | SBS | G | C |
| INDIA 8579 | LMNTD1 | chr12 | 25622712 | Intron | SBS | G | C |
| INDIA 8579 | ANO4 | chr12 | 101041460 | Intron | SBS | G | C |
| INDIA 8579 | PXDNL | chr8 | 51618004 | Intron | SBS | T | A |
| INDIA 8579 | Unknown | chr13 | 105818241 | IGR | SBS | G | C |
| INDIA 8579 | JPH3 | chr16 | 87692317 | Intron | DEL | CT | - |
| INDIA 8579 | NCAM1 | chr11 | 113037513 | Intron | SBS | G | T |

| | | | | | | | |
|---|---|---|---|---|---|---|---|
| INDIA 8579 | WAC-AS1 | chr10 | 28526760 | Intron | SBS | G | C |
| INDIA 8579 | NAALADL2 | chr3 | 175753935 | Intron | SBS | G | C |
| INDIA 8579 | ENST00000651336 | chr2 | 79291991 | Intron | SBS | C | A |
| INDIA 8579 | METTL4 | chr18 | 2536666 | 3'Flank | SBS | G | C |
| INDIA 8579 | CABLES1 | chr18 | 23166730 | Intron | SBS | G | C |
| INDIA 8579 | IQGAP3 | chr1 | 156524950 | 3'Flank | SBS | G | C |
| INDIA 8579 | Unknown | chr11 | 33844251 | IGR | SBS | C | G |
| INDIA 8579 | Unknown | chr2 | 149553342 | IGR | SBS | C | G |
| INDIA 8579 | ENST00000650982 | chr1 | 236127801 | Intron | SBS | C | G |
| INDIA 8579 | MECOM | chr3 | 169244534 | Intron | SBS | G | C |
| INDIA 8579 | MTMR4 | chr17 | 58504128 | Frame_Shift_Ins | INS | - | ACCT |
| INDIA 8579 | NLRP1 | chr17 | 5558460 | Missense_Mutation | SBS | C | G |
| INDIA 8579 | TMEM260 | chr14 | 56582764 | Intron | SBS | C | G |
| INDIA 8579 | SORL1 | chr11 | 121553530 | Intron | SBS | C | G |
| INDIA 8579 | ABLIM1 | chr10 | 114581781 | Intron | SBS | C | A |
| INDIA 8579 | TMEM266 | chr15 | 76192344 | Intron | SBS | G | C |
| INDIA 8579 | Unknown | chr12 | 106227819 | IGR | SBS | G | T |
| INDIA 8579 | HAUS3 | chr4 | 2240278 | Frame_Shift_Del | DEL | AAAGA | - |
| INDIA 8579 | EMX1 | chr2 | 72915402 | 5'Flank | SBS | C | G |
| INDIA 8579 | Unknown | chr2 | 66213691 | IGR | SBS | C | G |
| INDIA 8579 | SLC9A2 | chr2 | 102618181 | 5'Flank | SBS | C | G |
| INDIA 8579 | Unknown | chr16 | 82248074 | IGR | SBS | C | G |
| INDIA 8579 | ENST00000551940 | chr12 | 115611440 | Intron | SBS | G | C |
| INDIA 8579 | BBX | chr3 | 107682903 | Intron | SBS | G | C |
| INDIA 8579 | Unknown | chr4 | 110656867 | IGR | SBS | G | C |
| INDIA 8579 | ENST00000450677 | chr10 | 59587013 | 5'Flank | SBS | G | C |
| INDIA 8579 | VIPR2 | chr7 | 159127734 | Intron | SBS | C | G |
| INDIA 8579 | HEG1 | chr3 | 124969653 | 3'UTR | SBS | C | G |
| INDIA 8579 | RPRD2 | chr1 | 150381904 | Intron | SBS | C | G |
| INDIA 8579 | C1QL2 | chr2 | 119162296 | 5'Flank | SBS | T | A |
| INDIA 8579 | TBCD | chr17 | 82874972 | Intron | INS | - | A |
| INDIA 8579 | SEL1L2 | chr20 | 13877998 | Intron | SBS | C | G |
| INDIA 8579 | SELENOT | chr3 | 150609249 | Intron | SBS | C | G |
| INDIA 8579 | ADAMTS2 | chr5 | 179261458 | Intron | SBS | C | G |
| INDIA 8579 | DCUN1D4 | chr4 | 51842088 | 5'Flank | DEL | AGGACT | - |
| INDIA 8579 | COG4 | chr16 | 70512438 | Splice_Region | SBS | C | G |
| INDIA 8579 | MTMR14 | chr3 | 9683570 | Intron | SBS | G | C |
| INDIA 8579 | ENST00000592317 | chr17 | 60135380 | 3'Flank | SBS | C | A |
| INDIA 8579 | ADAMDEC1 | chr8 | 24408160 | 3'Flank | SBS | C | G |
| INDIA 8579 | RGL1 | chr1 | 183771028 | Intron | INS | - | G |
| INDIA 8579 | Unknown | chr4 | 171709407 | IGR | SBS | A | C |
| INDIA 8579 | Unknown | chr1 | 26666907 | IGR | SBS | G | C |
| INDIA 8579 | ENST00000653710 | chr2 | 183281238 | Intron | SBS | C | G |
| INDIA 8579 | ENST00000503323 | chr5 | 143672136 | Intron | SBS | G | C |
| INDIA 8579 | BRWD1 | chr21 | 39201478 | Intron | SBS | G | C |
| INDIA 8579 | CERK | chr22 | 46686280 | 3'UTR | SBS | G | C |
| INDIA 8579 | RIN2 | chr20 | 19757268 | 5'Flank | SBS | C | G |
| INDIA 8579 | CD226 | chr18 | 69913821 | Intron | SBS | G | C |
| INDIA 8579 | Unknown | chr17 | 12486442 | IGR | SBS | A | T |
| INDIA 8579 | Unknown | chr7 | 101717888 | IGR | INS | - | A |
| INDIA 8579 | RBP5 | chr12 | 7119672 | 3'Flank | SBS | C | G |
| INDIA 8579 | ENST00000517276 | chr1 | 13695081 | 3'Flank | SBS | G | C |
| INDIA 8579 | GPM6A | chr4 | 175772355 | Intron | SBS | C | A |
| INDIA 8580 | SPAAR | chr9 | 35908348 | 5'Flank | SBS | G | C |
| INDIA 8580 | RECK | chr9 | 36064590 | Intron | SBS | G | C |
| INDIA 8580 | ENST00000664438 | chr9 | 13500978 | Intron | SBS | C | G |
| INDIA 8580 | Unknown | chr9 | 4412078 | IGR | SBS | C | G |
| INDIA 8580 | RPP25L | chr9 | 34607377 | 3'Flank | SBS | G | C |
| INDIA 8580 | KANK1 | chr9 | 741235 | Intron | SBS | C | G |
| INDIA 8580 | LINGO2 | chr9 | 29159236 | Intron | SBS | T | A |
| INDIA 8580 | RFX3-DT | chr9 | 3671609 | RNA | SBS | C | G |
| INDIA 8580 | RRM2 | chr2 | 10125865 | Intron | SBS | G | C |
| INDIA 8580 | IRF2 | chr4 | 184411477 | Intron | SBS | C | G |
| INDIA 8580 | Unknown | chr17 | 13439644 | IGR | SBS | G | C |
| INDIA 8580 | Unknown | chr7 | 35590680 | IGR | SBS | G | C |
| INDIA 8580 | HS6ST3 | chr13 | 96502282 | Intron | SBS | G | C |

| | | | | | | | |
|---|---|---|---|---|---|---|---|
| INDIA 8580 | ASB11 | chrX | 15284420 | Intron | SBS | G | C |
| INDIA 8580 | NDUFB6 | chr9 | 32551016 | 3'Flank | SBS | G | C |
| INDIA 8580 | GET3 | chr19 | 12738426 | Intron | SBS | C | G |
| INDIA 8580 | DHX30 | chr3 | 47817580 | Intron | SBS | G | C |
| INDIA 8580 | ENST00000515213 | chr7 | 377198 | 5'Flank | SBS | G | C |
| INDIA 8580 | CMTM8 | chr3 | 32238242 | 5'Flank | SBS | C | G |
| INDIA 8580 | ENST00000453451 | chr2 | 35080673 | Intron | SBS | G | C |
| INDIA 8580 | COL4A1 | chr13 | 110155183 | Intron | SBS | G | C |
| INDIA 8580 | Unknown | chr2 | 49620609 | IGR | SBS | G | C |
| INDIA 8580 | Unknown | chr6 | 44704487 | IGR | SBS | C | G |
| INDIA 8580 | NHSL2 | chrX | 72026943 | Intron | SBS | G | C |
| INDIA 8580 | XYLT1 | chr16 | 17470101 | Intron | DEL | ACCT | - |
| INDIA 8580 | SPATS1 | chr6 | 44361310 | Intron | SBS | C | G |
| INDIA 8580 | EML1 | chr14 | 99781260 | Intron | SBS | C | G |
| INDIA 8580 | LINC02408 | chr12 | 67458543 | Intron | SBS | C | G |
| INDIA 8580 | ARHGAP10 | chr4 | 147732972 | Intron | SBS | C | G |
| INDIA 8580 | ENST00000561802 | chr16 | 73228831 | 5'Flank | SBS | C | G |
| INDIA 8580 | FANCD2OS | chr3 | 10110664 | 5'Flank | SBS | G | C |
| INDIA 8580 | ARHGAP21 | chr10 | 24647053 | Intron | SBS | C | G |
| INDIA 8580 | GALNT8 | chr12 | 4779943 | Intron | SBS | C | G |
| INDIA 8580 | SCNN1B | chr16 | 23367664 | Intron | SBS | C | G |
| INDIA 8580 | MTUS2 | chr13 | 29117552 | Intron | SBS | G | C |
| INDIA 8580 | ENST00000634305 | chr14 | 36398426 | Intron | SBS | G | C |
| INDIA 8580 | PSME4 | chr2 | 53885022 | Intron | SBS | C | G |
| INDIA 8580 | SATB1-AS1 | chr3 | 18787540 | Intron | SBS | G | C |
| INDIA 8580 | Unknown | chr7 | 41058725 | IGR | SBS | C | G |
| INDIA 8580 | Unknown | chr19 | 9172630 | IGR | SBS | G | C |
| INDIA 8580 | CFAP20 | chr16 | 58120494 | Intron | SBS | C | G |
| INDIA 8580 | CLYBL | chr13 | 99826494 | Intron | SBS | C | G |
| INDIA 8580 | CCM2 | chr7 | 45001244 | Intron | SBS | G | C |
| INDIA 8580 | ZGPAT | chr20 | 63735343 | Silent | SBS | G | C |
| INDIA 8580 | SRGAP3 | chr3 | 9031229 | Intron | SBS | G | C |
| INDIA 8580 | FILIP1 | chr6 | 75487389 | Intron | SBS | G | C |
| INDIA 8580 | RAD51D | chr17 | 35107673 | Intron | SBS | G | C |
| INDIA 8580 | CPNE2 | chr16 | 57097740 | Intron | SBS | C | G |
| INDIA 8580 | Unknown | chr20 | 55270180 | IGR | SBS | G | C |
| INDIA 8580 | ENST00000659453 | chr8 | 21073236 | Intron | SBS | G | C |
| INDIA 8580 | UBR1 | chr15 | 43038085 | Intron | SBS | C | G |
| INDIA 8580 | SHANK2 | chr11 | 70567140 | Intron | SBS | G | C |
| INDIA 8580 | ATP2B2 | chr3 | 10469694 | Intron | SBS | C | G |
| INDIA 8580 | PHACTR1 | chr6 | 13214411 | Intron | SBS | G | C |
| INDIA 8580 | Unknown | chr8 | 34916967 | IGR | SBS | G | C |
| INDIA 8580 | RBFOX1 | chr16 | 6982222 | Intron | SBS | C | G |
| INDIA 8580 | CDYL2 | chr16 | 80769518 | Intron | SBS | G | C |
| INDIA 8580 | GALNT9 | chr12 | 132297445 | Intron | SBS | C | G |
| INDIA 8580 | Unknown | chrX | 109558055 | IGR | SBS | G | C |
| INDIA 8580 | COL12A1 | chr6 | 75176024 | Intron | SBS | G | C |
| INDIA 8580 | TRIM67 | chr1 | 231194342 | Intron | SBS | C | G |
| INDIA 8580 | Unknown | chr5 | 35347179 | IGR | SBS | C | G |
| INDIA 8580 | RUFY3 | chr4 | 70790134 | Intron | SBS | C | G |
| INDIA 8580 | CNTNAP2 | chr7 | 146844698 | Intron | SBS | C | G |
| INDIA 8580 | CACNG2 | chr22 | 36644998 | Intron | SBS | C | G |
| INDIA 8580 | PLA2G12A | chr4 | 109730512 | 5'Flank | SBS | G | C |
| INDIA 8580 | DGKH | chr13 | 42080362 | Intron | SBS | G | C |
| INDIA 8580 | DCDC2 | chr6 | 24179175 | Intron | SBS | G | T |
| INDIA 8580 | DLEU1 | chr13 | 50353171 | Intron | SBS | C | G |
| INDIA 8580 | NRXN1 | chr2 | 50025408 | Intron | SBS | G | C |
| INDIA 8580 | SIAH3 | chr13 | 45847471 | Intron | SBS | C | G |
| INDIA 8580 | APC2 | chr19 | 1454250 | Intron | SBS | G | C |
| INDIA 8580 | CARM1 | chr19 | 10889133 | Intron | SBS | G | C |
| INDIA 8580 | LINC00862 | chr1 | 200337412 | Intron | SBS | G | C |
| INDIA 8580 | SGCD | chr5 | 156613789 | Intron | SBS | C | G |
| INDIA 8580 | HMCN1 | chr1 | 185971941 | Intron | SBS | G | C |
| INDIA 8580 | RNU5B-2P | chr3 | 40500936 | 5'Flank | SBS | C | G |
| INDIA 8580 | LINC02643 | chr10 | 21344377 | Intron | SBS | C | G |
| INDIA 8580 | LRFN2 | chr6 | 40491218 | Intron | SBS | C | G |

| | | | | | | | |
|---|---|---|---|---|---|---|---|
| INDIA 8580 | DGKD | chr2 | 233469048 | Intron | SBS | G | C |
| INDIA 8580 | TRIM13 | chr13 | 49996015 | 5'Flank | SBS | G | C |
| INDIA 8580 | Unknown | chr3 | 44026493 | IGR | SBS | C | G |
| INDIA 8580 | Unknown | chr5 | 10954136 | IGR | SBS | C | G |
| INDIA 8580 | OBI1-AS1 | chr13 | 78381482 | Intron | SBS | G | C |
| INDIA 8580 | FMO2 | chr1 | 171200197 | Intron | SBS | G | C |
| INDIA 8580 | RYR3 | chr15 | 33467926 | Intron | SBS | G | C |
| INDIA 8580 | Unknown | chr16 | 62538333 | IGR | SBS | C | G |
| INDIA 8580 | BACH1 | chr21 | 29410381 | Intron | SBS | C | G |
| INDIA 8580 | FCRLB | chr1 | 161717600 | 5'Flank | SBS | G | C |
| INDIA 8580 | PHACTR3 | chr20 | 59768381 | Intron | SBS | C | G |
| INDIA 8580 | DCHS2 | chr4 | 154370092 | Intron | SBS | C | G |
| INDIA 8580 | Unknown | chr8 | 138994339 | IGR | SBS | G | C |
| INDIA 8580 | GRIN2B | chr12 | 13498802 | Intron | SBS | C | G |
| INDIA 8581 | ENST00000431589 | chr20 | 42046703 | Intron | SBS | C | G |
| INDIA 8581 | Unknown | chr6 | 148611708 | IGR | SBS | G | C |
| INDIA 8581 | HOXC4 | chr12 | 54054549 | Intron | SBS | T | A |
| INDIA 8581 | RYR2 | chr1 | 237155931 | Intron | SBS | C | G |
| INDIA 8581 | TMEM132D | chr12 | 129454564 | Intron | SBS | G | T |
| INDIA 8581 | Unknown | chr14 | 83577428 | IGR | SBS | A | T |
| INDIA 8581 | ADGRG6 | chr6 | 142355676 | Intron | SBS | T | A |
| INDIA 8581 | ANKRD44 | chr2 | 197125965 | Missense_Mutation | SBS | G | C |
| INDIA 8581 | Unknown | chr3 | 45184951 | IGR | SBS | C | G |
| INDIA 8581 | PAPPA2 | chr1 | 176699733 | Intron | SBS | T | G |
| INDIA 8581 | TRHDE | chr12 | 72402941 | Intron | SBS | C | G |
| INDIA 8581 | ENST00000689451 | chr12 | 88777366 | 3'Flank | SBS | G | C |
| INDIA 8581 | ARIH2 | chr3 | 48969345 | Intron | SBS | G | C |
| INDIA 8581 | ROBO2 | chr3 | 76621501 | Intron | SBS | T | A |
| INDIA 8581 | TRAV39 | chr14 | 22300133 | 5'Flank | SBS | C | A |
| INDIA 8581 | CACNG7 | chr19 | 53943474 | 3'UTR | DEL | G | - |
| INDIA 8581 | NEK10 | chr3 | 27311152 | Intron | SBS | C | A |
| INDIA 8581 | ENST00000668367 | chr6 | 93654049 | Intron | SBS | G | C |
| INDIA 8581 | ARNTL | chr11 | 13358923 | Intron | SBS | C | G |
| INDIA 8581 | ENST00000649113 | chr6 | 122979476 | Intron | SBS | C | G |
| INDIA 8581 | Unknown | chr14 | 23600400 | IGR | SBS | C | G |
| INDIA 8581 | Unknown | chr21 | 23176295 | IGR | SBS | T | G |
| INDIA 8581 | KCNK10 | chr14 | 88272062 | Intron | DEL | C | - |
| INDIA 8581 | LARS2 | chr3 | 45474116 | Intron | SBS | G | C |
| INDIA 8581 | ENST00000414598 | chr20 | 52457466 | 5'Flank | DEL | C | - |
| INDIA 8581 | RSPH14 | chr22 | 23129356 | Intron | SBS | C | A |
| INDIA 8581 | TP53 | chr17 | 7670712 | Frame_Shift_Del | DEL | G | - |
| INDIA 8581 | NMI | chr2 | 151270185 | 3'Flank | SBS | C | G |
| INDIA 8581 | ENST00000556016 | chr14 | 85224430 | Intron | SBS | C | G |
| INDIA 8581 | Unknown | chr14 | 85668343 | IGR | SBS | A | T |
| INDIA 8581 | ZEB2 | chr2 | 144413324 | Intron | SBS | A | C |
| INDIA 8581 | GRM8 | chr7 | 126723475 | Intron | SBS | C | A |
| INDIA 8581 | FRMD5 | chr15 | 44125851 | Intron | SBS | T | G |
| INDIA 8581 | ENST00000661928 | chr4 | 148802973 | Intron | SBS | C | A |
| INDIA 8581 | C7orf77 | chr7 | 124774240 | 3'Flank | SBS | G | C |
| INDIA 8581 | Unknown | chr13 | 58152490 | IGR | SBS | C | G |
| INDIA 8581 | Unknown | chr4 | 7151734 | IGR | SBS | G | C |
| INDIA 8581 | ENST00000506305 | chr5 | 4028901 | 3'Flank | SBS | A | T |
| INDIA 8581 | Unknown | chr13 | 68552265 | IGR | SBS | G | C |
| INDIA 8581 | KCNH7 | chr2 | 162796997 | Intron | SBS | G | C |
| INDIA 8581 | Unknown | chr12 | 127855948 | IGR | SBS | C | G |
| INDIA 8581 | Unknown | chr13 | 69585125 | IGR | SBS | T | A |
| INDIA 8581 | Unknown | chr4 | 178741472 | IGR | SBS | G | C |
| INDIA 8581 | CIR1 | chr2 | 174344424 | 3'Flank | DEL | G | - |
| INDIA 8581 | ROBO2 | chr3 | 77298229 | Intron | SBS | C | G |
| INDIA 8581 | SLC30A10 | chr1 | 219926862 | Intron | SBS | C | G |
| INDIA 8581 | FAAH | chr1 | 46398435 | Intron | SBS | A | T |
| INDIA 8581 | PDE10A | chr6 | 165647328 | Intron | SBS | A | T |
| INDIA 8581 | OLFM4 | chr13 | 53050046 | Missense_Mutation | SBS | C | G |
| INDIA 8581 | Unknown | chr13 | 68558398 | IGR | DEL | G | - |
| INDIA 8581 | SP140 | chr2 | 230243918 | Intron | SBS | A | C |
| INDIA 8581 | ZNF804B | chr7 | 89335060 | Missense_Mutation | SBS | A | T |

| | | | | | | | |
|---|---|---|---|---|---|---|---|
| INDIA 8581 | ASNS | chr7 | 97853904 | Intron | SBS | C | G |
| INDIA 8581 | RFC3 | chr13 | 33881561 | Intron | SBS | C | G |
| INDIA 8581 | ARID3A | chr19 | 960042 | Intron | SBS | C | A |
| INDIA 8581 | Unknown | chr22 | 48095902 | IGR | SBS | G | C |
| INDIA 8581 | CLPB | chr11 | 72378119 | Intron | SBS | C | G |
| INDIA 8581 | CAND2 | chr3 | 12831306 | Intron | DEL | G | - |
| INDIA 8581 | DRC7 | chr16 | 57720601 | Intron | SBS | G | C |
| INDIA 8581 | Unknown | chr10 | 109119646 | IGR | SBS | A | T |
| INDIA 8581 | RN7SL363P | chr15 | 90861014 | 5'Flank | SBS | G | C |
| INDIA 8581 | ETFB | chr19 | 51343882 | 3'Flank | SBS | C | G |
| INDIA 8581 | Unknown | chr1 | 219568918 | IGR | DEL | C | - |
| INDIA 8581 | ZMIZ1 | chr10 | 79091804 | Intron | INS | - | G |
| INDIA 8581 | PTPA | chr9 | 129137905 | Intron | SBS | C | G |
| INDIA 8581 | FYB1 | chr5 | 39188262 | Intron | DEL | A | - |
| INDIA 8581 | ENST00000669768 | chr11 | 112821795 | 3'Flank | SBS | A | T |
| INDIA 8581 | NCAM1 | chr11 | 113076084 | Intron | SBS | C | G |
| INDIA 8581 | NCAM1 | chr11 | 113185650 | Intron | SBS | G | C |
| INDIA 8581 | Unknown | chr4 | 180195100 | IGR | SBS | T | G |
| INDIA 8581 | ENST00000671332 | chr12 | 17068499 | Intron | SBS | A | T |
| INDIA 8581 | TENM4 | chr11 | 78925576 | Intron | SBS | G | C |
| INDIA 8581 | PPFIBP2 | chr11 | 7642255 | Intron | INS | - | TG |
| INDIA 8581 | MECOM | chr3 | 169138299 | Intron | SBS | T | A |
| INDIA 8581 | CNTN5 | chr11 | 99849493 | Intron | SBS | C | G |
| INDIA 8581 | NUDT13 | chr10 | 73119365 | Intron | SBS | T | A |
| INDIA 8581 | ENST00000436492 | chr6 | 168717306 | Intron | DEL | C | - |
| INDIA 8581 | Unknown | chr13 | 74222115 | IGR | SBS | G | C |
| INDIA 8581 | Unknown | chr2 | 107065317 | IGR | SBS | C | G |
| INDIA 8581 | PLAC8L1 | chr5 | 146090162 | Intron | SBS | G | C |
| INDIA 8581 | LINC01376 | chr2 | 19132116 | Intron | SBS | C | G |
| INDIA 8581 | CRYBG3 | chr3 | 97825781 | Intron | SBS | T | A |
| INDIA 8581 | ENST00000429636 | chr2 | 164144964 | Intron | DEL | T | - |
| INDIA 8581 | Unknown | chr4 | 156270424 | IGR | DEL | C | - |
| INDIA 8581 | EML6 | chr2 | 54930705 | Intron | SBS | A | T |
| INDIA 8581 | GRM8 | chr7 | 126980096 | Intron | SBS | A | C |
| INDIA 8581 | NCAM1 | chr11 | 113154694 | Intron | SBS | A | T |
| INDIA 8581 | RERE | chr1 | 8396925 | Intron | DEL | AC | - |
| INDIA 8581 | SMIM20 | chr4 | 25905009 | Intron | SBS | G | C |
| INDIA 8581 | Unknown | chr8 | 65409620 | IGR | SBS | G | C |
| INDIA 8581 | GNG12 | chr1 | 67772709 | Intron | SBS | C | G |
| INDIA 8581 | KIAA1217 | chr10 | 23696654 | Intron | SBS | G | C |
| INDIA 8581 | TNR | chr1 | 175417526 | Intron | SBS | G | C |
| INDIA 8581 | CAMTA1 | chr1 | 7615614 | Intron | SBS | C | G |
| INDIA 8581 | LHPP | chr10 | 124516897 | Intron | DEL | C | - |
| INDIA 8582 | WWC3 | chrX | 10103434 | Intron | SBS | G | C |
| INDIA 8582 | ST8SIA5 | chr18 | 46685924 | Intron | SBS | C | G |
| INDIA 8582 | Unknown | chr9 | 108268013 | IGR | SBS | C | G |
| INDIA 8582 | PRSS51 | chr8 | 10496223 | 3'Flank | SBS | A | T |
| INDIA 8582 | PIGL | chr17 | 16326424 | 3'Flank | SBS | G | C |
| INDIA 8582 | LINC01509 | chr9 | 107420165 | 3'Flank | SBS | C | G |
| INDIA 8582 | Unknown | chr9 | 78349226 | IGR | SBS | C | G |
| INDIA 8582 | LINC01505 | chr9 | 106297450 | Intron | SBS | A | C |
| INDIA 8582 | C9orf92 | chr9 | 16339052 | Intron | SBS | G | C |
| INDIA 8582 | Unknown | chr10 | 112015165 | IGR | SBS | G | C |
| INDIA 8582 | HS3ST4 | chr16 | 26117502 | Intron | SBS | G | C |
| INDIA 8582 | NCOA7 | chr6 | 125819840 | Intron | SBS | G | C |
| INDIA 8582 | C4orf50 | chr4 | 5965729 | Intron | SBS | G | C |
| INDIA 8582 | ZNF337 | chr20 | 25670329 | 3'Flank | SBS | C | G |
| INDIA 8582 | TTC7B | chr14 | 90677743 | Intron | SBS | C | G |
| INDIA 8582 | Unknown | chr9 | 81877579 | IGR | SBS | C | G |
| INDIA 8582 | ENST00000590968 | chr18 | 61748130 | RNA | SBS | C | G |
| INDIA 8582 | PEX5L | chr3 | 180010721 | Intron | SBS | C | G |
| INDIA 8582 | ENST00000669430 | chr13 | 94993229 | Intron | SBS | G | C |
| INDIA 8582 | Unknown | chr18 | 28858822 | IGR | SBS | G | T |
| INDIA 8582 | DUSP5 | chr10 | 110507119 | Missense_Mutation | SBS | T | A |
| INDIA 8582 | MYO16 | chr13 | 109163349 | Intron | SBS | G | C |
| INDIA 8582 | ENST00000558587 | chr15 | 86942061 | Intron | SBS | A | T |

| | | | | | | | |
|---|---|---|---|---|---|---|---|
| INDIA 8582 | MECOM | chr3 | 169324272 | Intron | SBS | C | G |
| INDIA 8582 | LARGE1 | chr22 | 33613214 | Intron | SBS | G | C |
| INDIA 8582 | Unknown | chr5 | 166165830 | IGR | SBS | C | A |
| INDIA 8582 | RAB27B | chr18 | 54836179 | Intron | SBS | C | G |
| INDIA 8582 | USP6 | chr17 | 5113995 | 5'Flank | SBS | G | T |
| INDIA 8582 | Unknown | chr6 | 77250040 | IGR | SBS | G | C |
| INDIA 8582 | SH2B1 | chr16 | 28868892 | Intron | SBS | G | C |
| INDIA 8582 | SUSD4 | chr1 | 223227787 | Intron | DEL | GGT | - |
| INDIA 8582 | TENM2 | chr5 | 167803047 | Intron | SBS | G | C |
| INDIA 8582 | MEMO1 | chr2 | 32002887 | Intron | DEL | C | - |
| INDIA 8582 | DIRC3 | chr2 | 217749003 | Intron | SBS | C | G |
| INDIA 8582 | Unknown | chr3 | 127146823 | IGR | SBS | G | C |
| INDIA 8582 | SIAH2-AS1 | chr3 | 150800184 | Intron | SBS | C | G |
| INDIA 8582 | CAMK2N2 | chr3 | 184261804 | 5'Flank | SBS | G | C |
| INDIA 8582 | GBA3 | chr4 | 22766771 | Intron | SBS | T | A |
| INDIA 8582 | ENOPH1 | chr4 | 82459203 | Intron | SBS | G | T |
| INDIA 8582 | CCDC170 | chr6 | 151494847 | Intron | SBS | G | C |
| INDIA 8582 | LINC-PINT | chr7 | 130984933 | Intron | SBS | G | C |
| INDIA 8582 | Unknown | chr8 | 48753941 | IGR | SBS | G | C |
| INDIA 8582 | NUMA1 | chr11 | 72038034 | Intron | SBS | A | C |
| INDIA 8582 | EPIC1 | chr22 | 47861529 | Intron | SBS | C | G |
| INDIA 8582 | DNAI1 | chr9 | 34493227 | Missense_Mutation | SBS | G | C |
| INDIA 8582 | IL21 | chr4 | 122616428 | Intron | SBS | C | G |
| INDIA 8582 | CNST | chr1 | 246591133 | Intron | SBS | C | G |
| INDIA 8582 | ITGA9 | chr3 | 37533274 | Intron | SBS | C | G |
| INDIA 8582 | SUCLG2-DT | chr3 | 67898152 | Intron | SBS | C | G |
| INDIA 8582 | SPEN-AS1 | chr1 | 15836235 | RNA | SBS | C | G |
| INDIA 8582 | PLXNA2 | chr1 | 208102143 | Intron | SBS | C | G |
| INDIA 8582 | GABRB2 | chr5 | 161483216 | Intron | SBS | C | G |
| INDIA 8582 | SIGLEC7 | chr19 | 51138978 | 5'Flank | SBS | A | T |
| INDIA 8582 | NMNAT3 | chr3 | 139673093 | Intron | SBS | C | G |
| INDIA 8582 | LINC01208 | chr3 | 176659564 | Intron | SBS | G | C |
| INDIA 8582 | ASTN2 | chr9 | 116434296 | Intron | DEL | A | - |
| INDIA 8582 | ABLIM1 | chr10 | 114696045 | Intron | SBS | T | G |
| INDIA 8582 | CDH13 | chr16 | 83199967 | Intron | SBS | G | C |
| INDIA 8582 | CASC15 | chr6 | 22639566 | Intron | SBS | G | C |
| INDIA 8582 | SGCZ | chr8 | 14383577 | Intron | SBS | T | G |
| INDIA 8582 | Unknown | chr10 | 10585192 | IGR | SBS | T | A |
| INDIA 8582 | Unknown | chr12 | 49938100 | IGR | SBS | C | G |
| INDIA 8582 | Unknown | chr13 | 20132632 | IGR | SBS | C | G |
| INDIA 8582 | MIR3681HG | chr2 | 12238327 | Intron | SBS | A | T |
| INDIA 8582 | DAP | chr5 | 10706821 | Intron | INS | - | A |
| INDIA 8582 | PDE1C | chr7 | 32250931 | Intron | SBS | G | C |
| INDIA 8582 | ACTN2 | chr1 | 236711935 | Intron | SBS | T | A |
| INDIA 8582 | MYLK4 | chr6 | 2701407 | Intron | SBS | G | C |
| INDIA 8582 | TENT5A | chr6 | 81543607 | Intron | SBS | G | C |
| INDIA 8582 | SH2B1 | chr16 | 28867406 | Missense_Mutation | SBS | G | C |
| INDIA 8582 | LRRC4B | chr19 | 50563867 | Intron | SBS | G | C |
| INDIA 8582 | ENST00000663378 | chr2 | 104929322 | RNA | SBS | T | A |
| INDIA 8582 | Unknown | chr2 | 176459549 | IGR | SBS | G | C |
| INDIA 8582 | PLXNA1 | chr3 | 127021130 | Intron | SBS | G | T |
| INDIA 8582 | RTCA | chr1 | 100267824 | Intron | SBS | G | C |
| INDIA 8582 | Unknown | chr1 | 242724948 | IGR | SBS | C | G |
| INDIA 8582 | LINC01926 | chr18 | 58835051 | 3'Flank | SBS | C | G |
| INDIA 8582 | DOK6 | chr18 | 69464420 | Intron | SBS | T | A |
| INDIA 8582 | MTMR3 | chr22 | 29980957 | Intron | SBS | C | G |
| INDIA 8582 | Unknown | chr18 | 27621809 | IGR | SBS | G | C |
| INDIA 8582 | KCNJ5 | chr11 | 128924881 | 3'Flank | SBS | G | C |
| INDIA 8582 | ACSL1 | chr4 | 184773029 | Intron | SBS | C | G |
| INDIA 8582 | PDZD2 | chr5 | 32026607 | Intron | SBS | C | G |
| INDIA 8582 | Unknown | chr7 | 93938776 | IGR | SBS | G | C |
| INDIA 8582 | MAPKAP1 | chr9 | 125605553 | Intron | SBS | C | G |
| INDIA 8582 | LINC01029 | chr18 | 77970782 | RNA | SBS | C | G |
| INDIA 8582 | ENC1 | chr5 | 74629550 | 3'UTR | INS | - | G |
| INDIA 8582 | ADGRB3 | chr6 | 68956280 | Intron | SBS | G | C |
| INDIA 8582 | Unknown | chr14 | 83716441 | IGR | SBS | C | G |

| | | | | | | | |
|---|---|---|---|---|---|---|---|
| INDIA 8582 | Unknown | chr6 | 12277842 | IGR | SBS | G | C |
| INDIA 8582 | NPSR1-AS1 | chr7 | 34425826 | Intron | SBS | C | G |
| INDIA 8582 | MAGI2 | chr7 | 78173353 | Intron | SBS | C | G |
| INDIA 8582 | ENST00000554181 | chr14 | 86946766 | Intron | SBS | T | A |
| INDIA 8582 | DCC | chr18 | 52829192 | Intron | SBS | G | C |
| INDIA 8583 | DNM2 | chr19 | 10805912 | Splice_Region | SBS | T | G |
| INDIA 8583 | SLC9A4 | chr2 | 102489596 | Intron | SBS | T | A |
| INDIA 8583 | Unknown | chr2 | 120331829 | IGR | SBS | C | G |
| INDIA 8583 | SND1 | chr7 | 127973875 | Intron | SBS | G | C |
| INDIA 8583 | APCDD1L | chr20 | 58460779 | 3'UTR | SBS | C | G |
| INDIA 8583 | MEIS2 | chr15 | 36989032 | Intron | SBS | C | G |
| INDIA 8583 | CCDC150 | chr2 | 196678295 | Intron | SBS | G | C |
| INDIA 8583 | ENST00000671082 | chr2 | 172351814 | Intron | SBS | G | C |
| INDIA 8583 | Unknown | chr1 | 203782842 | IGR | SBS | G | C |
| INDIA 8583 | Unknown | chr10 | 84988346 | IGR | SBS | C | G |
| INDIA 8583 | Unknown | chr2 | 146317189 | IGR | SBS | T | A |
| INDIA 8583 | Unknown | chr4 | 159389298 | IGR | SBS | C | A |
| INDIA 8583 | RAP1GDS1 | chr4 | 98263977 | Intron | SBS | G | C |
| INDIA 8583 | Unknown | chr2 | 85995346 | IGR | SBS | C | G |
| INDIA 8583 | TMEM202 | chr15 | 72406738 | Silent | SBS | C | G |
| INDIA 8583 | Unknown | chr2 | 4361686 | IGR | SBS | C | A |
| INDIA 8583 | Unknown | chrX | 70077086 | IGR | SBS | G | C |
| INDIA 8583 | WIPI2 | chr7 | 5230430 | Intron | SBS | A | T |
| INDIA 8583 | TTC23L | chr5 | 34841112 | Intron | SBS | T | A |
| INDIA 8583 | FAM180A | chr7 | 135746210 | Intron | SBS | G | C |
| INDIA 8583 | MIR3976HG | chr18 | 5875707 | Intron | SBS | C | G |
| INDIA 8583 | Unknown | chr14 | 75393823 | IGR | SBS | C | G |
| INDIA 8583 | CA8 | chr8 | 60200348 | Intron | SBS | T | A |
| INDIA 8583 | MATN2 | chr8 | 97933726 | Intron | SBS | T | G |
| INDIA 8583 | Unknown | chr9 | 97965037 | IGR | SBS | G | C |
| INDIA 8583 | Unknown | chr1 | 166365940 | IGR | SBS | G | C |
| INDIA 8583 | CHST8 | chr19 | 33681601 | Intron | SBS | C | G |
| INDIA 8583 | OSBPL10 | chr3 | 31850964 | Intron | DEL | T | - |
| INDIA 8583 | Unknown | chr4 | 95620894 | IGR | SBS | G | C |
| INDIA 8583 | Unknown | chr5 | 150576117 | IGR | SBS | G | C |
| INDIA 8583 | POR | chr7 | 75916635 | Intron | SBS | G | C |
| INDIA 8583 | ENST00000562656 | chr16 | 65271795 | Intron | SBS | C | G |
| INDIA 8583 | KCNB1 | chr20 | 49464495 | Intron | SBS | C | G |
| INDIA 8583 | DCHS2 | chr4 | 154428606 | Intron | SBS | G | C |
| INDIA 8583 | ENST00000643626 | chr16 | 51286280 | Intron | SBS | T | A |
| INDIA 8583 | CENPO | chr2 | 24823324 | 3'Flank | SBS | C | G |
| INDIA 8583 | Unknown | chr3 | 19868943 | IGR | SBS | G | C |
| INDIA 8583 | NOSTRIN | chr2 | 168814261 | Intron | SBS | G | C |
| INDIA 8583 | HLCS | chr21 | 36834999 | Intron | SBS | G | C |
| INDIA 8583 | Unknown | chr10 | 8370299 | IGR | SBS | C | G |
| INDIA 8583 | Unknown | chr9 | 95619406 | IGR | SBS | T | A |
| INDIA 8583 | HBS1L | chr6 | 135059254 | 5'Flank | SBS | G | C |
| INDIA 8583 | EEF1A1-AS1 | chr6 | 73526956 | Intron | SBS | G | C |
| INDIA 8583 | Unknown | chr7 | 57292158 | IGR | SBS | C | A |
| INDIA 8583 | BRWD1 | chr21 | 39232869 | Intron | SBS | G | C |
| INDIA 8583 | RALGPS1 | chr9 | 126962380 | Intron | SBS | G | C |
| INDIA 8583 | LMO4 | chr1 | 87331939 | Intron | SBS | T | A |
| INDIA 8583 | RALGPS1 | chr9 | 127016301 | Intron | SBS | C | G |
| INDIA 8583 | Unknown | chr4 | 54213214 | IGR | SBS | G | C |
| INDIA 8583 | Unknown | chr6 | 13835902 | IGR | SBS | A | C |
| INDIA 8583 | ZMYND11 | chr10 | 257698 | 3'Flank | SBS | G | C |
| INDIA 8583 | MUC16 | chr19 | 8936600 | Missense_Mutation | SBS | A | T |
| INDIA 8583 | GLYATL3 | chr6 | 49528366 | 3'Flank | SBS | G | C |
| INDIA 8583 | ATP2B2 | chr3 | 10597560 | Intron | SBS | C | G |
| INDIA 8583 | VEPH1 | chr3 | 157291707 | Intron | SBS | C | G |
| INDIA 8583 | STEAP2-AS1 | chr7 | 90038673 | Intron | SBS | G | C |
| INDIA 8583 | PRKG1 | chr10 | 51198220 | Intron | SBS | C | G |
| INDIA 8583 | ZNF423 | chr16 | 49625840 | Intron | SBS | C | G |
| INDIA 8583 | Unknown | chr3 | 127147813 | IGR | SBS | C | G |
| INDIA 8583 | Unknown | chr9 | 97966096 | IGR | SBS | G | C |
| INDIA 8583 | Unknown | chr11 | 116050760 | IGR | SBS | C | G |

| | | | | | | | |
|---|---|---|---|---|---|---|---|
| INDIA 8583 | ZPLD1 | chr3 | 102383011 | Intron | SBS | C | G |
| INDIA 8583 | GRIK5 | chr19 | 42046403 | Intron | SBS | G | C |
| INDIA 8583 | LNX1 | chr4 | 53544175 | Intron | SBS | G | C |
| INDIA 8583 | Unknown | chr14 | 43631541 | IGR | SBS | C | G |
| INDIA 8583 | NFATC1 | chr18 | 79446099 | Intron | SBS | G | C |
| INDIA 8583 | LINC01425 | chr21 | 21762839 | Intron | SBS | G | C |
| INDIA 8583 | Unknown | chr13 | 87717101 | IGR | SBS | A | T |
| INDIA 8583 | Unknown | chr18 | 73862080 | IGR | SBS | G | C |
| INDIA 8583 | NFILZ | chr19 | 8627088 | 5'Flank | SBS | C | G |
| INDIA 8583 | MRPS18A | chr6 | 43683622 | Intron | SBS | G | C |
| INDIA 8583 | ADO | chr10 | 62804031 | 5'Flank | SBS | C | G |
| INDIA 8583 | Unknown | chr13 | 108189820 | IGR | SBS | G | C |
| INDIA 8583 | ADK | chr10 | 74650000 | Intron | SBS | G | C |
| INDIA 8583 | CPNE1 | chr20 | 35640996 | Intron | SBS | C | G |
| INDIA 8583 | PLCB4 | chr20 | 9393341 | Intron | SBS | G | T |
| INDIA 8583 | Unknown | chr1 | 36493050 | IGR | SBS | G | C |
| INDIA 8583 | FLNB | chr3 | 58064612 | Intron | SBS | C | G |
| INDIA 8583 | ELMO1 | chr7 | 36983472 | Intron | SBS | C | G |
| INDIA 8583 | COL26A1 | chr7 | 101535303 | Intron | SBS | G | C |
| INDIA 8583 | TBC1D15 | chr12 | 71839870 | Intron | SBS | C | G |
| INDIA 8583 | Unknown | chr12 | 78576722 | IGR | SBS | C | G |
| INDIA 8583 | PDE3A | chr12 | 20407507 | Intron | SBS | C | G |
| INDIA 8583 | Unknown | chr7 | 6285177 | IGR | SBS | G | T |
| INDIA 8583 | KCNB2 | chr8 | 72823389 | Intron | SBS | C | G |
| INDIA 8583 | MBP | chr18 | 77110309 | Intron | SBS | C | A |
| INDIA 8583 | Unknown | chr8 | 5547658 | IGR | SBS | C | G |
| INDIA 8583 | ACTA1 | chr1 | 229426762 | 3'Flank | SBS | C | G |
| INDIA 8584 | Unknown | chr7 | 88408779 | IGR | SBS | C | A |
| INDIA 8584 | CD101 | chr1 | 117040013 | 3'Flank | SBS | T | A |
| INDIA 8584 | FANK1 | chr10 | 125986699 | Intron | SBS | G | C |
| INDIA 8584 | HSPA8 | chr11 | 123061548 | Intron | DEL | ACCCAG | - |
| INDIA 8584 | GPR26 | chr10 | 123671103 | Intron | SBS | G | C |
| INDIA 8584 | CADM2 | chr3 | 85626908 | Intron | SBS | T | A |
| INDIA 8584 | FBN1 | chr15 | 48632671 | Intron | SBS | C | G |
| INDIA 8584 | CDS2 | chr20 | 5176800 | Intron | SBS | C | A |
| INDIA 8584 | CA12 | chr15 | 63359110 | Intron | DEL | C | - |
| INDIA 8584 | ENST00000656336 | chr14 | 26741393 | Intron | SBS | A | T |
| INDIA 8584 | GALNT17 | chr7 | 71566291 | Intron | DEL | G | - |
| INDIA 8584 | Unknown | chr10 | 108588809 | IGR | SBS | G | C |
| INDIA 8584 | SNX30 | chr9 | 112816623 | Intron | SBS | A | T |
| INDIA 8584 | Unknown | chr7 | 153688630 | IGR | SBS | C | A |
| INDIA 8584 | ENST00000515232 | chr4 | 153037103 | Intron | SBS | C | G |
| INDIA 8584 | Unknown | chr22 | 35642980 | IGR | DEL | C | - |
| INDIA 8584 | NREP | chr5 | 111775318 | Intron | SBS | G | C |
| INDIA 8584 | PCSK5 | chr9 | 76229770 | Intron | SBS | G | C |
| INDIA 8584 | ENST00000421324 | chr10 | 78415133 | Intron | SBS | T | A |
| INDIA 8584 | MAP2K6 | chr17 | 69478950 | Intron | DEL | T | - |
| INDIA 8584 | FTO | chr16 | 53946768 | Intron | SBS | C | G |
| INDIA 8584 | ST3GAL1 | chr8 | 133514303 | Intron | SBS | G | T |
| INDIA 8584 | ENST00000651539 | chr14 | 40760131 | Intron | SBS | C | A |
| INDIA 8584 | LINC01982 | chr17 | 52472586 | Intron | SBS | C | A |
| INDIA 8584 | RCAN2 | chr6 | 46473568 | Intron | SBS | A | T |
| INDIA 8584 | Unknown | chr6 | 99794303 | IGR | SBS | T | A |
| INDIA 8584 | Unknown | chr8 | 90177251 | IGR | SBS | G | T |
| INDIA 8584 | MBD5 | chr2 | 148075181 | Intron | SBS | G | C |
| INDIA 8584 | Unknown | chr10 | 123349065 | IGR | DEL | CATGTCT | - |
| INDIA 8584 | Unknown | chr2 | 160893869 | IGR | SBS | G | C |
| INDIA 8584 | DPP6 | chr7 | 154334988 | Intron | SBS | T | G |
| INDIA 8584 | PDE4D | chr5 | 59818749 | Intron | INS | - | CT |
| INDIA 8584 | EPG5 | chr18 | 45962974 | Intron | SBS | G | C |
| INDIA 8584 | Unknown | chr8 | 42114426 | IGR | SBS | C | G |
| INDIA 8584 | PCSK2 | chr20 | 17409301 | Silent | SBS | C | A |
| INDIA 8584 | GRAMD1B | chr11 | 123429295 | 5'Flank | SBS | G | C |
| INDIA 8584 | CCSER1 | chr4 | 91128539 | Intron | SBS | T | G |
| INDIA 8584 | BMPER | chr7 | 34086720 | Intron | SBS | C | G |
| INDIA 8584 | LINC01091 | chr4 | 123556596 | Intron | SBS | C | A |

| | | | | | | | |
|---|---|---|---|---|---|---|---|
| INDIA 8584 | Unknown | chr10 | 130492646 | IGR | SBS | T | A |
| INDIA 8584 | TRIO | chr5 | 14198326 | Intron | SBS | G | T |
| INDIA 8584 | LINC02188 | chr16 | 86708337 | 5'Flank | SBS | G | C |
| INDIA 8584 | LINC01247 | chr2 | 6362607 | 3'Flank | SBS | G | C |
| INDIA 8584 | Unknown | chr12 | 129037303 | IGR | SBS | C | G |
| INDIA 8584 | ENST00000622926 | chr8 | 48431674 | 5'Flank | SBS | A | C |
| INDIA 8584 | KLHL12 | chr1 | 202896675 | Intron | DEL | TCTTCTAATC | - |
| INDIA 8585 | STK3 | chr8 | 98542614 | Intron | SBS | G | C |
| INDIA 8585 | GRXCR2 | chr5 | 145861337 | Intron | SBS | C | G |
| INDIA 8585 | CCND1 | chr11 | 69656293 | 3'Flank | SBS | C | G |
| INDIA 8585 | ELP3 | chr8 | 28106760 | Missense_Mutation | SBS | C | G |
| INDIA 8585 | Unknown | chrX | 16547219 | IGR | SBS | C | G |
| INDIA 8585 | ENST00000562922 | chr9 | 27830015 | Intron | SBS | C | G |
| INDIA 8585 | LINC00309 | chr2 | 64200176 | Intron | SBS | G | C |
| INDIA 8585 | ENST00000651847 | chr5 | 126047920 | Intron | SBS | G | C |
| INDIA 8585 | SPTB | chr14 | 64783705 | Intron | SBS | G | C |
| INDIA 8585 | Unknown | chr5 | 2519952 | IGR | SBS | C | G |
| INDIA 8585 | Unknown | chr5 | 162195632 | IGR | SBS | G | C |
| INDIA 8585 | ZZEF1 | chr17 | 4084569 | Intron | SBS | C | G |
| INDIA 8585 | ZMAT4 | chr8 | 40872712 | Intron | SBS | G | C |
| INDIA 8585 | PTPRM | chr18 | 8011085 | Intron | SBS | G | C |
| INDIA 8585 | Unknown | chr8 | 101003734 | IGR | SBS | C | G |
| INDIA 8585 | Unknown | chr5 | 43998165 | IGR | SBS | C | G |
| INDIA 8585 | JAKMIP2 | chr5 | 147685269 | Intron | SBS | G | C |
| INDIA 8585 | Unknown | chr14 | 74221565 | IGR | SBS | C | G |
| INDIA 8585 | SLC5A10 | chr17 | 19014770 | Intron | SBS | C | G |
| INDIA 8585 | DNAH5 | chr5 | 13718524 | Intron | SBS | G | C |
| INDIA 8585 | KRBA2 | chr17 | 8369060 | 3'UTR | SBS | G | C |
| INDIA 8585 | CYP4Z1 | chr1 | 47068875 | Intron | SBS | G | C |
| INDIA 8585 | LINC01847 | chr5 | 159832522 | Intron | SBS | G | C |
| INDIA 8585 | LRP1B | chr2 | 141917705 | Intron | SBS | G | C |
| INDIA 8585 | Unknown | chr9 | 108402729 | IGR | SBS | C | G |
| INDIA 8585 | VAV2 | chr9 | 133809625 | Intron | SBS | C | G |
| INDIA 8585 | RETREG1 | chr5 | 16569420 | Intron | SBS | G | C |
| INDIA 8585 | MAP7D2 | chrX | 20100048 | Intron | SBS | T | A |
| INDIA 8585 | Unknown | chr9 | 13373953 | IGR | SBS | C | G |
| INDIA 8585 | LIMK2 | chr22 | 31235563 | Intron | SBS | G | C |
| INDIA 8585 | DENND1B | chr1 | 197680544 | Intron | SBS | G | C |
| INDIA 8585 | ENST00000521714 | chr8 | 33982003 | Intron | SBS | G | C |
| INDIA 8585 | FAM193B | chr5 | 177541318 | Intron | SBS | C | G |
| INDIA 8585 | TMEM53 | chr1 | 44673607 | Intron | SBS | G | C |
| INDIA 8585 | Unknown | chr5 | 173355153 | IGR | SBS | G | C |
| INDIA 8585 | SCARA5 | chr8 | 27871613 | 3'UTR | SBS | C | G |
| INDIA 8585 | Unknown | chrX | 17241780 | IGR | SBS | C | G |
| INDIA 8585 | TRIM9 | chr14 | 51052288 | Intron | SBS | G | C |
| INDIA 8585 | AP1B1 | chr22 | 29349916 | Intron | SBS | G | C |
| INDIA 8585 | Unknown | chr5 | 120141182 | IGR | SBS | G | C |
| INDIA 8585 | Unknown | chrX | 20613449 | IGR | SBS | C | G |
| INDIA 8585 | COL5A2 | chr2 | 189179634 | 5'UTR | SBS | G | C |
| INDIA 8585 | TMOD1 | chr9 | 97528141 | Intron | SBS | C | G |
| INDIA 8585 | CSMD2 | chr1 | 34158509 | Intron | SBS | G | C |
| INDIA 8585 | AVPR1B-DT | chr1 | 206131372 | 3'Flank | SBS | G | C |
| INDIA 8585 | ENST00000435237 | chr2 | 21364630 | Intron | SBS | C | G |
| INDIA 8585 | Unknown | chr9 | 108693081 | IGR | SBS | G | C |
| INDIA 8585 | OXCT1 | chr5 | 41825251 | Intron | SBS | G | C |
| INDIA 8585 | PTPRD | chr9 | 8942400 | Intron | SBS | G | C |
| INDIA 8585 | Unknown | chr14 | 83379198 | IGR | SBS | G | C |
| INDIA 8585 | GNA12 | chr7 | 2760294 | Intron | SBS | C | G |
| INDIA 8585 | Unknown | chrX | 39827445 | IGR | SBS | G | C |
| INDIA 8585 | COL23A1 | chr5 | 178287102 | Intron | SBS | G | T |
| INDIA 8585 | RAB31 | chr18 | 9743818 | Intron | SBS | G | C |
| INDIA 8585 | LINC01847 | chr5 | 159833645 | Intron | SBS | G | C |
| INDIA 8585 | Unknown | chr2 | 219676569 | IGR | SBS | C | G |
| INDIA 8585 | POLA1 | chrX | 24986275 | Intron | SBS | G | C |
| INDIA 8585 | SNTB1 | chr8 | 120588250 | Intron | SBS | C | G |
| INDIA 8585 | LINC01889 | chr2 | 125733648 | Intron | SBS | A | T |

| | | | | | | | |
|---|---|---|---|---|---|---|---|
| INDIA 8585 | Unknown | chr5 | 15969518 | IGR | SBS | G | C |
| INDIA 8585 | PALM2AKAP2 | chr9 | 110145114 | Intron | SBS | G | C |
| INDIA 8585 | THSD7B | chr2 | 137631030 | Intron | SBS | G | C |
| INDIA 8585 | Unknown | chr13 | 104992035 | IGR | SBS | G | C |
| INDIA 8585 | ENST00000442996 | chr2 | 174615931 | Intron | SBS | C | G |
| INDIA 8585 | RIPK2-DT | chr8 | 89718488 | Intron | INS | - | TT |
| INDIA 8585 | ZNF513 | chr2 | 27380282 | Intron | SBS | C | G |
| INDIA 8585 | Unknown | chrX | 63044810 | IGR | SBS | G | C |
| INDIA 8585 | FAT3 | chr11 | 92314993 | Intron | SBS | G | C |
| INDIA 8585 | Unknown | chr7 | 4383025 | IGR | SBS | C | G |
| INDIA 8585 | SCRN1 | chr7 | 29922388 | 3'UTR | SBS | C | G |
| INDIA 8585 | GRM6 | chr5 | 178982844 | Intron | SBS | C | G |
| INDIA 8585 | GLI3 | chr7 | 42017432 | Intron | SBS | C | G |
| INDIA 8585 | Unknown | chr8 | 134956677 | IGR | SBS | C | G |
| INDIA 8585 | Unknown | chr8 | 96398234 | IGR | SBS | G | C |
| INDIA 8585 | ENST00000667088 | chr7 | 36020072 | Intron | SBS | C | G |
| INDIA 8585 | PRICKLE1 | chr12 | 42527047 | Intron | SBS | G | C |
| INDIA 8585 | ADAM7-AS1 | chr8 | 24719907 | Intron | SBS | G | C |
| INDIA 8585 | CACNA1E | chr1 | 181550070 | Intron | SBS | G | C |
| INDIA 8585 | MPV17 | chr2 | 27310042 | Intron | SBS | G | C |
| INDIA 8585 | SDK1 | chr7 | 3953305 | Intron | SBS | C | G |
| INDIA 8585 | Unknown | chr2 | 30174776 | IGR | SBS | G | C |
| INDIA 8585 | Unknown | chr1 | 5826548 | IGR | SBS | G | C |
| INDIA 8585 | Unknown | chr5 | 100717664 | IGR | SBS | G | C |
| INDIA 8585 | SAMD4A | chr14 | 54728113 | Intron | SBS | C | G |
| INDIA 8585 | CALN1 | chr7 | 72403934 | Intron | SBS | C | G |
| INDIA 8585 | Unknown | chr1 | 61561023 | IGR | SBS | G | C |
| INDIA 8585 | RAB32 | chr6 | 146551075 | Intron | SBS | G | C |
| INDIA 8585 | Unknown | chr8 | 97588950 | IGR | SBS | G | C |
| INDIA 8585 | SHC3 | chr9 | 89009233 | 3'UTR | SBS | G | C |
| INDIA 8585 | GLRX5 | chr14 | 95537800 | Intron | SBS | G | C |
| INDIA 8585 | RHOBTB3 | chr5 | 95779318 | Intron | SBS | C | G |
| INDIA 8585 | Unknown | chr7 | 45122607 | IGR | SBS | C | G |
| INDIA 8585 | Unknown | chr7 | 46544527 | IGR | SBS | C | G |
| INDIA 8585 | ENST00000666810 | chr8 | 92669880 | Intron | SBS | G | C |
| INDIA 8585 | PSAT1 | chr9 | 78306129 | Intron | SBS | C | G |
| INDIA 8586 | CASC15 | chr6 | 21813580 | Intron | SBS | G | C |
| INDIA 8586 | ARHGEF4 | chr2 | 130883022 | Intron | SBS | C | G |
| INDIA 8586 | EPGN | chr4 | 74305371 | 5'Flank | SBS | G | C |
| INDIA 8586 | Unknown | chr13 | 57331917 | IGR | SBS | G | C |
| INDIA 8586 | APP | chr21 | 25891709 | Intron | SBS | C | G |
| INDIA 8586 | Unknown | chrX | 22022505 | IGR | SBS | G | C |
| INDIA 8586 | MORN4 | chr10 | 97616381 | Missense_Mutation | SBS | C | G |
| INDIA 8586 | LINC00927 | chr15 | 80270158 | Intron | SBS | C | G |
| INDIA 8586 | Unknown | chr10 | 121391906 | IGR | SBS | G | C |
| INDIA 8586 | ENST00000515232 | chr4 | 153080220 | Intron | SBS | C | G |
| INDIA 8586 | RTCB | chr22 | 32384367 | 3'Flank | SBS | C | G |
| INDIA 8586 | CLDN2 | chrX | 106929400 | 3'UTR | SBS | C | G |
| INDIA 8586 | TPM3 | chr1 | 154158773 | 3'Flank | SBS | C | G |
| INDIA 8586 | Unknown | chr14 | 96616536 | IGR | SBS | C | G |
| INDIA 8586 | DGKH | chr13 | 42145901 | Intron | SBS | C | G |
| INDIA 8586 | Unknown | chr16 | 8218568 | IGR | SBS | G | C |
| INDIA 8586 | SYN3 | chr22 | 32919103 | Intron | SBS | C | G |
| INDIA 8586 | BTRC | chr10 | 101526284 | Intron | SBS | G | C |
| INDIA 8586 | MARCHF1 | chr4 | 163612629 | Missense_Mutation | SBS | C | G |
| INDIA 8586 | UNC5C | chr4 | 95252409 | Intron | SBS | C | G |
| INDIA 8586 | Unknown | chr2 | 2378443 | IGR | SBS | G | C |
| INDIA 8586 | RASGEF1B | chr4 | 81882092 | Intron | SBS | C | G |
| INDIA 8586 | ENST00000657016 | chr13 | 53274428 | Intron | SBS | C | G |
| INDIA 8586 | Unknown | chr8 | 20625606 | IGR | SBS | C | G |
| INDIA 8586 | DMD | chrX | 32158640 | Intron | SBS | C | G |
| INDIA 8586 | Unknown | chr1 | 23226897 | IGR | SBS | T | G |
| INDIA 8586 | Unknown | chr4 | 124781946 | IGR | SBS | G | C |
| INDIA 8586 | ROS1 | chr6 | 117342115 | Intron | SBS | G | C |
| INDIA 8586 | Unknown | chr11 | 39961015 | IGR | SBS | G | C |
| INDIA 8586 | Unknown | chr5 | 24432337 | IGR | SBS | C | G |

| | | | | | | | |
|---|---|---|---|---|---|---|---|
| INDIA 8586 | Unknown | chr14 | 95982953 | IGR | SBS | C | G |
| INDIA 8586 | ENST00000578831 | chr18 | 22741785 | Intron | SBS | G | C |
| INDIA 8586 | Unknown | chr4 | 103248192 | IGR | SBS | C | G |
| INDIA 8586 | TRABD2B | chr1 | 47875789 | Intron | SBS | C | G |
| INDIA 8586 | LINC02994 | chr4 | 83880784 | Intron | SBS | G | C |
| INDIA 8586 | Unknown | chr4 | 132766517 | IGR | SBS | G | C |
| INDIA 8586 | U3 | chr5 | 149700582 | 5'Flank | SBS | C | A |
| INDIA 8586 | DISC1FP1 | chr11 | 90568188 | Intron | SBS | T | G |
| INDIA 8586 | KCNK3 | chr2 | 26723872 | Intron | SBS | G | C |
| INDIA 8586 | ENST00000681682 | chr4 | 111224792 | Intron | SBS | G | C |
| INDIA 8586 | Unknown | chr10 | 72248059 | IGR | SBS | C | A |
| INDIA 8586 | Unknown | chr12 | 94844245 | IGR | SBS | G | C |
| INDIA 8586 | ENST00000636248 | chr17 | 62911728 | Intron | SBS | C | G |
| INDIA 8586 | Unknown | chr8 | 25048419 | IGR | SBS | C | G |
| INDIA 8586 | Unknown | chr10 | 121099471 | IGR | SBS | G | C |
| INDIA 8586 | FAM149A | chr4 | 186172137 | 3'UTR | SBS | C | G |
| INDIA 8586 | KCTD8 | chr4 | 44217321 | Intron | SBS | T | G |
| INDIA 8586 | CACNA1C | chr12 | 2286085 | Intron | SBS | G | C |
| INDIA 8586 | PLA2G2E | chr1 | 19918890 | 3'Flank | SBS | C | G |
| INDIA 8586 | NRG2 | chr5 | 139968935 | Intron | SBS | G | C |
| INDIA 8586 | Unknown | chr9 | 120115494 | IGR | SBS | G | C |
| INDIA 8586 | ENST00000661809 | chr4 | 126457239 | Intron | SBS | G | C |
| INDIA 8586 | SCML4 | chr6 | 107825329 | 5'Flank | SBS | G | C |
| INDIA 8586 | RASSF8 | chr12 | 26045550 | Intron | SBS | G | C |
| INDIA 8586 | RILPL1 | chr12 | 123485818 | Intron | SBS | G | C |
| INDIA 8586 | ENST00000559600 | chr15 | 45919535 | Intron | SBS | G | C |
| INDIA 8586 | RFFL | chr17 | 35021882 | Intron | SBS | C | G |
| INDIA 8586 | Unknown | chr2 | 190585052 | IGR | SBS | G | C |
| INDIA 8586 | CCNY | chr10 | 35458221 | Intron | SBS | C | G |
| INDIA 8586 | CNTNAP2 | chr7 | 147225717 | Intron | SBS | C | G |
| INDIA 8586 | PRKCA | chr17 | 66447668 | Intron | SBS | C | G |
| INDIA 8586 | GLYAT | chr11 | 58708937 | 3'UTR | SBS | C | G |
| INDIA 8586 | GCDH | chr19 | 12889319 | 5'Flank | SBS | C | G |
| INDIA 8586 | DGKZ | chr11 | 46345702 | 5'Flank | SBS | G | C |
| INDIA 8586 | GMDS | chr6 | 1743252 | Intron | SBS | C | G |
| INDIA 8586 | Unknown | chr8 | 47030404 | IGR | SBS | C | G |
| INDIA 8586 | ENST00000649415 | chr11 | 39026128 | RNA | SBS | C | G |
| INDIA 8586 | MFGE8 | chr15 | 88912288 | Intron | SBS | G | C |
| INDIA 8586 | RNF175 | chr4 | 153743851 | Intron | SBS | C | G |
| INDIA 8586 | PKHD1 | chr6 | 51740937 | Intron | SBS | C | G |
| INDIA 8586 | TNPO3 | chr7 | 129030839 | Intron | SBS | C | G |
| INDIA 8586 | LINC01344 | chr1 | 182198042 | Intron | DEL | C | - |
| INDIA 8586 | DGKG | chr3 | 186275506 | Intron | SBS | G | C |
| INDIA 8586 | LINC02267 | chr4 | 96821958 | 3'Flank | SBS | G | C |
| INDIA 8586 | Unknown | chr4 | 176529301 | IGR | SBS | G | C |
| INDIA 8586 | Unknown | chr5 | 166598363 | IGR | SBS | C | G |
| INDIA 8586 | AGBL4 | chr1 | 49247458 | Intron | DEL | T | - |
| INDIA 8586 | CTNND2 | chr5 | 11360830 | Intron | SBS | C | G |
| INDIA 8586 | Unknown | chr13 | 44757823 | IGR | SBS | C | G |
| INDIA 8586 | FGF18 | chr5 | 171428324 | Intron | SBS | G | C |
| INDIA 8586 | UBQLN4 | chr1 | 156051695 | Intron | SBS | C | G |
| INDIA 8586 | PROC | chr2 | 127429658 | 3'Flank | SBS | G | C |
| INDIA 8586 | RAD21 | chr8 | 116850438 | Intron | INS | - | A |
| INDIA 8586 | LRRC4C | chr11 | 41344964 | Intron | SBS | C | G |
| INDIA 8586 | Unknown | chr10 | 84654189 | IGR | SBS | C | G |
| INDIA 8586 | DGKZ | chr11 | 46345782 | 5'Flank | SBS | G | C |
| INDIA 8586 | CNGA4 | chr11 | 6235198 | 5'Flank | SBS | G | C |
| INDIA 8586 | FMNL2 | chr2 | 152511801 | Intron | INS | - | G |
| INDIA 8586 | LAMA1 | chr18 | 6969709 | Intron | SBS | A | T |
| INDIA 8586 | SOX2-OT | chr3 | 181731058 | Intron | SBS | C | G |
| INDIA 8586 | ASAP1 | chr8 | 130229081 | Intron | SBS | C | G |
| INDIA 8586 | FAM120A | chr9 | 93527202 | Nonsense_Mutation | SBS | C | G |
| INDIA 8586 | HTR2A | chr13 | 46897050 | 5'UTR | SBS | C | G |
| INDIA 8586 | Unknown | chr1 | 41400763 | IGR | DEL | TACACT | - |
| INDIA 8586 | FCHSD2 | chr11 | 72902436 | Intron | SBS | G | C |
| INDIA 8586 | CHRDL1 | chrX | 110791588 | Intron | SBS | C | G |

| | | | | | | | |
|---|---|---|---|---|---|---|---|
| INDIA 8587 | MIR124-1HG | chr8 | 9897421 | 3'Flank | SBS | C | G |
| INDIA 8587 | Unknown | chr12 | 126791540 | IGR | SBS | C | G |
| INDIA 8587 | PTPN21 | chr14 | 88501143 | Intron | SBS | C | G |
| INDIA 8587 | BTBD9 | chr6 | 38233059 | Intron | SBS | C | G |
| INDIA 8587 | PAPLN | chr14 | 73239319 | Intron | SBS | G | C |
| INDIA 8587 | MAMLD1 | chrX | 150510863 | Intron | SBS | G | C |
| INDIA 8587 | ENST00000413066 | chr9 | 7938309 | 5'Flank | SBS | G | C |
| INDIA 8587 | SDC2 | chr8 | 96604178 | Intron | SBS | G | C |
| INDIA 8587 | Unknown | chr9 | 1287418 | IGR | SBS | C | G |
| INDIA 8587 | HUWE1 | chrX | 53585079 | Missense_Mutation | SBS | C | G |
| INDIA 8587 | Unknown | chr6 | 99786899 | IGR | SBS | C | G |
| INDIA 8587 | Unknown | chrX | 109353053 | IGR | SBS | G | C |
| INDIA 8587 | CCNK | chr14 | 99487905 | Intron | SBS | G | C |
| INDIA 8587 | NTM | chr11 | 132047938 | Intron | SBS | C | G |
| INDIA 8587 | Unknown | chr12 | 69053548 | IGR | SBS | C | A |
| INDIA 8587 | CERS6 | chr2 | 168735205 | Intron | SBS | C | G |
| INDIA 8587 | Unknown | chr9 | 7266922 | IGR | SBS | T | A |
| INDIA 8587 | Unknown | chr5 | 15342877 | IGR | SBS | G | C |
| INDIA 8587 | Unknown | chrX | 66448319 | IGR | SBS | C | G |
| INDIA 8587 | PDE6C | chr10 | 93654670 | Intron | SBS | C | G |
| INDIA 8587 | Unknown | chr6 | 51272276 | IGR | SBS | C | G |
| INDIA 8587 | ENST00000662174 | chr2 | 105611550 | Intron | SBS | G | C |
| INDIA 8587 | GPRC5A | chr12 | 12918922 | 3'Flank | SBS | C | G |
| INDIA 8587 | COX7C | chr5 | 86617580 | 5'Flank | SBS | G | C |
| INDIA 8587 | CLMN | chr14 | 95311793 | Intron | SBS | G | C |
| INDIA 8587 | C9orf92 | chr9 | 16389353 | Intron | SBS | A | T |
| INDIA 8587 | Unknown | chr16 | 1055084 | IGR | SBS | T | A |
| INDIA 8587 | MEG3 | chr14 | 100810788 | Intron | SBS | G | C |
| INDIA 8587 | WDR20 | chr14 | 102183604 | Intron | SBS | G | C |
| INDIA 8587 | MPRIP | chr17 | 17176834 | Intron | SBS | C | G |
| INDIA 8587 | KC6 | chr18 | 41521319 | Intron | SBS | T | A |
| INDIA 8587 | Unknown | chrX | 24256048 | IGR | SBS | C | G |
| INDIA 8587 | UVRAG | chr11 | 75901005 | Intron | SBS | C | G |
| INDIA 8587 | Unknown | chr12 | 5201504 | IGR | SBS | C | G |
| INDIA 8587 | Unknown | chr1 | 182751244 | IGR | SBS | G | C |
| INDIA 8587 | Unknown | chr9 | 119792464 | IGR | SBS | G | C |
| INDIA 8587 | Unknown | chr8 | 93073956 | IGR | SBS | G | C |
| INDIA 8587 | ENST00000485384 | chr3 | 109661393 | Intron | SBS | C | G |
| INDIA 8587 | Unknown | chrX | 30077946 | IGR | SBS | G | C |
| INDIA 8587 | AMPD3 | chr11 | 10437276 | Intron | SBS | C | G |
| INDIA 8587 | IQSEC1 | chr3 | 13220288 | Intron | SBS | C | G |
| INDIA 8587 | HUWE1 | chrX | 53562279 | Intron | SBS | G | C |
| INDIA 8587 | Unknown | chr8 | 116440841 | IGR | SBS | G | C |
| INDIA 8587 | FREM1 | chr9 | 14746406 | Missense_Mutation | SBS | C | G |
| INDIA 8587 | PAX5 | chr9 | 36968433 | Intron | SBS | T | A |
| INDIA 8587 | COX10 | chr17 | 14148030 | Intron | SBS | G | C |
| INDIA 8587 | PLEKHD1 | chr14 | 69494723 | Intron | SBS | G | C |
| INDIA 8587 | LINC02432 | chr4 | 141346909 | Intron | SBS | C | G |
| INDIA 8587 | LINC02089 | chr17 | 52902339 | 3'Flank | SBS | G | C |
| INDIA 8587 | PALM2AKAP2 | chr9 | 109998106 | Intron | SBS | G | C |
| INDIA 8587 | ASS1 | chr9 | 130464631 | Intron | SBS | G | C |
| INDIA 8587 | FGF13 | chrX | 138646440 | Intron | SBS | A | T |
| INDIA 8587 | CDK15 | chr2 | 201862820 | Intron | SBS | T | G |
| INDIA 8587 | Unknown | chr2 | 221774167 | IGR | SBS | G | C |
| INDIA 8587 | MPPED2 | chr11 | 30571098 | Intron | SBS | C | G |
| INDIA 8587 | RPIA | chr2 | 88687233 | 5'Flank | SBS | C | G |
| INDIA 8587 | LINC02325 | chr14 | 97524050 | Intron | SBS | C | G |
| INDIA 8587 | Unknown | chr9 | 90603657 | IGR | SBS | C | G |
| INDIA 8587 | NGEF | chr2 | 232894658 | Intron | SBS | G | C |
| INDIA 8587 | Unknown | chr2 | 88441719 | IGR | SBS | C | A |
| INDIA 8587 | CLMP | chr11 | 123091955 | Intron | SBS | C | G |
| INDIA 8587 | ATP6V1G1 | chr9 | 114587471 | 5'Flank | SBS | G | C |
| INDIA 8587 | Unknown | chr10 | 91090838 | IGR | SBS | C | G |
| INDIA 8587 | MDN1 | chr6 | 89745561 | Missense_Mutation | SBS | C | G |
| INDIA 8587 | SNX9 | chr6 | 157781440 | Intron | SBS | G | T |
| INDIA 8587 | NDUFAF6 | chr8 | 95051031 | Intron | SBS | G | C |

| | | | | | | | |
|---|---|---|---|---|---|---|---|
| INDIA 8587 | MATN2 | chr8 | 98036025 | 3'UTR | SBS | C | G |
| INDIA 8587 | LINGO2 | chr9 | 28786030 | Intron | SBS | A | T |
| INDIA 8587 | TJP2 | chr9 | 69164552 | Intron | SBS | G | C |
| INDIA 8587 | Unknown | chr9 | 107276537 | IGR | SBS | C | G |
| INDIA 8587 | SORCS1 | chr10 | 106715706 | Intron | SBS | C | G |
| INDIA 8587 | FRY | chr13 | 32142751 | Intron | SBS | C | G |
| INDIA 8587 | ENST00000580366 | chr18 | 32884877 | Intron | SBS | C | G |
| INDIA 8587 | TENM1 | chrX | 124464816 | Intron | SBS | C | G |
| INDIA 8587 | Unknown | chr10 | 9137513 | IGR | DEL | G | - |
| INDIA 8587 | Unknown | chr2 | 23213920 | IGR | SBS | G | C |
| INDIA 8587 | ENST00000664438 | chr9 | 13775473 | Intron | SBS | G | C |
| INDIA 8587 | IGSF22 | chr11 | 18730175 | 5'Flank | SBS | C | G |
| INDIA 8587 | FRMD3 | chr9 | 83466219 | Intron | SBS | G | C |
| INDIA 8587 | ENST00000382641 | chr22 | 26675685 | 5'Flank | SBS | G | T |
| INDIA 8587 | KCNJ8 | chr12 | 21763464 | 3'Flank | SBS | C | G |
| INDIA 8587 | CEMIP2 | chr9 | 71727258 | Intron | SBS | A | C |
| INDIA 8587 | LINC01440 | chr20 | 55682143 | Intron | SBS | A | T |
| INDIA 8587 | PARD3B | chr2 | 205402823 | Intron | SBS | G | C |
| INDIA 8587 | LINC02458 | chr12 | 89041758 | Intron | SBS | C | G |
| INDIA 8587 | Unknown | chr13 | 55268544 | IGR | SBS | C | G |
| INDIA 8587 | Unknown | chr13 | 91157336 | IGR | SBS | C | G |
| INDIA 8587 | Unknown | chr9 | 32109882 | IGR | SBS | G | C |
| INDIA 8587 | Unknown | chr14 | 97902695 | IGR | SBS | G | C |
| INDIA 8587 | FGGY | chr1 | 59692822 | Intron | SBS | G | C |
| INDIA 8587 | Unknown | chr8 | 102570125 | IGR | SBS | C | G |
| INDIA 8587 | ST8SIA5 | chr18 | 46677109 | 3'UTR | SBS | G | C |
| INDIA 8588 | SGCZ | chr8 | 14791807 | Intron | SBS | A | C |
| INDIA 8588 | Unknown | chr5 | 20950521 | IGR | SBS | T | G |
| INDIA 8588 | LINC02196 | chr5 | 7219173 | RNA | DEL | C | - |
| INDIA 8588 | Unknown | chr11 | 20193543 | IGR | SBS | T | A |
| INDIA 8588 | MDN1 | chr6 | 89712735 | Missense_Mutation | SBS | T | A |
| INDIA 8588 | ACACB | chr12 | 109115574 | 5'Flank | SBS | G | C |
| INDIA 8588 | CDH13 | chr16 | 82927911 | Intron | SBS | T | A |
| INDIA 8588 | Unknown | chr15 | 87213997 | IGR | SBS | C | G |
| INDIA 8588 | MYO10 | chr5 | 16828694 | Intron | SBS | C | G |
| INDIA 8588 | PAG1 | chr8 | 81110983 | Intron | SBS | T | A |
| INDIA 8588 | LDLRAD3 | chr11 | 36152721 | Intron | SBS | A | C |
| INDIA 8588 | Unknown | chr2 | 36061702 | IGR | SBS | T | A |
| INDIA 8588 | ARNT2 | chr15 | 80575259 | Intron | SBS | T | A |
| INDIA 8588 | Unknown | chr7 | 122286679 | IGR | SBS | T | A |
| INDIA 8588 | Unknown | chr21 | 14787481 | IGR | SBS | C | G |
| INDIA 8588 | Unknown | chr2 | 129290864 | IGR | SBS | G | C |
| INDIA 8588 | LINC01122 | chr2 | 58865468 | Intron | SBS | T | A |
| INDIA 8588 | Unknown | chr2 | 5131535 | IGR | SBS | A | T |
| INDIA 8588 | NTM | chr11 | 131822082 | Intron | SBS | C | G |
| INDIA 8588 | Unknown | chr6 | 76153253 | IGR | SBS | A | T |
| INDIA 8588 | Unknown | chr8 | 135812800 | IGR | SBS | C | A |
| INDIA 8588 | LINC02109 | chr5 | 29147485 | Intron | SBS | C | G |
| INDIA 8588 | GJD2 | chr15 | 34756261 | 5'Flank | SBS | C | G |
| INDIA 8588 | MEF2C | chr5 | 88733557 | Intron | SBS | C | A |
| INDIA 8588 | RPH3A | chr12 | 112837398 | Intron | SBS | T | A |
| INDIA 8588 | Unknown | chr5 | 22984332 | IGR | SBS | G | T |
| INDIA 8588 | TMEM164 | chrX | 110053298 | Intron | SBS | T | A |
| INDIA 8588 | Unknown | chr5 | 2312315 | IGR | SBS | C | G |
| INDIA 8588 | Unknown | chr14 | 96183938 | IGR | SBS | C | G |
| INDIA 8588 | Unknown | chr3 | 194974808 | IGR | SBS | T | A |
| INDIA 8588 | Unknown | chr2 | 139969560 | IGR | SBS | T | G |
| INDIA 8588 | Unknown | chr2 | 84376872 | IGR | SBS | T | A |
| INDIA 8588 | Unknown | chr4 | 178552891 | IGR | SBS | A | C |
| INDIA 8588 | ENST00000357045 | chr2 | 177602651 | 5'Flank | SBS | C | A |
| INDIA 8588 | Unknown | chr8 | 73226033 | IGR | SBS | C | G |
| INDIA 8588 | Unknown | chr9 | 12620648 | IGR | DEL | C | - |
| INDIA 8588 | ENST00000664749 | chr2 | 62569404 | Intron | SBS | A | T |
| INDIA 8588 | Unknown | chr5 | 5355912 | IGR | SBS | C | G |
| INDIA 8588 | Unknown | chr7 | 38956689 | IGR | SBS | T | A |
| INDIA 8588 | LINC02955 | chr12 | 23082683 | Intron | SBS | T | A |

| | | | | | | | |
|---|---|---|---|---|---|---|---|
| INDIA 8588 | ENST00000562182 | chr17 | 22421367 | RNA | SBS | T | A |
| INDIA 8588 | Unknown | chr2 | 10971967 | IGR | SBS | G | C |
| INDIA 8588 | Unknown | chr2 | 186975415 | IGR | DEL | G | - |
| INDIA 8588 | LINC02008 | chr3 | 82100644 | Intron | SBS | T | A |
| INDIA 8588 | MECOM | chr3 | 169402356 | Intron | SBS | G | C |
| INDIA 8588 | SCFD2 | chr4 | 53024940 | Intron | SBS | T | A |
| INDIA 8588 | Unknown | chr5 | 3121985 | IGR | SBS | C | G |
| INDIA 8588 | LINC02162 | chr5 | 3461409 | RNA | DEL | C | - |
| INDIA 8588 | Unknown | chr6 | 132236103 | IGR | SBS | A | T |
| INDIA 8588 | PLXDC2 | chr10 | 19828144 | Intron | SBS | C | G |
| INDIA 8588 | NEURL1 | chr10 | 103568394 | Intron | SBS | C | G |
| INDIA 8588 | TF | chr3 | 133754239 | Intron | SBS | A | T |
| INDIA 8588 | ATOX1 | chr5 | 151758421 | Intron | SBS | G | C |
| INDIA 8588 | Unknown | chr9 | 103806393 | IGR | SBS | C | A |
| INDIA 8588 | CDH13 | chr16 | 83347862 | Intron | SBS | C | G |
| INDIA 8588 | MIR3976HG | chr18 | 5741974 | Intron | SBS | G | C |
| INDIA 8588 | SNTG2 | chr2 | 1293824 | Intron | SBS | A | T |
| INDIA 8588 | URI1 | chr19 | 29928517 | Intron | SBS | T | A |
| INDIA 8588 | Unknown | chr12 | 128160682 | IGR | SBS | C | A |
| INDIA 8588 | IFT43 | chr14 | 76071099 | Intron | SBS | A | T |
| INDIA 8588 | ENST00000580524 | chr18 | 1679490 | Intron | SBS | A | T |
| INDIA 8588 | MSRB3-AS1 | chr12 | 65531385 | Intron | SBS | C | A |
| INDIA 8588 | Unknown | chr14 | 42945177 | IGR | SBS | A | T |
| INDIA 8588 | LRP11 | chr6 | 149823598 | Intron | INS | - | GC |
| INDIA 8588 | LINC00299 | chr2 | 8344413 | Intron | SBS | C | G |
| INDIA 8588 | GALNT17 | chr7 | 71182463 | Intron | SBS | G | C |
| INDIA 8588 | Unknown | chr21 | 19930500 | IGR | SBS | C | G |
| INDIA 8588 | Unknown | chr1 | 84773327 | IGR | SBS | G | C |
| INDIA 8588 | Unknown | chr2 | 59078906 | IGR | DEL | C | - |
| INDIA 8588 | Unknown | chr5 | 2914930 | IGR | SBS | T | A |
| INDIA 8588 | CD2AP | chr6 | 47600497 | Intron | SBS | G | C |
| INDIA 8588 | SLC2A13 | chr12 | 39975849 | Intron | SBS | T | A |
| INDIA 8588 | Unknown | chr3 | 135740510 | IGR | SBS | G | C |
| INDIA 8588 | LINC02986 | chr8 | 80544744 | 3'Flank | SBS | G | C |
| INDIA 8588 | KRT75 | chr12 | 52437916 | 5'Flank | DEL | C | - |
| INDIA 8588 | Unknown | chr2 | 118536384 | IGR | SBS | A | C |
| INDIA 8588 | IRF4 | chr6 | 392544 | Intron | DEL | G | - |
| INDIA 8588 | Unknown | chr4 | 133461592 | IGR | SBS | C | G |
| INDIA 8588 | Unknown | chr12 | 68502314 | IGR | SBS | C | A |
| INDIA 8588 | MGC4859 | chr7 | 10470993 | Intron | SBS | C | A |
| INDIA 8588 | FAT3 | chr11 | 92275191 | Intron | SBS | C | A |
| INDIA 8588 | Unknown | chr3 | 73192206 | IGR | SBS | A | T |
| INDIA 8588 | Unknown | chr1 | 101199295 | IGR | SBS | C | A |
| INDIA 8588 | ENST00000435333 | chr1 | 247600418 | Intron | SBS | T | A |
| INDIA 8588 | UNC5C | chr4 | 95515724 | Intron | SBS | G | T |
| INDIA 8588 | ENST00000435148 | chr7 | 69229875 | Intron | INS | - | T |
| INDIA 8588 | MBTPS1 | chr16 | 84094278 | Intron | SBS | G | C |
| INDIA 8588 | Unknown | chr5 | 3749213 | IGR | SBS | G | C |
| INDIA 8588 | ENST00000622671 | chr12 | 25388292 | 5'Flank | INS | - | A |
| INDIA 8588 | ENST00000681880 | chr4 | 111537919 | Intron | SBS | C | G |
| INDIA 8588 | ADAM12 | chr10 | 126092338 | Intron | SBS | G | C |
| INDIA 8588 | LAMA5 | chr20 | 62363933 | Intron | SBS | C | A |
| INDIA 8590 | Unknown | chr3 | 39784055 | IGR | SBS | A | T |
| INDIA 8590 | CCDC175 | chr14 | 59562004 | Intron | SBS | C | G |
| INDIA 8590 | LRMDA | chr10 | 76119092 | Intron | SBS | G | C |
| INDIA 8590 | NAV3 | chr12 | 78130790 | Intron | SBS | A | T |
| INDIA 8590 | SFRP4 | chr7 | 37908633 | Intron | SBS | A | T |
| INDIA 8590 | NHEG1 | chr6 | 136977479 | 3'Flank | SBS | T | G |
| INDIA 8590 | STEEP1 | chrX | 119560287 | Missense_Mutation | SBS | C | G |
| INDIA 8590 | EPB41L4A | chr5 | 112194487 | Intron | DEL | TT | - |
| INDIA 8590 | ENST00000412197 | chr7 | 25548328 | Intron | SBS | T | G |
| INDIA 8590 | EPHA3 | chr3 | 89402274 | Intron | SBS | C | G |
| INDIA 8590 | TMEM232 | chr5 | 110564791 | Intron | SBS | A | C |
| INDIA 8590 | ENST00000673514 | chr4 | 33625913 | Intron | SBS | T | A |
| INDIA 8590 | Unknown | chr3 | 94330160 | IGR | SBS | G | C |
| INDIA 8590 | ENST00000446187 | chr7 | 68183307 | Intron | SBS | G | C |

| | | | | | | | |
|---|---|---|---|---|---|---|---|
| INDIA 8590 | MCU | chr10 | 72764336 | Intron | SBS | C | G |
| INDIA 8590 | RBFOX1 | chr16 | 6786360 | Intron | SBS | C | A |
| INDIA 8590 | NF2 | chr22 | 29604341 | Intron | SBS | T | A |
| INDIA 8590 | Unknown | chrX | 27969373 | IGR | SBS | G | C |
| INDIA 8590 | Unknown | chr7 | 30225973 | IGR | INS | - | AG |
| INDIA 8590 | LINC02006 | chr3 | 153848636 | Intron | SBS | T | A |
| INDIA 8590 | Unknown | chrX | 55337040 | IGR | SBS | G | C |
| INDIA 8590 | Unknown | chr8 | 21380458 | IGR | SBS | T | A |
| INDIA 8590 | Unknown | chr1 | 30185092 | IGR | SBS | C | A |
| INDIA 8590 | ST6GALNAC3 | chr1 | 76623551 | Intron | SBS | A | T |
| INDIA 8590 | Unknown | chr6 | 68165100 | IGR | SBS | A | T |
| INDIA 8590 | ENST00000438247 | chr2 | 1605245 | Intron | SBS | C | G |
| INDIA 8590 | GSN | chr9 | 121308952 | Intron | SBS | G | T |
| INDIA 8590 | Unknown | chr4 | 181669710 | IGR | SBS | G | T |
| INDIA 8590 | ZNRF3 | chr22 | 28956624 | Intron | SBS | G | C |
| INDIA 8590 | TULP4 | chr6 | 158507765 | 3'UTR | SBS | G | C |
| INDIA 8590 | NKAIN3 | chr8 | 62584466 | Intron | SBS | C | G |
| INDIA 8590 | Unknown | chrX | 83982557 | IGR | SBS | G | T |
| INDIA 8590 | FZD5 | chr2 | 207766794 | 3'UTR | SBS | G | C |
| INDIA 8590 | Unknown | chr22 | 49331336 | IGR | SBS | G | T |
| INDIA 8590 | Unknown | chr6 | 77412085 | IGR | SBS | G | C |
| INDIA 8590 | LINC01239 | chr9 | 22681500 | Intron | SBS | C | G |
| INDIA 8590 | Unknown | chr7 | 113726785 | IGR | DEL | C | - |
| INDIA 8590 | CDK14 | chr7 | 90693384 | Intron | DEL | TACTAG | - |
| INDIA 8590 | NRIP1 | chr21 | 15066841 | 5'Flank | SBS | T | G |
| INDIA 8590 | LINC01661 | chr1 | 106816022 | 5'Flank | SBS | T | A |
| INDIA 8590 | Unknown | chr2 | 102553117 | IGR | SBS | G | T |
| INDIA 8590 | GLRA1 | chr5 | 151870465 | Intron | SBS | C | G |
| INDIA 8590 | Unknown | chr6 | 76417342 | IGR | SBS | C | G |
| INDIA 8590 | Unknown | chr7 | 68958134 | IGR | DEL | C | - |
| INDIA 8590 | Unknown | chrX | 78409524 | IGR | SBS | C | G |
| INDIA 8590 | RAD54B | chr8 | 94378934 | Intron | SBS | C | G |
| INDIA 8590 | SLC7A6 | chr16 | 68264162 | 5'Flank | DEL | AGGGGGCGC | - |
| INDIA 8590 | LINC01138 | chr1 | 148350700 | Intron | DEL | G | - |
| INDIA 8590 | PRKN | chr6 | 162495562 | Intron | SBS | G | C |
| INDIA 8590 | Unknown | chr7 | 54006729 | IGR | SBS | A | C |
| INDIA 8590 | ENST00000655342 | chr13 | 39107827 | Intron | SBS | T | G |
| INDIA 8590 | Unknown | chr14 | 54915014 | IGR | SBS | A | C |
| INDIA 8590 | Unknown | chrX | 137280800 | IGR | SBS | G | T |
| INDIA 8590 | Unknown | chrX | 9958665 | IGR | SBS | A | C |
| INDIA 8590 | Unknown | chr11 | 87425879 | IGR | SBS | G | T |
| INDIA 8590 | Unknown | chr5 | 3058819 | IGR | SBS | A | T |
| INDIA 8590 | RBL1 | chr20 | 37007400 | Intron | DEL | AT | - |
| INDIA 8590 | Unknown | chrX | 109274442 | IGR | SBS | T | A |
| INDIA 8590 | MPV17L2 | chr19 | 18193024 | 5'Flank | DEL | AGGTC | - |
| INDIA 8590 | Unknown | chr1 | 198995881 | IGR | SBS | T | A |
| INDIA 8590 | TRPC6 | chr11 | 101468708 | Intron | SBS | A | T |
| INDIA 8590 | MB21D2 | chr3 | 192802967 | Intron | SBS | T | G |
| INDIA 8590 | CFAP47 | chrX | 36100894 | Intron | SBS | T | A |
| INDIA 8590 | Unknown | chrX | 148437946 | IGR | SBS | A | C |
| INDIA 8590 | MEIS1 | chr2 | 66510282 | Intron | DEL | T | - |
| INDIA 8590 | CCDC141 | chr2 | 178961764 | Intron | SBS | T | A |
| INDIA 8590 | KIAA1210 | chrX | 119144435 | Intron | SBS | G | C |
| INDIA 8590 | TRABD2B | chr1 | 47793099 | Intron | SBS | T | A |
| INDIA 8590 | FOXG1-AS1 | chr14 | 28737718 | Intron | SBS | C | A |
| INDIA 8590 | LINGO1 | chr15 | 77732503 | Intron | DEL | G | - |
| INDIA 8590 | FTO | chr16 | 53979990 | Intron | SBS | C | G |
| INDIA 8590 | TTN | chr2 | 178728728 | Missense_Mutation | SBS | C | G |
| INDIA 8590 | VEPH1 | chr3 | 157464333 | Intron | SBS | C | A |
| INDIA 8590 | Unknown | chr3 | 135569468 | IGR | SBS | G | C |
| INDIA 8590 | Unknown | chr5 | 160507432 | IGR | SBS | G | C |
| INDIA 8590 | MIR3681HG | chr2 | 12451986 | Intron | DEL | G | - |
| INDIA 8590 | CDKN2AIP | chr4 | 183446604 | Missense_Mutation | SBS | C | G |
| INDIA 8590 | Unknown | chr10 | 76758712 | IGR | SBS | G | C |
| INDIA 8590 | GABRA2 | chr4 | 46379671 | Intron | SBS | G | C |
| INDIA 8590 | THBS4 | chr5 | 80012239 | Intron | SBS | C | G |

| | | | | | | | |
|---|---|---|---|---|---|---|---|
| INDIA 8590 | UNC5D | chr8 | 35413718 | Intron | SBS | G | C |
| INDIA 8590 | FAH | chr15 | 80188282 | 3'Flank | DEL | GGAAATG | - |
| INDIA 8590 | BNC2 | chr9 | 16664198 | Intron | DEL | G | - |
| INDIA 8590 | ENST00000665445 | chr2 | 76632793 | 3'Flank | SBS | C | G |
| INDIA 8590 | Unknown | chr2 | 154925668 | IGR | SBS | A | T |
| INDIA 8590 | ENST00000639259 | chr2 | 210179945 | Intron | SBS | G | C |
| INDIA 8590 | LINC01017 | chr5 | 3503300 | Intron | SBS | C | A |
| INDIA 8590 | Unknown | chr7 | 135343992 | IGR | SBS | C | G |
| INDIA 8590 | DACH1 | chr13 | 71864383 | Intron | DEL | AGA | - |
| INDIA 8590 | PTPRS | chr19 | 5261532 | Intron | SBS | G | C |
| INDIA 8590 | LINC02208 | chr5 | 118504282 | Intron | DEL | G | - |
| INDIA 8590 | CACNA1A | chr19 | 13244761 | Intron | SBS | C | G |
| INDIA 8590 | TMEM131 | chr2 | 97789740 | Intron | SBS | C | G |
| INDIA 8590 | MRAP2 | chr6 | 84060052 | Intron | SBS | G | C |
| INDIA 8590 | Unknown | chr11 | 114915811 | IGR | DEL | A | - |
| INDIA 8591 | CTNNA2 | chr2 | 79248782 | Intron | SBS | C | G |
| INDIA 8591 | LINC02011 | chr3 | 14802576 | RNA | DEL | G | - |
| INDIA 8591 | LINC01885 | chr2 | 107540634 | Intron | SBS | C | G |
| INDIA 8591 | GRM7 | chr3 | 7463512 | Intron | SBS | G | C |
| INDIA 8591 | ENST00000522815 | chr8 | 125479406 | Intron | SBS | G | T |
| INDIA 8591 | ENST00000637529 | chr12 | 3342494 | Intron | DEL | C | - |
| INDIA 8591 | ENST00000484092 | chr3 | 117857444 | Intron | SBS | T | G |
| INDIA 8591 | CDH18 | chr5 | 20118843 | Intron | SBS | G | C |
| INDIA 8591 | LGR4-AS1 | chr11 | 27486485 | 3'Flank | SBS | G | C |
| INDIA 8591 | LINC01655 | chr1 | 221825359 | Intron | DEL | T | - |
| INDIA 8591 | CDH10 | chr5 | 24623161 | Intron | SBS | C | G |
| INDIA 8591 | Unknown | chr13 | 23001738 | IGR | SBS | A | C |
| INDIA 8591 | WWTR1 | chr3 | 149656107 | Intron | SBS | G | C |
| INDIA 8591 | SMARCC1 | chr3 | 47689359 | Intron | SBS | C | G |
| INDIA 8591 | ABCC4 | chr13 | 95206486 | Intron | SBS | C | G |
| INDIA 8591 | PDE4B | chr1 | 66163898 | Intron | SBS | T | A |
| INDIA 8591 | EXT1 | chr8 | 118052022 | Intron | SBS | G | C |
| INDIA 8591 | Unknown | chr11 | 8336446 | IGR | SBS | G | C |
| INDIA 8591 | RIMS1 | chr6 | 72300222 | Intron | SBS | G | C |
| INDIA 8591 | Unknown | chr10 | 61628181 | IGR | SBS | G | C |
| INDIA 8591 | Unknown | chr11 | 11629557 | IGR | SBS | A | T |
| INDIA 8591 | NRXN3 | chr14 | 79337909 | Intron | SBS | C | G |
| INDIA 8591 | LINC01320 | chr2 | 33824353 | Intron | SBS | G | C |
| INDIA 8591 | LINC02506 | chr4 | 32147861 | Intron | DEL | G | - |
| INDIA 8591 | Unknown | chr3 | 168223864 | IGR | DEL | G | - |
| INDIA 8591 | GALNTL6 | chr4 | 172548457 | Intron | SBS | T | A |
| INDIA 8591 | TAAR5 | chr6 | 132594356 | 5'Flank | SBS | A | T |
| INDIA 8591 | Unknown | chr18 | 70811069 | IGR | SBS | C | G |
| INDIA 8591 | MEF2C | chr5 | 88822808 | Intron | SBS | A | T |
| INDIA 8591 | Unknown | chr6 | 82334175 | IGR | DEL | A | - |
| INDIA 8591 | MIDEAS | chr14 | 73762559 | 5'Flank | SBS | C | G |
| INDIA 8591 | L3MBTL4 | chr18 | 6241789 | Intron | INS | - | C |
| INDIA 8591 | KIF15 | chr3 | 44830134 | Intron | SBS | C | G |
| INDIA 8591 | Unknown | chr7 | 9859322 | IGR | SBS | C | G |
| INDIA 8591 | Unknown | chr15 | 40121989 | IGR | SBS | G | C |
| INDIA 8591 | CETN1 | chr18 | 577744 | 5'Flank | SBS | A | T |
| INDIA 8591 | SETBP1 | chr18 | 44708096 | Intron | SBS | G | C |
| INDIA 8591 | Unknown | chr17 | 70751536 | IGR | SBS | T | A |
| INDIA 8591 | SIRPD | chr20 | 1554232 | Intron | SBS | C | G |
| INDIA 8591 | NTN1 | chr17 | 9176357 | Intron | DEL | GA | - |
| INDIA 8591 | RBFOX1 | chr16 | 7573386 | Intron | SBS | C | A |
| INDIA 8591 | PPP1R17 | chr7 | 31686301 | 5'Flank | SBS | C | A |
| INDIA 8591 | ENST00000435539 | chr10 | 2074985 | Intron | SBS | T | A |
| INDIA 8591 | WWTR1 | chr3 | 149563570 | Intron | SBS | A | C |
| INDIA 8591 | SNF8 | chr17 | 48940402 | Intron | SBS | C | G |
| INDIA 8591 | EPDR1 | chr7 | 37944384 | Intron | SBS | C | G |
| INDIA 8591 | CAPN15 | chr16 | 544085 | Intron | SBS | A | T |
| INDIA 8591 | UBE2E2 | chr3 | 23525698 | Intron | SBS | A | C |
| INDIA 8591 | LRMDA | chr10 | 76549468 | Intron | SBS | C | G |
| INDIA 8591 | Unknown | chr13 | 106841601 | IGR | SBS | C | A |
| INDIA 8591 | RBFOX1 | chr16 | 7522205 | Intron | DEL | G | - |

| | | | | | | | |
|---|---|---|---|---|---|---|---|
| INDIA 8591 | TP53 | chr17 | 7675136 | Frame_Shift_Del | DEL | GC | - |
| INDIA 8591 | LINC01549 | chr21 | 17435460 | 5'Flank | SBS | C | G |
| INDIA 8591 | IL36A | chr2 | 113005427 | 5'Flank | SBS | G | C |
| INDIA 8591 | EPHB1 | chr3 | 135162719 | Intron | SBS | C | G |
| INDIA 8591 | DISC1FP1 | chr11 | 90523688 | Intron | SBS | T | G |
| INDIA 8591 | CTNNA2 | chr2 | 79872834 | Intron | SBS | T | A |
| INDIA 8591 | GSTM3 | chr1 | 109732988 | 3'Flank | SBS | G | C |
| INDIA 8591 | Unknown | chr13 | 88826264 | IGR | SBS | C | G |
| INDIA 8591 | PLEKHA6 | chr1 | 204285999 | Intron | SBS | G | C |
| INDIA 8591 | PRIMA1 | chr14 | 93766998 | Intron | SBS | C | G |
| INDIA 8591 | Unknown | chr20 | 48539065 | IGR | SBS | G | C |
| INDIA 8591 | ENST00000575911 | chr17 | 1242188 | RNA | SBS | C | G |
| INDIA 8591 | MIR3681HG | chr2 | 12037389 | Intron | SBS | C | G |
| INDIA 8591 | KCNK12 | chr2 | 47504741 | 3'Flank | SBS | G | T |
| INDIA 8591 | LINC01800 | chr2 | 64845468 | 3'Flank | SBS | C | G |
| INDIA 8591 | ENST00000664181 | chr3 | 188143035 | Intron | SBS | C | G |
| INDIA 8591 | ENST00000509641 | chr4 | 57598956 | Intron | SBS | A | T |
| INDIA 8591 | Unknown | chr5 | 89831516 | IGR | SBS | C | G |
| INDIA 8591 | ENST00000659832 | chr5 | 145228096 | Intron | SBS | C | G |
| INDIA 8591 | FARS2 | chr6 | 5322764 | Intron | SBS | C | G |
| INDIA 8591 | NEAT1 | chr11 | 65430302 | RNA | SBS | G | T |
| INDIA 8591 | TTI1 | chr20 | 38004231 | Intron | SBS | G | C |
| INDIA 8591 | NKAIN4 | chr20 | 63236630 | 3'Flank | SBS | G | T |
| INDIA 8591 | LINC02792 | chr1 | 79342992 | Intron | SBS | C | G |
| INDIA 8591 | NKAIN2 | chr6 | 124517525 | Intron | SBS | T | G |
| INDIA 8591 | AMPH | chr7 | 38622554 | Intron | SBS | C | A |
| INDIA 8591 | BICD1 | chr12 | 32139242 | Intron | SBS | C | G |
| INDIA 8591 | MFN2 | chr1 | 11996666 | Intron | SBS | C | G |
| INDIA 8591 | MAP3K11 | chr11 | 65600943 | Intron | SBS | C | G |
| INDIA 8591 | Unknown | chr10 | 30211678 | IGR | SBS | C | G |
| INDIA 8591 | AEBP2 | chr12 | 19670031 | Intron | SBS | C | G |
| INDIA 8591 | Unknown | chr20 | 41917791 | IGR | SBS | C | G |
| INDIA 8591 | Unknown | chr2 | 22560217 | IGR | SBS | A | T |
| INDIA 8591 | KALRN | chr3 | 124194738 | Intron | SBS | C | A |
| INDIA 8591 | PLCL1 | chr2 | 198168755 | Intron | SBS | T | A |
| INDIA 8591 | ZER1 | chr9 | 128733700 | Intron | SBS | T | A |
| INDIA 8591 | TENM3 | chr4 | 182199078 | Intron | SBS | A | C |
| INDIA 8591 | TMEM167A | chr5 | 83051784 | 3'Flank | SBS | C | G |
| INDIA 8591 | LINC02200 | chr5 | 112627783 | 5'Flank | SBS | A | C |
| INDIA 8591 | WAKMAR2 | chr6 | 137850636 | Intron | SBS | G | C |
| INDIA 8591 | Unknown | chr11 | 79809890 | IGR | SBS | G | C |
| INDIA 8591 | Unknown | chr13 | 72665248 | IGR | SBS | G | C |
| INDIA 8592 | Unknown | chrX | 99974134 | IGR | SBS | T | G |
| INDIA 8592 | ENST00000617436 | chr9 | 89664985 | RNA | SBS | G | C |
| INDIA 8592 | Unknown | chr9 | 71632312 | IGR | SBS | G | C |
| INDIA 8592 | MID1 | chrX | 10620506 | 5'UTR | SBS | C | G |
| INDIA 8592 | Unknown | chrX | 103297380 | IGR | SBS | G | T |
| INDIA 8592 | LINC02196 | chr5 | 7181615 | 3'Flank | SBS | A | C |
| INDIA 8592 | TUSC3 | chr8 | 15491819 | Intron | SBS | A | C |
| INDIA 8592 | DNAH2 | chr17 | 7717617 | 5'Flank | SBS | G | C |
| INDIA 8592 | ROPN1L | chr5 | 10453639 | Intron | SBS | C | G |
| INDIA 8592 | Unknown | chr5 | 10145479 | IGR | SBS | G | C |
| INDIA 8592 | ENST00000507435 | chr5 | 4662155 | Intron | SBS | C | G |
| INDIA 8592 | Unknown | chr5 | 31614750 | IGR | INS | - | CCACTAACTG |
| INDIA 8592 | Unknown | chrX | 145863243 | IGR | SBS | C | G |
| INDIA 8592 | RGL1 | chr1 | 183881799 | Intron | SBS | C | G |
| INDIA 8592 | TJP2 | chr9 | 69248739 | Intron | SBS | A | T |
| INDIA 8592 | ENOX1 | chr13 | 43557056 | Intron | SBS | T | G |
| INDIA 8592 | Unknown | chr1 | 153291827 | IGR | DEL | C | - |
| INDIA 8592 | RGS9 | chr17 | 65147414 | Intron | SBS | C | G |
| INDIA 8592 | Unknown | chr17 | 77847760 | IGR | SBS | C | G |
| INDIA 8592 | Unknown | chr5 | 3200536 | IGR | SBS | A | T |
| INDIA 8592 | Unknown | chr2 | 208041986 | IGR | SBS | G | C |
| INDIA 8592 | SNORD116-14 | chr15 | 25076707 | 5'Flank | SBS | G | C |
| INDIA 8592 | FSHR | chr2 | 49089874 | Intron | SBS | C | G |
| INDIA 8592 | TOR3A | chr1 | 179096214 | 3'Flank | SBS | C | G |

| | | | | | | | |
|---|---|---|---|---|---|---|---|
| INDIA 8592 | Unknown | chr17 | 83115046 | IGR | SBS | C | A |
| INDIA 8592 | SORBS1 | chr10 | 95361932 | Intron | SBS | C | G |
| INDIA 8592 | Unknown | chrX | 110906368 | IGR | SBS | C | A |
| INDIA 8592 | FSD1L | chr9 | 105532796 | Intron | SBS | G | C |
| INDIA 8592 | Unknown | chr2 | 118678115 | IGR | SBS | C | G |
| INDIA 8592 | AGBL1 | chr15 | 86864072 | Intron | SBS | C | G |
| INDIA 8592 | TMEM94 | chr17 | 75471360 | Intron | SBS | T | G |
| INDIA 8592 | Unknown | chr9 | 78807985 | IGR | SBS | A | T |
| INDIA 8592 | PIAS1 | chr15 | 68193929 | 3'Flank | SBS | C | G |
| INDIA 8592 | Unknown | chr17 | 55233840 | IGR | SBS | C | G |
| INDIA 8592 | DDR2 | chr1 | 162728783 | Intron | SBS | G | C |
| INDIA 8592 | ARHGAP29-AS1 | chr1 | 94340539 | Intron | SBS | G | T |
| INDIA 8592 | WDR47 | chr1 | 109043183 | 5'Flank | SBS | G | C |
| INDIA 8592 | ENST00000697639 | chr9 | 118278553 | Intron | SBS | A | T |
| INDIA 8592 | PTH2R | chr2 | 208406761 | 5'Flank | DEL | CGCAGGCGC | - |
| INDIA 8592 | Unknown | chr2 | 143838607 | IGR | SBS | T | A |
| INDIA 8592 | PKN2-AS1 | chr1 | 88006988 | Intron | SBS | G | C |
| INDIA 8592 | TMEM132D | chr12 | 129135108 | Intron | SBS | G | T |
| INDIA 8592 | SLC2A1 | chr1 | 42944583 | Intron | SBS | T | A |
| INDIA 8592 | Unknown | chr1 | 117249688 | IGR | SBS | G | T |
| INDIA 8592 | ACYP2 | chr2 | 54021949 | Intron | SBS | A | T |
| INDIA 8592 | Unknown | chr2 | 82116973 | IGR | SBS | C | G |
| INDIA 8592 | LINC01824 | chr2 | 6506702 | Intron | DEL | T | - |
| INDIA 8592 | CDH18 | chr5 | 19543705 | Intron | SBS | A | T |
| INDIA 8592 | AGBL1 | chr15 | 86546756 | Intron | SBS | G | C |
| INDIA 8592 | LINC01532 | chr19 | 29014714 | RNA | SBS | C | G |
| INDIA 8592 | PDE4D | chr5 | 59807036 | Intron | SBS | G | C |
| INDIA 8592 | ENST00000522661 | chr8 | 8564597 | Intron | SBS | A | T |
| INDIA 8592 | FYB2 | chr1 | 56821433 | 5'Flank | DEL | CC | - |
| INDIA 8592 | OR52B2 | chr11 | 6173841 | 5'Flank | SBS | T | G |
| INDIA 8592 | THSD7B | chr2 | 137003496 | Intron | SBS | G | C |
| INDIA 8592 | CALHM2 | chr10 | 103451130 | 5'UTR | SBS | G | T |
| INDIA 8592 | LYPD6B | chr2 | 149081084 | Intron | SBS | A | C |
| INDIA 8592 | PYY | chr17 | 43977097 | Intron | SBS | G | C |
| INDIA 8592 | Unknown | chr2 | 69280336 | IGR | SBS | G | C |
| INDIA 8592 | RBFOX1 | chr16 | 5887336 | Intron | DEL | C | - |
| INDIA 8592 | ARID1A | chr1 | 26779111 | Frame_Shift_Ins | INS | - | C |
| INDIA 8592 | IFI16 | chr1 | 159042093 | Intron | DEL | CCGGTACAG | - |
| INDIA 8592 | Unknown | chr4 | 116578220 | IGR | SBS | T | A |
| INDIA 8592 | YTHDF2 | chr1 | 28739762 | Intron | SBS | T | G |
| INDIA 8592 | Unknown | chr2 | 158945414 | IGR | SBS | G | C |
| INDIA 8592 | LIPH | chr3 | 185544996 | Intron | SBS | A | T |
| INDIA 8592 | ENST00000606382 | chr2 | 59746910 | Intron | SBS | G | C |
| INDIA 8592 | LINC00989 | chr4 | 79624565 | 3'Flank | SBS | A | C |
| INDIA 8592 | PTGFRN | chr1 | 116911511 | Intron | SBS | G | C |
| INDIA 8592 | APOL5 | chr22 | 35713992 | 5'Flank | SBS | C | G |
| INDIA 8592 | LINC02490 | chr15 | 53103233 | Intron | SBS | T | A |
| INDIA 8592 | CD58 | chr1 | 116538869 | Intron | SBS | G | C |
| INDIA 8592 | Unknown | chr5 | 174546797 | IGR | SBS | C | G |
| INDIA 8592 | ENST00000423781 | chr7 | 46981464 | Intron | DEL | C | - |
| INDIA 8592 | SLC9A1 | chr1 | 27097580 | 3'Flank | SBS | T | A |
| INDIA 8592 | CTNNA2 | chr2 | 80193386 | Intron | SBS | T | A |
| INDIA 8592 | R3HDM2 | chr12 | 57280315 | Intron | SBS | T | G |
| INDIA 8592 | DPYSL5 | chr2 | 26850139 | Intron | SBS | G | C |
| INDIA 8592 | STON1 | chr2 | 48541980 | Intron | SBS | A | C |
| INDIA 8592 | SLC12A8 | chr3 | 125183752 | Intron | SBS | G | C |
| INDIA 8592 | CCDC171 | chr9 | 15916896 | Intron | SBS | C | G |
| INDIA 8592 | Unknown | chr7 | 155163495 | IGR | SBS | C | G |
| INDIA 8592 | EBF3 | chr10 | 129925931 | Intron | SBS | G | T |
| INDIA 8592 | HOXD9 | chr2 | 176121273 | 5'Flank | SBS | G | C |
| INDIA 8592 | TCF7L1 | chr2 | 85277535 | Intron | SBS | A | C |
| INDIA 8592 | SDR42E2 | chr16 | 22195334 | 3'Flank | SBS | C | G |
| INDIA 8592 | SOX10 | chr22 | 37981735 | Intron | SBS | G | T |
| INDIA 8592 | ENST00000641006 | chr1 | 9416387 | 3'Flank | SBS | C | G |
| INDIA 8592 | Unknown | chr2 | 116376354 | IGR | DEL | G | - |
| INDIA 8592 | PGM1 | chr1 | 63605882 | Intron | SBS | T | A |

| | | | | | | | |
|---|---|---|---|---|---|---|---|
| INDIA 8592 | Unknown | chr5 | 4059176 | IGR | SBS | C | G |
| INDIA 8592 | TRIP6 | chr7 | 100875681 | 3'Flank | SBS | G | C |
| INDIA 8592 | C5orf67 | chr5 | 56516552 | Intron | SBS | C | G |
| INDIA 8593 | PSMD7-DT | chr16 | 74007294 | Intron | SBS | G | C |
| INDIA 8593 | Unknown | chrX | 12736684 | IGR | SBS | G | C |
| INDIA 8593 | TMEM255A | chrX | 120310521 | Intron | SBS | C | G |
| INDIA 8593 | Unknown | chrX | 149567201 | IGR | SBS | G | C |
| INDIA 8593 | LINC01696 | chr1 | 209333127 | 3'Flank | SBS | G | C |
| INDIA 8593 | ENST00000663262 | chr5 | 173381048 | 5'Flank | SBS | C | G |
| INDIA 8593 | Unknown | chr3 | 96804752 | IGR | SBS | C | G |
| INDIA 8593 | NSFL1C | chr20 | 1455966 | Intron | SBS | G | C |
| INDIA 8593 | IGLV7-46 | chr22 | 22367915 | 5'Flank | SBS | G | C |
| INDIA 8593 | VGLL1 | chrX | 136532449 | Intron | SBS | C | G |
| INDIA 8593 | MGAT5 | chr2 | 134425632 | Intron | SBS | G | C |
| INDIA 8593 | ANK3 | chr10 | 60517808 | Intron | SBS | C | G |
| INDIA 8593 | KALRN | chr3 | 124472184 | Intron | SBS | C | G |
| INDIA 8593 | LRP6 | chr12 | 12250230 | Intron | SBS | C | G |
| INDIA 8593 | Unknown | chr4 | 130342651 | IGR | SBS | G | C |
| INDIA 8593 | PKD1L1 | chr7 | 47826677 | Intron | SBS | G | C |
| INDIA 8593 | Unknown | chr20 | 22861736 | IGR | SBS | G | C |
| INDIA 8593 | Unknown | chr20 | 43283912 | IGR | SBS | G | C |
| INDIA 8593 | Unknown | chr1 | 21491871 | IGR | SBS | C | G |
| INDIA 8593 | C1orf112 | chr1 | 169848464 | Intron | SBS | G | C |
| INDIA 8593 | Unknown | chr2 | 150785084 | IGR | SBS | C | G |
| INDIA 8593 | PCDH10-DT | chr4 | 133072323 | 3'Flank | SBS | G | C |
| INDIA 8593 | DMGDH | chr5 | 79033456 | Intron | SBS | C | G |
| INDIA 8593 | SAMD12-AS1 | chr8 | 118782020 | Intron | SBS | C | G |
| INDIA 8593 | RRP12 | chr10 | 97355975 | 3'Flank | SBS | G | C |
| INDIA 8593 | VTI1A | chr10 | 112626117 | Intron | SBS | C | G |
| INDIA 8593 | ENST00000530492 | chr11 | 13882597 | 5'Flank | SBS | G | C |
| INDIA 8593 | Unknown | chr11 | 42555646 | IGR | SBS | G | C |
| INDIA 8593 | SMARCD1 | chr12 | 50098957 | Missense_Mutation | SBS | G | C |
| INDIA 8593 | PITPNM2 | chr12 | 123050775 | Intron | SBS | C | G |
| INDIA 8593 | CMIP | chr16 | 81607298 | Intron | SBS | C | G |
| INDIA 8593 | LINC01917 | chr18 | 53563447 | Intron | SBS | C | G |
| INDIA 8593 | HNF4A | chr20 | 44362097 | Intron | SBS | G | C |
| INDIA 8593 | Unknown | chrX | 93926681 | IGR | SBS | G | C |
| INDIA 8593 | GRM8 | chr7 | 126981185 | Intron | DEL | T | - |
| INDIA 8593 | ENST00000612444 | chr20 | 32350887 | 5'Flank | SBS | C | G |
| INDIA 8593 | TNIP3 | chr4 | 121231544 | 5'Flank | SBS | C | G |
| INDIA 8593 | CTNND2 | chr5 | 11902309 | Intron | SBS | G | C |
| INDIA 8593 | Unknown | chr20 | 59382931 | IGR | SBS | C | G |
| INDIA 8593 | PRR14L | chr22 | 31682418 | 3'UTR | SBS | C | G |
| INDIA 8593 | FZD4-DT | chr11 | 86994282 | Intron | SBS | G | C |
| INDIA 8593 | MED12L | chr3 | 151423304 | Intron | SBS | G | C |
| INDIA 8593 | Unknown | chr10 | 44472688 | IGR | SBS | G | C |
| INDIA 8593 | PWRN1 | chr15 | 24612057 | Intron | SBS | G | C |
| INDIA 8593 | RBFOX1 | chr16 | 7302698 | Intron | SBS | C | G |
| INDIA 8593 | Unknown | chr12 | 33673378 | IGR | SBS | C | G |
| INDIA 8593 | TAFA1 | chr3 | 68537152 | Intron | SBS | C | G |
| INDIA 8593 | DLGAP4 | chr20 | 36496780 | Nonsense_Mutation | SBS | C | G |
| INDIA 8593 | Unknown | chr4 | 163421777 | IGR | SBS | G | C |
| INDIA 8593 | ENST00000664438 | chr9 | 13673657 | Intron | SBS | G | C |
| INDIA 8593 | ENST00000659397 | chr9 | 89869979 | Intron | SBS | G | C |
| INDIA 8593 | Unknown | chrX | 49675635 | IGR | SBS | G | C |
| INDIA 8593 | NISCH | chr3 | 52481143 | Intron | SBS | G | C |
| INDIA 8593 | Unknown | chr10 | 124262178 | IGR | SBS | C | G |
| INDIA 8593 | TPRG1 | chr3 | 189202067 | Intron | SBS | G | C |
| INDIA 8593 | ARHGAP24 | chr4 | 85607860 | Intron | SBS | G | C |
| INDIA 8593 | LINC00640 | chr14 | 51333977 | Intron | SBS | G | C |
| INDIA 8593 | ZFHX3 | chr16 | 73711851 | Intron | SBS | C | G |
| INDIA 8593 | SMIM21 | chr18 | 75428456 | 5'Flank | SBS | G | C |
| INDIA 8593 | Unknown | chr11 | 19314751 | IGR | SBS | G | C |
| INDIA 8593 | SAE1 | chr19 | 47172272 | Intron | SBS | G | C |
| INDIA 8593 | PROKR2 | chr20 | 5321311 | 5'Flank | SBS | G | C |
| INDIA 8593 | DLGAP4 | chr20 | 36436476 | Intron | SBS | G | C |

| | | | | | | | |
|---|---|---|---|---|---|---|---|
| INDIA 8593 | KIF13A | chr6 | 17985075 | Intron | SBS | G | C |
| INDIA 8593 | Unknown | chr10 | 78084903 | IGR | SBS | G | C |
| INDIA 8593 | Unknown | chr10 | 100627822 | IGR | SBS | C | G |
| INDIA 8593 | VSNL1 | chr2 | 17611637 | Intron | SBS | C | G |
| INDIA 8593 | Unknown | chr4 | 10998850 | IGR | SBS | G | C |
| INDIA 8593 | Unknown | chr4 | 129357247 | IGR | SBS | G | C |
| INDIA 8593 | EIF4A3 | chr17 | 80151516 | 5'Flank | SBS | G | C |
| INDIA 8593 | NOS1AP | chr1 | 162137417 | Intron | SBS | C | G |
| INDIA 8593 | PCDH7 | chr4 | 30790178 | Intron | SBS | G | C |
| INDIA 8593 | Unknown | chr5 | 155339800 | IGR | SBS | C | G |
| INDIA 8593 | Unknown | chr20 | 51992585 | IGR | SBS | G | C |
| INDIA 8593 | TBL1X | chrX | 9647857 | Intron | SBS | G | C |
| INDIA 8593 | TENM1 | chrX | 124530933 | Intron | SBS | G | C |
| INDIA 8593 | CUX1 | chr7 | 101947873 | Intron | SBS | C | G |
| INDIA 8593 | Unknown | chr2 | 104553000 | IGR | SBS | C | G |
| INDIA 8593 | ENST00000667725 | chr2 | 172283988 | Intron | SBS | C | G |
| INDIA 8593 | RNU1-55P | chr20 | 5888196 | 5'Flank | SBS | C | G |
| INDIA 8593 | Unknown | chr20 | 37636627 | IGR | SBS | C | G |
| INDIA 8593 | Unknown | chr3 | 28167955 | IGR | SBS | C | G |
| INDIA 8593 | Unknown | chr9 | 81832873 | IGR | SBS | C | G |
| INDIA 8593 | ENST00000668454 | chr5 | 88116533 | Intron | SBS | C | G |
| INDIA 8593 | Unknown | chr10 | 88238966 | IGR | SBS | C | G |
| INDIA 8593 | Unknown | chr10 | 98300849 | IGR | SBS | C | G |
| INDIA 8593 | Unknown | chr17 | 13489007 | IGR | SBS | C | G |
| INDIA 8593 | Unknown | chr13 | 64203328 | IGR | SBS | G | C |
| INDIA 8593 | HS6ST2 | chrX | 132836482 | Intron | SBS | G | C |
| INDIA 8593 | GPR50 | chrX | 151175491 | 5'Flank | SBS | C | G |
| INDIA 8593 | ZNF593 | chr1 | 26165198 | 5'Flank | SBS | G | C |
| INDIA 8593 | KCNT2 | chr1 | 196347112 | Intron | SBS | G | C |
| INDIA 8593 | ENST00000523264 | chr8 | 110608956 | 5'Flank | SBS | C | G |
| INDIA 8593 | LINC00692 | chr3 | 25874303 | 5'Flank | SBS | C | G |
| INDIA 8593 | SLC2A9 | chr4 | 9910792 | Intron | SBS | G | C |
| INDIA 8593 | ENST00000656970 | chr8 | 20672340 | Intron | SBS | G | C |
| INDIA 8594 | ENST00000667181 | chr1 | 230898551 | 3'Flank | SBS | C | A |
| INDIA 8594 | ANKS6 | chr9 | 98731013 | 3'Flank | SBS | C | G |
| INDIA 8594 | DOCK3 | chr3 | 50950765 | Intron | SBS | C | G |
| INDIA 8594 | Unknown | chr11 | 127490862 | IGR | SBS | G | C |
| INDIA 8594 | GGTA1 | chr9 | 121471739 | Intron | SBS | C | G |
| INDIA 8594 | RERGL | chr12 | 18124067 | Intron | SBS | G | C |
| INDIA 8594 | NFIA | chr1 | 61292685 | Intron | SBS | G | C |
| INDIA 8594 | Unknown | chr11 | 87357683 | IGR | SBS | C | G |
| INDIA 8594 | Unknown | chr10 | 111780062 | IGR | SBS | G | C |
| INDIA 8594 | Unknown | chr12 | 27852371 | IGR | SBS | C | G |
| INDIA 8594 | NTNG2 | chr9 | 132245462 | 3'Flank | SBS | C | G |
| INDIA 8594 | DISC1 | chr1 | 231691116 | Intron | SBS | G | C |
| INDIA 8594 | ENST00000438371 | chr1 | 235940657 | 3'Flank | SBS | C | G |
| INDIA 8594 | Unknown | chr21 | 41046127 | IGR | SBS | G | C |
| INDIA 8594 | SHROOM2 | chrX | 9903427 | Intron | SBS | G | C |
| INDIA 8594 | CSNK2A2 | chr16 | 58159638 | Intron | SBS | C | G |
| INDIA 8594 | RIPK4 | chr21 | 41753489 | Intron | SBS | C | G |
| INDIA 8594 | Unknown | chr17 | 12534644 | IGR | SBS | G | C |
| INDIA 8594 | GPR137B | chr1 | 236176409 | Intron | SBS | C | G |
| INDIA 8594 | GIGYF2 | chr2 | 232860674 | 3'Flank | SBS | C | G |
| INDIA 8594 | ENST00000624331 | chr8 | 19275576 | 5'Flank | SBS | G | T |
| INDIA 8594 | NFRKB | chr11 | 129885639 | Intron | SBS | C | G |
| INDIA 8594 | ENST00000669454 | chr15 | 46351256 | Intron | SBS | C | G |
| INDIA 8594 | LINC00670 | chr17 | 12581333 | Intron | SBS | G | C |
| INDIA 8594 | CD48 | chr1 | 160707647 | Intron | SBS | G | C |
| INDIA 8594 | PCNX2 | chr1 | 233060010 | Intron | SBS | G | C |
| INDIA 8594 | Unknown | chr9 | 12319490 | IGR | SBS | G | C |
| INDIA 8594 | ETV6 | chr12 | 11838651 | Intron | SBS | C | G |
| INDIA 8594 | Unknown | chr14 | 49138850 | IGR | SBS | G | C |
| INDIA 8594 | S100B | chr21 | 46604943 | Intron | SBS | C | G |
| INDIA 8594 | MTMR7 | chr8 | 17371134 | Silent | SBS | C | G |
| INDIA 8594 | Unknown | chr9 | 26535402 | IGR | SBS | G | C |
| INDIA 8594 | Unknown | chrX | 13000738 | IGR | SBS | C | G |

| | | | | | | | |
|---|---|---|---|---|---|---|---|
| INDIA 8594 | PTPRG | chr3 | 61688285 | Intron | SBS | C | G |
| INDIA 8594 | HCN1 | chr5 | 45634463 | Intron | SBS | G | C |
| INDIA 8594 | CSMD1 | chr8 | 4087166 | Intron | SBS | C | G |
| INDIA 8594 | MITF | chr3 | 69896800 | Intron | SBS | G | C |
| INDIA 8594 | Unknown | chr6 | 90642634 | IGR | SBS | G | C |
| INDIA 8594 | ENST00000521143 | chr8 | 9347064 | 3'Flank | SBS | C | G |
| INDIA 8594 | SPIDR | chr8 | 47560527 | Intron | SBS | C | G |
| INDIA 8594 | Unknown | chr8 | 125636161 | IGR | SBS | C | G |
| INDIA 8594 | Unknown | chr9 | 108284255 | IGR | SBS | C | A |
| INDIA 8594 | EGLN3 | chr14 | 34133889 | Intron | SBS | C | G |
| INDIA 8594 | ARMCX6 | chrX | 101617058 | Intron | SBS | C | G |
| INDIA 8594 | ENST00000653000 | chr5 | 2032457 | Intron | SBS | C | G |
| INDIA 8594 | STK3 | chr8 | 98403877 | Intron | SBS | C | G |
| INDIA 8594 | ENST00000645181 | chr4 | 42238922 | Intron | SBS | G | C |
| INDIA 8594 | LINC02913 | chr9 | 129322893 | RNA | SBS | G | C |
| INDIA 8594 | ENST00000671048 | chr8 | 34199747 | Intron | SBS | C | G |
| INDIA 8594 | Unknown | chr8 | 48705230 | IGR | SBS | C | G |
| INDIA 8594 | USP40 | chr2 | 233546301 | Intron | SBS | G | C |
| INDIA 8594 | Unknown | chr4 | 27305395 | IGR | SBS | C | G |
| INDIA 8594 | CDX1 | chr5 | 150171046 | Intron | SBS | G | C |
| INDIA 8594 | Unknown | chr7 | 51950487 | IGR | SBS | G | C |
| INDIA 8594 | Unknown | chr8 | 5525695 | IGR | SBS | G | C |
| INDIA 8594 | UMOD | chr16 | 20346455 | Intron | SBS | G | C |
| INDIA 8594 | WDR25 | chr14 | 100396444 | Intron | SBS | C | G |
| INDIA 8594 | RXRA | chr9 | 134366859 | Intron | SBS | C | G |
| INDIA 8594 | ENST00000655054 | chr11 | 94281038 | 3'Flank | SBS | G | C |
| INDIA 8594 | NFIB-AS1 | chr9 | 14363305 | 3'Flank | SBS | C | G |
| INDIA 8594 | TRIO | chr5 | 14473118 | Intron | SBS | G | C |
| INDIA 8594 | Unknown | chr12 | 5198818 | IGR | SBS | C | G |
| INDIA 8594 | ZNF672 | chr1 | 248844548 | 5'UTR | SBS | G | C |
| INDIA 8594 | LINC02017 | chr3 | 55183991 | Intron | SBS | G | C |
| INDIA 8594 | ERC2 | chr3 | 55689187 | Intron | SBS | C | G |
| INDIA 8594 | ENST00000666856 | chr9 | 105237012 | Intron | SBS | C | G |
| INDIA 8594 | KLK1 | chr19 | 50828435 | 5'Flank | SBS | C | G |
| INDIA 8594 | Unknown | chr3 | 49347658 | IGR | SBS | G | C |
| INDIA 8594 | Unknown | chr5 | 166344958 | IGR | SBS | G | C |
| INDIA 8594 | HDAC9 | chr7 | 18506221 | Intron | SBS | C | G |
| INDIA 8594 | Unknown | chr7 | 85351043 | IGR | SBS | C | G |
| INDIA 8594 | VLDLR-AS1 | chr9 | 2503817 | Intron | SBS | G | C |
| INDIA 8594 | ENST00000664549 | chr10 | 7038035 | Intron | SBS | C | G |
| INDIA 8594 | LINC02706 | chr11 | 134713805 | RNA | SBS | C | G |
| INDIA 8594 | OBI1-AS1 | chr13 | 78459080 | Intron | SBS | C | G |
| INDIA 8594 | GPC5 | chr13 | 92746074 | Intron | SBS | C | G |
| INDIA 8594 | ENST00000520375 | chr8 | 23745180 | 5'Flank | SBS | C | G |
| INDIA 8594 | Unknown | chr1 | 5403615 | IGR | SBS | G | C |
| INDIA 8594 | TRNAU1AP | chr1 | 28564642 | Intron | SBS | G | C |
| INDIA 8594 | AOPEP | chr9 | 94981579 | Intron | SBS | C | G |
| INDIA 8594 | Unknown | chrX | 43981412 | IGR | SBS | C | G |
| INDIA 8594 | Unknown | chr11 | 130745261 | IGR | SBS | C | G |
| INDIA 8594 | Unknown | chr1 | 38821412 | IGR | SBS | G | C |
| INDIA 8594 | ENST00000646142 | chr1 | 41621380 | Intron | SBS | C | G |
| INDIA 8594 | Unknown | chr3 | 28089059 | IGR | SBS | G | C |
| INDIA 8594 | SH2D3C | chr9 | 127767603 | Intron | SBS | G | C |
| INDIA 8594 | Unknown | chrX | 16551386 | IGR | SBS | G | C |
| INDIA 8594 | ENST00000652827 | chr9 | 99776644 | Intron | DEL | TTG | - |
| INDIA 8594 | LINC01622 | chr6 | 967720 | Intron | SBS | G | C |
| INDIA 8594 | LRMDA | chr10 | 75661035 | Intron | SBS | C | G |
| INDIA 8594 | BLK | chr8 | 11563375 | Intron | SBS | C | G |
| INDIA 8594 | Unknown | chr2 | 222006691 | IGR | SBS | G | C |
| INDIA 8594 | Unknown | chr4 | 10186261 | IGR | SBS | C | G |
| INDIA 8594 | ATP6V0D2 | chr8 | 86125353 | Intron | SBS | G | C |
| INDIA 8595 | WDR7 | chr18 | 56844759 | Intron | SBS | G | C |
| INDIA 8595 | Unknown | chr18 | 61564598 | IGR | SBS | G | C |
| INDIA 8595 | ENST00000667627 | chr3 | 34148881 | 5'Flank | SBS | C | G |
| INDIA 8595 | Unknown | chrX | 127772675 | IGR | DEL | TC | - |
| INDIA 8595 | Unknown | chr5 | 60859496 | IGR | SBS | C | G |

| | | | | | | | |
|---|---|---|---|---|---|---|---|
| INDIA 8595 | Unknown | chr18 | 59945684 | IGR | SBS | A | C |
| INDIA 8595 | RFTN1 | chr3 | 16468453 | Intron | SBS | G | C |
| INDIA 8595 | Unknown | chr18 | 43635399 | IGR | SBS | T | A |
| INDIA 8595 | HACD1 | chr10 | 17598528 | Intron | DEL | AG | - |
| INDIA 8595 | CDH1 | chr16 | 68810211 | Frame_Shift_Del | DEL | TG | - |
| INDIA 8595 | PDE6A | chr5 | 149906187 | Intron | DEL | TA | - |
| INDIA 8595 | CACNB2 | chr10 | 18333314 | Intron | SBS | G | C |
| INDIA 8595 | ENST00000643167 | chr1 | 55719739 | Intron | SBS | C | G |
| INDIA 8595 | CASC6 | chr6 | 91683465 | Intron | SBS | C | G |
| INDIA 8595 | ENST00000564561 | chr16 | 76122774 | Intron | DEL | AG | - |
| INDIA 8595 | LINC01807 | chr2 | 228577896 | Intron | SBS | C | G |
| INDIA 8595 | EPM2A | chr6 | 145631506 | Intron | SBS | C | G |
| INDIA 8595 | Unknown | chr16 | 26205805 | IGR | SBS | A | C |
| INDIA 8595 | ADARB2 | chr10 | 1600326 | Intron | SBS | C | G |
| INDIA 8595 | LINC01646 | chr1 | 4583263 | Intron | SBS | G | C |
| INDIA 8595 | WWOX | chr16 | 79119471 | Intron | SBS | G | C |
| INDIA 8595 | MAP4 | chr3 | 47855443 | Intron | SBS | C | G |
| INDIA 8595 | PITX1-AS1 | chr5 | 135110904 | Intron | SBS | C | G |
| INDIA 8595 | Unknown | chr6 | 76133467 | IGR | INS | - | A |
| INDIA 8595 | MALT1 | chr18 | 58716915 | Intron | DEL | AA | - |
| INDIA 8595 | CARMIL2 | chr16 | 67649450 | Missense_Mutation | SBS | C | G |
| INDIA 8595 | RBMS3 | chr3 | 28757503 | RNA | SBS | G | C |
| INDIA 8595 | ENST00000653535 | chr20 | 56559411 | 3'Flank | SBS | C | G |
| INDIA 8595 | FTO | chr16 | 54061098 | Intron | SBS | G | C |
| INDIA 8595 | Unknown | chr5 | 149702968 | IGR | DEL | TAAC | - |
| INDIA 8595 | Unknown | chr10 | 69071814 | IGR | DEL | AA | - |
| INDIA 8595 | CAMTA1 | chr1 | 7257835 | Intron | SBS | G | C |
| INDIA 8595 | PDIA5 | chr3 | 123081835 | Intron | SBS | G | C |
| INDIA 8595 | COQ10B | chr2 | 197461263 | Intron | DEL | AG | - |
| INDIA 8595 | SPATA18 | chr4 | 52079767 | Silent | SBS | C | G |
| INDIA 8595 | Unknown | chr10 | 124376685 | IGR | SBS | G | C |
| INDIA 8595 | TCF7L1 | chr2 | 85289411 | Intron | DEL | AG | - |
| INDIA 8595 | SNRK | chr3 | 43320453 | Intron | SBS | C | G |
| INDIA 8595 | NPSR1 | chr7 | 34769810 | Intron | SBS | G | C |
| INDIA 8595 | DNAJC16 | chr1 | 15537379 | Intron | SBS | G | C |
| INDIA 8595 | SLC6A6 | chr3 | 14422753 | Intron | DEL | TG | - |
| INDIA 8595 | MYOF | chr10 | 93437544 | Intron | DEL | AAATC | - |
| INDIA 8595 | TIFAB | chr5 | 135452085 | Intron | SBS | C | G |
| INDIA 8595 | ZFPM2 | chr8 | 104780614 | Intron | SBS | G | C |
| INDIA 8595 | LRRC36 | chr16 | 67347518 | Frame_Shift_Del | DEL | GA | - |
| INDIA 8595 | Unknown | chr16 | 82373076 | IGR | SBS | C | G |
| INDIA 8595 | LINC02955 | chr12 | 22846297 | Intron | SBS | C | G |
| INDIA 8595 | GTSE1 | chr22 | 46308434 | Frame_Shift_Del | DEL | GA | - |
| INDIA 8595 | PCDH7 | chr4 | 30906624 | Intron | SBS | G | C |
| INDIA 8595 | SNCAIP | chr5 | 122420089 | Intron | SBS | G | C |
| INDIA 8595 | PER2 | chr2 | 238286903 | Intron | SBS | C | G |
| INDIA 8595 | TNRC6B | chr22 | 40302912 | Intron | SBS | G | C |
| INDIA 8595 | LINC01829 | chr2 | 67310735 | Intron | SBS | A | T |
| INDIA 8595 | GPR37 | chr7 | 124762883 | Intron | DEL | AC | - |
| INDIA 8595 | Unknown | chr10 | 121215146 | IGR | SBS | T | G |
| INDIA 8595 | AKAP13 | chr15 | 85704259 | Intron | INS | - | G |
| INDIA 8595 | PHF21B | chr22 | 44885081 | Intron | SBS | G | C |
| INDIA 8595 | EFNA5 | chr5 | 107606778 | Intron | DEL | CT | - |
| INDIA 8595 | Unknown | chr2 | 153142189 | IGR | SBS | G | C |
| INDIA 8595 | RHOBTB1 | chr10 | 60887570 | Intron | DEL | AT | - |
| INDIA 8595 | ROBO2 | chr3 | 76309142 | 5'Flank | DEL | CT | - |
| INDIA 8595 | SORBS1 | chr10 | 95535953 | Intron | SBS | A | C |
| INDIA 8595 | Unknown | chr22 | 26405585 | IGR | SBS | G | C |
| INDIA 8595 | MYH9 | chr22 | 36355091 | Intron | SBS | G | C |
| INDIA 8595 | Unknown | chr10 | 34102338 | IGR | DEL | GA | - |
| INDIA 8595 | ACTA2 | chr10 | 88932720 | 3'Flank | SBS | C | G |
| INDIA 8595 | Unknown | chr16 | 87028243 | IGR | SBS | G | C |
| INDIA 8595 | Unknown | chr18 | 27203009 | IGR | SBS | C | G |
| INDIA 8595 | Unknown | chr2 | 10473164 | IGR | SBS | G | C |
| INDIA 8595 | PACSIN2 | chr22 | 42884260 | Intron | SBS | G | C |
| INDIA 8595 | ADARB2 | chr10 | 1213774 | Intron | DEL | AGG | - |

| | | | | | | | |
|---|---|---|---|---|---|---|---|
| INDIA 8595 | DCAF6 | chr1 | 168008805 | Intron | DEL | AG | - |
| INDIA 8595 | Unknown | chr6 | 76355764 | IGR | DEL | AC | - |
| INDIA 8595 | LINC00624 | chr1 | 147385155 | Intron | DEL | CT | - |
| INDIA 8595 | CAMK1G | chr1 | 209590386 | Intron | DEL | GT | - |
| INDIA 8595 | LINC01885 | chr2 | 107505790 | Intron | DEL | TTCT | - |
| INDIA 8595 | P4HTM | chr3 | 49006537 | Intron | SBS | G | C |
| INDIA 8595 | LINC00882 | chr3 | 106728421 | Intron | SBS | G | C |
| INDIA 8595 | Unknown | chr3 | 177405060 | IGR | DEL | GA | - |
| INDIA 8595 | CCDC39 | chr3 | 180647978 | Intron | SBS | G | C |
| INDIA 8595 | UNC5C | chr4 | 95506332 | Intron | SBS | C | G |
| INDIA 8595 | ENST00000667497 | chr4 | 125854439 | Intron | DEL | CT | - |
| INDIA 8595 | FAM83B | chr6 | 54914665 | Intron | SBS | G | C |
| INDIA 8595 | BCKDHB | chr6 | 80332185 | Intron | SBS | G | C |
| INDIA 8595 | TENT5A | chr6 | 81744939 | 3'Flank | DEL | CA | - |
| INDIA 8595 | PRKN | chr6 | 161826620 | Intron | SBS | C | G |
| INDIA 8595 | Unknown | chr8 | 55860836 | IGR | SBS | G | C |
| INDIA 8595 | ZNF248 | chr10 | 37831725 | Frame_Shift_Del | DEL | CT | - |
| INDIA 8595 | DOCK1 | chr10 | 126998086 | Splice_Region | DEL | AC | - |
| INDIA 8595 | Unknown | chr11 | 20142624 | IGR | SBS | C | G |
| INDIA 8595 | Unknown | chr13 | 39962030 | IGR | SBS | C | G |
| INDIA 8595 | LINC01586 | chr15 | 88608224 | 5'Flank | SBS | G | C |
| INDIA 8595 | ENST00000564193 | chr16 | 47029383 | Intron | SBS | C | G |
| INDIA 8595 | Unknown | chr16 | 82263738 | IGR | SBS | G | C |
| INDIA 8596 | Unknown | chr9 | 13380200 | IGR | SBS | C | G |
| INDIA 8596 | Unknown | chr9 | 94683031 | IGR | SBS | C | G |
| INDIA 8596 | Unknown | chr8 | 25708093 | IGR | SBS | C | G |
| INDIA 8596 | STXBP6 | chr14 | 24955225 | Intron | SBS | C | G |
| INDIA 8596 | ADAMTSL1 | chr9 | 18817626 | Intron | SBS | G | C |
| INDIA 8596 | ENST00000687892 | chrX | 38951294 | Intron | SBS | C | G |
| INDIA 8596 | NFIB | chr9 | 14275208 | Intron | SBS | G | C |
| INDIA 8596 | OTOF | chr2 | 26514279 | Intron | SBS | C | G |
| INDIA 8596 | Unknown | chr9 | 16133345 | IGR | SBS | G | C |
| INDIA 8596 | ENAH | chr1 | 225506568 | Intron | SBS | G | C |
| INDIA 8596 | RNU6-205P | chr4 | 110275266 | 5'Flank | SBS | C | G |
| INDIA 8596 | Unknown | chr8 | 15305293 | IGR | SBS | G | C |
| INDIA 8596 | ADAMTSL1 | chr9 | 18800494 | Intron | SBS | G | C |
| INDIA 8596 | Unknown | chr9 | 90172312 | IGR | SBS | C | G |
| INDIA 8596 | ENST00000399342 | chr17 | 10384186 | Intron | SBS | G | C |
| INDIA 8596 | Unknown | chr13 | 51308802 | IGR | SBS | G | C |
| INDIA 8596 | ENST00000686463 | chr9 | 91073193 | 3'Flank | SBS | C | G |
| INDIA 8596 | Unknown | chr9 | 94680157 | IGR | SBS | C | G |
| INDIA 8596 | Unknown | chr9 | 20637854 | IGR | SBS | G | C |
| INDIA 8596 | RCSD1 | chr1 | 167702456 | Intron | SBS | C | G |
| INDIA 8596 | CLBA1 | chr14 | 104981073 | 5'Flank | SBS | C | G |
| INDIA 8596 | SLC24A2 | chr9 | 19579999 | Intron | SBS | C | G |
| INDIA 8596 | MFSD4A | chr1 | 205566919 | 5'Flank | SBS | G | C |
| INDIA 8596 | TG | chr8 | 132877829 | Intron | SBS | G | C |
| INDIA 8596 | Unknown | chr9 | 33189179 | IGR | SBS | G | C |
| INDIA 8596 | NXNL2 | chr9 | 88587434 | 3'Flank | SBS | C | G |
| INDIA 8596 | CACNA1E | chr1 | 181499260 | Intron | SBS | G | C |
| INDIA 8596 | TRPV2 | chr17 | 16427538 | Silent | SBS | C | A |
| INDIA 8596 | PNPLA1 | chr6 | 36290920 | Intron | SBS | C | G |
| INDIA 8596 | Unknown | chr9 | 117730008 | IGR | SBS | C | G |
| INDIA 8596 | Unknown | chr9 | 14075250 | IGR | SBS | G | C |
| INDIA 8596 | SPATA3 | chr2 | 231001932 | Intron | SBS | G | C |
| INDIA 8596 | CMIP | chr16 | 81639034 | Intron | SBS | G | C |
| INDIA 8596 | PIP5K1B | chr9 | 68832645 | Intron | SBS | C | G |
| INDIA 8596 | Unknown | chr12 | 13167166 | IGR | SBS | G | C |
| INDIA 8596 | Unknown | chr8 | 108112364 | IGR | SBS | C | G |
| INDIA 8596 | ENST00000652827 | chr9 | 99718243 | RNA | SBS | C | G |
| INDIA 8596 | GBGT1 | chr9 | 133153881 | Missense_Mutation | SBS | G | C |
| INDIA 8596 | ENST00000685728 | chrX | 146857060 | Intron | SBS | C | G |
| INDIA 8596 | FOXN2 | chr2 | 48332516 | Intron | SBS | C | G |
| INDIA 8596 | Unknown | chr2 | 42929965 | IGR | SBS | G | T |
| INDIA 8596 | RUBCN | chr3 | 197735546 | Intron | SBS | C | G |
| INDIA 8596 | DNAH14 | chr1 | 225389851 | Silent | SBS | C | G |

| | | | | | | | |
|---|---|---|---|---|---|---|---|
| INDIA 8596 | ENST00000664438 | chr9 | 13706007 | Intron | SBS | G | C |
| INDIA 8596 | MIRLET7F1 | chr9 | 94176348 | RNA | SBS | C | G |
| INDIA 8596 | Unknown | chr14 | 74635292 | IGR | SBS | G | C |
| INDIA 8596 | NTRK2 | chr9 | 84886730 | Intron | SBS | G | C |
| INDIA 8596 | ENST00000663507 | chr2 | 12983710 | Intron | SBS | C | G |
| INDIA 8596 | PTPRD | chr9 | 8403952 | Intron | SBS | C | G |
| INDIA 8596 | TTC6 | chr14 | 37820010 | Intron | SBS | C | G |
| INDIA 8596 | TMED6 | chr16 | 69343318 | 3'UTR | SBS | G | C |
| INDIA 8596 | ENST00000666628 | chrX | 36689321 | Intron | SBS | C | G |
| INDIA 8596 | TCF7L1 | chr2 | 85284544 | Intron | SBS | G | C |
| INDIA 8596 | SNTB1 | chr8 | 120578421 | Intron | SBS | C | G |
| INDIA 8596 | NSMCE2 | chr8 | 125253392 | Intron | SBS | G | C |
| INDIA 8596 | PTPRD | chr9 | 10612575 | Intron | SBS | G | C |
| INDIA 8596 | ETV7 | chr6 | 36386724 | Intron | SBS | C | G |
| INDIA 8596 | LINC01588 | chr14 | 49954759 | Intron | SBS | G | C |
| INDIA 8596 | FHOD3 | chr18 | 36721291 | Intron | SBS | G | C |
| INDIA 8596 | ENST00000419809 | chr2 | 67569998 | Intron | SBS | C | G |
| INDIA 8596 | ASAP2 | chr2 | 9212498 | Intron | SBS | C | G |
| INDIA 8596 | DPYSL5 | chr2 | 26919181 | Intron | SBS | G | C |
| INDIA 8596 | FANCC | chr9 | 95273172 | Intron | SBS | G | C |
| INDIA 8596 | Unknown | chr2 | 14599979 | IGR | SBS | C | G |
| INDIA 8596 | PARTICL | chr2 | 85533768 | 3'Flank | SBS | C | G |
| INDIA 8596 | Unknown | chr2 | 129163004 | IGR | SBS | G | C |
| INDIA 8596 | CDH12 | chr5 | 22057532 | Intron | SBS | G | C |
| INDIA 8596 | Unknown | chr6 | 1405157 | IGR | SBS | C | G |
| INDIA 8596 | Unknown | chr6 | 10318405 | IGR | DEL | C | - |
| INDIA 8596 | Unknown | chr6 | 27337140 | IGR | SBS | C | G |
| INDIA 8596 | STMN2 | chr8 | 79621757 | Intron | SBS | G | C |
| INDIA 8596 | AGO2 | chr8 | 140560781 | Intron | SBS | C | G |
| INDIA 8596 | GABBR2 | chr9 | 98611713 | Intron | SBS | C | G |
| INDIA 8596 | Unknown | chrX | 13252411 | IGR | SBS | G | C |
| INDIA 8596 | ANXA1 | chr9 | 73165194 | Missense_Mutation | SBS | C | G |
| INDIA 8596 | ZNF684 | chr1 | 40541948 | Intron | SBS | C | G |
| INDIA 8596 | TPPP | chr5 | 673078 | Intron | SBS | C | G |
| INDIA 8596 | Unknown | chr6 | 33960992 | IGR | SBS | G | C |
| INDIA 8596 | Unknown | chr9 | 83132952 | IGR | SBS | C | G |
| INDIA 8596 | SCARA5 | chr8 | 27890900 | Intron | SBS | G | C |
| INDIA 8596 | FUCA1 | chr1 | 23869019 | 5'Flank | SBS | G | C |
| INDIA 8596 | Unknown | chr2 | 382402 | IGR | SBS | G | C |
| INDIA 8596 | Unknown | chr8 | 37135635 | IGR | SBS | G | C |
| INDIA 8596 | ODF2 | chr9 | 128485108 | Intron | SBS | G | C |
| INDIA 8596 | MGST1 | chr12 | 16347277 | 5'Flank | DEL | G | - |
| INDIA 8596 | CDH13 | chr16 | 83467728 | Intron | SBS | G | C |
| INDIA 8596 | RFX3 | chr9 | 3527710 | 5'Flank | SBS | G | C |
| INDIA 8596 | DIRC3 | chr2 | 217572087 | Intron | SBS | G | C |
| INDIA 8596 | AP3B1 | chr5 | 78240434 | Intron | SBS | G | C |
| INDIA 8596 | Unknown | chr17 | 5675677 | IGR | SBS | C | G |
| INDIA 8596 | Unknown | chr9 | 71534085 | IGR | SBS | C | G |
| INDIA 8596 | ENST00000670133 | chr16 | 86952450 | Intron | SBS | G | C |
| INDIA 8596 | LINC01804 | chr2 | 15712802 | Intron | SBS | G | C |
| INDIA 8596 | Unknown | chr4 | 11006010 | IGR | SBS | C | G |
| INDIA 8596 | ANK2 | chr4 | 113131933 | Intron | SBS | C | G |
| INDIA 8597 | SBF2 | chr11 | 10269393 | Intron | SBS | G | C |
| INDIA 8597 | Unknown | chr8 | 6276650 | IGR | SBS | C | G |
| INDIA 8597 | Unknown | chr3 | 83465198 | IGR | SBS | G | C |
| INDIA 8597 | CSMD1 | chr8 | 4943148 | Intron | SBS | C | G |
| INDIA 8597 | Unknown | chr3 | 26078180 | IGR | SBS | C | G |
| INDIA 8597 | TENM2 | chr5 | 167626760 | Intron | SBS | C | G |
| INDIA 8597 | POU4F3 | chr5 | 146342321 | 3'Flank | SBS | C | G |
| INDIA 8597 | ENST00000656455 | chr8 | 36526474 | Intron | SBS | G | C |
| INDIA 8597 | TFAP2B | chr6 | 50818975 | Intron | SBS | G | C |
| INDIA 8597 | OCRL | chrX | 129566743 | Intron | SBS | G | C |
| INDIA 8597 | Unknown | chr6 | 1495592 | IGR | SBS | C | G |
| INDIA 8597 | Unknown | chr5 | 58323281 | IGR | SBS | G | C |
| INDIA 8597 | ENST00000629853 | chr6 | 14826727 | Intron | SBS | G | C |
| INDIA 8597 | Unknown | chr9 | 1686325 | IGR | DEL | G | - |

| | | | | | | | |
|---|---|---|---|---|---|---|---|
| INDIA 8597 | ENST00000656032 | chr11 | 22497578 | Intron | SBS | G | C |
| INDIA 8597 | ENST00000414774 | chr10 | 120970690 | Intron | SBS | G | C |
| INDIA 8597 | FYN | chr6 | 111746701 | Intron | SBS | C | G |
| INDIA 8597 | TP53 | chr17 | 7673781 | Missense_Mutation | SBS | C | G |
| INDIA 8597 | TRPM6 | chr9 | 74798495 | Intron | SBS | G | C |
| INDIA 8597 | TRANK1 | chr3 | 36831860 | Missense_Mutation | SBS | C | G |
| INDIA 8597 | TMEM245 | chr9 | 109099716 | Intron | SBS | C | G |
| INDIA 8597 | CARMIL1 | chr6 | 25621369 | 3'Flank | SBS | G | C |
| INDIA 8597 | LINC00845 | chr10 | 61153653 | Intron | SBS | C | G |
| INDIA 8597 | ENST00000640307 | chr9 | 23628538 | Intron | SBS | C | G |
| INDIA 8597 | FBXL17 | chr5 | 108035782 | Intron | SBS | G | C |
| INDIA 8597 | CDKAL1 | chr6 | 20824199 | Intron | SBS | C | G |
| INDIA 8597 | GABBR2 | chr9 | 98641677 | Intron | SBS | G | C |
| INDIA 8597 | Unknown | chr11 | 8369744 | IGR | SBS | A | T |
| INDIA 8597 | KRBA2 | chr17 | 8354304 | 3'Flank | SBS | C | G |
| INDIA 8597 | LINC01919 | chr18 | 53602306 | Intron | SBS | C | A |
| INDIA 8597 | Unknown | chr20 | 12037334 | IGR | SBS | G | C |
| INDIA 8597 | PMP22 | chr17 | 15237311 | Intron | DEL | T | - |
| INDIA 8597 | Unknown | chr6 | 40610262 | IGR | SBS | C | G |
| INDIA 8597 | Unknown | chr9 | 104391603 | IGR | SBS | G | C |
| INDIA 8597 | Unknown | chr17 | 15180270 | IGR | SBS | G | C |
| INDIA 8597 | NAV2 | chr11 | 19800159 | Intron | SBS | C | G |
| INDIA 8597 | Unknown | chr9 | 26166772 | IGR | SBS | T | A |
| INDIA 8597 | DNAH12 | chr3 | 57508526 | Missense_Mutation | SBS | G | C |
| INDIA 8597 | Unknown | chr11 | 26175374 | IGR | SBS | C | G |
| INDIA 8597 | SAMMSON | chr3 | 70081022 | Intron | SBS | T | A |
| INDIA 8597 | ENST00000646617 | chr5 | 163288220 | Intron | SBS | G | C |
| INDIA 8597 | GRIPAP1 | chrX | 48999464 | Nonsense_Mutation | SBS | G | C |
| INDIA 8597 | Unknown | chr8 | 26709014 | IGR | SBS | C | G |
| INDIA 8597 | DZANK1 | chr20 | 18459447 | Intron | SBS | C | A |
| INDIA 8597 | POU4F3 | chr5 | 146342990 | 3'Flank | SBS | C | G |
| INDIA 8597 | SHB | chr9 | 38009631 | Intron | SBS | C | G |
| INDIA 8597 | GPSM1 | chr9 | 136350199 | Intron | SBS | G | C |
| INDIA 8597 | Unknown | chr9 | 90062949 | IGR | SBS | G | C |
| INDIA 8597 | SYCP2L | chr6 | 10929435 | Intron | SBS | C | G |
| INDIA 8597 | VCP | chr9 | 35065574 | Intron | SBS | G | C |
| INDIA 8597 | GABRE | chrX | 151958763 | Intron | SBS | G | C |
| INDIA 8597 | SH3TC2 | chr5 | 149008972 | Missense_Mutation | SBS | C | G |
| INDIA 8597 | ENST00000563763 | chr17 | 5785092 | Intron | SBS | G | C |
| INDIA 8597 | Unknown | chr8 | 29571515 | IGR | SBS | G | C |
| INDIA 8597 | GNE | chr9 | 36276921 | Missense_Mutation | SBS | C | G |
| INDIA 8597 | Unknown | chr10 | 72325414 | IGR | SBS | C | G |
| INDIA 8597 | PIGN | chr18 | 62033324 | Intron | SBS | G | C |
| INDIA 8597 | Unknown | chr6 | 47202331 | IGR | SBS | C | G |
| INDIA 8597 | Unknown | chr10 | 117845035 | IGR | SBS | C | G |
| INDIA 8597 | SUSD6 | chr14 | 69626387 | Intron | SBS | T | A |
| INDIA 8597 | Unknown | chr6 | 115459713 | IGR | SBS | G | C |
| INDIA 8597 | TBC1D2 | chr9 | 98225071 | Intron | SBS | T | A |
| INDIA 8597 | Unknown | chr11 | 39747096 | IGR | SBS | C | G |
| INDIA 8597 | COL23A1 | chr5 | 178312641 | Intron | SBS | C | G |
| INDIA 8597 | CASC15 | chr6 | 21730667 | Intron | DEL | TTTGCAGAA | - |
| INDIA 8597 | Unknown | chr5 | 132837823 | IGR | DEL | G | - |
| INDIA 8597 | Unknown | chr9 | 26669832 | IGR | SBS | G | C |
| INDIA 8597 | GRID1 | chr10 | 85601127 | 3'UTR | DEL | A | - |
| INDIA 8597 | Unknown | chr6 | 90700550 | IGR | SBS | A | T |
| INDIA 8597 | ODF2 | chr9 | 128459502 | Intron | SBS | G | C |
| INDIA 8597 | DCDC2 | chr6 | 24330304 | Intron | SBS | G | C |
| INDIA 8597 | BSN | chr3 | 49557882 | Intron | SBS | G | C |
| INDIA 8597 | Unknown | chr6 | 113400082 | IGR | SBS | G | C |
| INDIA 8597 | NCOA7 | chr6 | 125813406 | Intron | DEL | C | - |
| INDIA 8597 | Unknown | chrX | 58279108 | IGR | SBS | C | A |
| INDIA 8597 | Unknown | chr18 | 41263395 | IGR | SBS | G | C |
| INDIA 8597 | PITX1-AS1 | chr5 | 135142119 | Intron | SBS | G | T |
| INDIA 8597 | UNC5D | chr8 | 35587454 | Intron | SBS | G | C |
| INDIA 8597 | Unknown | chr9 | 82923935 | IGR | SBS | G | C |
| INDIA 8597 | LINC03000 | chr5 | 164433161 | Intron | SBS | C | G |

| | | | | | | | |
|---|---|---|---|---|---|---|---|
| INDIA 8597 | Unknown | chr9 | 73979496 | IGR | SBS | G | T |
| INDIA 8597 | Unknown | chr10 | 61863284 | IGR | SBS | G | C |
| INDIA 8597 | Unknown | chr9 | 81516413 | IGR | SBS | G | C |
| INDIA 8597 | FRRS1L | chr9 | 109148601 | Intron | SBS | G | C |
| INDIA 8597 | LRRC75A | chr17 | 16451052 | Intron | DEL | G | - |
| INDIA 8597 | CSMD1 | chr8 | 4415215 | Intron | SBS | C | G |
| INDIA 8597 | MPRIP | chr17 | 17133261 | Intron | DEL | TC | - |
| INDIA 8597 | TSPAN18 | chr11 | 44758924 | Intron | SBS | G | C |
| INDIA 8597 | COL4A4 | chr2 | 227138883 | Intron | SBS | G | C |
| INDIA 8597 | FBXL17 | chr5 | 107892839 | Intron | SBS | C | G |
| INDIA 8597 | Unknown | chr9 | 111955558 | IGR | SBS | C | G |
| INDIA 8597 | RAPGEF1 | chr9 | 131667697 | Intron | SBS | C | G |
| INDIA 8597 | ENST00000454882 | chr6 | 5865660 | Intron | SBS | C | G |
| INDIA 8597 | ENST00000446811 | chr6 | 168260720 | RNA | SBS | C | G |
| INDIA 8597 | BCOR | chrX | 40179215 | 5'Flank | SBS | G | C |
| INDIA 8597 | ERC2 | chr3 | 55581480 | Intron | SBS | G | C |
| INDIA 8598 | TMEM63C | chr14 | 77253233 | Intron | SBS | G | C |
| INDIA 8598 | DLEU1 | chr13 | 50279765 | Intron | SBS | C | G |
| INDIA 8598 | DARS1 | chr2 | 135945018 | Intron | SBS | C | A |
| INDIA 8598 | Unknown | chr3 | 81407328 | IGR | SBS | G | C |
| INDIA 8598 | NRAD1 | chr13 | 43991755 | Intron | DEL | C | - |
| INDIA 8598 | RASGRF1 | chr15 | 79073011 | Intron | DEL | G | - |
| INDIA 8598 | Unknown | chr2 | 237851553 | IGR | SBS | C | G |
| INDIA 8598 | LINC02572 | chr2 | 129823071 | 3'Flank | SBS | G | C |
| INDIA 8598 | CRACDL | chr2 | 98896633 | Intron | DEL | G | - |
| INDIA 8598 | INA | chr10 | 103290205 | 3'UTR | DEL | C | - |
| INDIA 8598 | CPB2 | chr13 | 46078899 | Missense_Mutation | SBS | G | C |
| INDIA 8598 | TRPM2 | chr21 | 44426735 | Missense_Mutation | SBS | G | C |
| INDIA 8598 | Unknown | chr2 | 126749691 | IGR | SBS | A | T |
| INDIA 8598 | HOXD9 | chr2 | 176125319 | 3'Flank | SBS | C | G |
| INDIA 8598 | ENST00000701990 | chr6 | 1130553 | 3'Flank | SBS | C | G |
| INDIA 8598 | ENST00000431708 | chr2 | 188752534 | Intron | SBS | G | T |
| INDIA 8598 | LHFPL6 | chr13 | 39396222 | Intron | SBS | G | T |
| INDIA 8598 | Unknown | chr9 | 72440501 | IGR | SBS | T | A |
| INDIA 8598 | ENST00000456384 | chr2 | 204505675 | Intron | SBS | G | C |
| INDIA 8598 | LIPC | chr15 | 58429159 | 5'Flank | SBS | G | C |
| INDIA 8598 | CNTNAP5 | chr2 | 124667996 | Intron | SBS | C | G |
| INDIA 8598 | LRP1B | chr2 | 141942730 | Intron | DEL | T | - |
| INDIA 8598 | Unknown | chr10 | 133034634 | IGR | SBS | G | C |
| INDIA 8598 | LINC01877 | chr2 | 199620478 | Intron | SBS | G | C |
| INDIA 8598 | ENST00000690652 | chr6 | 23493197 | Intron | SBS | A | T |
| INDIA 8598 | MIR888 | chrX | 145995314 | 5'Flank | SBS | G | C |
| INDIA 8598 | Unknown | chr2 | 204135988 | IGR | DEL | T | - |
| INDIA 8598 | CRACDL | chr2 | 98794748 | Intron | SBS | C | G |
| INDIA 8598 | Unknown | chr15 | 94626164 | IGR | SBS | G | C |
| INDIA 8598 | TRPC4 | chr13 | 37722389 | Intron | SBS | G | C |
| INDIA 8598 | SNX13 | chr7 | 17908536 | Intron | SBS | C | G |
| INDIA 8598 | LINC00540 | chr13 | 22222218 | Intron | SBS | T | A |
| INDIA 8598 | OLA1 | chr2 | 174082629 | Intron | SBS | T | A |
| INDIA 8598 | TENM2 | chr5 | 167512047 | Intron | SBS | T | A |
| INDIA 8598 | XKR6 | chr8 | 11069547 | Intron | SBS | G | C |
| INDIA 8598 | DSCAM | chr21 | 40254560 | Intron | SBS | C | G |
| INDIA 8598 | Unknown | chr18 | 59125518 | IGR | SBS | G | C |
| INDIA 8598 | TTLL11 | chr9 | 121875503 | Intron | SBS | G | C |
| INDIA 8598 | PTH1R | chr3 | 46903874 | 3'Flank | SBS | T | A |
| INDIA 8598 | Unknown | chr13 | 22402797 | IGR | SBS | C | G |
| INDIA 8598 | ENST00000565523 | chr16 | 35150595 | 3'Flank | SBS | T | A |
| INDIA 8598 | FLT1 | chr13 | 28319119 | Intron | SBS | G | C |
| INDIA 8598 | Unknown | chr2 | 100756027 | IGR | SBS | G | C |
| INDIA 8598 | SDK1 | chr7 | 3538725 | Intron | SBS | C | G |
| INDIA 8598 | TMEM229B | chr14 | 67528113 | Intron | SBS | G | C |
| INDIA 8598 | Unknown | chr3 | 134472038 | IGR | SBS | C | G |
| INDIA 8598 | Unknown | chr2 | 35930869 | IGR | SBS | C | G |
| INDIA 8598 | Unknown | chr2 | 184009912 | IGR | SBS | C | G |
| INDIA 8598 | LINC02015 | chr3 | 177830726 | Intron | SBS | G | C |
| INDIA 8598 | Unknown | chr7 | 20795782 | IGR | SBS | C | G |

| | | | | | | | |
|---|---|---|---|---|---|---|---|
| INDIA 8598 | TRPM3 | chr9 | 71186488 | Intron | SBS | T | A |
| INDIA 8598 | KMT2D | chr12 | 49053647 | Splice_Region | SBS | G | C |
| INDIA 8598 | NDEL1 | chr17 | 8451881 | Intron | SBS | C | G |
| INDIA 8598 | CCT8 | chr21 | 29076024 | 5'Flank | SBS | C | G |
| INDIA 8598 | SVEP1 | chr9 | 110538775 | Intron | SBS | C | G |
| INDIA 8598 | LYPD4 | chr19 | 41838887 | Missense_Mutation | SBS | C | G |
| INDIA 8598 | LINC00540 | chr13 | 22121749 | Intron | SBS | C | A |
| INDIA 8598 | ENST00000440698 | chr2 | 51586608 | Intron | SBS | A | C |
| INDIA 8598 | LINC00882 | chr3 | 106781470 | Intron | SBS | G | C |
| INDIA 8598 | DCDC2C | chr2 | 3807401 | Intron | SBS | C | G |
| INDIA 8598 | Unknown | chr5 | 6679632 | IGR | DEL | G | - |
| INDIA 8598 | PIK3C3 | chr18 | 41956081 | Intron | SBS | G | C |
| INDIA 8598 | NAALADL2 | chr3 | 174789948 | Intron | SBS | G | T |
| INDIA 8598 | SLC35F3 | chr1 | 233949752 | Intron | SBS | G | C |
| INDIA 8598 | VAV3 | chr1 | 107694375 | Intron | SBS | T | G |
| INDIA 8598 | MYT1L | chr2 | 1788372 | 3'Flank | SBS | C | G |
| INDIA 8598 | LPP | chr3 | 188452643 | Intron | SBS | C | G |
| INDIA 8598 | SNTG2 | chr2 | 1097907 | Intron | INS | - | T |
| INDIA 8598 | CAMKMT | chr2 | 44670923 | Intron | SBS | C | G |
| INDIA 8598 | MAP3K1 | chr5 | 56873008 | Intron | INS | - | ACC |
| INDIA 8598 | Unknown | chr5 | 143241153 | IGR | SBS | C | G |
| INDIA 8598 | LOXL4 | chr10 | 98257177 | Intron | SBS | C | G |
| INDIA 8598 | LINC00550 | chr13 | 68876223 | Intron | SBS | A | T |
| INDIA 8598 | RAP2A | chr13 | 97434557 | Missense_Mutation | SBS | C | G |
| INDIA 8598 | NRXN3 | chr14 | 79473621 | Intron | SBS | G | C |
| INDIA 8598 | CDH4 | chr20 | 61823960 | Intron | DEL | T | - |
| INDIA 8598 | ENST00000454622 | chr21 | 33014145 | Intron | SBS | C | A |
| INDIA 8598 | TRIO | chr5 | 14491478 | Intron | SBS | G | C |
| INDIA 8598 | LRRC37A5P | chr9 | 111616005 | Intron | SBS | G | C |
| INDIA 8598 | QSOX2 | chr9 | 136235564 | Intron | SBS | C | G |
| INDIA 8598 | MAST2 | chr1 | 46026594 | Intron | SBS | G | C |
| INDIA 8598 | LINC01414 | chr8 | 64089884 | Intron | SBS | A | T |
| INDIA 8598 | Unknown | chr14 | 82107617 | IGR | SBS | C | G |
| INDIA 8598 | ENST00000671274 | chr18 | 70501534 | Intron | SBS | G | C |
| INDIA 8598 | HRG-AS1 | chr3 | 186741560 | Intron | SBS | C | G |
| INDIA 8598 | KLHL29 | chr2 | 23484251 | Intron | SBS | C | G |
| INDIA 8598 | Unknown | chr1 | 69388789 | IGR | SBS | A | T |
| INDIA 8598 | KLHL29 | chr2 | 23484497 | Intron | SBS | C | G |
| INDIA 8598 | Unknown | chr1 | 208374622 | IGR | SBS | G | C |
| INDIA 8598 | ADAMTS2 | chr5 | 179155672 | Intron | SBS | T | G |
| INDIA 8598 | MAGI1 | chr3 | 65709536 | Intron | SBS | G | C |
| INDIA 8598 | EFNA5 | chr5 | 107674914 | 5'Flank | SBS | G | C |
| INDIA 8598 | Unknown | chr14 | 95810903 | IGR | SBS | G | C |
| INDIA 8599 | Unknown | chr1 | 219342118 | IGR | SBS | C | G |
| INDIA 8599 | FAT2 | chr5 | 151576085 | Intron | SBS | C | G |
| INDIA 8599 | Unknown | chr1 | 240712264 | IGR | SBS | C | G |
| INDIA 8599 | COP1-DT | chr1 | 176444659 | Intron | DEL | ATC | - |
| INDIA 8599 | Unknown | chr8 | 87905449 | IGR | SBS | T | G |
| INDIA 8599 | SPAG16 | chr2 | 214082886 | Intron | SBS | C | G |
| INDIA 8599 | LINC02497 | chr4 | 31205700 | Intron | DEL | T | - |
| INDIA 8599 | Unknown | chr3 | 74628161 | IGR | SBS | G | C |
| INDIA 8599 | APP | chr21 | 26049746 | Intron | SBS | G | C |
| INDIA 8599 | OXR1 | chr8 | 106381454 | Intron | SBS | C | G |
| INDIA 8599 | SEMA5A | chr5 | 9041805 | 3'UTR | DEL | TAC | - |
| INDIA 8599 | KCNE5 | chrX | 109626673 | 5'Flank | INS | - | A |
| INDIA 8599 | ERC1 | chr12 | 1387931 | Intron | SBS | G | C |
| INDIA 8599 | ENST00000653962 | chr2 | 157045502 | Intron | DEL | T | - |
| INDIA 8599 | PTH2R | chr2 | 208483276 | Intron | SBS | A | C |
| INDIA 8599 | FCRL4 | chr1 | 157601899 | 5'Flank | SBS | A | T |
| INDIA 8599 | Unknown | chr3 | 167839588 | IGR | SBS | T | G |
| INDIA 8599 | TBC1D10A | chr22 | 30295592 | Intron | SBS | C | G |
| INDIA 8599 | LINC02162 | chr5 | 3449408 | 5'Flank | SBS | G | C |
| INDIA 8599 | DZIP3 | chr3 | 108610513 | Intron | SBS | C | G |
| INDIA 8599 | DOCK4 | chr7 | 111915050 | Intron | SBS | C | G |
| INDIA 8599 | ENST00000651429 | chr2 | 48092742 | Intron | SBS | G | C |
| INDIA 8599 | Unknown | chr8 | 128203539 | IGR | SBS | C | G |

| | | | | | | | |
|---|---|---|---|---|---|---|---|
| INDIA 8599 | Unknown | chr1 | 231303605 | IGR | SBS | C | G |
| INDIA 8599 | LINC02645 | chr10 | 2494056 | Intron | SBS | G | C |
| INDIA 8599 | CPEB2 | chr4 | 15056716 | Intron | SBS | G | C |
| INDIA 8599 | Unknown | chr8 | 137508195 | IGR | SBS | G | T |
| INDIA 8599 | GPR149 | chr3 | 154382150 | Intron | SBS | G | C |
| INDIA 8599 | Unknown | chr12 | 114612569 | IGR | SBS | G | C |
| INDIA 8599 | Unknown | chr20 | 40530605 | IGR | SBS | C | G |
| INDIA 8599 | CHCHD6 | chr3 | 126711212 | Intron | SBS | G | C |
| INDIA 8599 | EXT1 | chr8 | 117810118 | Intron | SBS | C | G |
| INDIA 8599 | SLC4A4 | chr4 | 71454166 | Intron | SBS | T | G |
| INDIA 8599 | Unknown | chr5 | 74053009 | IGR | SBS | G | C |
| INDIA 8599 | CREB5 | chr7 | 28435738 | Intron | SBS | C | G |
| INDIA 8599 | DPYD | chr1 | 97258694 | Intron | SBS | G | C |
| INDIA 8599 | CR1L | chr1 | 207706675 | Intron | SBS | G | C |
| INDIA 8599 | Unknown | chr3 | 163516261 | IGR | DEL | A | - |
| INDIA 8599 | Unknown | chr3 | 182734258 | IGR | SBS | G | C |
| INDIA 8599 | TBC1D9B | chr5 | 179899648 | Intron | SBS | C | G |
| INDIA 8599 | ZNF395 | chr8 | 28383228 | Intron | SBS | C | G |
| INDIA 8599 | Unknown | chr12 | 90523989 | IGR | SBS | G | C |
| INDIA 8599 | NEMP2-DT | chr2 | 190547988 | Intron | SBS | C | G |
| INDIA 8599 | MYO10 | chr5 | 16692911 | Intron | DEL | TGTAC | - |
| INDIA 8599 | GHR | chr5 | 42726159 | 3'Flank | DEL | TT | - |
| INDIA 8599 | TRAPPC9 | chr8 | 140139168 | Intron | SBS | G | C |
| INDIA 8599 | VIPR2 | chr7 | 159057878 | Intron | SBS | C | G |
| INDIA 8599 | Metazoa_SRP | chr12 | 68578967 | 5'Flank | SBS | G | C |
| INDIA 8599 | Unknown | chr10 | 20342484 | IGR | SBS | C | G |
| INDIA 8599 | SAMD4B | chr19 | 39346410 | Intron | SBS | G | C |
| INDIA 8599 | KCNJ15 | chr21 | 38250268 | Intron | SBS | G | C |
| INDIA 8599 | PCBP3 | chr21 | 45911045 | Intron | SBS | C | G |
| INDIA 8599 | BASP1 | chr5 | 17267319 | Intron | SBS | G | C |
| INDIA 8599 | SHCBP1 | chr16 | 46599967 | Splice_Region | DEL | G | - |
| INDIA 8599 | OVOL2 | chr20 | 18023476 | 3'Flank | SBS | G | C |
| INDIA 8599 | DHRS3 | chr1 | 12581842 | Intron | SBS | A | C |
| INDIA 8599 | DSCAML1 | chr11 | 117623122 | Intron | SBS | C | G |
| INDIA 8599 | CAMSAP2 | chr1 | 200853350 | Missense_Mutation | SBS | G | C |
| INDIA 8599 | IL12A-AS1 | chr3 | 160092776 | Intron | SBS | C | G |
| INDIA 8599 | KCNH7 | chr2 | 162446204 | Missense_Mutation | SBS | C | G |
| INDIA 8599 | Unknown | chr16 | 48903816 | IGR | SBS | C | G |
| INDIA 8599 | PLCB1 | chr20 | 8329791 | Intron | SBS | C | G |
| INDIA 8599 | PBX1 | chr1 | 164688210 | Intron | SBS | C | G |
| INDIA 8599 | LINC02897 | chr1 | 247112805 | 5'Flank | SBS | C | G |
| INDIA 8599 | IGF2BP2 | chr3 | 185828408 | 5'Flank | DEL | ATCTTG | - |
| INDIA 8599 | ATPSCKMT | chr5 | 10232817 | Intron | SBS | G | C |
| INDIA 8599 | Unknown | chr11 | 57834826 | IGR | SBS | G | C |
| INDIA 8599 | RERE | chr1 | 8411457 | Intron | SBS | G | C |
| INDIA 8599 | EFHD1 | chr2 | 232640136 | Intron | SBS | C | G |
| INDIA 8599 | ZNF678 | chr1 | 227616260 | Intron | SBS | C | G |
| INDIA 8599 | ENST00000560484 | chr15 | 38887597 | Intron | SBS | C | G |
| INDIA 8599 | TP63 | chr3 | 189732305 | Intron | SBS | G | C |
| INDIA 8599 | VPS37B | chr12 | 122874070 | Intron | SBS | G | C |
| INDIA 8599 | ENST00000648066 | chr14 | 53628814 | Intron | SBS | G | C |
| INDIA 8599 | LINC01675 | chr1 | 166485460 | Intron | SBS | C | G |
| INDIA 8599 | LINC02934 | chr2 | 66126268 | Intron | SBS | C | G |
| INDIA 8599 | Unknown | chr9 | 1947408 | IGR | SBS | G | C |
| INDIA 8599 | LINC01423 | chr21 | 38325001 | Intron | SBS | C | G |
| INDIA 8599 | Unknown | chr2 | 31304680 | IGR | SBS | C | G |
| INDIA 8599 | ABI2 | chr2 | 203408765 | Intron | SBS | C | G |
| INDIA 8599 | Unknown | chr17 | 78957416 | IGR | SBS | G | C |
| INDIA 8599 | DAB1 | chr1 | 58396287 | Intron | DEL | AACA | - |
| INDIA 8599 | ITGA6 | chr2 | 172481057 | Intron | SBS | C | G |
| INDIA 8599 | Unknown | chr8 | 141821561 | IGR | SBS | C | G |
| INDIA 8599 | APOD | chr3 | 195563747 | 3'Flank | SBS | G | C |
| INDIA 8599 | Unknown | chr7 | 135355188 | IGR | SBS | C | G |
| INDIA 8599 | LRRC4C | chr11 | 41165375 | Intron | SBS | G | C |
| INDIA 8599 | Unknown | chr22 | 46142738 | IGR | SBS | C | G |
| INDIA 8599 | INC01358 | chr1 | 58982951 | Intron | SBS | G | C |

| | | | | | | | |
|---|---|---|---|---|---|---|---|
| INDIA 8599 | PLA1A | chr3 | 119628947 | Intron | SBS | G | C |
| INDIA 8599 | CDKAL1 | chr6 | 20589423 | Intron | SBS | C | G |
| INDIA 8599 | CA8 | chr8 | 60260296 | Intron | SBS | G | C |
| INDIA 8600 | DYM | chr18 | 49412635 | Intron | SBS | G | C |
| INDIA 8600 | CTIF | chr18 | 48659024 | Intron | SBS | G | C |
| INDIA 8600 | METTL16 | chr17 | 2512255 | 5'Flank | SBS | G | C |
| INDIA 8600 | WARS2-AS1 | chr1 | 119230723 | Intron | SBS | G | C |
| INDIA 8600 | FIBIN | chr11 | 26991857 | 5'Flank | SBS | G | C |
| INDIA 8600 | LINC02854 | chr7 | 53931821 | Intron | DEL | A | - |
| INDIA 8600 | WSCD1 | chr17 | 6101058 | Intron | SBS | G | C |
| INDIA 8600 | Unknown | chr8 | 79726036 | IGR | SBS | G | C |
| INDIA 8600 | ARHGAP44 | chr17 | 12918044 | Intron | SBS | C | G |
| INDIA 8600 | DCC | chr18 | 53081011 | Intron | SBS | A | C |
| INDIA 8600 | ARHGAP44 | chr17 | 12917910 | Intron | SBS | C | G |
| INDIA 8600 | SLC9A7 | chrX | 46634842 | Intron | SBS | G | C |
| INDIA 8600 | MAOB | chrX | 43791287 | Intron | SBS | C | G |
| INDIA 8600 | Unknown | chr2 | 221206646 | IGR | SBS | C | G |
| INDIA 8600 | ENST00000685138 | chr20 | 41655565 | Intron | SBS | C | G |
| INDIA 8600 | SHROOM2 | chrX | 9934248 | Intron | SBS | G | C |
| INDIA 8600 | Unknown | chr8 | 54254157 | IGR | SBS | G | C |
| INDIA 8600 | Unknown | chr5 | 16948486 | IGR | SBS | G | C |
| INDIA 8600 | Unknown | chr14 | 45445161 | IGR | SBS | C | G |
| INDIA 8600 | GPC3 | chrX | 133681795 | Intron | SBS | G | C |
| INDIA 8600 | ENST00000651527 | chr1 | 212527300 | Intron | SBS | G | C |
| INDIA 8600 | C19orf12 | chr19 | 29707460 | Intron | SBS | G | C |
| INDIA 8600 | ENST00000568659 | chr16 | 65600238 | 3'Flank | SBS | T | A |
| INDIA 8600 | Unknown | chr18 | 41074430 | IGR | SBS | C | G |
| INDIA 8600 | FGGY | chr1 | 59633593 | Intron | SBS | G | C |
| INDIA 8600 | Unknown | chr2 | 181438902 | IGR | SBS | C | G |
| INDIA 8600 | ENST00000697639 | chr9 | 118110819 | Intron | SBS | G | C |
| INDIA 8600 | SGCZ | chr8 | 15180244 | Intron | SBS | G | C |
| INDIA 8600 | PEMT | chr17 | 17557022 | Intron | SBS | C | G |
| INDIA 8600 | Unknown | chr1 | 3006095 | IGR | SBS | G | C |
| INDIA 8600 | Unknown | chr8 | 107805386 | IGR | SBS | C | G |
| INDIA 8600 | Unknown | chr18 | 38024028 | IGR | SBS | G | T |
| INDIA 8600 | LINC00670 | chr17 | 12584345 | Intron | SBS | G | C |
| INDIA 8600 | DIS3L2 | chr2 | 232249835 | Intron | SBS | G | C |
| INDIA 8600 | WDR5 | chr9 | 134155777 | Intron | SBS | A | T |
| INDIA 8600 | Unknown | chr12 | 114046530 | IGR | DEL | A | - |
| INDIA 8600 | LINC00407 | chr13 | 44655539 | Intron | SBS | G | C |
| INDIA 8600 | DCDC1 | chr11 | 31127708 | Intron | SBS | G | C |
| INDIA 8600 | Unknown | chr1 | 186805558 | IGR | SBS | G | C |
| INDIA 8600 | ATP2B4 | chr1 | 203628184 | Intron | SBS | G | C |
| INDIA 8600 | GPT2 | chr16 | 46931117 | 3'UTR | SBS | C | A |
| INDIA 8600 | FREM3 | chr4 | 143698733 | Missense_Mutation | SBS | C | G |
| INDIA 8600 | Unknown | chr17 | 11094578 | IGR | SBS | C | G |
| INDIA 8600 | Unknown | chr8 | 119155453 | IGR | SBS | G | C |
| INDIA 8600 | ATG4B | chr2 | 241638201 | Intron | SBS | G | C |
| INDIA 8600 | TMEM243 | chr7 | 87199173 | Intron | SBS | G | C |
| INDIA 8600 | Unknown | chr13 | 58088171 | IGR | SBS | G | C |
| INDIA 8600 | LINC00670 | chr17 | 12551488 | Intron | SBS | T | A |
| INDIA 8600 | ST8SIA3 | chr18 | 57368791 | 3'UTR | SBS | G | C |
| INDIA 8600 | SNTG1 | chr8 | 50332886 | Intron | SBS | C | G |
| INDIA 8600 | Unknown | chr8 | 56728291 | IGR | SBS | T | A |
| INDIA 8600 | NUP210 | chr3 | 13385283 | Intron | SBS | C | G |
| INDIA 8600 | CRTAC1 | chr10 | 98032085 | 5'Flank | SBS | C | G |
| INDIA 8600 | ZNF99 | chr19 | 22754046 | 3'UTR | SBS | G | C |
| INDIA 8600 | ENST00000644134 | chr1 | 192831544 | Intron | SBS | G | T |
| INDIA 8600 | Unknown | chr14 | 92281158 | IGR | SBS | G | C |
| INDIA 8600 | Unknown | chr1 | 17712231 | IGR | SBS | C | G |
| INDIA 8600 | MYH7B | chr20 | 34960109 | Intron | SBS | G | C |
| INDIA 8600 | C1orf127 | chr1 | 10961390 | Intron | SBS | C | G |
| INDIA 8600 | RNF180 | chr5 | 64286388 | Intron | DEL | T | - |
| INDIA 8600 | Unknown | chr1 | 33437276 | IGR | SBS | C | G |
| INDIA 8600 | ALG6 | chr1 | 63404486 | Silent | SBS | C | G |
| INDIA 8600 | ENST00000456031 | chr2 | 199054513 | Intron | SBS | G | C |

| | | | | | | | |
|---|---|---|---|---|---|---|---|
| INDIA 8600 | Unknown | chr2 | 238803899 | IGR | SBS | G | C |
| INDIA 8600 | LRRC14B | chr5 | 189385 | 5'Flank | SBS | C | G |
| INDIA 8600 | Unknown | chr5 | 3312489 | IGR | SBS | C | G |
| INDIA 8600 | Unknown | chr6 | 80723341 | IGR | SBS | A | T |
| INDIA 8600 | GPR141 | chr7 | 37747324 | 3'Flank | SBS | C | G |
| INDIA 8600 | Unknown | chr7 | 57438343 | IGR | SBS | G | T |
| INDIA 8600 | CDK6 | chr7 | 92805079 | Intron | SBS | C | G |
| INDIA 8600 | Unknown | chr8 | 119155532 | IGR | SBS | G | C |
| INDIA 8600 | NRG3 | chr10 | 82052665 | Intron | SBS | A | T |
| INDIA 8600 | LINC01435 | chr10 | 108002861 | Intron | SBS | G | C |
| INDIA 8600 | BTBD11 | chr12 | 107468197 | Intron | SBS | C | G |
| INDIA 8600 | RIMBP2 | chr12 | 130568100 | Intron | SBS | G | C |
| INDIA 8600 | MIR17HG | chr13 | 91358003 | 3'Flank | SBS | G | C |
| INDIA 8600 | CLEC14A | chr14 | 38255323 | Missense_Mutation | SBS | G | C |
| INDIA 8600 | ENST00000647580 | chr14 | 77001142 | 5'Flank | SBS | C | G |
| INDIA 8600 | Unknown | chr14 | 95382690 | IGR | SBS | C | G |
| INDIA 8600 | TANC2 | chr17 | 63153332 | Intron | SBS | A | T |
| INDIA 8600 | FAM9B | chrX | 9090586 | Intron | SBS | G | C |
| INDIA 8600 | SGCZ | chr8 | 15179869 | Intron | SBS | G | C |
| INDIA 8600 | FKBP9 | chr7 | 32964103 | Intron | SBS | C | G |
| INDIA 8600 | IGSF21 | chr1 | 18257686 | Intron | SBS | C | G |
| INDIA 8600 | Unknown | chr2 | 165413154 | IGR | SBS | C | G |
| INDIA 8600 | ADAMTSL2 | chr9 | 133541953 | Intron | SBS | C | G |
| INDIA 8600 | DSCAML1 | chr11 | 117442623 | Intron | SBS | C | G |
| INDIA 8600 | ANO4 | chr12 | 100834402 | Intron | SBS | G | C |
| INDIA 8600 | TMEM132D | chr12 | 129354857 | Intron | SBS | C | G |
| INDIA 8600 | DNAH11 | chr7 | 21888010 | Intron | SBS | G | C |
| INDIA 8600 | Unknown | chr8 | 30957137 | IGR | SBS | C | G |
| INDIA 8600 | DRAIC | chr15 | 69722085 | Intron | SBS | G | C |
| INDIA 8601 | PXMP4 | chr20 | 33723473 | 5'Flank | SBS | C | G |
| INDIA 8601 | CTDSPL | chr3 | 37936953 | Intron | SBS | G | C |
| INDIA 8601 | Unknown | chr6 | 87935877 | IGR | SBS | C | G |
| INDIA 8601 | OSBPL1A | chr18 | 24333263 | Intron | SBS | C | G |
| INDIA 8601 | APH1B | chr15 | 63308515 | 3'UTR | SBS | C | G |
| INDIA 8601 | Unknown | chr12 | 97405087 | IGR | SBS | A | C |
| INDIA 8601 | CNTNAP2 | chr7 | 147569193 | Intron | SBS | G | C |
| INDIA 8601 | TMEM108 | chr3 | 133384739 | Intron | SBS | G | C |
| INDIA 8601 | ZBTB20 | chr3 | 114701041 | Intron | SBS | G | C |
| INDIA 8601 | MECOM | chr3 | 169368594 | Intron | SBS | C | G |
| INDIA 8601 | NKAIN2 | chr6 | 123867171 | Intron | SBS | C | G |
| INDIA 8601 | TNS3 | chr7 | 47304907 | Missense_Mutation | SBS | G | C |
| INDIA 8601 | TRIM44 | chr11 | 35676567 | Intron | SBS | G | C |
| INDIA 8601 | KMT5B | chr11 | 68216731 | 5'Flank | SBS | G | C |
| INDIA 8601 | ENST00000653491 | chr8 | 49234013 | 3'Flank | SBS | A | C |
| INDIA 8601 | EIF4G3 | chr1 | 21025387 | Intron | SBS | C | G |
| INDIA 8601 | Unknown | chr2 | 191510922 | IGR | SBS | G | C |
| INDIA 8601 | RARB | chr3 | 25486075 | Intron | SBS | G | C |
| INDIA 8601 | GMPPB | chr3 | 49722828 | Intron | SBS | C | G |
| INDIA 8601 | ELMO1 | chr7 | 36919355 | Intron | SBS | C | G |
| INDIA 8601 | ENST00000656697 | chr7 | 112722451 | Intron | SBS | G | C |
| INDIA 8601 | SDC2 | chr8 | 96498415 | Intron | SBS | C | G |
| INDIA 8601 | DIS3 | chr13 | 72780820 | Intron | SBS | G | C |
| INDIA 8601 | SH3GL3 | chr15 | 83614845 | Intron | SBS | C | G |
| INDIA 8601 | ENST00000412410 | chr7 | 109589869 | Intron | SBS | C | G |
| INDIA 8601 | DAB2 | chr5 | 39398639 | Intron | SBS | G | C |
| INDIA 8601 | SRGAP3 | chr3 | 8981040 | 3'UTR | SBS | G | C |
| INDIA 8601 | Unknown | chr1 | 6737809 | IGR | SBS | C | G |
| INDIA 8601 | NUP210L | chr1 | 153999266 | Intron | SBS | C | G |
| INDIA 8601 | GMDS | chr6 | 1932098 | Intron | SBS | G | C |
| INDIA 8601 | Unknown | chr9 | 131791382 | IGR | SBS | C | G |
| INDIA 8601 | DLG2 | chr11 | 85223052 | Intron | SBS | G | C |
| INDIA 8601 | PTGS1 | chr9 | 122387381 | Intron | SBS | C | G |
| INDIA 8601 | Unknown | chr13 | 80210677 | IGR | SBS | C | G |
| INDIA 8601 | ENST00000665142 | chr10 | 22235315 | 5'Flank | SBS | C | G |
| INDIA 8601 | ENST00000652412 | chr12 | 67092313 | Intron | SBS | G | C |
| INDIA 8601 | TTLL13 | chr15 | 90251766 | Intron | SBS | G | C |

| | | | | | | | |
|---|---|---|---|---|---|---|---|
| INDIA 8601 | Unknown | chr11 | 106635371 | IGR | SBS | C | G |
| INDIA 8601 | Unknown | chr13 | 103278860 | IGR | SBS | G | C |
| INDIA 8601 | STK35 | chr20 | 2143400 | Intron | SBS | G | C |
| INDIA 8601 | Unknown | chr10 | 111342249 | IGR | SBS | C | G |
| INDIA 8601 | PRELID3A | chr18 | 12411532 | Intron | SBS | G | T |
| INDIA 8601 | MAD1L1 | chr7 | 2001930 | Intron | SBS | C | G |
| INDIA 8601 | Unknown | chr1 | 90351575 | IGR | SBS | A | T |
| INDIA 8601 | ENST00000425894 | chr3 | 6083314 | Intron | SBS | C | G |
| INDIA 8601 | ENST00000511579 | chr5 | 33518160 | 3'Flank | SBS | G | C |
| INDIA 8601 | Unknown | chr8 | 135686815 | IGR | SBS | C | G |
| INDIA 8601 | Unknown | chr5 | 85952776 | IGR | SBS | G | C |
| INDIA 8601 | SLC19A1 | chr21 | 45515673 | Missense_Mutation | SBS | C | G |
| INDIA 8601 | DAB1 | chr1 | 57828448 | Intron | SBS | G | C |
| INDIA 8601 | ENST00000669290 | chr15 | 28876745 | 3'Flank | SBS | C | G |
| INDIA 8601 | Unknown | chr13 | 74979039 | IGR | SBS | G | C |
| INDIA 8601 | ELL | chr19 | 18500874 | Intron | SBS | G | C |
| INDIA 8601 | TWIST1 | chr7 | 19095077 | Intron | SBS | C | A |
| INDIA 8601 | SMPD4P1 | chr22 | 20641099 | Intron | SBS | G | C |
| INDIA 8601 | CNTNAP2 | chr7 | 146591539 | Intron | SBS | G | C |
| INDIA 8601 | LINC02227 | chr5 | 158381994 | Intron | SBS | C | G |
| INDIA 8601 | Unknown | chr6 | 45835357 | IGR | SBS | C | G |
| INDIA 8601 | LINC02932 | chr7 | 91436873 | Intron | SBS | T | A |
| INDIA 8601 | GAB4 | chr22 | 16963413 | Intron | SBS | G | C |
| INDIA 8601 | ENST00000515213 | chr7 | 378538 | 5'Flank | SBS | G | T |
| INDIA 8603 | ENST00000649628 | chr8 | 30203095 | 3'Flank | SBS | C | G |
| INDIA 8603 | Unknown | chr4 | 117984521 | IGR | DEL | A | - |
| INDIA 8603 | ENST00000688155 | chr13 | 53432308 | RNA | SBS | C | A |
| INDIA 8603 | RBFOX1 | chr16 | 6520483 | Intron | SBS | T | A |
| INDIA 8603 | Unknown | chr16 | 60545904 | IGR | DEL | G | - |
| INDIA 8603 | CSMD3 | chr8 | 113436635 | Intron | SBS | C | A |
| INDIA 8603 | CDH4 | chr20 | 61293323 | Intron | SBS | C | A |
| INDIA 8603 | ZNF81 | chrX | 47889508 | Intron | SBS | G | T |
| INDIA 8603 | Unknown | chr6 | 62663326 | IGR | DEL | TG | - |
| INDIA 8603 | HOXC11 | chr12 | 53971302 | 5'Flank | SBS | G | C |
| INDIA 8603 | PXK | chr3 | 58358821 | Intron | INS | - | T |
| INDIA 8603 | NRG3 | chr10 | 82858717 | Intron | SBS | T | A |
| INDIA 8603 | Unknown | chr14 | 46791894 | IGR | SBS | T | G |
| INDIA 8603 | ATRNL1 | chr10 | 115859602 | Intron | SBS | A | C |
| INDIA 8603 | Unknown | chr18 | 63085852 | IGR | SBS | G | T |
| INDIA 8603 | SLC45A4 | chr8 | 141300899 | Intron | SBS | G | T |
| INDIA 8603 | ENST00000702159 | chr3 | 164533465 | Intron | SBS | A | T |
| INDIA 8603 | UBALD1 | chr16 | 4614064 | Intron | SBS | G | C |
| INDIA 8603 | Unknown | chr16 | 64766323 | IGR | SBS | C | G |
| INDIA 8603 | ENST00000443163 | chr9 | 86643647 | Intron | SBS | T | A |
| INDIA 8603 | UNC5D | chr8 | 35378461 | Intron | SBS | G | C |
| INDIA 8603 | B3GAT1-DT | chr11 | 134420988 | Intron | SBS | G | C |
| INDIA 8603 | ENST00000636095 | chrX | 137970617 | Intron | SBS | T | A |
| INDIA 8603 | Unknown | chr10 | 108434977 | IGR | DEL | G | - |
| INDIA 8603 | Unknown | chr13 | 111913354 | IGR | SBS | T | A |
| INDIA 8603 | DENND1A | chr9 | 123669638 | Intron | SBS | C | A |
| INDIA 8603 | SMARCD3 | chr7 | 151242583 | Silent | SBS | G | C |
| INDIA 8603 | ACSL4 | chrX | 109651723 | Intron | SBS | C | G |
| INDIA 8603 | ENST00000638199 | chr2 | 164979469 | Intron | DEL | C | - |
| INDIA 8603 | Unknown | chrX | 16419671 | IGR | SBS | A | T |
| INDIA 8603 | KCNIP4 | chr4 | 21213138 | Intron | SBS | G | T |
| INDIA 8603 | Unknown | chr18 | 8679269 | IGR | SBS | G | T |
| INDIA 8603 | Unknown | chr20 | 6351267 | IGR | SBS | A | T |
| INDIA 8603 | ENST00000440698 | chr2 | 51050381 | Intron | DEL | G | - |
| INDIA 8603 | ENST00000657880 | chr2 | 122035090 | Intron | SBS | C | A |
| INDIA 8603 | Unknown | chr2 | 129091446 | IGR | SBS | C | A |
| INDIA 8603 | SCN2A | chr2 | 165362978 | Intron | SBS | G | C |
| INDIA 8603 | Unknown | chr3 | 81426996 | IGR | SBS | C | G |
| INDIA 8603 | ZNF385D | chr3 | 22019176 | Intron | SBS | C | G |
| INDIA 8603 | EIF2S2 | chr20 | 34097240 | Intron | DEL | GG | - |
| INDIA 8603 | MYH9 | chr22 | 36327505 | Intron | DEL | ATCAGAT | - |
| INDIA 8603 | Unknown | chr5 | 29779489 | IGR | SBS | C | G |

| | | | | | | | |
|---|---|---|---|---|---|---|---|
| INDIA 8603 | MYLK | chr3 | 123691217 | Intron | SBS | C | G |
| INDIA 8603 | Unknown | chr2 | 123602765 | IGR | SBS | C | G |
| INDIA 8603 | ENST00000646532 | chr11 | 127162317 | Intron | SBS | C | G |
| INDIA 8603 | DMD | chrX | 32889946 | Intron | SBS | A | T |
| INDIA 8603 | Unknown | chr2 | 107096966 | IGR | SBS | T | G |
| INDIA 8603 | SLC24A3 | chr20 | 19668653 | Intron | DEL | TTCT | - |
| INDIA 8603 | Unknown | chrX | 145314899 | IGR | DEL | C | - |
| INDIA 8603 | RABGAP1L | chr1 | 174658584 | Intron | INS | - | AA |
| INDIA 8603 | Unknown | chr5 | 3100068 | IGR | SBS | G | C |
| INDIA 8603 | CTNND2 | chr5 | 11770313 | Intron | SBS | C | G |
| INDIA 8603 | IL1RAPL2 | chrX | 104917598 | Intron | SBS | A | C |
| INDIA 8603 | ENST00000659527 | chr2 | 5709056 | Intron | SBS | G | C |
| INDIA 8603 | PRKCE | chr2 | 45988840 | Intron | SBS | G | T |
| INDIA 8603 | Unknown | chr4 | 122467190 | IGR | SBS | G | T |
| INDIA 8603 | ENST00000649380 | chr9 | 24581094 | Intron | SBS | G | C |
| INDIA 8603 | PPP1R21 | chr2 | 48516144 | 3'Flank | INS | - | T |
| INDIA 8603 | ENST00000651788 | chrX | 121107705 | Intron | SBS | C | G |
| INDIA 8603 | FAM20B | chr1 | 179027566 | Intron | SBS | G | T |
| INDIA 8603 | Unknown | chr2 | 147222458 | IGR | SBS | T | A |
| INDIA 8603 | ENST00000415393 | chr7 | 131930270 | Intron | SBS | G | T |
| INDIA 8603 | ZNF837 | chr19 | 58372602 | Intron | SBS | C | G |
| INDIA 8603 | CPQ | chr8 | 97080033 | Intron | SBS | T | A |
| INDIA 8603 | Unknown | chr13 | 54178519 | IGR | DEL | A | - |
| INDIA 8603 | DAPL1 | chr2 | 158817828 | 3'Flank | SBS | A | C |
| INDIA 8603 | Unknown | chr4 | 126909299 | IGR | SBS | C | A |
| INDIA 8603 | LINGO2 | chr9 | 28434633 | Intron | SBS | C | G |
| INDIA 8603 | ENST00000648066 | chr14 | 53403584 | Intron | SBS | C | A |
| INDIA 8603 | RAMP2 | chr17 | 42763388 | 3'Flank | SBS | C | A |
| INDIA 8603 | LINC00639 | chr14 | 38816443 | Intron | SBS | C | G |
| INDIA 8603 | PTPRT | chr20 | 42582796 | Intron | SBS | C | G |
| INDIA 8603 | TTC4 | chr1 | 54713095 | 5'Flank | SBS | G | C |
| INDIA 8603 | Unknown | chr13 | 68242116 | IGR | SBS | G | C |
| INDIA 8603 | OCIAD1 | chr4 | 48851922 | Frame_Shift_Del | DEL | GT | - |
| INDIA 8603 | Unknown | chr2 | 82666062 | IGR | SBS | C | G |
| INDIA 8603 | MGC4859 | chr7 | 10768144 | Intron | SBS | A | T |
| INDIA 8603 | SOX3 | chrX | 140506540 | 5'Flank | SBS | A | T |
| INDIA 8603 | SIPA1L1 | chr14 | 71482529 | Intron | SBS | C | G |
| INDIA 8603 | LINC02008 | chr3 | 82214256 | Intron | SBS | C | A |
| INDIA 8603 | SLC14A2 | chr18 | 45563156 | Intron | DEL | TGTAGAA | - |
| INDIA 8603 | MID2 | chrX | 107904453 | Intron | SBS | G | T |
| INDIA 8604 | APOBEC1 | chr12 | 7667412 | 5'Flank | SBS | C | G |
| INDIA 8604 | FLT4 | chr5 | 180612394 | Intron | SBS | C | G |
| INDIA 8604 | EPS15-AS1 | chr1 | 51560545 | Intron | SBS | G | C |
| INDIA 8604 | JAK1 | chr1 | 64935005 | Intron | SBS | C | G |
| INDIA 8604 | ENST00000426204 | chr9 | 111186947 | Intron | SBS | G | C |
| INDIA 8604 | PRRG3 | chrX | 151703610 | 3'UTR | SBS | G | C |
| INDIA 8604 | ENST00000430748 | chr1 | 81604839 | Intron | SBS | G | C |
| INDIA 8604 | PKN2-AS1 | chr1 | 87797781 | Intron | SBS | C | G |
| INDIA 8604 | Unknown | chr9 | 7660396 | IGR | SBS | A | T |
| INDIA 8604 | Unknown | chrX | 106347660 | IGR | SBS | G | T |
| INDIA 8604 | Unknown | chr2 | 80949392 | IGR | SBS | G | C |
| INDIA 8604 | ENST00000652827 | chr9 | 99425093 | Intron | SBS | C | G |
| INDIA 8604 | NCAPD3 | chr11 | 134168529 | Missense_Mutation | SBS | C | G |
| INDIA 8604 | HSD17B4 | chr5 | 119493988 | Intron | SBS | G | C |
| INDIA 8604 | Unknown | chr9 | 90241898 | IGR | SBS | C | G |
| INDIA 8604 | CARD18 | chr11 | 105148373 | Intron | SBS | G | C |
| INDIA 8604 | ENST00000516487 | chrX | 100104767 | 3'Flank | SBS | G | C |
| INDIA 8604 | FGD3 | chr9 | 93030590 | Intron | SBS | G | C |
| INDIA 8604 | TXNDC12 | chr1 | 52033849 | Intron | SBS | C | G |
| INDIA 8604 | RBBP7 | chrX | 16869263 | Intron | SBS | G | C |
| INDIA 8604 | DDAH1 | chr1 | 85330449 | Intron | SBS | C | G |
| INDIA 8604 | ENST00000636819 | chr5 | 159777863 | Intron | SBS | G | C |
| INDIA 8604 | PTCHD1 | chrX | 23392354 | Intron | SBS | A | T |
| INDIA 8604 | Unknown | chr9 | 117628948 | IGR | SBS | C | G |
| INDIA 8604 | TEAD1 | chr11 | 12803279 | Intron | SBS | G | C |
| INDIA 8604 | FAR2 | chr12 | 29312427 | Intron | SBS | C | G |

| | | | | | | | |
|---|---|---|---|---|---|---|---|
| INDIA 8604 | LINC01362 | chr1 | 83062987 | Intron | SBS | G | C |
| INDIA 8604 | CA10 | chr17 | 51868574 | Intron | SBS | C | G |
| INDIA 8604 | DUSP16 | chr12 | 12473762 | 3'UTR | SBS | G | C |
| INDIA 8604 | PPAN | chr19 | 10106150 | 5'Flank | SBS | G | C |
| INDIA 8604 | ENST00000419596 | chr1 | 58077114 | Intron | SBS | C | G |
| INDIA 8604 | Unknown | chr5 | 53450683 | IGR | SBS | G | C |
| INDIA 8604 | Unknown | chr9 | 23905161 | IGR | SBS | G | C |
| INDIA 8604 | ERCC6L2 | chr9 | 95942081 | Intron | SBS | G | C |
| INDIA 8604 | ZNF555 | chr19 | 2849291 | Intron | SBS | G | C |
| INDIA 8604 | LINC01863 | chr5 | 173647614 | Intron | SBS | G | C |
| INDIA 8604 | TMEM255A | chrX | 120285198 | Silent | SBS | G | C |
| INDIA 8604 | ANKFN1 | chr17 | 56512719 | 3'UTR | SBS | G | C |
| INDIA 8604 | LINC01933 | chr5 | 152178068 | Intron | SBS | C | G |
| INDIA 8604 | LINC01411 | chr5 | 174461051 | Intron | SBS | C | G |
| INDIA 8604 | NFX1 | chr9 | 33313861 | Intron | SBS | G | C |
| INDIA 8604 | CRB2 | chr9 | 123378620 | 3'UTR | SBS | C | G |
| INDIA 8604 | LRP6 | chr12 | 12160560 | Intron | SBS | G | C |
| INDIA 8604 | RBM33 | chr7 | 155731118 | Intron | SBS | C | G |
| INDIA 8604 | RAB40AL | chrX | 102941069 | 3'Flank | SBS | C | G |
| INDIA 8604 | ENST00000659022 | chrX | 7938360 | Intron | SBS | C | G |
| INDIA 8604 | DAB2IP | chr9 | 121597540 | 5'Flank | SBS | G | C |
| INDIA 8604 | CACNA1A | chr19 | 13292931 | Intron | SBS | C | G |
| INDIA 8604 | GABRA2 | chr4 | 46337324 | Intron | SBS | G | T |
| INDIA 8604 | ATRNL1 | chr10 | 115953435 | 3'Flank | SBS | G | C |
| INDIA 8604 | KDM1A | chr1 | 23066401 | Intron | SBS | C | G |
| INDIA 8604 | GTDC1 | chr2 | 144081495 | Intron | SBS | C | G |
| INDIA 8604 | ZNF682 | chr19 | 20034309 | Intron | SBS | C | G |
| INDIA 8604 | CCBE1 | chr18 | 59545332 | Intron | SBS | C | G |
| INDIA 8604 | Unknown | chr9 | 107778691 | IGR | SBS | C | G |
| INDIA 8604 | ENST00000457975 | chr10 | 55379745 | Intron | DEL | G | - |
| INDIA 8604 | Unknown | chr11 | 110532005 | IGR | SBS | C | G |
| INDIA 8604 | Unknown | chr17 | 5679773 | IGR | SBS | A | C |
| INDIA 8604 | MAST4 | chr5 | 67010821 | Intron | SBS | G | C |
| INDIA 8604 | PRTFDC1 | chr10 | 24913221 | Intron | SBS | G | C |
| INDIA 8604 | Unknown | chr3 | 168983076 | IGR | SBS | C | G |
| INDIA 8604 | ENST00000510621 | chr5 | 67879640 | Intron | SBS | C | G |
| INDIA 8604 | CKMT2-AS1 | chr5 | 81286644 | Intron | SBS | C | G |
| INDIA 8604 | Unknown | chr9 | 108590604 | IGR | SBS | G | C |
| INDIA 8604 | ENST00000544515 | chr12 | 118853697 | Intron | SBS | G | C |
| INDIA 8604 | ZZEF1 | chr17 | 4058422 | Intron | SBS | C | G |
| INDIA 8604 | ANKFY1 | chr17 | 4263053 | Intron | SBS | G | C |
| INDIA 8604 | APMAP | chr20 | 24960505 | 3'Flank | SBS | C | A |
| INDIA 8604 | Unknown | chr10 | 105923050 | IGR | SBS | G | C |
| INDIA 8604 | Unknown | chr6 | 61269323 | IGR | SBS | A | C |
| INDIA 8604 | Unknown | chr7 | 81870405 | IGR | SBS | C | G |
| INDIA 8604 | SPAG16 | chr2 | 213383733 | Intron | SBS | C | G |
| INDIA 8604 | ENST00000701990 | chr6 | 1132648 | 3'Flank | SBS | G | C |
| INDIA 8604 | AP5B1 | chr11 | 65781618 | 5'Flank | SBS | G | C |
| INDIA 8604 | MGMT | chr10 | 129581308 | Intron | SBS | C | G |
| INDIA 8604 | MEF2C | chr5 | 88769281 | Intron | SBS | C | G |
| INDIA 8604 | Unknown | chr5 | 115641483 | IGR | SBS | C | G |
| INDIA 8604 | RPTOR | chr17 | 80619007 | Intron | SBS | C | G |
| INDIA 8604 | CPNE9 | chr3 | 9716312 | Intron | SBS | G | C |
| INDIA 8604 | ENST00000487840 | chr3 | 150116743 | Intron | SBS | G | C |
| INDIA 8604 | TGFBR2 | chr3 | 30665681 | Intron | SBS | C | G |
| INDIA 8604 | ATP10A | chr15 | 25812742 | Intron | SBS | G | C |
| INDIA 8604 | Unknown | chr18 | 51255322 | IGR | SBS | C | G |
| INDIA 8604 | Unknown | chr18 | 53875331 | IGR | SBS | G | C |
| INDIA 8604 | ENST00000664519 | chrX | 142130740 | Intron | SBS | T | A |
| INDIA 8604 | FFAR4 | chr10 | 93587169 | Intron | SBS | C | G |
| INDIA 8604 | EFCAB6 | chr22 | 43774334 | Intron | SBS | G | C |
| INDIA 8604 | RGS7 | chr1 | 240801731 | Intron | SBS | C | G |
| INDIA 8604 | ADAMTS12 | chr5 | 33789599 | Intron | SBS | C | G |
| INDIA 8604 | KHDRBS2-OT1 | chr6 | 61642080 | Intron | SBS | G | C |
| INDIA 8604 | MAML2 | chr11 | 96195540 | Intron | DEL | G | - |
| INDIA 8604 | LINC02694 | chr15 | 38639693 | Intron | DEL | C | - |

| | | | | | | | |
|---|---|---|---|---|---|---|---|
| INDIA 8604 | PMM2 | chr16 | 8808068 | Intron | SBS | C | G |
| INDIA 8604 | LINC02911 | chr16 | 52051600 | Intron | SBS | G | C |
| INDIA 8604 | ADAMTS10 | chr19 | 8608520 | Intron | SBS | C | G |
| INDIA 8605 | GRIN2A | chr16 | 9779860 | Intron | DEL | C | - |
| INDIA 8605 | Unknown | chr5 | 168278854 | IGR | SBS | C | G |
| INDIA 8605 | LINC01177 | chr16 | 9439852 | 3'Flank | SBS | C | G |
| INDIA 8605 | JPT1 | chr17 | 75153478 | Intron | INS | - | CAG |
| INDIA 8605 | REEP3 | chr10 | 63521612 | Intron | SBS | G | C |
| INDIA 8605 | MRTFB | chr16 | 14136421 | Intron | SBS | G | C |
| INDIA 8605 | PPIG | chr2 | 169586706 | Intron | SBS | C | G |
| INDIA 8605 | Unknown | chrX | 151753586 | IGR | SBS | G | C |
| INDIA 8605 | WDFY4 | chr10 | 48792961 | Intron | SBS | A | T |
| INDIA 8605 | CDKN2B-AS1 | chr9 | 22124748 | Intron | SBS | C | G |
| INDIA 8605 | TMEM174 | chr5 | 73176310 | 3'Flank | SBS | A | T |
| INDIA 8605 | CPLX2 | chr5 | 175832674 | Intron | SBS | G | C |
| INDIA 8605 | CDKAL1 | chr6 | 21015075 | Intron | SBS | C | G |
| INDIA 8605 | Unknown | chr18 | 78191240 | IGR | SBS | A | C |
| INDIA 8605 | TENM2 | chr5 | 167566763 | Intron | SBS | G | C |
| INDIA 8605 | Unknown | chr8 | 135781587 | IGR | SBS | G | C |
| INDIA 8605 | ARMH3 | chr10 | 101965538 | Intron | SBS | C | G |
| INDIA 8605 | TMEM171 | chr5 | 73133377 | 3'Flank | SBS | G | C |
| INDIA 8605 | Unknown | chr20 | 60662909 | IGR | DEL | C | - |
| INDIA 8605 | Unknown | chr10 | 117223340 | IGR | SBS | G | T |
| INDIA 8605 | Unknown | chr15 | 55115310 | IGR | SBS | G | C |
| INDIA 8605 | DISP3 | chr1 | 11519615 | Intron | SBS | G | C |
| INDIA 8605 | GRM1 | chr6 | 146341088 | Intron | DEL | C | - |
| INDIA 8605 | RPTOR | chr17 | 80913911 | Intron | SBS | C | G |
| INDIA 8605 | FANCA | chr16 | 89805212 | Intron | SBS | G | C |
| INDIA 8605 | ATP6V0A4 | chr7 | 138759430 | Intron | SBS | C | G |
| INDIA 8605 | PKN2-AS1 | chr1 | 88192011 | Intron | SBS | G | C |
| INDIA 8605 | ENST00000641979 | chr1 | 11321807 | 5'Flank | SBS | C | G |
| INDIA 8605 | Unknown | chr4 | 187130402 | IGR | SBS | C | G |
| INDIA 8605 | FAT3 | chr11 | 92780811 | Intron | SBS | C | G |
| INDIA 8605 | ENST00000671933 | chr4 | 25552795 | Intron | SBS | C | G |
| INDIA 8605 | ENST00000565523 | chr16 | 35146809 | 3'Flank | SBS | C | G |
| INDIA 8605 | FBN1 | chr15 | 48478084 | Intron | SBS | C | G |
| INDIA 8605 | ENST00000577437 | chrX | 152128070 | 3'Flank | SBS | C | G |
| INDIA 8605 | Unknown | chr8 | 133630980 | IGR | SBS | C | G |
| INDIA 8605 | Unknown | chr5 | 130979130 | IGR | DEL | T | - |
| INDIA 8605 | SORCS2 | chr4 | 7214020 | Intron | DEL | C | - |
| INDIA 8605 | H2BC3 | chr6 | 26046005 | 5'Flank | SBS | G | C |
| INDIA 8605 | ERGIC2 | chr12 | 29382637 | 5'Flank | SBS | G | C |
| INDIA 8605 | KLHL32 | chr6 | 97113379 | Intron | SBS | T | G |
| INDIA 8605 | Unknown | chr6 | 167452186 | IGR | SBS | G | C |
| INDIA 8605 | ANO3 | chr11 | 26328490 | 5'Flank | SBS | C | A |
| INDIA 8605 | Unknown | chr11 | 104389316 | IGR | SBS | A | T |
| INDIA 8605 | ENST00000593558 | chr19 | 6950108 | 5'Flank | SBS | G | C |
| INDIA 8605 | ENST00000649978 | chr2 | 40618690 | Intron | SBS | C | G |
| INDIA 8605 | Unknown | chr3 | 174348792 | IGR | SBS | G | C |
| INDIA 8605 | Unknown | chr2 | 81690988 | IGR | SBS | C | G |
| INDIA 8605 | Unknown | chr2 | 56545880 | IGR | DEL | G | - |
| INDIA 8605 | SLC35F3 | chr1 | 233914993 | Intron | SBS | G | C |
| INDIA 8605 | LARGE1 | chr22 | 33189336 | Intron | SBS | T | A |
| INDIA 8605 | CYRIA | chr2 | 16546332 | 3'Flank | SBS | C | G |
| INDIA 8605 | Unknown | chr2 | 222361894 | IGR | SBS | G | C |
| INDIA 8605 | AK8 | chr9 | 132848448 | Intron | SBS | G | C |
| INDIA 8605 | Unknown | chr16 | 13841105 | IGR | SBS | C | G |
| INDIA 8605 | MAML2 | chr11 | 96140616 | Intron | SBS | G | C |
| INDIA 8605 | Unknown | chr3 | 13788616 | IGR | SBS | G | C |
| INDIA 8605 | Unknown | chr5 | 176517565 | IGR | SBS | G | C |
| INDIA 8605 | PIEZO2 | chr18 | 10937467 | Intron | SBS | G | C |
| INDIA 8605 | TCERG1L | chr10 | 131146081 | Intron | SBS | C | G |
| INDIA 8605 | Unknown | chr4 | 180465341 | IGR | SBS | G | C |
| INDIA 8605 | LINC02646 | chr10 | 130455609 | Intron | DEL | A | - |
| INDIA 8605 | CREBBP | chr16 | 3748577 | Intron | SBS | T | A |
| INDIA 8605 | ENST00000641035 | chr1 | 56374183 | Intron | SBS | C | G |

| | | | | | | | |
|---|---|---|---|---|---|---|---|
| INDIA 8605 | NQO2 | chr6 | 3019511 | Silent | SBS | C | G |
| INDIA 8605 | ENST00000654696 | chr5 | 73263281 | Intron | SBS | G | C |
| INDIA 8605 | TCAIM | chr3 | 44367141 | Intron | SBS | G | C |
| INDIA 8605 | SGIP1 | chr1 | 66645292 | Intron | SBS | A | T |
| INDIA 8605 | TTC28 | chr22 | 28206808 | Intron | SBS | G | C |
| INDIA 8605 | RAB20 | chr13 | 110538969 | Intron | SBS | G | C |
| INDIA 8605 | PTGER3 | chr1 | 70865260 | Intron | SBS | C | G |
| INDIA 8605 | Unknown | chr5 | 104015106 | IGR | SBS | T | A |
| INDIA 8605 | KDM4D | chr11 | 94998664 | Nonsense_Mutation | SBS | C | G |
| INDIA 8605 | ENST00000659127 | chr4 | 188806865 | Intron | SBS | G | C |
| INDIA 8605 | Unknown | chr9 | 81814875 | IGR | SBS | G | C |
| INDIA 8605 | RYR3 | chr15 | 33501107 | Intron | SBS | G | C |
| INDIA 8605 | DPH6-DT | chr15 | 35761048 | Intron | DEL | GCC | - |
| INDIA 8605 | ZFHX3 | chr16 | 72818326 | Intron | SBS | G | C |
| INDIA 8605 | ZNF709 | chr19 | 12513371 | Intron | SBS | G | C |
| INDIA 8605 | POU2F2 | chr19 | 42184484 | Intron | SBS | C | G |
| INDIA 8605 | CDK5RAP2 | chr9 | 120463569 | Intron | SBS | G | C |
| INDIA 8605 | YWHAH | chr22 | 31956912 | 3'UTR | SBS | C | G |
| INDIA 8605 | PPIG | chr2 | 169584885 | Intron | SBS | C | G |
| INDIA 8605 | NCAM1 | chr11 | 112973262 | Intron | SBS | G | C |
| INDIA 8605 | Unknown | chr3 | 187439448 | IGR | SBS | G | C |
| INDIA 8605 | TG | chr8 | 133091513 | Intron | SBS | G | C |
| INDIA 8605 | LINC01933 | chr5 | 152008493 | Intron | SBS | G | C |
| INDIA 8605 | LINC01787 | chr1 | 96366044 | Intron | DEL | T | - |
| INDIA 8605 | Unknown | chrX | 41246886 | IGR | SBS | C | G |
| INDIA 8605 | ENST00000664549 | chr10 | 6954549 | Intron | SBS | C | G |
| INDIA 8605 | Unknown | chr11 | 101380283 | IGR | SBS | G | C |
| INDIA 8605 | SPARCL1 | chr4 | 87529294 | 5'UTR | SBS | C | G |
| INDIA 8605 | Unknown | chr11 | 11254820 | IGR | SBS | G | C |
| INDIA 8605 | STAB2 | chr12 | 103683247 | Missense_Mutation | SBS | G | C |
| INDIA 8605 | HSPBP1 | chr19 | 55270346 | Intron | SBS | C | G |
| INDIA 8607 | CDH12 | chr5 | 21805630 | Intron | SBS | C | A |
| INDIA 8607 | Unknown | chr3 | 127247332 | IGR | SBS | G | C |
| INDIA 8607 | ENST00000701429 | chr6 | 105458343 | Intron | SBS | C | G |
| INDIA 8607 | Unknown | chr6 | 18326499 | IGR | SBS | T | A |
| INDIA 8607 | SRBD1 | chr2 | 45505080 | Intron | SBS | G | C |
| INDIA 8607 | Unknown | chr8 | 98301499 | IGR | SBS | C | G |
| INDIA 8607 | Unknown | chr9 | 33435923 | IGR | SBS | C | G |
| INDIA 8607 | ASIC2 | chr17 | 33142659 | Intron | SBS | A | T |
| INDIA 8607 | SHROOM4 | chrX | 50708292 | Intron | SBS | C | G |
| INDIA 8607 | LIN54 | chr4 | 82936015 | Nonsense_Mutation | SBS | G | C |
| INDIA 8607 | LINC01933 | chr5 | 151945055 | 5'Flank | DEL | G | - |
| INDIA 8607 | TSPAN9 | chr12 | 3111275 | Intron | SBS | G | C |
| INDIA 8607 | ERBB2 | chr17 | 39709724 | Intron | SBS | C | A |
| INDIA 8607 | Unknown | chr2 | 88332623 | IGR | SBS | G | C |
| INDIA 8607 | RBMS1 | chr2 | 160385569 | Intron | INS | - | AA |
| INDIA 8607 | PAPOLG | chr2 | 60806100 | 3'Flank | SBS | G | C |
| INDIA 8607 | ENST00000702169 | chr17 | 66957763 | 3'Flank | SBS | G | C |
| INDIA 8607 | EML6 | chr2 | 54725335 | Intron | SBS | C | A |
| INDIA 8607 | KAT6B | chr10 | 74907388 | Intron | SBS | G | C |
| INDIA 8607 | Unknown | chr16 | 17791936 | IGR | SBS | G | T |
| INDIA 8607 | RPIA | chr2 | 88740063 | Intron | SBS | G | C |
| INDIA 8607 | ST3GAL3 | chr1 | 43835907 | Intron | DEL | TC | - |
| INDIA 8607 | FBP1 | chr9 | 94622745 | Intron | SBS | G | C |
| INDIA 8607 | Unknown | chr18 | 10230933 | IGR | DEL | G | - |
| INDIA 8607 | MAPK8IP3 | chr16 | 1760258 | Intron | SBS | C | G |
| INDIA 8607 | ERBB2 | chr17 | 39709482 | Intron | SBS | C | G |
| INDIA 8607 | SERPIND1 | chr22 | 20779430 | Frame_Shift_Del | DEL | CCCC | - |
| INDIA 8607 | Unknown | chr20 | 57415771 | IGR | SBS | G | C |
| INDIA 8607 | PBX1 | chr1 | 164699547 | Intron | SBS | C | G |
| INDIA 8607 | GALNT13 | chr2 | 154321126 | Intron | SBS | C | G |
| INDIA 8607 | CALCRL | chr2 | 187448370 | 5'Flank | SBS | C | G |
| INDIA 8607 | PITPNM3 | chr17 | 6559036 | 5'Flank | SBS | C | G |
| INDIA 8607 | ENST00000421324 | chr10 | 78464923 | Intron | SBS | C | A |
| INDIA 8607 | DGAT1 | chr8 | 144317280 | Intron | SBS | C | G |
| INDIA 8607 | NRROS | chr3 | 196641664 | Intron | SBS | G | C |

| | | | | | | | |
|---|---|---|---|---|---|---|---|
| INDIA 8607 | NRROS | chr3 | 196641665 | Intron | SBS | G | T |
| INDIA 8607 | EIF3H | chr8 | 116712072 | Intron | SBS | G | C |
| INDIA 8607 | CUBN | chr10 | 17006361 | Intron | SBS | G | C |
| INDIA 8607 | NXF1 | chr11 | 62804463 | Intron | SBS | C | G |
| INDIA 8607 | CCDC88C | chr14 | 91336661 | Intron | SBS | G | T |
| INDIA 8607 | SEH1L | chr18 | 12947264 | 5'Flank | SBS | C | G |
| INDIA 8607 | PCIF1 | chr20 | 45944544 | Intron | DEL | ATC | - |
| INDIA 8607 | RGS12 | chr4 | 3430484 | Missense_Mutation | SBS | G | C |
| INDIA 8607 | HSD17B2 | chr16 | 82035725 | Intron | DEL | TGATC | - |
| INDIA 8607 | NXF1 | chr11 | 62803850 | Missense_Mutation | SBS | C | G |
| INDIA 8607 | NCK2 | chr2 | 105855565 | Intron | SBS | G | C |
| INDIA 8607 | C8orf33 | chr8 | 145058014 | 3'Flank | SBS | C | G |
| INDIA 8607 | TEDC1 | chr14 | 105496942 | Intron | SBS | G | T |
| INDIA 8607 | MAPK8IP2 | chr22 | 50607384 | Intron | SBS | C | G |
| INDIA 8607 | CAPN12 | chr19 | 38731807 | Intron | SBS | C | G |
| INDIA 8607 | RTN4RL1 | chr17 | 2022691 | Intron | SBS | C | G |
| INDIA 8607 | LGALS9 | chr17 | 27646420 | Intron | SBS | C | G |
| INDIA 8607 | Unknown | chr2 | 42973827 | IGR | SBS | G | T |
| INDIA 8607 | SAXO1 | chr9 | 18938783 | Intron | SBS | G | C |
| INDIA 8607 | ENST00000651161 | chr19 | 19749803 | RNA | SBS | G | C |
| INDIA 8607 | RTN4RL1 | chr17 | 2022672 | Intron | SBS | C | G |
| INDIA 8607 | FIGNL2 | chr12 | 51847167 | Intron | SBS | G | C |
| INDIA 8607 | PLEKHO1 | chr1 | 150162863 | 3'Flank | SBS | G | C |
| INDIA 8607 | LINC02381 | chr12 | 54142943 | Intron | DEL | AA | - |
| INDIA 8607 | DOCK4 | chr7 | 112005862 | Intron | SBS | G | C |
| INDIA 8607 | MICAL2 | chr11 | 12179051 | Intron | SBS | G | C |
| INDIA 8607 | MZF1 | chr19 | 58570801 | Intron | DEL | TGCAGG | - |
| INDIA 8607 | IZUMO1 | chr19 | 48745710 | Missense_Mutation | SBS | G | C |
| INDIA 8607 | ZFAND2B | chr2 | 219206923 | 5'UTR | SBS | G | C |
| INDIA 8607 | P4HA2 | chr5 | 132279438 | Intron | DEL | G | - |
| INDIA 8607 | LINC01805 | chr2 | 64503109 | 3'Flank | SBS | C | A |
| INDIA 8607 | Unknown | chr6 | 115883556 | IGR | INS | - | T |
| INDIA 8607 | TFIP11 | chr22 | 26497096 | Intron | DEL | CAGTCTC | - |
| INDIA 8607 | CNNM2 | chr10 | 102919395 | Missense_Mutation | SBS | C | G |
| INDIA 8607 | DPAGT1 | chr11 | 119101852 | 5'UTR | SBS | C | G |
| INDIA 8607 | PPM1N | chr19 | 45500666 | Missense_Mutation | SBS | G | C |
| INDIA 8607 | FOS | chr14 | 75280286 | Intron | SBS | C | G |
| INDIA 8607 | NEK6 | chr9 | 124294565 | Intron | SBS | C | G |
| INDIA 8607 | CELF6 | chr15 | 72318860 | Intron | SBS | C | G |
| INDIA 8607 | ARPC3 | chr12 | 110431526 | 3'Flank | SBS | G | T |
| INDIA 8607 | Unknown | chr1 | 39932288 | IGR | SBS | C | A |
| INDIA 8607 | DIP2B | chr12 | 50555722 | Intron | DEL | CA | - |
| INDIA 8607 | LGI4 | chr19 | 35134765 | 5'UTR | SBS | G | T |
| INDIA 8607 | ATG101 | chr12 | 52077778 | 3'Flank | DEL | AG | - |
| INDIA 8607 | LINC01220 | chr14 | 75296443 | RNA | DEL | G | - |
| INDIA 8607 | DNAAF2 | chr14 | 49634190 | Silent | SBS | G | C |
| INDIA 8607 | ENST00000668450 | chr2 | 64838372 | Intron | DEL | CG | - |
| INDIA 8608 | DSG3 | chr18 | 31474307 | Missense_Mutation | SBS | C | G |
| INDIA 8608 | CSTPP1 | chr11 | 47109527 | Intron | SBS | G | T |
| INDIA 8608 | DPYD | chr1 | 97348511 | Intron | SBS | T | G |
| INDIA 8608 | Unknown | chr20 | 55000607 | IGR | SBS | G | C |
| INDIA 8608 | ZNF71 | chr19 | 56617818 | Intron | SBS | C | G |
| INDIA 8608 | Unknown | chr12 | 14015540 | IGR | INS | - | A |
| INDIA 8608 | LINC02580 | chr2 | 43154884 | Intron | DEL | TC | - |
| INDIA 8608 | WAC | chr10 | 28571599 | Intron | SBS | A | C |
| INDIA 8608 | EPHB1 | chr3 | 135261413 | 3'Flank | SBS | G | C |
| INDIA 8608 | Unknown | chr6 | 15927653 | IGR | SBS | G | T |
| INDIA 8608 | TMEM178B | chr7 | 141327959 | Intron | SBS | C | G |
| INDIA 8608 | ZFPM2 | chr8 | 104802088 | Intron | INS | - | CAT |
| INDIA 8608 | ENST00000650712 | chr2 | 239809221 | Intron | DEL | CTG | - |
| INDIA 8608 | CSMD1 | chr8 | 4185570 | Intron | SBS | C | G |
| INDIA 8608 | RICTOR | chr5 | 39076805 | 5'Flank | DEL | G | - |
| INDIA 8608 | OR2S2 | chr9 | 35961137 | 5'Flank | INS | - | T |
| INDIA 8608 | Unknown | chr5 | 63675851 | IGR | SBS | C | G |
| INDIA 8608 | FAM184B | chr4 | 17752543 | Intron | SBS | C | G |
| INDIA 8608 | Unknown | chr3 | 100588481 | IGR | SBS | A | T |

| | | | | | | | |
|---|---|---|---|---|---|---|---|
| INDIA 8608 | GZMK | chr5 | 55027419 | Intron | SBS | C | A |
| INDIA 8608 | SPACA1 | chr6 | 88047478 | 5'Flank | SBS | A | T |
| INDIA 8608 | Unknown | chr3 | 84376586 | IGR | DEL | A | - |
| INDIA 8608 | PKD1L1 | chr7 | 47833701 | Intron | SBS | C | A |
| INDIA 8608 | KIRREL3 | chr11 | 126774626 | Intron | SBS | T | A |
| INDIA 8608 | Unknown | chr20 | 55874994 | IGR | SBS | C | G |
| INDIA 8608 | SPA17 | chr11 | 124696564 | 3'UTR | DEL | TAT | - |
| INDIA 8608 | LIX1-AS1 | chr5 | 97427068 | Intron | SBS | A | T |
| INDIA 8608 | Unknown | chr12 | 103006503 | IGR | SBS | C | G |
| INDIA 8608 | TNKS | chr8 | 9668783 | Intron | SBS | G | C |
| INDIA 8608 | PPP1R3B-DT | chr8 | 9303098 | Intron | DEL | CAC | - |
| INDIA 8608 | Unknown | chr2 | 7635826 | IGR | SBS | C | A |
| INDIA 8608 | MIR4432HG | chr2 | 60356061 | 3'Flank | SBS | T | G |
| INDIA 8608 | LINC00615 | chr12 | 90919547 | Intron | SBS | A | T |
| INDIA 8608 | TTC6 | chr14 | 37611365 | Intron | DEL | A | - |
| INDIA 8608 | SCN2A | chr2 | 165293764 | Intron | SBS | A | C |
| INDIA 8608 | AGAP1 | chr2 | 235747134 | Intron | SBS | G | T |
| INDIA 8608 | GLTP | chr12 | 109858350 | Intron | DEL | AGA | - |
| INDIA 8608 | Unknown | chr19 | 31072659 | IGR | DEL | GA | - |
| INDIA 8608 | DENND1A | chr9 | 123570490 | Intron | DEL | T | - |
| INDIA 8608 | SMARCA4 | chr19 | 11020022 | Intron | SBS | G | C |
| INDIA 8608 | ZFYVE26 | chr14 | 67753009 | Intron | SBS | C | A |
| INDIA 8608 | DRC1 | chr2 | 26441801 | Intron | SBS | T | G |
| INDIA 8608 | Unknown | chr12 | 114916334 | IGR | DEL | GCT | - |
| INDIA 8608 | COX7A2L | chr2 | 42368103 | Intron | SBS | C | G |
| INDIA 8608 | CADPS2 | chr7 | 122387064 | Frame_Shift_Del | DEL | GCTTTTTG | - |
| INDIA 8608 | ABCC4 | chr13 | 95207961 | Intron | SBS | G | C |
| INDIA 8608 | ABAT | chr16 | 8782581 | 3'UTR | INS | - | CT |
| INDIA 8608 | PTPN1 | chr20 | 50509992 | 5'Flank | SBS | A | T |
| INDIA 8608 | PTPN1 | chr20 | 50509993 | 5'Flank | SBS | C | G |
| INDIA 8608 | PCSK5 | chr9 | 75912887 | Intron | DEL | TCCCCA | - |
| INDIA 8608 | ANO2 | chr12 | 5605901 | Intron | SBS | C | G |
| INDIA 8608 | Unknown | chr5 | 132028540 | IGR | SBS | T | G |
| INDIA 8608 | TMEM178B | chr7 | 141330049 | Intron | SBS | C | G |
| INDIA 8608 | ZNF804A | chr2 | 184835758 | Intron | SBS | T | A |
| INDIA 8608 | NSMCE2 | chr8 | 125345726 | Intron | SBS | C | G |
| INDIA 8608 | KIAA0408 | chr6 | 127453920 | Frame_Shift_Del | DEL | AA | - |
| INDIA 8608 | PRKG1 | chr10 | 51604320 | Intron | SBS | G | C |
| INDIA 8608 | LINC02715 | chr11 | 109757765 | Intron | SBS | C | G |
| INDIA 8608 | Unknown | chr5 | 114730130 | IGR | SBS | G | C |
| INDIA 8608 | ARL4C | chr2 | 234492027 | 3'Flank | DEL | A | - |
| INDIA 8608 | PKIG | chr20 | 44547153 | Intron | DEL | TAT | - |
| INDIA 8608 | ATP10A | chr15 | 25763408 | Intron | DEL | CCAGACTCA | - |
| INDIA 8608 | SLC66A1L | chr3 | 157554625 | Intron | SBS | G | C |
| INDIA 8608 | Unknown | chr8 | 137803727 | IGR | SBS | C | G |
| INDIA 8609 | SYTL5 | chrX | 38107986 | Intron | SBS | G | C |
| INDIA 8609 | ENST00000669987 | chr9 | 36791051 | Intron | SBS | C | G |
| INDIA 8609 | OR3A2 | chr17 | 3343677 | Intron | SBS | G | C |
| INDIA 8609 | STX8 | chr17 | 9461302 | Intron | SBS | G | C |
| INDIA 8609 | Unknown | chr21 | 41602675 | IGR | SBS | C | G |
| INDIA 8609 | Unknown | chr9 | 117637753 | IGR | SBS | T | G |
| INDIA 8609 | EXD3 | chr9 | 137317289 | Intron | SBS | C | G |
| INDIA 8609 | Unknown | chrX | 100430903 | IGR | INS | - | T |
| INDIA 8609 | ANKRD33B | chr5 | 10651455 | 3'UTR | DEL | T | - |
| INDIA 8609 | Unknown | chr9 | 80421289 | IGR | SBS | C | A |
| INDIA 8609 | PIP5K1B | chr9 | 68714711 | Intron | SBS | C | G |
| INDIA 8609 | Unknown | chr14 | 85104607 | IGR | SBS | G | C |
| INDIA 8609 | ENST00000692740 | chr2 | 175697262 | Intron | SBS | G | C |
| INDIA 8609 | CACNA1B | chr9 | 137876036 | 5'Flank | SBS | C | G |
| INDIA 8609 | Unknown | chr18 | 77168684 | IGR | SBS | G | C |
| INDIA 8609 | LINC02208 | chr5 | 118451310 | Intron | SBS | G | C |
| INDIA 8609 | UNC5D | chr8 | 35502406 | Intron | SBS | G | C |
| INDIA 8609 | Unknown | chr5 | 136794564 | IGR | SBS | G | C |
| INDIA 8609 | PSMA2 | chr7 | 42916912 | 3'UTR | SBS | G | C |
| INDIA 8609 | POLR2M | chr15 | 57710909 | Intron | SBS | T | A |
| INDIA 8609 | CELF2 | chr10 | 11201485 | Intron | SBS | G | C |

| | | | | | | | |
|---|---|---|---|---|---|---|---|
| INDIA 8609 | SCNN1G | chr16 | 23191280 | Intron | SBS | G | C |
| INDIA 8609 | ABCA9 | chr17 | 68975920 | Missense_Mutation | SBS | G | C |
| INDIA 8609 | GALNT9 | chr12 | 132254468 | Intron | SBS | G | C |
| INDIA 8609 | RPS6KA2 | chr6 | 166484733 | Intron | SBS | C | G |
| INDIA 8609 | NCKAP1 | chr2 | 183037509 | Intron | SBS | C | G |
| INDIA 8609 | NDUFA10 | chr2 | 240018833 | Intron | SBS | G | C |
| INDIA 8609 | TLN2 | chr15 | 62405173 | Intron | DEL | ATTCAGGG | - |
| INDIA 8609 | SCP2 | chr1 | 53049158 | Intron | SBS | G | C |
| INDIA 8609 | GRM7 | chr3 | 7368126 | Intron | SBS | G | C |
| INDIA 8609 | ADAMTSL3 | chr15 | 83785086 | Intron | SBS | A | T |
| INDIA 8609 | CSMD3 | chr8 | 112483932 | Intron | SBS | G | C |
| INDIA 8609 | MARCHF1 | chr4 | 163612151 | Intron | SBS | C | G |
| INDIA 8609 | Unknown | chr8 | 48779959 | IGR | SBS | C | G |
| INDIA 8609 | Unknown | chr20 | 46528134 | IGR | SBS | G | C |
| INDIA 8609 | ENTPD4 | chr8 | 23454974 | Intron | SBS | A | T |
| INDIA 8609 | KIRREL3 | chr11 | 126543172 | Intron | SBS | C | G |
| INDIA 8609 | GGACT | chr13 | 100552453 | Intron | SBS | G | C |
| INDIA 8609 | Unknown | chr17 | 56711206 | IGR | SBS | T | A |
| INDIA 8609 | SLC4A7 | chr3 | 27409503 | Missense_Mutation | SBS | G | C |
| INDIA 8609 | Unknown | chr8 | 21325616 | IGR | SBS | G | C |
| INDIA 8609 | NIBAN1 | chr1 | 184827451 | Intron | SBS | A | C |
| INDIA 8609 | MMADHC-DT | chr2 | 149848572 | Intron | SBS | G | C |
| INDIA 8609 | Unknown | chr2 | 35351406 | IGR | SBS | C | G |
| INDIA 8609 | LINC02261 | chr4 | 27268378 | Intron | SBS | C | G |
| INDIA 8609 | Unknown | chr9 | 26535194 | IGR | SBS | G | T |
| INDIA 8609 | ACHE | chr7 | 100888252 | 3'Flank | SBS | C | G |
| INDIA 8609 | DLGAP1 | chr18 | 4427548 | Intron | SBS | C | G |
| INDIA 8609 | KIRREL3 | chr11 | 126756902 | Intron | SBS | C | G |
| INDIA 8609 | Unknown | chr10 | 56087284 | IGR | SBS | T | A |
| INDIA 8609 | GLRA3 | chr4 | 174674841 | Intron | SBS | C | G |
| INDIA 8609 | KAT6A | chr8 | 41951906 | Intron | SBS | G | C |
| INDIA 8609 | ADCK1 | chr14 | 77895084 | Intron | SBS | C | G |
| INDIA 8609 | FHOD3 | chr18 | 36760974 | Intron | SBS | C | G |
| INDIA 8609 | ENST00000641892 | chr1 | 18021472 | Intron | SBS | C | G |
| INDIA 8609 | ARHGAP15 | chr2 | 143451768 | Intron | SBS | C | G |
| INDIA 8609 | EYS | chr6 | 64045017 | Intron | SBS | C | G |
| INDIA 8609 | Unknown | chr10 | 84839411 | IGR | SBS | C | G |
| INDIA 8609 | FHIT | chr3 | 59804225 | Intron | SBS | C | G |
| INDIA 8609 | GABRB1 | chr4 | 47038255 | Intron | SBS | T | A |
| INDIA 8609 | Unknown | chr5 | 132032375 | IGR | SBS | G | C |
| INDIA 8609 | ZNF880 | chr19 | 52383784 | Intron | SBS | C | G |
| INDIA 8609 | POC1A | chr3 | 52076276 | Intron | SBS | C | G |
| INDIA 8609 | TG | chr8 | 132894162 | Intron | SBS | G | C |
| INDIA 8609 | ADCY5 | chr3 | 123319776 | Missense_Mutation | SBS | C | G |
| INDIA 8609 | Unknown | chr8 | 77206607 | IGR | SBS | G | C |
| INDIA 8609 | Unknown | chr13 | 28745846 | IGR | SBS | G | C |
| INDIA 8609 | CD55 | chr1 | 207353786 | Intron | SBS | C | G |
| INDIA 8609 | PEX1 | chr7 | 92490833 | Intron | SBS | A | C |
| INDIA 8609 | CDH4 | chr20 | 61460023 | Intron | SBS | G | C |
| INDIA 8609 | Unknown | chr11 | 91908112 | IGR | SBS | A | T |
| INDIA 8609 | PRDM13 | chr6 | 99618378 | 3'Flank | SBS | C | G |
| INDIA 8609 | Unknown | chr5 | 166564713 | IGR | SBS | T | A |
| INDIA 8609 | ENST00000439455 | chr1 | 94143489 | 5'Flank | SBS | G | C |
| INDIA 8609 | KIT | chr4 | 54731527 | Intron | SBS | C | G |
| INDIA 8609 | TNPO3 | chr7 | 128980249 | Intron | SBS | C | G |
| INDIA 8609 | GATA3 | chr10 | 8076110 | 3'Flank | INS | - | TC |
| INDIA 8609 | WWC2 | chr4 | 183323418 | 3'Flank | SBS | C | G |
| INDIA 8609 | BCAR3 | chr1 | 93781924 | Intron | SBS | G | C |
| INDIA 8609 | ST7L | chr1 | 112547040 | Intron | SBS | G | C |
| INDIA 8609 | ENST00000664948 | chr18 | 75476883 | 3'Flank | SBS | G | C |
| INDIA 8609 | SCN1A | chr2 | 166086974 | Intron | SBS | G | C |
| INDIA 8609 | ENST00000658087 | chr2 | 106953835 | Intron | SBS | C | G |
| INDIA 8609 | MIR4778 | chr2 | 66354458 | 3'Flank | SBS | C | G |
| INDIA 8609 | OLFM3 | chr1 | 101847039 | Intron | SBS | G | T |
| INDIA 8609 | Unknown | chr5 | 67985842 | IGR | SBS | G | C |
| INDIA 8609 | GRIA1 | chr5 | 153630380 | Intron | SBS | G | C |

| | | | | | | | |
|---|---|---|---|---|---|---|---|
| INDIA 8609 | ENST00000584611 | chr18 | 21375640 | 3'Flank | SBS | G | C |
| INDIA 8609 | Unknown | chr20 | 43239327 | IGR | SBS | G | C |
| INDIA 8609 | LINC02934 | chr2 | 65768018 | Intron | DEL | CTC | - |
| INDIA 8609 | MYH15 | chr3 | 108511774 | 5'Flank | SBS | C | G |
| INDIA 8609 | CENPE | chr4 | 103179348 | Intron | SBS | C | G |
| INDIA 8610 | Unknown | chr11 | 69892552 | IGR | SBS | G | C |
| INDIA 8610 | GALNT17 | chr7 | 71579127 | Intron | DEL | G | - |
| INDIA 8610 | CFAP77 | chr9 | 132441733 | Intron | SBS | A | T |
| INDIA 8610 | ENST00000419809 | chr2 | 67628624 | Intron | DEL | AGAACCA | - |
| INDIA 8610 | Unknown | chr9 | 93711341 | IGR | SBS | G | T |
| INDIA 8610 | SDC1 | chr2 | 20207387 | Intron | DEL | A | - |
| INDIA 8610 | THSD4 | chr15 | 71749466 | Intron | SBS | G | C |
| INDIA 8610 | TFAP2A | chr6 | 10415836 | 5'Flank | SBS | C | G |
| INDIA 8610 | BEX1 | chrX | 103065068 | 5'Flank | SBS | G | T |
| INDIA 8610 | ENST00000505002 | chr5 | 109469246 | 5'Flank | DEL | T | - |
| INDIA 8610 | ENST00000663591 | chr6 | 156158749 | Intron | SBS | C | G |
| INDIA 8610 | Unknown | chr10 | 26420519 | IGR | SBS | G | C |
| INDIA 8610 | Unknown | chr2 | 129027327 | IGR | INS | - | A |
| INDIA 8610 | Unknown | chr9 | 7770544 | IGR | DEL | G | - |
| INDIA 8610 | ATXN2 | chr12 | 111509334 | Intron | DEL | C | - |
| INDIA 8610 | RFLNA | chr12 | 124141128 | Intron | SBS | G | C |
| INDIA 8610 | KIF26A | chr14 | 104148412 | Intron | SBS | C | G |
| INDIA 8610 | TMEM117 | chr12 | 43836526 | Intron | INS | - | A |
| INDIA 8610 | Unknown | chr12 | 76197506 | IGR | SBS | C | G |
| INDIA 8610 | PRKCH | chr14 | 61536534 | Intron | SBS | G | C |
| INDIA 8610 | ADGRE1 | chr19 | 6902121 | Intron | SBS | C | G |
| INDIA 8610 | Unknown | chr4 | 181336400 | IGR | SBS | C | G |
| INDIA 8610 | TENM3-AS1 | chr4 | 182126403 | Intron | SBS | A | C |
| INDIA 8610 | SPOCK1 | chr5 | 137400484 | Intron | SBS | C | G |
| INDIA 8610 | CACNA2D1 | chr7 | 82073226 | Intron | SBS | C | G |
| INDIA 8610 | C13orf42 | chr13 | 51106702 | Intron | SBS | G | C |
| INDIA 8610 | ENST00000624591 | chr19 | 29922632 | 3'Flank | SBS | C | G |
| INDIA 8610 | Unknown | chr2 | 120577000 | IGR | SBS | C | G |
| INDIA 8610 | ENST00000652330 | chr6 | 126792922 | Intron | DEL | G | - |
| INDIA 8610 | CCNJL | chr5 | 160294972 | Intron | DEL | A | - |
| INDIA 8610 | HECA | chr6 | 139131410 | 5'Flank | DEL | GGTGTA | - |
| INDIA 8610 | GALNT16 | chr14 | 69314468 | Intron | SBS | G | C |
| INDIA 8610 | ENST00000533459 | chr11 | 103862262 | Intron | SBS | C | G |
| INDIA 8610 | LINC02775 | chr1 | 214147042 | Intron | DEL | C | - |
| INDIA 8610 | DAB1 | chr1 | 58157037 | Intron | DEL | TGGTGATTT | - |
| INDIA 8610 | MS4A6A | chr11 | 60176926 | Intron | SBS | G | T |
| INDIA 8610 | JPH4 | chr14 | 23582307 | 5'Flank | INS | - | G |
| INDIA 8610 | Unknown | chr11 | 133988494 | IGR | SBS | C | G |
| INDIA 8610 | CRPPA | chr7 | 16212656 | Intron | SBS | T | A |
| INDIA 8610 | CMTM7 | chr3 | 32421431 | Intron | SBS | C | G |
| INDIA 8610 | FAM76A | chr1 | 27725845 | 5'Flank | DEL | GT | - |
| INDIA 8610 | LINC02458 | chr12 | 89115683 | Intron | DEL | G | - |
| INDIA 8610 | Unknown | chr1 | 226215415 | IGR | SBS | C | G |
| INDIA 8610 | SLC8A1 | chr2 | 40105726 | 3'UTR | SBS | C | G |
| INDIA 8610 | GMDS | chr6 | 2111066 | Intron | DEL | C | - |
| INDIA 8610 | UMAD1 | chr7 | 7890909 | Intron | SBS | C | G |
| INDIA 8610 | Unknown | chr12 | 29998424 | IGR | SBS | A | C |
| INDIA 8610 | Unknown | chr12 | 98070053 | IGR | SBS | A | C |
| INDIA 8610 | Unknown | chr14 | 48328283 | IGR | SBS | A | T |
| INDIA 8610 | MIER2 | chr19 | 327827 | Intron | DEL | C | - |
| INDIA 8610 | Unknown | chr19 | 29795884 | IGR | SBS | G | C |
| INDIA 8610 | TTC6 | chr14 | 37851303 | Intron | SBS | G | C |
| INDIA 8610 | Unknown | chr15 | 87145137 | IGR | SBS | A | C |
| INDIA 8610 | FAM193A | chr4 | 2647652 | Intron | DEL | GAGCAG | - |
| INDIA 8610 | ST6GALNAC3 | chr1 | 76616880 | Intron | SBS | C | G |
| INDIA 8610 | ZC3H6 | chr2 | 112335068 | 3'UTR | SBS | G | C |
| INDIA 8610 | Unknown | chr18 | 78046219 | IGR | SBS | A | T |
| INDIA 8610 | Unknown | chrX | 69401533 | IGR | SBS | G | C |
| INDIA 8610 | MED15 | chr22 | 20568905 | Intron | DEL | TGAG | - |
| INDIA 8610 | Unknown | chr12 | 83310910 | IGR | SBS | G | T |
| INDIA 8610 | CRTC3 | chr15 | 90629356 | Missense_Mutation | SBS | C | G |

| | | | | | | | |
|---|---|---|---|---|---|---|---|
| INDIA 8610 | LASP1 | chr17 | 38868376 | 5'Flank | SBS | G | C |
| INDIA 8610 | DNER | chr2 | 229433401 | Intron | DEL | CTAC | - |
| INDIA 8610 | KCTD8 | chr4 | 44241116 | Intron | SBS | C | G |
| INDIA 8610 | ENST00000659958 | chr17 | 33876895 | Intron | SBS | G | C |
| INDIA 8610 | NIN | chr14 | 50825828 | Intron | SBS | A | C |
| INDIA 8610 | EYA3 | chr1 | 27997660 | Intron | DEL | CTTACA | - |
| INDIA 8610 | MKNK1 | chr1 | 46563235 | Intron | SBS | A | C |
| INDIA 8610 | LINC02607 | chr1 | 95550685 | Intron | SBS | G | C |
| INDIA 8610 | LMX1A | chr1 | 165315630 | Intron | SBS | C | A |
| INDIA 8610 | GALNTL6 | chr4 | 172352975 | Intron | SBS | G | T |
| INDIA 8610 | TENM3 | chr4 | 182303570 | Intron | SBS | T | G |
| INDIA 8610 | Unknown | chr13 | 63351772 | IGR | SBS | G | C |
| INDIA 8610 | MYO9B | chr19 | 17152854 | Intron | SBS | C | G |
| INDIA 8610 | SLC26A9 | chr1 | 205914671 | 3'UTR | SBS | G | C |
| INDIA 8610 | ETS1 | chr11 | 128489119 | Intron | DEL | CTAAGAGTA( | - |
| INDIA 8610 | Unknown | chr3 | 22522011 | IGR | SBS | G | C |
| INDIA 8610 | Unknown | chr16 | 54432370 | IGR | SBS | G | C |
| INDIA 8610 | VMP1 | chr17 | 59756605 | Intron | SBS | C | G |
| INDIA 8610 | EXOC4 | chr7 | 133855652 | Intron | SBS | C | G |
| INDIA 8610 | UNC45B | chr17 | 35166631 | Intron | DEL | C | - |
| INDIA 8610 | Unknown | chr2 | 166759529 | IGR | SBS | A | T |
| INDIA 8610 | ENST00000420000 | chr3 | 3469129 | Intron | DEL | GTCGCCT | - |
| INDIA 8610 | Unknown | chr20 | 56723993 | IGR | SBS | G | C |
| INDIA 8610 | ENST00000701907 | chr12 | 3961888 | 5'Flank | SBS | C | G |
| INDIA 8610 | ENST00000654829 | chr18 | 55608094 | Intron | DEL | C | - |
| INDIA 8610 | UCK2 | chr1 | 165828507 | Intron | DEL | T | - |
| INDIA 8610 | Unknown | chr12 | 5102597 | IGR | DEL | T | - |
| INDIA 8611 | CLCN5 | chrX | 50075689 | Intron | SBS | G | C |
| INDIA 8611 | ENST00000653599 | chrX | 96313924 | Intron | DEL | AATGT | - |
| INDIA 8611 | NALF2 | chrX | 69531363 | 3'UTR | SBS | G | C |
| INDIA 8611 | PLXNA4 | chr7 | 132149308 | Intron | DEL | C | - |
| INDIA 8611 | Unknown | chr5 | 146533496 | IGR | INS | - | A |
| INDIA 8611 | CCDC7 | chr10 | 32565584 | Missense_Mutation | SBS | C | G |
| INDIA 8611 | ENST00000415387 | chr2 | 212979906 | Intron | SBS | A | C |
| INDIA 8611 | GJA8 | chr1 | 147898063 | 5'Flank | SBS | A | T |
| INDIA 8611 | ENST00000635130 | chr20 | 22371934 | Intron | SBS | A | T |
| INDIA 8611 | Unknown | chr12 | 39523755 | IGR | SBS | T | G |
| INDIA 8611 | TMEM132D | chr12 | 129374956 | Intron | DEL | C | - |
| INDIA 8611 | NIPAL1 | chr4 | 48024627 | Intron | DEL | TCTC | - |
| INDIA 8611 | ACTBL2 | chr5 | 57486345 | 5'Flank | SBS | C | G |
| INDIA 8611 | Unknown | chr2 | 180155196 | IGR | SBS | G | T |
| INDIA 8611 | Unknown | chr9 | 119486955 | IGR | SBS | G | C |
| INDIA 8611 | RASGRF1 | chr15 | 79047675 | Intron | SBS | G | C |
| INDIA 8611 | Unknown | chrX | 18226188 | IGR | SBS | G | C |
| INDIA 8611 | Unknown | chr5 | 3695114 | IGR | SBS | G | C |
| INDIA 8611 | ENST00000573220 | chr16 | 4177134 | 3'Flank | SBS | G | C |
| INDIA 8611 | ENST00000473299 | chr3 | 146986658 | Intron | DEL | ATG | - |
| INDIA 8611 | GPC5 | chr13 | 92213147 | Intron | SBS | C | G |
| INDIA 8611 | Unknown | chr9 | 116020043 | IGR | SBS | G | C |
| INDIA 8611 | OPCML | chr11 | 133254466 | Intron | SBS | A | T |
| INDIA 8611 | NDNF | chr4 | 121057811 | Intron | SBS | C | G |
| INDIA 8611 | SORCS3 | chr10 | 105246164 | Intron | SBS | C | G |
| INDIA 8611 | ENST00000664648 | chr20 | 1048455 | Intron | SBS | G | C |
| INDIA 8611 | POLDIP3 | chr22 | 42602038 | Frame_Shift_Ins | INS | - | G |
| INDIA 8611 | MGAT4C | chr12 | 86371479 | Intron | SBS | G | C |
| INDIA 8611 | ROBO1 | chr3 | 79082590 | Intron | SBS | T | A |
| INDIA 8611 | ENST00000655342 | chr13 | 39093498 | Intron | SBS | G | C |
| INDIA 8611 | LNC-LBCS | chr6 | 19422806 | Intron | SBS | G | C |
| INDIA 8611 | SORCS2 | chr4 | 7664243 | Intron | DEL | G | - |
| INDIA 8611 | COPS9 | chr2 | 240127843 | 3'Flank | SBS | G | C |
| INDIA 8611 | LINC02955 | chr12 | 23040350 | Intron | DEL | G | - |
| INDIA 8611 | B3GNT2 | chr2 | 62197895 | Intron | INS | - | AG |
| INDIA 8611 | PLXNA4 | chr7 | 132401591 | Intron | SBS | A | C |
| INDIA 8611 | Unknown | chr17 | 54793499 | IGR | SBS | G | C |
| INDIA 8611 | THSD4 | chr15 | 71181985 | Intron | DEL | C | - |
| INDIA 8611 | LRBA | chr4 | 150354445 | Intron | SBS | C | G |

| | | | | | | | |
|---|---|---|---|---|---|---|---|
| INDIA 8611 | SLC38A2 | chr12 | 46367596 | Intron | DEL | CAAA | - |
| INDIA 8611 | ETV6 | chr12 | 11723650 | Intron | SBS | C | G |
| INDIA 8611 | Unknown | chr4 | 179843536 | IGR | DEL | CA | - |
| INDIA 8611 | ABCA3 | chr16 | 2290105 | Intron | DEL | GCCC | - |
| INDIA 8611 | LINC01376 | chr2 | 19230994 | Intron | SBS | C | G |
| INDIA 8611 | POMC | chr2 | 25165622 | Intron | SBS | C | G |
| INDIA 8611 | Unknown | chr8 | 113557703 | IGR | SBS | G | C |
| INDIA 8611 | RUNX1 | chr21 | 35371587 | 3'Flank | SBS | C | G |
| INDIA 8611 | HPYR1 | chr8 | 132524348 | Intron | SBS | G | C |
| INDIA 8611 | TRPM3 | chr9 | 70962237 | Intron | SBS | G | C |
| INDIA 8611 | TMEM104 | chr17 | 74827112 | Intron | SBS | G | C |
| INDIA 8611 | VMAC | chr19 | 5904613 | 5'Flank | SBS | G | C |
| INDIA 8611 | LTBP1 | chr2 | 33299676 | Intron | SBS | C | G |
| INDIA 8611 | ENST00000651429 | chr2 | 48066340 | Intron | SBS | G | C |
| INDIA 8611 | FLNB | chr3 | 58157991 | Intron | SBS | C | G |
| INDIA 8611 | INPP4B | chr4 | 142683565 | Intron | SBS | T | G |
| INDIA 8611 | PDE4D | chr5 | 59766691 | Intron | SBS | C | G |
| INDIA 8611 | Unknown | chr6 | 95105529 | IGR | SBS | G | C |
| INDIA 8611 | Unknown | chr7 | 125516447 | IGR | SBS | G | T |
| INDIA 8611 | NIPAL2 | chr8 | 98196514 | Intron | SBS | C | G |
| INDIA 8611 | LINC00536 | chr8 | 116131627 | Intron | DEL | C | - |
| INDIA 8611 | MLLT3 | chr9 | 20441564 | Intron | SBS | C | G |
| INDIA 8611 | Unknown | chr11 | 55766689 | IGR | SBS | A | T |
| INDIA 8611 | Unknown | chr15 | 94750241 | IGR | SBS | C | G |
| INDIA 8611 | LDLRAD4 | chr18 | 13492105 | Intron | SBS | C | G |
| INDIA 8611 | ENST00000654150 | chr18 | 37986460 | Intron | SBS | C | A |
| INDIA 8611 | IL27RA | chr19 | 14042781 | Missense_Mutation | SBS | C | G |
| INDIA 8611 | SEPTIN6 | chrX | 119663579 | Missense_Mutation | SBS | G | C |
| INDIA 8611 | Unknown | chr7 | 46351836 | IGR | DEL | TTATATAC | - |
| INDIA 8611 | PRDM5 | chr4 | 120740921 | Intron | SBS | C | G |
| INDIA 8611 | CDK6 | chr7 | 92623474 | Intron | SBS | C | G |
| INDIA 8611 | STAG1 | chr3 | 136618562 | Intron | SBS | T | A |
| INDIA 8611 | ENST00000673788 | chr21 | 14699380 | Intron | SBS | C | G |
| INDIA 8611 | ENST00000606382 | chr2 | 60023766 | Intron | DEL | G | - |
| INDIA 8611 | CAMK2G | chr10 | 73872121 | Intron | INS | - | AG |
| INDIA 8611 | DR1 | chr1 | 93350339 | Intron | DEL | C | - |
| INDIA 8611 | Unknown | chr1 | 192113688 | IGR | SBS | G | C |
| INDIA 8611 | ENST00000657641 | chr9 | 88751650 | Intron | SBS | C | G |
| INDIA 8611 | MICU3 | chr8 | 17113005 | Intron | SBS | G | C |
| INDIA 8611 | S100A10 | chr1 | 151980588 | 3'Flank | SBS | C | G |
| INDIA 8611 | LINC00578 | chr3 | 177745121 | Intron | SBS | G | C |
| INDIA 8611 | ENST00000427911 | chr21 | 32844956 | Intron | SBS | G | T |
| INDIA 8611 | LINC01250 | chr2 | 3077304 | Intron | DEL | G | - |
| INDIA 8611 | Unknown | chr4 | 52241187 | IGR | SBS | A | T |
| INDIA 8611 | AQP3 | chr9 | 33448138 | 5'Flank | SBS | G | C |
| INDIA 8611 | MIR920 | chr12 | 24207770 | 5'Flank | DEL | C | - |
| INDIA 8611 | RNU6-1265P | chr1 | 14119531 | 3'Flank | SBS | C | G |
| INDIA 8611 | ENST00000456240 | chr1 | 213778914 | Intron | SBS | A | T |
| INDIA 8611 | C1orf68 | chr1 | 152720523 | 3'Flank | SBS | C | G |
| INDIA 8611 | TENM3-AS1 | chr4 | 181981984 | Intron | SBS | G | T |
| INDIA 8611 | PKHD1 | chr6 | 51631388 | Intron | SBS | G | C |
| INDIA 8611 | PABPN1 | chr14 | 23320780 | 5'Flank | DEL | G | - |
| INDIA 8612 | MID1 | chrX | 10506101 | Intron | SBS | T | A |
| INDIA 8612 | Y_RNA | chrX | 106423196 | 5'Flank | SBS | G | C |
| INDIA 8612 | Unknown | chrX | 39585872 | IGR | SBS | G | C |
| INDIA 8612 | GNAQ | chr9 | 77839022 | Intron | SBS | C | G |
| INDIA 8612 | KRT6B | chr12 | 52445986 | 3'Flank | SBS | C | G |
| INDIA 8612 | Unknown | chr10 | 59969506 | IGR | SBS | C | G |
| INDIA 8612 | Unknown | chrX | 14136357 | IGR | SBS | C | G |
| INDIA 8612 | Unknown | chrX | 104262658 | IGR | SBS | C | G |
| INDIA 8612 | AGBL1 | chr15 | 86146725 | Intron | SBS | C | G |
| INDIA 8612 | Unknown | chrX | 66899548 | IGR | SBS | G | C |
| INDIA 8612 | VPS13D | chr1 | 12377896 | Intron | SBS | G | C |
| INDIA 8612 | EXOC6 | chr10 | 92940230 | Intron | SBS | G | C |
| INDIA 8612 | SORCS1 | chr10 | 107033949 | Intron | SBS | C | G |
| INDIA 8612 | Unknown | chr10 | 65394705 | IGR | SBS | G | C |

| | | | | | | | |
|---|---|---|---|---|---|---|---|
| INDIA 8612 | ENST00000560484 | chr15 | 39054244 | Intron | SBS | G | C |
| INDIA 8612 | USP6NL | chr10 | 11540218 | Intron | SBS | G | C |
| INDIA 8612 | FTX | chrX | 74207757 | Intron | SBS | G | C |
| INDIA 8612 | WTAP | chr6 | 159755054 | Missense_Mutation | SBS | G | C |
| INDIA 8612 | ENST00000422842 | chr10 | 54527137 | Intron | SBS | G | C |
| INDIA 8612 | LINC00523 | chr14 | 100673330 | 3'Flank | SBS | C | G |
| INDIA 8612 | CCDC136 | chr7 | 128815135 | Intron | SBS | G | C |
| INDIA 8612 | ANK3 | chr10 | 60138979 | Missense_Mutation | SBS | C | G |
| INDIA 8612 | MID1 | chrX | 10576872 | Intron | SBS | C | A |
| INDIA 8612 | VCPKMT | chr14 | 50111968 | Intron | DEL | AA | - |
| INDIA 8612 | FBXO39 | chr17 | 6781332 | Intron | SBS | G | C |
| INDIA 8612 | SORT1 | chr1 | 109368580 | Intron | SBS | C | G |
| INDIA 8612 | FHOD3 | chr18 | 36296418 | 5'Flank | SBS | C | G |
| INDIA 8612 | TAFA1 | chr3 | 68150263 | Intron | DEL | TT | - |
| INDIA 8612 | Unknown | chr4 | 12009794 | IGR | SBS | C | G |
| INDIA 8612 | ANKHD1 | chr5 | 140471414 | Intron | SBS | G | C |
| INDIA 8612 | OCRL | chrX | 129562815 | Intron | SBS | G | C |
| INDIA 8612 | FTCD | chr21 | 46137634 | Intron | SBS | G | C |
| INDIA 8612 | Unknown | chr20 | 21457466 | IGR | SBS | G | C |
| INDIA 8612 | Unknown | chr5 | 121935926 | IGR | SBS | G | C |
| INDIA 8612 | SH3PXD2A | chr10 | 103759927 | Intron | SBS | C | G |
| INDIA 8612 | ZNF510 | chr9 | 96757394 | 3'UTR | SBS | C | G |
| INDIA 8612 | PLEKHA4 | chr19 | 48869144 | 5'Flank | SBS | C | G |
| INDIA 8612 | C9orf92 | chr9 | 16218060 | Intron | SBS | G | C |
| INDIA 8612 | Unknown | chr15 | 94768353 | IGR | SBS | C | G |
| INDIA 8612 | Unknown | chr16 | 86691671 | IGR | SBS | C | G |
| INDIA 8612 | SPATA8-AS1 | chr15 | 96771372 | 3'Flank | SBS | C | G |
| INDIA 8612 | PTPRM | chr18 | 7809950 | Intron | SBS | C | G |
| INDIA 8612 | ZNF385D | chr3 | 21903521 | Intron | SBS | C | G |
| INDIA 8612 | MCC | chr5 | 113201812 | Intron | SBS | G | C |
| INDIA 8612 | AUTS2 | chr7 | 70060345 | Intron | SBS | T | A |
| INDIA 8612 | IFNAR1 | chr21 | 33345344 | Missense_Mutation | SBS | C | G |
| INDIA 8612 | SOX17 | chr8 | 54458787 | Intron | SBS | T | A |
| INDIA 8612 | Unknown | chr5 | 16960867 | IGR | SBS | G | C |
| INDIA 8612 | Unknown | chr6 | 116949790 | IGR | SBS | G | C |
| INDIA 8612 | DSCAM | chr21 | 40642370 | Intron | SBS | G | T |
| INDIA 8612 | Unknown | chr7 | 12263080 | IGR | DEL | A | - |
| INDIA 8612 | Unknown | chr2 | 168426118 | IGR | SBS | C | G |
| INDIA 8612 | OGDH | chr7 | 44678569 | Intron | SBS | G | C |
| INDIA 8612 | PYROXD2 | chr10 | 98385590 | Intron | SBS | C | G |
| INDIA 8612 | SLCO1C1 | chr12 | 20710526 | Intron | SBS | G | C |
| INDIA 8612 | Unknown | chr6 | 168174420 | IGR | SBS | C | G |
| INDIA 8612 | RGS7 | chr1 | 241229932 | Intron | SBS | G | C |
| INDIA 8612 | LINC01885 | chr2 | 107553052 | Intron | SBS | G | C |
| INDIA 8612 | TCF7L2 | chr10 | 113113312 | Intron | SBS | G | C |
| INDIA 8612 | RAD51B | chr14 | 68392785 | Intron | SBS | G | C |
| INDIA 8612 | ZNF536 | chr19 | 30657351 | Intron | SBS | G | C |
| INDIA 8612 | SNTA1 | chr20 | 33444631 | 5'Flank | SBS | C | G |
| INDIA 8612 | EPHB6 | chr7 | 142868216 | Intron | SBS | C | G |
| INDIA 8612 | GHET1 | chr7 | 148987133 | 5'Flank | SBS | C | G |
| INDIA 8612 | Unknown | chr8 | 127540711 | IGR | SBS | C | G |
| INDIA 8612 | KCNIP4 | chr4 | 20986200 | Intron | SBS | C | G |
| INDIA 8612 | Unknown | chr21 | 23192943 | IGR | SBS | C | G |
| INDIA 8612 | RASGRP1 | chr15 | 38532898 | Intron | SBS | G | C |
| INDIA 8612 | Unknown | chr4 | 102156735 | IGR | SBS | A | T |
| INDIA 8612 | SCAPER | chr15 | 76453983 | Intron | SBS | C | G |
| INDIA 8612 | LINC02100 | chr5 | 18701663 | 3'Flank | SBS | C | G |
| INDIA 8612 | CAMK1D | chr10 | 12634779 | Intron | SBS | C | G |
| INDIA 8612 | SENP8 | chr15 | 72117140 | 5'Flank | SBS | G | C |
| INDIA 8612 | ASCL4 | chr12 | 107776293 | 3'UTR | SBS | G | C |
| INDIA 8612 | LINC01885 | chr2 | 107491217 | Intron | SBS | T | G |
| INDIA 8612 | Unknown | chr1 | 54477552 | IGR | SBS | C | G |
| INDIA 8612 | Unknown | chr6 | 118354279 | IGR | SBS | G | C |
| INDIA 8612 | LINC01423 | chr21 | 38338068 | 5'Flank | SBS | C | G |
| INDIA 8612 | OBSCN | chr1 | 228264441 | Intron | SBS | C | G |
| INDIA 8612 | EDEM1 | chr3 | 5222931 | 3'Flank | SBS | C | G |

| | | | | | | | |
|---|---|---|---|---|---|---|---|
| INDIA 8612 | SUCLA2 | chr13 | 47755885 | Intron | SBS | G | C |
| INDIA 8612 | Unknown | chr5 | 106264613 | IGR | SBS | G | C |
| INDIA 8612 | CCDC136 | chr7 | 128815556 | Intron | SBS | G | C |
| INDIA 8612 | ITGB6 | chr2 | 160144566 | Intron | SBS | G | C |
| INDIA 8612 | BAHD1 | chr15 | 40443938 | Intron | SBS | C | G |
| INDIA 8612 | LINC01441 | chr20 | 55421732 | RNA | DEL | G | - |
| INDIA 8612 | GCSAML | chr1 | 247559481 | Intron | SBS | C | G |
| INDIA 8612 | ARMH3 | chr10 | 101933751 | Intron | SBS | G | C |
| INDIA 8612 | ENST00000605292 | chr20 | 10934606 | Intron | SBS | C | G |
| INDIA 8612 | LRIG1 | chr3 | 66424041 | Intron | SBS | C | G |
| INDIA 8612 | Unknown | chr4 | 59248619 | IGR | SBS | C | G |
| INDIA 8612 | LINC01933 | chr5 | 152004144 | Intron | SBS | G | C |
| INDIA 8612 | MYO18B | chr22 | 25909659 | Intron | SBS | C | G |
| INDIA 8612 | DAAM1 | chr14 | 59360911 | Intron | SBS | C | G |
| INDIA 8612 | Unknown | chr21 | 17407644 | IGR | SBS | C | G |
| INDIA 8612 | MGAT5 | chr2 | 134300520 | Intron | SBS | C | G |
| INDIA 8613 | ENST00000649962 | chr1 | 73901116 | Intron | SBS | G | C |
| INDIA 8613 | LINC01140 | chr1 | 87153702 | Intron | INS | - | T |
| INDIA 8613 | Unknown | chr1 | 153246040 | IGR | DEL | G | - |
| INDIA 8613 | Unknown | chr1 | 178048860 | IGR | SBS | G | C |
| INDIA 8613 | BABAM2 | chr2 | 28233329 | Intron | SBS | G | C |
| INDIA 8613 | CRIM1 | chr2 | 36529297 | Intron | SBS | C | G |
| INDIA 8613 | Unknown | chr2 | 37417924 | IGR | SBS | T | A |
| INDIA 8613 | Unknown | chr2 | 82731290 | IGR | DEL | G | - |
| INDIA 8613 | Unknown | chr2 | 88475315 | IGR | SBS | G | C |
| INDIA 8613 | Unknown | chr2 | 139444402 | IGR | DEL | C | - |
| INDIA 8613 | IFIH1 | chr2 | 162267629 | Intron | SBS | T | A |
| INDIA 8613 | KCNH7 | chr2 | 162813498 | Intron | SBS | G | T |
| INDIA 8613 | ANKAR | chr2 | 189735085 | Intron | SBS | C | G |
| INDIA 8613 | ROBO2 | chr3 | 76003090 | Intron | SBS | G | T |
| INDIA 8613 | CBLB | chr3 | 105713661 | Intron | SBS | C | G |
| INDIA 8613 | AGTR1 | chr3 | 148708830 | Intron | SBS | C | A |
| INDIA 8613 | Unknown | chr3 | 167097571 | IGR | SBS | G | C |
| INDIA 8613 | Unknown | chr3 | 194378778 | IGR | SBS | G | C |
| INDIA 8613 | Unknown | chr4 | 42762341 | IGR | SBS | C | G |
| INDIA 8613 | KCTD8 | chr4 | 44280444 | Intron | SBS | C | G |
| INDIA 8613 | Unknown | chr4 | 44591126 | IGR | SBS | T | A |
| INDIA 8613 | CWH43 | chr4 | 49046975 | Intron | SBS | G | T |
| INDIA 8613 | ARHGAP24 | chr4 | 85683032 | Intron | SBS | G | C |
| INDIA 8613 | ENST00000513332 | chr4 | 136144921 | Intron | DEL | T | - |
| INDIA 8613 | ENST00000660197 | chr4 | 154047935 | Intron | DEL | T | - |
| INDIA 8613 | Unknown | chr4 | 164854041 | IGR | SBS | G | C |
| INDIA 8613 | MARCHF11 | chr5 | 16178730 | Intron | DEL | G | - |
| INDIA 8613 | RAI14 | chr5 | 34755801 | Intron | SBS | G | C |
| INDIA 8613 | Unknown | chr5 | 58322977 | IGR | SBS | C | G |
| INDIA 8613 | Unknown | chr5 | 62004769 | IGR | SBS | G | C |
| INDIA 8613 | Unknown | chr6 | 71853274 | IGR | SBS | T | G |
| INDIA 8613 | Unknown | chr6 | 113465393 | IGR | SBS | A | T |
| INDIA 8613 | SGK1 | chr6 | 134198268 | Intron | SBS | C | G |
| INDIA 8613 | Unknown | chr7 | 18046822 | IGR | SBS | G | C |
| INDIA 8613 | RAPGEF5 | chr7 | 22256259 | Intron | SBS | A | C |
| INDIA 8613 | NPSR1-AS1 | chr7 | 34568485 | Intron | SBS | G | T |
| INDIA 8613 | Unknown | chr7 | 41224532 | IGR | DEL | G | - |
| INDIA 8613 | ENST00000447158 | chr7 | 69025312 | 5'Flank | SBS | C | G |
| INDIA 8613 | NPTX2 | chr7 | 98629837 | 3'UTR | DEL | G | - |
| INDIA 8613 | ENST00000656697 | chr7 | 112681458 | Intron | SBS | C | G |
| INDIA 8613 | KCND2 | chr7 | 120470188 | Intron | SBS | A | T |
| INDIA 8613 | Unknown | chr7 | 145734219 | IGR | SBS | T | G |
| INDIA 8613 | Unknown | chr7 | 153018684 | IGR | SBS | C | G |
| INDIA 8613 | Unknown | chr8 | 89261126 | IGR | SBS | G | C |
| INDIA 8613 | SHC3 | chr9 | 89090955 | Intron | SBS | A | T |
| INDIA 8613 | ENST00000652874 | chr10 | 22479916 | 5'Flank | DEL | T | - |
| INDIA 8613 | Unknown | chr10 | 25869809 | IGR | SBS | C | G |
| INDIA 8613 | PCDH15 | chr10 | 54955760 | Intron | DEL | G | - |
| INDIA 8613 | Unknown | chr10 | 117776485 | IGR | SBS | G | C |
| INDIA 8613 | Unknown | chr11 | 80804938 | IGR | SBS | G | C |

| | | | | | | | |
|---|---|---|---|---|---|---|---|
| INDIA 8613 | OPCML | chr11 | 133325291 | Intron | SBS | T | A |
| INDIA 8613 | Unknown | chr12 | 16893074 | IGR | SBS | C | G |
| INDIA 8613 | Unknown | chr12 | 74492069 | IGR | SBS | T | G |
| INDIA 8613 | PPFIA2 | chr12 | 81628887 | Intron | SBS | C | G |
| INDIA 8613 | EEA1 | chr12 | 92769410 | 3'Flank | SBS | G | C |
| INDIA 8613 | Unknown | chr13 | 105437377 | IGR | SBS | C | G |
| INDIA 8613 | MMP14 | chr14 | 22835285 | 5'Flank | SBS | C | G |
| INDIA 8613 | Unknown | chr14 | 25443343 | IGR | SBS | C | G |
| INDIA 8613 | LINC02306 | chr14 | 25909286 | Intron | DEL | C | - |
| INDIA 8613 | Unknown | chr14 | 27936496 | IGR | SBS | C | G |
| INDIA 8613 | NPAS3 | chr14 | 33396445 | Intron | SBS | C | G |
| INDIA 8613 | NIN | chr14 | 50716256 | 3'Flank | SBS | C | G |
| INDIA 8613 | LINC00640 | chr14 | 51352113 | Intron | SBS | C | G |
| INDIA 8613 | RNU7-116P | chr14 | 63736316 | 5'Flank | SBS | G | C |
| INDIA 8613 | MTHFD1 | chr14 | 64410710 | Intron | SBS | C | G |
| INDIA 8613 | TMEM63C | chr14 | 77124726 | Intron | SBS | A | T |
| INDIA 8613 | NRXN3 | chr14 | 79178700 | Intron | SBS | A | T |
| INDIA 8613 | RN7SL714P | chr14 | 98353354 | 5'Flank | SBS | C | G |
| INDIA 8613 | RYR3 | chr15 | 33532140 | Intron | SBS | G | C |
| INDIA 8613 | Unknown | chr15 | 60298018 | IGR | SBS | C | G |
| INDIA 8613 | PSTPIP1 | chr15 | 77012848 | Intron | SBS | C | G |
| INDIA 8613 | PCSK6 | chr15 | 101478711 | Intron | SBS | G | C |
| INDIA 8613 | HNRNPA1L3 | chr16 | 51571362 | Intron | SBS | A | C |
| INDIA 8613 | WWOX | chr16 | 78342621 | Intron | SBS | G | C |
| INDIA 8613 | ARHGAP44 | chr17 | 12820496 | Intron | SBS | G | C |
| INDIA 8613 | Unknown | chr17 | 13086175 | IGR | SBS | C | G |
| INDIA 8613 | PRR11 | chr17 | 59199759 | Intron | SBS | G | C |
| INDIA 8613 | ENST00000581165 | chr17 | 64420300 | 3'Flank | SBS | T | A |
| INDIA 8613 | Unknown | chr18 | 1443332 | IGR | SBS | C | G |
| INDIA 8613 | Unknown | chr18 | 40768183 | IGR | SBS | G | C |
| INDIA 8613 | ENST00000660208 | chr18 | 45113755 | Intron | SBS | G | C |
| INDIA 8613 | Unknown | chr18 | 65351879 | IGR | SBS | C | A |
| INDIA 8613 | Unknown | chr18 | 77903833 | IGR | SBS | T | A |
| INDIA 8613 | MED25 | chr19 | 49828387 | Intron | SBS | C | G |
| INDIA 8613 | PLCB1 | chr20 | 8658229 | Intron | SBS | C | G |
| INDIA 8613 | RALGAPA2 | chr20 | 20689895 | Intron | SBS | C | G |
| INDIA 8613 | OSBPL2 | chr20 | 62279237 | Missense_Mutation | SBS | A | T |
| INDIA 8613 | BRWD1 | chr21 | 39317304 | 5'Flank | SBS | G | C |
| INDIA 8613 | Y_RNA | chrX | 16914939 | 5'Flank | SBS | G | C |
| INDIA 8613 | LINC01456 | chrX | 18079781 | Intron | SBS | G | T |
| INDIA 8613 | Unknown | chrX | 27987093 | IGR | SBS | G | C |
| INDIA 8613 | Unknown | chrX | 40483769 | IGR | SBS | G | C |
| INDIA 8613 | Unknown | chrX | 46830448 | IGR | INS | - | T |
| INDIA 8613 | PCDH19 | chrX | 100354612 | Intron | SBS | G | C |
| INDIA 8613 | GUCY2F | chrX | 109453948 | Intron | SBS | C | A |
| INDIA 8614 | Unknown | chr16 | 17636773 | IGR | SBS | A | T |
| INDIA 8614 | Unknown | chr10 | 58085036 | IGR | SBS | A | C |
| INDIA 8614 | SYNE3 | chr14 | 95442731 | Intron | DEL | AGGCCT | - |
| INDIA 8614 | SLC35F4 | chr14 | 57965984 | Intron | SBS | G | C |
| INDIA 8614 | Unknown | chr7 | 49673364 | IGR | SBS | C | G |
| INDIA 8614 | SNTG2 | chr2 | 984012 | Intron | SBS | G | T |
| INDIA 8614 | LINC00378 | chr13 | 61150400 | Intron | SBS | G | C |
| INDIA 8614 | ENST00000624219 | chr22 | 48054135 | RNA | SBS | G | C |
| INDIA 8614 | Unknown | chr11 | 116131989 | IGR | SBS | A | T |
| INDIA 8614 | MROH2B | chr5 | 41066153 | Intron | SBS | G | C |
| INDIA 8614 | Unknown | chr11 | 15323480 | IGR | SBS | G | C |
| INDIA 8614 | VAC14 | chr16 | 70738461 | Intron | DEL | C | - |
| INDIA 8614 | MICAL3 | chr22 | 17829047 | Intron | DEL | G | - |
| INDIA 8614 | Unknown | chr9 | 100955290 | IGR | SBS | A | T |
| INDIA 8614 | Unknown | chr5 | 10909026 | IGR | SBS | C | G |
| INDIA 8614 | Unknown | chr18 | 60979993 | IGR | SBS | C | G |
| INDIA 8614 | ADARB1 | chr21 | 45131898 | Intron | SBS | C | G |
| INDIA 8614 | ARHGEF10L | chr1 | 17685029 | Intron | SBS | G | T |
| INDIA 8614 | NELL1 | chr11 | 21248432 | Intron | SBS | G | C |
| INDIA 8614 | CDH13 | chr16 | 83464009 | Intron | SBS | T | A |
| INDIA 8614 | PRKG1 | chr10 | 51844757 | Intron | SBS | T | G |

| | | | | | | | |
|---|---|---|---|---|---|---|---|
| INDIA 8614 | SIL1 | chr5 | 139178365 | Intron | DEL | C | - |
| INDIA 8614 | KSR2 | chr12 | 117586121 | Intron | INS | - | A |
| INDIA 8614 | OVOL2 | chr20 | 18053868 | Intron | SBS | A | C |
| INDIA 8616 | PTPRM | chr18 | 7822450 | Intron | DEL | CTAT | - |
| INDIA 8616 | FRMD4A | chr10 | 13841211 | Intron | SBS | C | G |
| INDIA 8616 | CTBP2 | chr10 | 125089573 | Intron | SBS | C | G |
| INDIA 8616 | SPIRE1 | chr18 | 12478183 | Intron | SBS | C | G |
| INDIA 8616 | SH3BGRL | chrX | 81263304 | Intron | DEL | A | - |
| INDIA 8616 | MAP3K15 | chrX | 19484555 | Intron | SBS | G | C |
| INDIA 8616 | MTCL1 | chr18 | 8801563 | Intron | SBS | T | G |
| INDIA 8616 | LINC00710 | chr10 | 10922339 | Intron | SBS | G | C |
| INDIA 8616 | ENST00000563503 | chr18 | 6638609 | 3'Flank | SBS | G | C |
| INDIA 8616 | Unknown | chr9 | 135038675 | IGR | SBS | G | C |
| INDIA 8616 | Unknown | chr19 | 29042883 | IGR | SBS | T | A |
| INDIA 8616 | LINC02223 | chr5 | 17703840 | Intron | SBS | C | G |
| INDIA 8616 | Unknown | chr13 | 55925486 | IGR | INS | - | A |
| INDIA 8616 | ESX1 | chrX | 104257867 | 5'Flank | SBS | G | T |
| INDIA 8616 | Unknown | chr13 | 81554819 | IGR | SBS | G | T |
| INDIA 8616 | CDH20 | chr18 | 61456324 | Intron | DEL | C | - |
| INDIA 8616 | MARCHF3 | chr5 | 126878270 | Missense_Mutation | SBS | T | A |
| INDIA 8616 | KIAA0825 | chr5 | 94157421 | Intron | SBS | T | A |
| INDIA 8616 | LINC02950 | chr8 | 8452417 | 3'Flank | SBS | C | G |
| INDIA 8616 | ENST00000639316 | chr18 | 2284807 | Intron | SBS | C | G |
| INDIA 8616 | ENST00000425150 | chrX | 51222561 | Intron | SBS | C | G |
| INDIA 8616 | Unknown | chr1 | 218776030 | IGR | SBS | G | C |
| INDIA 8616 | UBR4 | chr1 | 19171310 | Intron | INS | - | A |
| INDIA 8616 | Unknown | chr7 | 10787536 | IGR | SBS | G | C |
| INDIA 8616 | CEP104 | chr1 | 3847594 | Missense_Mutation | SBS | T | G |
| INDIA 8616 | CEP78 | chr9 | 78269168 | Intron | SBS | G | C |
| INDIA 8616 | LINC01235 | chr9 | 13456210 | Intron | SBS | C | G |
| INDIA 8616 | Unknown | chr7 | 51896215 | IGR | SBS | G | C |
| INDIA 8616 | CARD18 | chr11 | 105204157 | Intron | SBS | G | C |
| INDIA 8616 | Unknown | chr10 | 129185375 | IGR | SBS | G | C |
| INDIA 8616 | B3GALT1 | chr2 | 167652281 | Intron | SBS | C | G |
| INDIA 8616 | LINC01320 | chr2 | 34238873 | Intron | SBS | G | C |
| INDIA 8616 | RUBCN | chr3 | 197728813 | Intron | SBS | C | G |
| INDIA 8616 | Unknown | chr1 | 115810091 | IGR | SBS | C | G |
| INDIA 8616 | KCNIP1 | chr5 | 170441372 | Intron | SBS | G | C |
| INDIA 8616 | LINC02780 | chr1 | 3972104 | 5'Flank | SBS | C | G |
| INDIA 8616 | NOL9 | chr1 | 6519221 | 3'Flank | SBS | C | G |
| INDIA 8616 | KCNMB2 | chr3 | 178629220 | Intron | SBS | T | A |
| INDIA 8616 | AK8 | chr9 | 132806902 | Intron | SBS | C | G |
| INDIA 8616 | KLF5 | chr13 | 73076192 | 3'UTR | SBS | G | C |
| INDIA 8616 | ZNF106 | chr15 | 42450968 | Missense_Mutation | SBS | C | G |
| INDIA 8616 | MAP2K1 | chr15 | 66455999 | Intron | SBS | C | G |
| INDIA 8616 | MIR3976HG | chr18 | 5793568 | Intron | SBS | C | G |
| INDIA 8616 | FBN3 | chr19 | 8123502 | Missense_Mutation | SBS | C | G |
| INDIA 8616 | Unknown | chr20 | 11430440 | IGR | SBS | T | G |
| INDIA 8616 | GREM2 | chr1 | 240547092 | Intron | SBS | T | A |
| INDIA 8616 | JCAD | chr10 | 30042881 | Intron | SBS | G | C |
| INDIA 8616 | SPTSSB | chr3 | 161349955 | Intron | SBS | G | C |
| INDIA 8616 | Unknown | chr8 | 69221081 | IGR | SBS | C | G |
| INDIA 8616 | Unknown | chr10 | 2294059 | IGR | SBS | G | C |
| INDIA 8616 | OXCT1 | chr5 | 41767626 | Intron | SBS | C | G |
| INDIA 8616 | Unknown | chr5 | 8591096 | IGR | SBS | C | A |
| INDIA 8616 | C20orf96 | chr20 | 278478 | Intron | SBS | G | C |
| INDIA 8616 | HNF1A-AS1 | chr12 | 120961809 | Intron | SBS | G | T |
| INDIA 8616 | COL19A1 | chr6 | 70169308 | Intron | SBS | G | C |
| INDIA 8616 | Unknown | chr1 | 217886941 | IGR | SBS | C | G |
| INDIA 8616 | CNBD1 | chr8 | 87294347 | Intron | SBS | C | G |
| INDIA 8616 | SLC16A9 | chr10 | 59657733 | Intron | SBS | G | C |
| INDIA 8616 | CTIF | chr18 | 48859081 | Intron | SBS | G | C |
| INDIA 8616 | PEX6 | chr6 | 42969607 | Intron | SBS | G | C |
| INDIA 8616 | Unknown | chr1 | 5460153 | IGR | SBS | G | C |
| INDIA 8616 | SND1 | chr7 | 127978321 | Intron | SBS | G | C |
| INDIA 8616 | SPTAN1 | chr9 | 128566876 | Frame_Shift_Del | DEL | T | - |

| | | | | | | | |
|---|---|---|---|---|---|---|---|
| INDIA 8616 | ITGB1-DT | chr10 | 33071238 | Intron | SBS | C | G |
| INDIA 8616 | Unknown | chr7 | 134623485 | IGR | SBS | G | C |
| INDIA 8616 | SMC5 | chr9 | 70351603 | Intron | SBS | G | C |
| INDIA 8616 | Unknown | chr11 | 39917322 | IGR | SBS | C | A |
| INDIA 8616 | ENST00000665445 | chr2 | 76668111 | Intron | SBS | C | G |
| INDIA 8616 | BAZ1B | chr7 | 73438517 | 3'Flank | SBS | G | C |
| INDIA 8616 | ENST00000578650 | chr17 | 31604616 | Intron | SBS | C | G |
| INDIA 8616 | TGM3 | chr20 | 2320693 | Intron | SBS | G | C |
| INDIA 8616 | PLCB1 | chr20 | 8500583 | Intron | SBS | G | C |
| INDIA 8616 | LINC02026 | chr3 | 193965922 | Intron | SBS | C | G |
| INDIA 8616 | SPTSSB | chr3 | 161350017 | Intron | SBS | G | C |
| INDIA 8616 | Unknown | chr4 | 46025085 | IGR | SBS | T | G |
| INDIA 8616 | LINC01954 | chr2 | 10887699 | 3'Flank | SBS | C | G |
| INDIA 8616 | LINC01806 | chr2 | 161242108 | 5'Flank | SBS | G | T |
| INDIA 8616 | FAT3 | chr11 | 92506068 | Intron | DEL | A | - |
| INDIA 8616 | SATB2 | chr2 | 199271993 | 3'UTR | SBS | G | C |
| INDIA 8616 | INPP4A | chr2 | 98451751 | Intron | SBS | T | G |
| INDIA 8616 | Unknown | chr2 | 112805742 | IGR | SBS | C | G |
| INDIA 8616 | Unknown | chr6 | 91046177 | IGR | SBS | T | A |
| INDIA 8616 | LINC00645 | chr14 | 27625301 | Intron | SBS | A | C |
| INDIA 8616 | PRKCH-AS1 | chr14 | 61316519 | Intron | SBS | C | G |
| INDIA 8616 | PAK5 | chr20 | 9821940 | Intron | SBS | G | C |
| INDIA 8616 | Unknown | chr7 | 46604964 | IGR | SBS | C | G |
| INDIA 8616 | ARMC9 | chr2 | 231196196 | 5'Flank | SBS | A | T |
| INDIA 8616 | RPRD1A | chr18 | 36041814 | Intron | SBS | C | G |
| INDIA 8616 | Unknown | chr1 | 232077916 | IGR | SBS | G | C |
| INDIA 8616 | CALD1 | chr7 | 134859872 | Intron | SBS | C | G |
| INDIA 8616 | LINC00589 | chr8 | 29738581 | 5'Flank | SBS | G | T |
| INDIA 8616 | TBXAS1 | chr7 | 139814137 | Intron | SBS | G | C |
| INDIA 8616 | CNTNAP2 | chr7 | 148170692 | Intron | SBS | G | C |
| INDIA 8616 | Unknown | chr10 | 25845401 | IGR | SBS | C | G |
| INDIA 8616 | AP1G1 | chr16 | 71754973 | Intron | SBS | T | G |
| INDIA 8616 | MAML3 | chr4 | 139969734 | Intron | SBS | G | C |
| INDIA 8617 | PLA2G5 | chr1 | 20091298 | 3'UTR | SBS | G | C |
| INDIA 8617 | LINC01635 | chr1 | 22030514 | 5'Flank | SBS | C | G |
| INDIA 8617 | HMGB4 | chr1 | 33866224 | 3'Flank | SBS | T | A |
| INDIA 8617 | EPHA10 | chr1 | 37719089 | Intron | SBS | C | G |
| INDIA 8617 | CTPS1 | chr1 | 41007357 | Intron | SBS | T | G |
| INDIA 8617 | CMPK1 | chr1 | 47333684 | 5'Flank | DEL | A | - |
| INDIA 8617 | CACHD1 | chr1 | 64482930 | Intron | SBS | C | G |
| INDIA 8617 | Unknown | chr1 | 119816607 | IGR | SBS | G | C |
| INDIA 8617 | LINC01527 | chr1 | 152934417 | Intron | SBS | C | G |
| INDIA 8617 | PBX1 | chr1 | 164672129 | Intron | SBS | C | G |
| INDIA 8617 | Unknown | chr1 | 165143690 | IGR | SBS | G | C |
| INDIA 8617 | Unknown | chr1 | 167714972 | IGR | SBS | T | A |
| INDIA 8617 | Unknown | chr2 | 67835589 | IGR | SBS | T | A |
| INDIA 8617 | TACR1 | chr2 | 75096359 | Intron | SBS | C | G |
| INDIA 8617 | Unknown | chr2 | 82183562 | IGR | SBS | C | G |
| INDIA 8617 | KDM3A | chr2 | 86489687 | Intron | SBS | G | C |
| INDIA 8617 | CERS6 | chr2 | 168517980 | Intron | SBS | A | C |
| INDIA 8617 | ENST00000665393 | chr2 | 200084866 | 5'Flank | SBS | C | G |
| INDIA 8617 | ALS2 | chr2 | 201707932 | Missense_Mutation | SBS | G | C |
| INDIA 8617 | NHEJ1 | chr2 | 219120905 | Intron | INS | - | A |
| INDIA 8617 | USP40 | chr2 | 233560378 | Intron | SBS | G | C |
| INDIA 8617 | EPHA6 | chr3 | 96850561 | Intron | SBS | G | C |
| INDIA 8617 | ENST00000465477 | chr3 | 158771374 | Intron | SBS | C | G |
| INDIA 8617 | ENST00000658869 | chr3 | 170692716 | RNA | SBS | C | G |
| INDIA 8617 | ENST00000657847 | chr3 | 171471312 | 3'Flank | SBS | G | C |
| INDIA 8617 | TP63 | chr3 | 189831134 | Intron | SBS | T | G |
| INDIA 8617 | RBM47 | chr4 | 40449020 | Intron | SBS | A | C |
| INDIA 8617 | ANTXR2 | chr4 | 79897348 | 3'Flank | SBS | G | C |
| INDIA 8617 | Unknown | chr4 | 115611834 | IGR | SBS | C | A |
| INDIA 8617 | PALLD | chr4 | 168785647 | Intron | SBS | G | T |
| INDIA 8617 | SNX25 | chr4 | 185351591 | In_Frame_Del | DEL | CTT | - |
| INDIA 8617 | Unknown | chr5 | 3254613 | IGR | SBS | G | C |
| INDIA 8617 | ADCY2 | chr5 | 7502992 | Intron | SBS | A | C |

| | | | | | | | |
|---|---|---|---|---|---|---|---|
| INDIA 8617 | ADCY2 | chr5 | 7803308 | Intron | SBS | T | A |
| INDIA 8617 | LINC02112 | chr5 | 9651665 | Intron | SBS | C | G |
| INDIA 8617 | Unknown | chr5 | 57778925 | IGR | SBS | C | G |
| INDIA 8617 | ERCC8 | chr5 | 60891876 | Intron | SBS | C | G |
| INDIA 8617 | ANKDD1B | chr5 | 75667327 | Intron | SBS | G | C |
| INDIA 8617 | EFNA5 | chr5 | 107592819 | Intron | SBS | A | C |
| INDIA 8617 | MEGF10 | chr5 | 127314117 | Intron | SBS | G | C |
| INDIA 8617 | Unknown | chr5 | 155333208 | IGR | SBS | C | G |
| INDIA 8617 | MIR3142HG | chr5 | 160473829 | Intron | SBS | C | G |
| INDIA 8617 | CDKAL1 | chr6 | 20768894 | Intron | SBS | G | C |
| INDIA 8617 | ZKSCAN8P1 | chr6 | 28166538 | 5'UTR | SBS | G | C |
| INDIA 8617 | Unknown | chr6 | 87869185 | IGR | SBS | G | C |
| INDIA 8617 | LAMA2 | chr6 | 129501632 | Intron | SBS | C | A |
| INDIA 8617 | Unknown | chr7 | 19201938 | IGR | SBS | T | G |
| INDIA 8617 | ENST00000659273 | chr7 | 25758781 | 3'Flank | SBS | G | C |
| INDIA 8617 | Unknown | chr7 | 81808077 | IGR | SBS | T | A |
| INDIA 8617 | DOCK4 | chr7 | 111970986 | Intron | SBS | T | G |
| INDIA 8617 | Unknown | chr7 | 123386411 | IGR | SBS | T | A |
| INDIA 8617 | ENST00000415393 | chr7 | 131908995 | 5'Flank | SBS | G | C |
| INDIA 8617 | PLXNA4 | chr7 | 132145699 | Intron | SBS | G | C |
| INDIA 8617 | ENST00000656533 | chr7 | 134432835 | 5'Flank | SBS | C | G |
| INDIA 8617 | CNTNAP2 | chr7 | 147672677 | Intron | SBS | A | T |
| INDIA 8617 | Unknown | chr8 | 2373056 | IGR | SBS | G | C |
| INDIA 8617 | SOX7 | chr8 | 10730981 | 5'Flank | SBS | T | A |
| INDIA 8617 | Unknown | chr8 | 11876838 | IGR | SBS | C | G |
| INDIA 8617 | ENST00000661295 | chr8 | 37335915 | 5'Flank | SBS | G | C |
| INDIA 8617 | Unknown | chr8 | 55008974 | IGR | SBS | G | C |
| INDIA 8617 | ASAP1 | chr8 | 130219508 | Intron | SBS | T | G |
| INDIA 8617 | Unknown | chr9 | 71527384 | IGR | SBS | G | C |
| INDIA 8617 | LNCARSR | chr9 | 79553940 | Intron | SBS | G | T |
| INDIA 8617 | AOPEP | chr9 | 94941405 | Intron | SBS | T | G |
| INDIA 8617 | SHOC1 | chr9 | 111746302 | Missense_Mutation | SBS | G | C |
| INDIA 8617 | SORBS1 | chr10 | 95429767 | Intron | SBS | C | G |
| INDIA 8617 | PLEKHA7 | chr11 | 16895293 | Intron | DEL | TTA | - |
| INDIA 8617 | CD44 | chr11 | 35174415 | Intron | SBS | G | C |
| INDIA 8617 | Unknown | chr11 | 80192917 | IGR | SBS | G | C |
| INDIA 8617 | PANX1 | chr11 | 94132469 | Intron | SBS | G | C |
| INDIA 8617 | WNT5B | chr12 | 1604687 | Intron | DEL | C | - |
| INDIA 8617 | PIK3C2G | chr12 | 18422103 | Intron | SBS | C | G |
| INDIA 8617 | MED13L | chr12 | 115954294 | 3'Flank | SBS | G | T |
| INDIA 8617 | NEK5 | chr13 | 52083348 | Frame_Shift_Del | DEL | T | - |
| INDIA 8617 | Unknown | chr13 | 78748308 | IGR | SBS | G | C |
| INDIA 8617 | Unknown | chr14 | 89701435 | IGR | SBS | C | G |
| INDIA 8617 | HDC | chr15 | 50248562 | Intron | SBS | G | C |
| INDIA 8617 | ARIH1 | chr15 | 72549017 | Intron | SBS | T | G |
| INDIA 8617 | TMC3 | chr15 | 81371271 | Intron | SBS | C | G |
| INDIA 8617 | SYNM-AS1 | chr15 | 99035764 | Intron | SBS | G | C |
| INDIA 8617 | ANKFN1 | chr17 | 56159699 | Intron | SBS | T | A |
| INDIA 8617 | ERN1 | chr17 | 64081236 | Intron | SBS | A | T |
| INDIA 8617 | SLC39A11 | chr17 | 72727141 | Intron | SBS | G | C |
| INDIA 8617 | SEPTIN9 | chr17 | 77495246 | Intron | SBS | G | C |
| INDIA 8617 | RPTOR | chr17 | 80891795 | Missense_Mutation | SBS | G | C |
| INDIA 8617 | Unknown | chr18 | 59837696 | IGR | SBS | T | G |
| INDIA 8617 | PARD6G | chr18 | 80182708 | Intron | SBS | G | C |
| INDIA 8617 | GPATCH1 | chr19 | 33109846 | Frame_Shift_Del | DEL | GC | - |
| INDIA 8617 | Unknown | chr20 | 47382659 | IGR | SBS | C | G |
| INDIA 8617 | SPATA2 | chr20 | 49918952 | 5'Flank | SBS | C | G |
| INDIA 8617 | SEZ6L | chr22 | 26214528 | Intron | SBS | A | T |
| INDIA 8617 | DIAPH2 | chrX | 97308418 | Intron | SBS | C | G |

**Table S2: ctDNA and utDNA results for samples included in study.**

| Patient | Sample Type | Sample description | # Mutations Tracked | # Mutations identified | Total # Mutant Molecules | Total # Molecules Assessed | Mutant Allele Fraction (%) |
|---|---|---|---|---|---|---|---|
| INDIA 8600 | Urine | Restaging | 92 | 0 | 0 | 119592 | 0.00000% |
| INDIA 8549 | Urine supernatant | Restaging | 96 | 0 | 0 | 436528 | 0.00000% |
| INDIA 8565 | Urine supernatant | Restaging | 94 | 0 | 0 | 373193 | 0.00000% |
| INDIA 8575 | Urine | Restaging | 93 | 0 | 0 | 93041 | 0.00000% |
| INDIA 8577 | Urine | Restaging | 96 | 0 | 0 | 148563 | 0.00000% |
| INDIA 8580 | Urine supernatant | Restaging | 93 | 1 | 1 | 73368 | 0.00136% |
| INDIA 8587 | Urine | Restaging | 92 | 0 | 0 | 10450 | 0.00000% |
| INDIA 8592 | Urine | Restaging | 93 | 0 | 0 | 138679 | 0.00000% |
| INDIA 8613 | Urine | Restaging | 95 | 0 | 0 | 580197 | 0.00000% |
| INDIA 8601 | Urine | Restaging | 61 | 1 | 1 | 612509 | 0.00016% |
| INDIA 8593 | Urine | Restaging | 96 | 0 | 0 | 1270 | 0.00000% |
| INDIA 8564 | Urine supernatant | Restaging | 90 | 0 | 0 | 45524 | 0.00000% |
| INDIA 8588 | Urine | Restaging | 92 | 1 | 1 | 134831 | 0.00074% |
| INDIA 8595 | Urine | Restaging | 94 | 0 | 0 | 65835 | 0.00000% |
| INDIA 8548 | Urine supernatant | Restaging | 85 | 4 | 4 | 140952 | 0.00284% |
| INDIA 8579 | Urine supernatant | Restaging | 94 | 85 | 248 | 261959 | 0.09467% |
| INDIA 8611 | Urine | Restaging | 91 | 6 | 6 | 233293 | 0.00257% |
| INDIA 8544 | Urine | Restaging | 75 | 27 | 36 | 245967 | 0.01464% |
| INDIA 8550 | Urine | Restaging | 84 | 0 | 0 | 65138 | 0.00000% |
| INDIA 8561 | Urine supernatant | Restaging | 93 | 17 | 20 | 144831 | 0.01381% |
| INDIA 8566 | Urine supernatant | Restaging | 93 | 1 | 1 | 18568 | 0.00539% |
| INDIA 8568 | Urine supernatant | Restaging | 96 | 34 | 58 | 793190 | 0.00731% |
| INDIA 8571 | Urine | Restaging | 92 | 27 | 91 | 137336 | 0.06626% |
| INDIA 8610 | Urine | Restaging | 88 | 15 | 25 | 16291 | 0.15346% |
| INDIA 8556 | Urine pellet | Restaging | 39 | 0 | 0 | 131140 | 0.00000% |
| INDIA 8590 | Urine | Restaging | 95 | 2 | 2 | 305865 | 0.00065% |
| INDIA 8554 | Urine pellet | Restaging | 91 | 1 | 1 | 269490 | 0.00037% |
| INDIA 8596 | Urine | Restaging | 95 | 91 | 2731 | 691385 | 0.39500% |
| INDIA 8597 | Urine | Restaging | 96 | 63 | 6004 | 168079 | 3.57213% |
| INDIA 8547 | Urine pellet | Restaging | 71 | 16 | 25 | 289611 | 0.00863% |
| INDIA 8552 | Urine pellet | Restaging | 94 | 65 | 243 | 305298 | 0.07959% |
| INDIA 8553 | Urine | Restaging | 95 | 1 | 1 | 385509 | 0.00026% |
| INDIA 8555 | Urine | Restaging | 92 | 0 | 0 | 29717 | 0.00000% |
| INDIA 8563 | Urine pellet | Restaging | 94 | 72 | 224 | 511584 | 0.04379% |
| INDIA 8569 | Urine pellet | Restaging | 93 | 43 | 10952 | 59997 | 18.25425% |
| INDIA 8570 | Urine pellet | Restaging | 92 | 44 | 91 | 158667 | 0.05735% |
| INDIA 8572 | Urine | Restaging | 93 | 91 | 57350 | 778879 | 7.36315% |
| INDIA 8573 | Urine pellet | Restaging | 89 | 23 | 29 | 53284 | 0.05443% |
| INDIA 8574 | Urine pellet | Restaging | 91 | 66 | 10125 | 89682 | 11.28989% |
| INDIA 8576 | Urine pellet | Restaging | 90 | 87 | 7237 | 92909 | 7.78934% |
| INDIA 8578 | Urine pellet | Restaging | 96 | 75 | 5950 | 69400 | 8.57349% |
| INDIA 8581 | Urine pellet | Restaging | 95 | 62 | 168 | 36222 | 0.46381% |
| INDIA 8582 | Urine | Restaging | 94 | 22 | 37 | 8955 | 0.41318% |
| INDIA 8585 | Urine | Restaging | 95 | 25 | 45 | 76213 | 0.05905% |
| INDIA 8586 | Urine | Restaging | 96 | 19 | 29 | 59156 | 0.04902% |
| INDIA 8591 | Urine | Restaging | 93 | 92 | 19761 | 837496 | 2.35953% |
| INDIA 8598 | Urine | Restaging | 93 | 63 | 160 | 43578 | 0.36716% |
| INDIA 8599 | Urine | Restaging | 92 | 61 | 237 | 20291 | 1.16801% |
| INDIA 8614 | Urine | Restaging | 24 | 6 | 8 | 14693 | 0.05445% |
| INDIA 8605 | Urine | Restaging | 94 | 3 | 4 | 151889 | 0.00263% |
| INDIA 8607 | Urine | Restaging | 82 | 2 | 3 | 37113 | 0.00808% |
| INDIA 8609 | Urine | Restaging | 92 | 89 | 1642 | 106773 | 1.53784% |
| INDIA 8612 | Urine | Restaging | 96 | 6 | 6 | 1536 | 0.39063% |
| INDIA 8616 | Urine | Restaging | 96 | 27 | 41 | 30928 | 0.13257% |
| INDIA 8548 | Plasma | End of treatment | 85 | 0 | 0 | 265919 | 0.00000% |
| INDIA 8548 | PBMC | Baseline | 85 | 0 | 0 | 881737 | 0.00000% |
| INDIA 8549 | PBMC | Baseline | 96 | 0 | 0 | 1001413 | 0.00000% |
| INDIA 8559 | PBMC | Baseline | 90 | 0 | 0 | 940836 | 0.00000% |
| INDIA 8565 | PBMC | Baseline | 94 | 0 | 0 | 265258 | 0.00000% |
| INDIA 8575 | PBMC | Baseline | 93 | 0 | 0 | 776758 | 0.00000% |
| INDIA 8577 | PBMC | Baseline | 96 | 0 | 0 | 716796 | 0.00000% |
| INDIA 8579 | PBMC | Baseline | 94 | 0 | 0 | 638649 | 0.00000% |
| INDIA 8580 | PBMC | Baseline | 93 | 0 | 0 | 652496 | 0.00000% |
| INDIA 8583 | PBMC | Baseline | 88 | 0 | 0 | 670480 | 0.00000% |

| | | | | | | | |
|---|---|---|---|---|---|---|---|
| INDIA 8587 | PBMC | Baseline | 92 | 0 | 0 | 914179 | 0.00000% |
| INDIA 8592 | PBMC | Baseline | 93 | 0 | 0 | 727078 | 0.00000% |
| INDIA 8594 | PBMC | Baseline | 94 | 0 | 0 | 821160 | 0.00000% |
| INDIA 8613 | PBMC | Baseline | 95 | 0 | 0 | 817463 | 0.00000% |
| INDIA 8600 | PBMC | Baseline | 92 | 0 | 0 | 792224 | 0.00000% |
| INDIA 8601 | PBMC | Baseline | 61 | 0 | 0 | 826568 | 0.00000% |
| INDIA 8604 | PBMC | Baseline | 95 | 0 | 0 | 1482601 | 0.00000% |
| INDIA 8611 | PBMC | Baseline | 91 | 0 | 0 | 1165753 | 0.00000% |
| INDIA 8544 | PBMC | Baseline | 75 | 0 | 0 | 738495 | 0.00000% |
| INDIA 8550 | PBMC | Baseline | 84 | 0 | 0 | 823984 | 0.00000% |
| INDIA 8561 | PBMC | Baseline | 93 | 0 | 0 | 608566 | 0.00000% |
| INDIA 8564 | PBMC | Baseline | 90 | 0 | 0 | 494880 | 0.00000% |
| INDIA 8566 | PBMC | Baseline | 93 | 0 | 0 | 810602 | 0.00000% |
| INDIA 8568 | PBMC | Baseline | 96 | 0 | 0 | 976958 | 0.00000% |
| INDIA 8571 | PBMC | Baseline | 92 | 0 | 0 | 797053 | 0.00000% |
| INDIA 8588 | PBMC | Baseline | 92 | 0 | 0 | 870557 | 0.00000% |
| INDIA 8593 | PBMC | Baseline | 96 | 0 | 0 | 856319 | 0.00000% |
| INDIA 8595 | PBMC | Baseline | 94 | 0 | 0 | 819297 | 0.00000% |
| INDIA 8610 | PBMC | Baseline | 88 | 0 | 0 | 1189200 | 0.00000% |
| INDIA 8556 | PBMC | Baseline | 39 | 0 | 0 | 354872 | 0.00000% |
| INDIA 8560 | PBMC | Baseline | 89 | 0 | 0 | 902521 | 0.00000% |
| INDIA 8584 | PBMC | Baseline | 46 | 0 | 0 | 421740 | 0.00000% |
| INDIA 8590 | Plasma | Restaging | 95 | 25 | 32 | 2697894 | 0.00000% |
| INDIA 8547 | PBMC | Baseline | 71 | 0 | 0 | 811853 | 0.00000% |
| INDIA 8551 | PBMC | Baseline | 83 | 0 | 0 | 831403 | 0.00000% |
| INDIA 8552 | PBMC | Baseline | 94 | 0 | 0 | 768951 | 0.00000% |
| INDIA 8553 | PBMC | Baseline | 95 | 0 | 0 | 761823 | 0.00000% |
| INDIA 8554 | PBMC | Baseline | 91 | 0 | 0 | 627607 | 0.00000% |
| INDIA 8555 | PBMC | Baseline | 92 | 0 | 0 | 939664 | 0.00000% |
| INDIA 8557 | PBMC | Baseline | 93 | 0 | 0 | 497812 | 0.00000% |
| INDIA 8563 | PBMC | Baseline | 94 | 0 | 0 | 975699 | 0.00000% |
| INDIA 8569 | PBMC | Baseline | 93 | 0 | 0 | 881195 | 0.00000% |
| INDIA 8570 | PBMC | Baseline | 92 | 0 | 0 | 886927 | 0.00000% |
| INDIA 8572 | PBMC | Baseline | 93 | 0 | 0 | 837389 | 0.00000% |
| INDIA 8573 | PBMC | Baseline | 89 | 0 | 0 | 929383 | 0.00000% |
| INDIA 8574 | PBMC | Baseline | 91 | 0 | 0 | 1000922 | 0.00000% |
| INDIA 8576 | PBMC | Baseline | 90 | 0 | 0 | 934969 | 0.00000% |
| INDIA 8578 | PBMC | Baseline | 96 | 0 | 0 | 747810 | 0.00000% |
| INDIA 8581 | PBMC | Baseline | 95 | 0 | 0 | 953873 | 0.00000% |
| INDIA 8582 | PBMC | Baseline | 94 | 0 | 0 | 1049386 | 0.00000% |
| INDIA 8585 | PBMC | Baseline | 95 | 0 | 0 | 797546 | 0.00000% |
| INDIA 8586 | PBMC | Baseline | 96 | 0 | 0 | 711917 | 0.00000% |
| INDIA 8591 | PBMC | Baseline | 93 | 0 | 0 | 697974 | 0.00000% |
| INDIA 8596 | PBMC | Baseline | 95 | 0 | 0 | 941008 | 0.00000% |
| INDIA 8597 | PBMC | Baseline | 96 | 0 | 0 | 870452 | 0.00000% |
| INDIA 8598 | PBMC | Baseline | 93 | 0 | 0 | 856502 | 0.00000% |
| INDIA 8599 | PBMC | Baseline | 92 | 0 | 0 | 609550 | 0.00000% |
| INDIA 8614 | PBMC | Baseline | 24 | 0 | 0 | 259929 | 0.00000% |
| INDIA 8603 | PBMC | Baseline | 82 | 0 | 0 | 1241466 | 0.00000% |
| INDIA 8605 | PBMC | Baseline | 94 | 0 | 0 | 1035148 | 0.00000% |
| INDIA 8607 | PBMC | Baseline | 82 | 0 | 0 | 570141 | 0.00000% |
| INDIA 8608 | PBMC | Baseline | 64 | 0 | 0 | 515896 | 0.00000% |
| INDIA 8609 | PBMC | Baseline | 92 | 0 | 0 | 1110066 | 0.00000% |
| INDIA 8612 | PBMC | Baseline | 96 | 0 | 0 | 485529 | 0.00000% |
| INDIA 8616 | PBMC | Baseline | 96 | 0 | 0 | 1556217 | 0.00000% |
| INDIA 8617 | PBMC | Baseline | 92 | 0 | 0 | 93292 | 0.00000% |
| INDIA 8544 | Urine supernatant | Restaging | 75 | 25 | 37 | 348208 | 0.01063% |
| INDIA 8547 | Urine supernatant | Restaging | 71 | 48 | 117 | 626920 | 0.01866% |
| INDIA 8547 | Urine | Restaging | 71 | 15 | 23 | 163827 | 0.01404% |
| INDIA 8548 | Urine | Restaging | 85 | 4 | 6 | 420537 | 0.00143% |
| INDIA 8548 | Urine pellet | Restaging | 85 | 7 | 8 | 49273 | 0.01624% |
| INDIA 8549 | Urine | Restaging | 96 | 0 | 0 | 263161 | 0.00000% |
| INDIA 8549 | Urine pellet | Restaging | 96 | 0 | 0 | 139531 | 0.00000% |
| INDIA 8550 | Urine pellet | Restaging | 84 | 5 | 6 | 163906 | 0.00366% |
| INDIA 8552 | Urine supernatant | Restaging | 94 | 35 | 51 | 54401 | 0.09375% |
| INDIA 8552 | Urine | Restaging | 94 | 66 | 252 | 288561 | 0.08733% |
| INDIA 8553 | Urine supernatant | Restaging | 95 | 3 | 3 | 445270 | 0.00067% |

| | | | | | | | |
|---|---|---|---|---|---|---|---|
| INDIA 8554 | Urine supernatant | Restaging | 91 | 1 | 1 | 295599 | 0.00034% |
| INDIA 8554 | Urine | Restaging | 91 | 1 | 1 | 499544 | 0.00020% |
| INDIA 8555 | Urine pellet | Restaging | 92 | 4 | 7 | 245335 | 0.00285% |
| INDIA 8556 | Urine supernatant | Restaging | 39 | 0 | 0 | 26683 | 0.00000% |
| INDIA 8556 | Urine | Restaging | 39 | 0 | 0 | 66688 | 0.00000% |
| INDIA 8561 | Urine | Restaging | 93 | 19 | 22 | 211075 | 0.01042% |
| INDIA 8561 | Urine pellet | Restaging | 93 | 25 | 36 | 153222 | 0.02350% |
| INDIA 8563 | Urine supernatant | Restaging | 94 | 89 | 748 | 598361 | 0.12501% |
| INDIA 8563 | Urine | Restaging | 94 | 91 | 3183 | 1016267 | 0.31321% |
| INDIA 8564 | Urine | Restaging | 90 | 0 | 0 | 59831 | 0.00000% |
| INDIA 8564 | Urine pellet | Restaging | 90 | 0 | 0 | 5859 | 0.00000% |
| INDIA 8565 | Urine | Restaging | 94 | 0 | 0 | 308868 | 0.00000% |
| INDIA 8565 | Urine pellet | Restaging | 94 | 0 | 0 | 176828 | 0.00000% |
| INDIA 8566 | Urine | Restaging | 93 | 0 | 0 | 31413 | 0.00000% |
| INDIA 8566 | Urine pellet | Restaging | 93 | 1 | 3 | 7057 | 0.04251% |
| INDIA 8568 | Urine | Restaging | 96 | 29 | 45 | 920730 | 0.00489% |
| INDIA 8568 | Urine pellet | Restaging | 96 | 35 | 49 | 463737 | 0.01057% |
| INDIA 8569 | Urine supernatant | Restaging | 93 | 51 | 37451 | 235802 | 15.88239% |
| INDIA 8569 | Urine | Restaging | 93 | 58 | 75369 | 467272 | 16.12958% |
| INDIA 8570 | Urine supernatant | Restaging | 92 | 74 | 261 | 430267 | 0.06066% |
| INDIA 8570 | Urine | Restaging | 92 | 72 | 297 | 640614 | 0.04636% |
| INDIA 8571 | Urine pellet | Restaging | 92 | 31 | 539 | 219651 | 0.24539% |
| INDIA 8572 | Urine supernatant | Restaging | 93 | 91 | 37352 | 506028 | 7.38141% |
| INDIA 8573 | Urine supernatant | Restaging | 89 | 57 | 151 | 523868 | 0.02882% |
| INDIA 8573 | Urine | Restaging | 89 | 55 | 195 | 780257 | 0.02499% |
| INDIA 8574 | Urine supernatant | Restaging | 91 | 30 | 62 | 1588 | 3.90428% |
| INDIA 8574 | Urine | Restaging | 91 | 63 | 718 | 21615 | 3.32177% |
| INDIA 8575 | Urine pellet | Restaging | 93 | 0 | 0 | 554566 | 0.00000% |
| INDIA 8576 | Urine supernatant | Restaging | 90 | 2 | 2 | 164 | 1.21951% |
| INDIA 8576 | Urine | Restaging | 90 | 44 | 68 | 2205 | 3.08390% |
| INDIA 8577 | Urine pellet | Restaging | 96 | 0 | 0 | 534381 | 0.00000% |
| INDIA 8578 | Urine supernatant | Restaging | 96 | 57 | 157 | 2702 | 5.81051% |
| INDIA 8578 | Urine | Restaging | 96 | 65 | 217 | 4296 | 5.05121% |
| INDIA 8579 | Urine | Restaging | 94 | 81 | 248 | 297457 | 0.08337% |
| INDIA 8579 | Urine pellet | Restaging | 94 | 28 | 38 | 61149 | 0.06214% |
| INDIA 8580 | Urine | Restaging | 93 | 0 | 0 | 103342 | 0.00000% |
| INDIA 8580 | Urine pellet | Restaging | 93 | 0 | 0 | 88898 | 0.00000% |
| INDIA 8581 | Urine supernatant | Restaging | 95 | 24 | 28 | 12620 | 0.22187% |
| INDIA 8581 | Urine | Restaging | 95 | 68 | 213 | 70552 | 0.30190% |
| INDIA 8582 | Urine pellet | Restaging | 94 | 54 | 291 | 524178 | 0.05552% |
| INDIA 8585 | Urine pellet | Restaging | 95 | 45 | 80 | 19023 | 0.42054% |
| INDIA 8586 | Urine pellet | Restaging | 96 | 49 | 282 | 332300 | 0.08486% |
| INDIA 8587 | Urine pellet | Restaging | 92 | 0 | 0 | 236544 | 0.00000% |
| INDIA 8588 | Urine pellet | Restaging | 92 | 0 | 0 | 907976 | 0.00000% |
| INDIA 8590 | Urine pellet | Restaging | 95 | 0 | 0 | 605637 | 0.00000% |
| INDIA 8591 | Urine pellet | Restaging | 93 | 92 | 6432 | 788849 | 0.81537% |
| INDIA 8592 | Urine pellet | Restaging | 93 | 0 | 0 | 37099 | 0.00000% |
| INDIA 8613 | Urine pellet | Restaging | 95 | 0 | 0 | 769375 | 0.00000% |
| INDIA 8595 | Urine pellet | Restaging | 94 | 0 | 0 | 19840 | 0.00000% |
| INDIA 8598 | Urine pellet | Restaging | 93 | 91 | 8755 | 966809 | 0.90556% |
| INDIA 8599 | Urine pellet | Restaging | 92 | 74 | 1620 | 256781 | 0.63089% |
| INDIA 8601 | Urine pellet | Restaging | 61 | 0 | 0 | 182010 | 0.00000% |
| INDIA 8614 | Urine pellet | Restaging | 24 | 20 | 216 | 277132 | 0.07794% |
| INDIA 8605 | Urine pellet | Restaging | 94 | 2 | 2 | 1030343 | 0.00019% |
| INDIA 8607 | Urine pellet | Restaging | 82 | 11 | 33 | 89459 | 0.03689% |
| INDIA 8609 | Urine pellet | Restaging | 92 | 90 | 40554 | 1110446 | 3.65205% |
| INDIA 8610 | Urine pellet | Restaging | 88 | 27 | 37 | 32991 | 0.11215% |
| INDIA 8611 | Urine pellet | Restaging | 91 | 48 | 115 | 781244 | 0.01472% |
| INDIA 8612 | Urine pellet | Restaging | 96 | 83 | 2528 | 833273 | 0.30338% |
| INDIA 8616 | Urine pellet | Restaging | 96 | 22 | 32 | 1499244 | 0.00213% |
| INDIA 8565 | Plasma | Baseline | 94 | 0 | 0 | 659904 | 0.00000% |
| INDIA 8577 | Plasma | Baseline | 96 | 0 | 0 | 872872 | 0.00000% |
| INDIA 8587 | Plasma | Baseline | 92 | 0 | 0 | 693574 | 0.00000% |
| INDIA 8592 | Plasma | Baseline | 93 | 0 | 0 | 294174 | 0.00000% |
| INDIA 8594 | Plasma | Baseline | 94 | 0 | 0 | 462470 | 0.00000% |
| INDIA 8559 | Plasma | Baseline | 90 | 0 | 0 | 650803 | 0.00000% |
| INDIA 8604 | Plasma | Baseline | 95 | 0 | 0 | 641828 | 0.00000% |

| | | | | | | | |
|---|---|---|---|---|---|---|---|
| INDIA 8588 | Plasma | Baseline | 92 | 0 | 0 | 647640 | 0.00000% |
| INDIA 8593 | Plasma | Baseline | 96 | 0 | 0 | 528727 | 0.00000% |
| INDIA 8544 | Plasma | Baseline | 75 | 0 | 0 | 373219 | 0.00000% |
| INDIA 8550 | Plasma | Baseline | 84 | 0 | 0 | 637805 | 0.00000% |
| INDIA 8564 | Plasma | Baseline | 90 | 0 | 0 | 301365 | 0.00000% |
| INDIA 8610 | Plasma | Baseline | 88 | 0 | 0 | 1316234 | 0.00000% |
| INDIA 8549 | Plasma | Baseline | 96 | 6 | 6 | 730590 | 0.00000% |
| INDIA 8579 | Plasma | Baseline | 94 | 12 | 15 | 314980 | 0.00000% |
| INDIA 8580 | Plasma | Baseline | 93 | 2 | 2 | 351997 | 0.00000% |
| INDIA 8583 | Plasma | Baseline | 88 | 27 | 37 | 394210 | 0.00000% |
| INDIA 8600 | Plasma | Baseline | 92 | 91 | 980 | 590115 | 0.00000% |
| INDIA 8601 | Plasma | Baseline | 61 | 16 | 22 | 285079 | 0.00000% |
| INDIA 8611 | Plasma | Baseline | 91 | 9 | 11 | 826999 | 0.00000% |
| INDIA 8575 | Plasma | Baseline | 93 | 93 | 7882 | 1478216 | 0.00000% |
| INDIA 8595 | Plasma | Baseline | 94 | 55 | 376 | 376230 | 0.00000% |
| INDIA 8561 | Plasma | Baseline | 93 | 68 | 204 | 1158461 | 0.00000% |
| INDIA 8566 | Plasma | Baseline | 93 | 4 | 4 | 248728 | 0.00000% |
| INDIA 8568 | Plasma | Baseline | 96 | 77 | 224 | 896424 | 0.00000% |
| INDIA 8571 | Plasma | Baseline | 92 | 38 | 63 | 480268 | 0.00000% |
| INDIA 8548 | Plasma | Baseline | 85 | 0 | 0 | 63767 | 0.00000% |
| INDIA 8548 | Plasma | Baseline | 85 | 1 | 1 | 363869 | 0.00000% |
| INDIA 8549 | Plasma | End of treatment | 96 | 0 | 0 | 350593 | 0.00000% |
| INDIA 8565 | Plasma | End of treatment | 94 | 0 | 0 | 395489 | 0.00000% |
| INDIA 8577 | Plasma | End of treatment | 96 | 1 | 1 | 658860 | 0.00000% |
| INDIA 8579 | Plasma | End of treatment | 94 | 0 | 0 | 430058 | 0.00000% |
| INDIA 8580 | Plasma | End of treatment | 93 | 0 | 0 | 332028 | 0.00000% |
| INDIA 8583 | Plasma | End of treatment | 88 | 0 | 0 | 213378 | 0.00000% |
| INDIA 8587 | Plasma | End of treatment | 92 | 0 | 0 | 843228 | 0.00000% |
| INDIA 8592 | Plasma | End of treatment | 93 | 0 | 0 | 775428 | 0.00000% |
| INDIA 8613 | Plasma | End of treatment | 95 | 0 | 0 | 426526 | 0.00000% |
| INDIA 8600 | Plasma | End of treatment | 92 | 1 | 1 | 849319 | 0.00000% |
| INDIA 8588 | Plasma | End of treatment | 92 | 0 | 0 | 807129 | 0.00000% |
| INDIA 8593 | Plasma | End of treatment | 96 | 0 | 0 | 216497 | 0.00000% |
| INDIA 8595 | Plasma | End of treatment | 94 | 0 | 0 | 377884 | 0.00000% |
| INDIA 8559 | Plasma | End of treatment | 90 | 0 | 0 | 775435 | 0.00000% |
| INDIA 8575 | Plasma | End of treatment | 93 | 0 | 0 | 2368558 | 0.00000% |
| INDIA 8544 | Plasma | End of treatment | 75 | 0 | 0 | 110403 | 0.00000% |
| INDIA 8561 | Plasma | End of treatment | 93 | 0 | 0 | 348085 | 0.00000% |
| INDIA 8564 | Plasma | End of treatment | 90 | 0 | 0 | 519029 | 0.00000% |
| INDIA 8566 | Plasma | End of treatment | 93 | 0 | 0 | 297704 | 0.00000% |
| INDIA 8568 | Plasma | End of treatment | 96 | 0 | 0 | 447372 | 0.00000% |
| INDIA 8610 | Plasma | End of treatment | 88 | 0 | 0 | 241607 | 0.00000% |
| INDIA 8548 | Plasma | Restaging | 85 | 0 | 0 | 598569 | 0.00000% |
| INDIA 8549 | Plasma | Restaging | 96 | 1 | 1 | 566247 | 0.00000% |
| INDIA 8565 | Plasma | Restaging | 94 | 0 | 0 | 709277 | 0.00000% |
| INDIA 8575 | Plasma | Restaging | 93 | 0 | 0 | 1070982 | 0.00000% |
| INDIA 8577 | Plasma | Restaging | 96 | 1 | 1 | 1247794 | 0.00000% |
| INDIA 8579 | Plasma | Restaging | 94 | 1 | 1 | 1911890 | 0.00000% |
| INDIA 8580 | Plasma | Restaging | 93 | 0 | 0 | 882972 | 0.00000% |
| INDIA 8587 | Plasma | Restaging | 92 | 0 | 0 | 1613180 | 0.00000% |
| INDIA 8592 | Plasma | Restaging | 93 | 0 | 0 | 908801 | 0.00000% |
| INDIA 8613 | Plasma | Restaging | 95 | 0 | 0 | 919358 | 0.00000% |
| INDIA 8600 | Plasma | Restaging | 92 | 1 | 1 | 712834 | 0.00000% |
| INDIA 8601 | Plasma | Restaging | 61 | 0 | 0 | 138851 | 0.00000% |
| INDIA 8611 | Plasma | Restaging | 91 | 0 | 0 | 988590 | 0.00000% |
| INDIA 8544 | Plasma | Restaging | 75 | 0 | 0 | 314092 | 0.00000% |
| INDIA 8550 | Plasma | Restaging | 84 | 0 | 0 | 618564 | 0.00000% |
| INDIA 8561 | Plasma | Restaging | 93 | 0 | 0 | 488898 | 0.00000% |
| INDIA 8564 | Plasma | Restaging | 90 | 0 | 0 | 824056 | 0.00000% |
| INDIA 8566 | Plasma | Restaging | 93 | 0 | 0 | 471697 | 0.00000% |
| INDIA 8568 | Plasma | Restaging | 96 | 0 | 0 | 1551525 | 0.00000% |
| INDIA 8571 | Plasma | Restaging | 92 | 0 | 0 | 1365992 | 0.00000% |
| INDIA 8588 | Plasma | Restaging | 92 | 0 | 0 | 4095401 | 0.00000% |
| INDIA 8593 | Plasma | Restaging | 96 | 0 | 0 | 787836 | 0.00000% |
| INDIA 8595 | Plasma | Restaging | 94 | 0 | 0 | 792900 | 0.00000% |
| INDIA 8610 | Plasma | Restaging | 88 | 0 | 0 | 5870684 | 0.00000% |
| INDIA 8548 | Tumor | Baseline | 85 | 85 | 623 | 2753 | 22.62986% |

| | | | | | | | |
|---|---|---|---|---|---|---|---|
| INDIA 8549 | Tumor | Baseline | 96 | 3 | 3 | 5 | 60.00000% |
| INDIA 8559 | Tumor | Baseline | 90 | 90 | 9532 | 40966 | 23.26808% |
| INDIA 8565 | Tumor | Baseline | 94 | 94 | 47718 | 118696 | 40.20186% |
| INDIA 8575 | Tumor | Baseline | 93 | 93 | 47034 | 131636 | 35.73035% |
| INDIA 8577 | Tumor | Baseline | 96 | 96 | 993 | 2504 | 39.65655% |
| INDIA 8579 | Tumor | Baseline | 94 | 94 | 41533 | 159950 | 25.96624% |
| INDIA 8580 | Tumor | Baseline | 93 | 93 | 123818 | 313688 | 39.47170% |
| INDIA 8583 | Tumor | Baseline | 88 | 88 | 8469 | 31772 | 26.65555% |
| INDIA 8587 | Tumor | Baseline | 92 | 92 | 81138 | 276596 | 29.33448% |
| INDIA 8592 | Tumor | Baseline | 93 | 93 | 286608 | 435498 | 65.81155% |
| INDIA 8594 | Tumor | Baseline | 94 | 94 | 41638 | 59054 | 70.50835% |
| INDIA 8613 | Tumor | Baseline | 95 | 95 | 71580 | 204864 | 34.94025% |
| INDIA 8600 | Tumor | Baseline | 92 | 92 | 61897 | 132669 | 46.65521% |
| INDIA 8601 | Tumor | Baseline | 61 | 58 | 271 | 1345 | 20.14870% |
| INDIA 8604 | Tumor | Baseline | 95 | 95 | 118567 | 197337 | 60.08351% |
| INDIA 8611 | Tumor | Baseline | 91 | 90 | 826 | 1819 | 45.40957% |
| INDIA 8544 | Tumor | Baseline | 75 | 75 | 2178 | 12150 | 17.92593% |
| INDIA 8550 | Tumor | Baseline | 84 | 84 | 25073 | 87244 | 28.73894% |
| INDIA 8561 | Tumor | Baseline | 93 | 93 | 1861 | 5572 | 33.39914% |
| INDIA 8564 | Tumor | Baseline | 90 | 89 | 164955 | 198454 | 83.12002% |
| INDIA 8566 | Tumor | Baseline | 93 | 92 | 453 | 1194 | 37.93970% |
| INDIA 8568 | Tumor | Baseline | 96 | 96 | 107641 | 298248 | 36.09111% |
| INDIA 8571 | Tumor | Baseline | 92 | 92 | 784 | 2300 | 34.08696% |
| INDIA 8588 | Tumor | Baseline | 92 | 92 | 31041 | 168270 | 18.44714% |
| INDIA 8593 | Tumor | Baseline | 96 | 96 | 46952 | 188564 | 24.89977% |
| INDIA 8595 | Tumor | Baseline | 94 | 94 | 169876 | 398068 | 42.67512% |
| INDIA 8610 | Tumor | Baseline | 88 | 86 | 501 | 1013 | 49.45706% |
| INDIA 8584 | Plasma | Baseline | 46 | 18 | 74 | 335221 | 0.00000% |
| INDIA 8590 | Plasma | Baseline | 95 | 87 | 543 | 1133621 | 0.00000% |
| INDIA 8556 | Plasma | Baseline | 39 | 0 | 0 | 243385 | 0.00000% |
| INDIA 8560 | Plasma | Baseline | 89 | 88 | 2518 | 1704861 | 0.00000% |
| INDIA 8556 | Tumor | Baseline | 39 | 39 | 2379 | 122445 | 1.94291% |
| INDIA 8560 | Tumor | Baseline | 89 | 89 | 28823 | 59603 | 48.35830% |
| INDIA 8584 | Tumor | Baseline | 46 | 46 | 759 | 2849 | 26.64093% |
| INDIA 8590 | Tumor | Baseline | 95 | 95 | 84925 | 231327 | 36.71210% |
| INDIA 8590 | PBMC | Baseline | 95 | 0 | 0 | 496550 | 0.00000% |
| INDIA 8557 | Plasma | Baseline | 93 | 0 | 0 | 322084 | 0.00000% |
| INDIA 8572 | Plasma | Baseline | 93 | 1 | 1 | 311209 | 0.00032% |
| INDIA 8581 | Plasma | Baseline | 95 | 0 | 0 | 260745 | 0.00000% |
| INDIA 8599 | Plasma | Baseline | 92 | 0 | 0 | 675111 | 0.00000% |
| INDIA 8603 | Plasma | Baseline | 82 | 1 | 1 | 1168019 | 0.00009% |
| INDIA 8605 | Plasma | Baseline | 94 | 0 | 0 | 789379 | 0.00000% |
| INDIA 8607 | Plasma | Baseline | 82 | 0 | 0 | 1132051 | 0.00000% |
| INDIA 8552 | Plasma | Baseline | 94 | 93 | 2299 | 1552803 | 0.00000% |
| INDIA 8553 | Plasma | Baseline | 95 | 89 | 428 | 524997 | 0.00000% |
| INDIA 8563 | Plasma | Baseline | 94 | 72 | 207 | 328891 | 0.00000% |
| INDIA 8569 | Plasma | Baseline | 93 | 13 | 18 | 370868 | 0.00000% |
| INDIA 8570 | Plasma | Baseline | 92 | 69 | 180 | 1203490 | 0.00000% |
| INDIA 8573 | Plasma | Baseline | 89 | 9 | 10 | 605741 | 0.00000% |
| INDIA 8574 | Plasma | Baseline | 91 | 7 | 7 | 389857 | 0.00000% |
| INDIA 8576 | Plasma | Baseline | 90 | 2 | 2 | 794358 | 0.00000% |
| INDIA 8578 | Plasma | Baseline | 96 | 32 | 47 | 350521 | 0.00000% |
| INDIA 8582 | Plasma | Baseline | 94 | 2 | 2 | 295859 | 0.00000% |
| INDIA 8585 | Plasma | Baseline | 95 | 24 | 32 | 1173991 | 0.00000% |
| INDIA 8586 | Plasma | Baseline | 96 | 90 | 445 | 1389350 | 0.00000% |
| INDIA 8596 | Plasma | Baseline | 95 | 55 | 90 | 357725 | 0.00000% |
| INDIA 8598 | Plasma | Baseline | 93 | 4 | 15 | 1204035 | 0.00000% |
| INDIA 8609 | Plasma | Baseline | 92 | 4 | 7 | 640479 | 0.00000% |
| INDIA 8612 | Plasma | Baseline | 96 | 28 | 41 | 414770 | 0.00000% |
| INDIA 8608 | Plasma | Baseline | 64 | 7 | 7 | 514817 | 0.00000% |
| INDIA 8547 | Plasma | Baseline | 71 | 63 | 50315 | 691682 | 0.00000% |
| INDIA 8551 | Plasma | Baseline | 83 | 82 | 2908 | 728173 | 0.00000% |
| INDIA 8554 | Plasma | Baseline | 91 | 28 | 41 | 211025 | 0.00000% |
| INDIA 8555 | Plasma | Baseline | 92 | 26 | 29 | 495771 | 0.00000% |
| INDIA 8591 | Plasma | Baseline | 93 | 41 | 57 | 247814 | 0.00000% |
| INDIA 8597 | Plasma | Baseline | 96 | 6 | 7 | 351776 | 0.00000% |
| INDIA 8617 | Plasma | Baseline | 92 | 25 | 32 | 664888 | 0.00000% |

| | | | | | | | |
|---|---|---|---|---|---|---|---|
| INDIA 8547 | Tumor | Baseline | 71 | 71 | 16722 | 35976 | 46.48099% |
| INDIA 8551 | Tumor | Baseline | 83 | 83 | 6674 | 44597 | 14.96513% |
| INDIA 8552 | Tumor | Baseline | 94 | 94 | 49389 | 119898 | 41.19251% |
| INDIA 8553 | Tumor | Baseline | 95 | 95 | 2466 | 5558 | 44.36848% |
| INDIA 8554 | Tumor | Baseline | 91 | 91 | 20430 | 47978 | 42.58202% |
| INDIA 8555 | Tumor | Baseline | 92 | 92 | 15604 | 124011 | 12.58275% |
| INDIA 8557 | Tumor | Baseline | 93 | 93 | 13877 | 31520 | 44.02602% |
| INDIA 8563 | Tumor | Baseline | 94 | 94 | 28737 | 87483 | 32.84867% |
| INDIA 8569 | Tumor | Baseline | 93 | 93 | 67597 | 188322 | 35.89437% |
| INDIA 8570 | Tumor | Baseline | 92 | 92 | 459 | 923 | 49.72914% |
| INDIA 8572 | Tumor | Baseline | 93 | 93 | 49872 | 80058 | 62.29484% |
| INDIA 8573 | Tumor | Baseline | 89 | 89 | 10847 | 39488 | 27.46910% |
| INDIA 8574 | Tumor | Baseline | 91 | 91 | 56079 | 242350 | 23.13967% |
| INDIA 8576 | Tumor | Baseline | 90 | 90 | 6311 | 20426 | 30.89690% |
| INDIA 8578 | Tumor | Baseline | 96 | 96 | 125378 | 321774 | 38.96461% |
| INDIA 8581 | Tumor | Baseline | 95 | 95 | 169695 | 288432 | 58.83362% |
| INDIA 8582 | Tumor | Baseline | 94 | 94 | 5642 | 14940 | 37.76439% |
| INDIA 8585 | Tumor | Baseline | 95 | 95 | 200152 | 233766 | 85.62066% |
| INDIA 8586 | Tumor | Baseline | 96 | 96 | 6712 | 11650 | 57.61373% |
| INDIA 8591 | Tumor | Baseline | 93 | 90 | 551 | 1111 | 49.59496% |
| INDIA 8596 | Tumor | Baseline | 95 | 95 | 18417 | 27177 | 67.76686% |
| INDIA 8597 | Tumor | Baseline | 96 | 96 | 49713 | 64082 | 77.57717% |
| INDIA 8598 | Tumor | Baseline | 93 | 93 | 119159 | 163369 | 72.93856% |
| INDIA 8599 | Tumor | Baseline | 92 | 91 | 927 | 7182 | 12.90727% |
| INDIA 8614 | Tumor | Baseline | 24 | 23 | 137 | 3934 | 3.48246% |
| INDIA 8603 | Tumor | Baseline | 82 | 82 | 1826 | 9171 | 19.91059% |
| INDIA 8605 | Tumor | Baseline | 94 | 94 | 70357 | 109448 | 64.28350% |
| INDIA 8607 | Tumor | Baseline | 82 | 41 | 106 | 707 | 14.99293% |
| INDIA 8608 | Tumor | Baseline | 64 | 64 | 48372 | 133785 | 36.15652% |
| INDIA 8609 | Tumor | Baseline | 92 | 92 | 56124 | 110239 | 50.91120% |
| INDIA 8612 | Tumor | Baseline | 96 | 96 | 91770 | 166862 | 54.99754% |
| INDIA 8616 | Tumor | Baseline | 96 | 96 | 35635 | 86321 | 41.28196% |
| INDIA 8617 | Tumor | Baseline | 92 | 92 | 23705 | 88027 | 26.92924% |
| INDIA 8596 | Plasma | Restaging | 95 | 30 | 40 | 647428 | 0.00618% |
| INDIA 8616 | Plasma | Restaging | 96 | 60 | 139 | 3022436 | 0.00460% |
| INDIA 8598 | Plasma | Restaging | 93 | 8 | 69 | 3115169 | 0.00221% |
| INDIA 8569 | Plasma | Restaging | 93 | 4 | 4 | 233100 | 0.00172% |
| INDIA 8582 | Plasma | Restaging | 94 | 16 | 19 | 6263631 | 0.00030% |
| INDIA 8591 | Plasma | Restaging | 93 | 90 | 961 | 343296 | 0.27993% |
| INDIA 8597 | Plasma | Restaging | 96 | 31 | 1108 | 500501 | 0.22138% |
| INDIA 8555 | Plasma | Restaging | 92 | 25 | 28 | 526635 | 0.00532% |
| INDIA 8551 | Plasma | Restaging | 83 | 12 | 13 | 683524 | 0.00190% |
| INDIA 8547 | Plasma | Restaging | 71 | 3 | 3 | 446463 | 0.00067% |
| INDIA 8554 | Plasma | Restaging | 91 | 1 | 1 | 507476 | 0.00020% |
| INDIA 8599 | Plasma | Restaging | 92 | 1 | 1 | 869370 | 0.00012% |
| INDIA 8557 | Plasma | Restaging | 93 | 0 | 0 | 538349 | 0.00000% |
| INDIA 8572 | Plasma | Restaging | 93 | 0 | 0 | 374545 | 0.00000% |
| INDIA 8552 | Plasma | Restaging | 94 | 0 | 0 | 1860007 | 0.00000% |
| INDIA 8553 | Plasma | Restaging | 95 | 0 | 0 | 1707867 | 0.00000% |
| INDIA 8563 | Plasma | Restaging | 94 | 0 | 0 | 611061 | 0.00000% |
| INDIA 8570 | Plasma | Restaging | 92 | 0 | 0 | 2019152 | 0.00000% |
| INDIA 8573 | Plasma | Restaging | 89 | 0 | 0 | 778694 | 0.00000% |
| INDIA 8574 | Plasma | Restaging | 91 | 0 | 0 | 3130136 | 0.00000% |
| INDIA 8576 | Plasma | Restaging | 90 | 0 | 0 | 5056336 | 0.00000% |
| INDIA 8581 | Plasma | Restaging | 95 | 0 | 0 | 800348 | 0.00000% |
| INDIA 8585 | Plasma | Restaging | 95 | 0 | 0 | 773005 | 0.00000% |
| INDIA 8586 | Plasma | Restaging | 96 | 0 | 0 | 1394594 | 0.00000% |
| INDIA 8605 | Plasma | Restaging | 94 | 0 | 0 | 1928595 | 0.00000% |
| INDIA 8607 | Plasma | Restaging | 82 | 0 | 0 | 821168 | 0.00000% |
| INDIA 8609 | Plasma | Restaging | 92 | 0 | 0 | 1074190 | 0.00000% |
| INDIA 8612 | Plasma | Restaging | 96 | 0 | 0 | 450862 | 0.00000% |
| INDIA 8614 | Plasma | Restaging | 24 | 0 | 0 | 137356 | 0.00000% |
| INDIA 8617 | Plasma | Restaging | 92 | 0 | 0 | 806365 | 0.00000% |
